# Calibrating machine learning approaches for probability estimation without calibration data

**DOI:** 10.64898/2026.07.10.26357723

**Authors:** Eleonora Di Carluccio, Georgios Koliopanos, Francisco M. Ojeda, Christian Weimar, Andreas Ziegler

**Author notes:** **Correspondence** Corresponding author Andreas Ziegler, Cardio-CARE, Medizincampus Davos, Herman-Burchard-Str. 12, 7265 Davos, Wolfgang, Switzerland.

## Abstract

Statistical prediction models for binary outcomes are becoming increasingly popular. One significant challenge is calibrating these models to suit the characteristics of a target population that is structurally different from the original population. Calibration is especially challenging when there is no training data available from the target population. To address this problem, we propose a novel calibration method, SimCal, which uses synthetic data generated from the model development data in conjunction with marginal statistics from the calibration cohort. We show that expert judgment modeling (EJM) may be used for calibration if cross-sectional data from the target population are available comprising expert judgments about the potential outcome and the covariates. We describe three alternative calibration approaches when calibration data are lacking: similarity-binning averaging (SBA), adaptive calibration of predictions (ACP), and Elkan calibration. In a simulation study, we compare SBA, ACP, Elkan calibration, and SimCal. R code for applying these methods is provided from the re-analysis of data on coronary artery disease. We illustrate all 5 calibration approaches with a real data set for predicting functional outcome after stroke and all approaches but EJM in the re-analysis of the Cleveland Clinic data. None of the approaches performed convincingly well in all situations. SimCal performed well when model parameters were correctly specified. EJM failed on the stroke data. Further research is urgently required for calibration in the absence of calibration data.

## 1 INTRODUCTION

Estimating outcome probabilities is crucial in life sciences. Such prediction models can be developed using classical statistical methods, such as logistic regression (LogReg). Alternatively, machine learning methods for probability estimation can be used, which are referred to as probability machines ^1,2^. Before the developed model can be applied to a sample of new patients in different centers or clinical settings, it generally needs to be calibrated to the new population. For example, if the model was developed with patient data from a university medical center, but is now to be applied to patients who are cared for by an emergency doctor on an ambulance, the adequacy of the model predictions should be checked. Calibration thus denotes the process of adjusting a model’s predicted probabilities so that they reflect the true probabilities of outcomes in a new population. This process is also referred to as recalibration in the literature ^3^. Phrased differently, a well-calibrated model outputs probabilities that closely correspond to actual observed relative frequencies. Calibration can be performed with many different statistical approaches, which have recently been evaluated and summarized ^4–6^.

The standard algorithm for calibration is identical for all classical calibration approaches, and it is laid out in algorithm 1. The key assumption of this general calibration algorithm is that validation data are available ^5^ (Figure 1). These are, in turn, used to adapt the prediction model to the population structure represented by the validation data.

**FIGURE 1.**
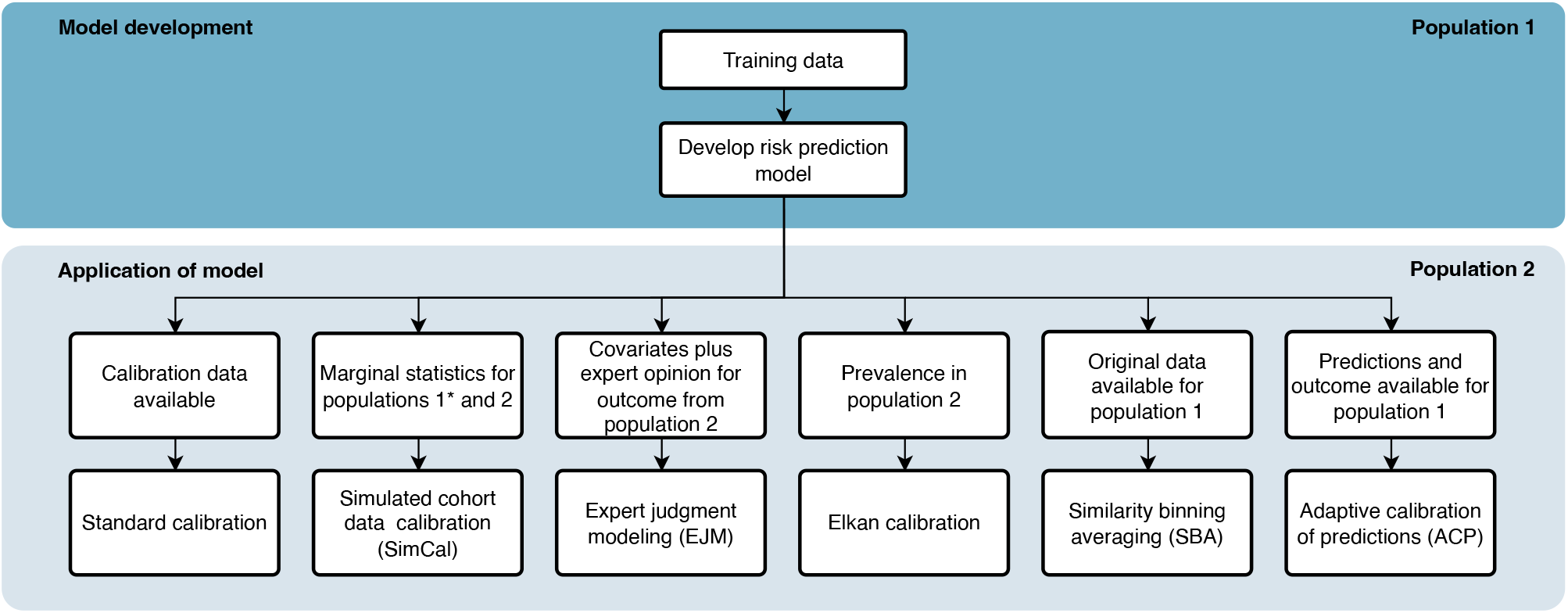
Calibration strategies depending on the availability of calibration data. ^∗^: The original data from population 1 may alternatively be used if available.

However, what can be done in the many cases where calibration data is not available? Some researchers have argued that the training data itself can serve as the calibration dataset ^7^. For example, Platt ^8^ proposed using either cross-validation or a hold-out set to estimate the calibration function, a transformation function that maps the originally estimated probabilities to calibrated probability estimates ^9^. The disadvantage of either approach, when relying solely on data from population 1, is that such data may not reflect the distributional structure of individuals in population 2. These approaches may thus not be well suited to address the conceptual demands of the task.

### Algorithm 1

The general calibration approach. Adapted from Ojeda and colleagues ^5^.

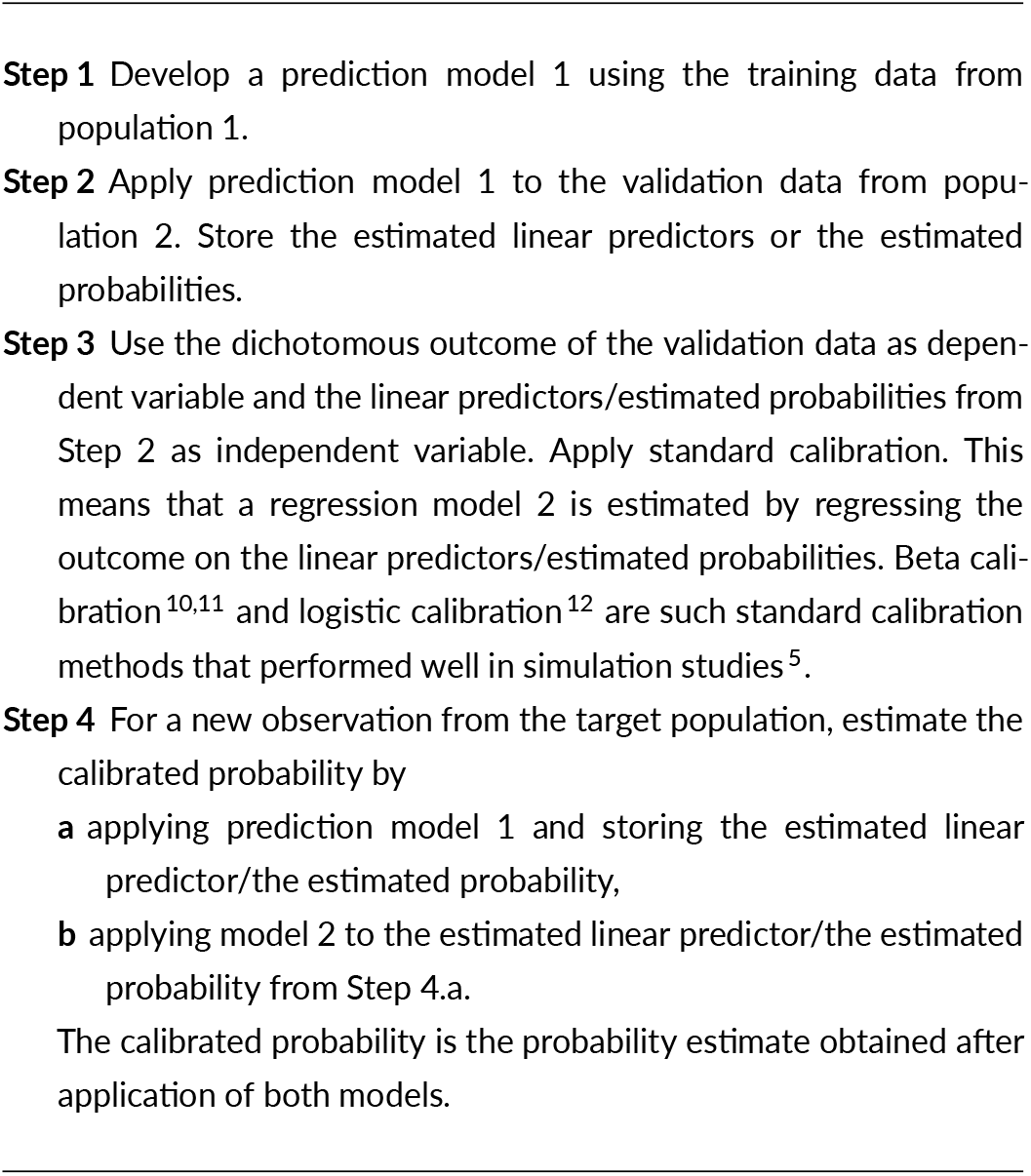

The aim of this work is four-fold. First, we propose a novel approach (simulated calibration; SimCal) for calibration in the absence of calibration data, i.e., for out-of-population predictions (Figure 1). For SimCal, a synthetic cohort is generated in the first step by simulating a large cohort that mimics the properties of the training data. In the second step, the marginal statistics from the simulated cohort are transferred to the assumed marginal statistics of the target population. In the last step, a calibration model is estimated using the transferred simulated data and may then be applied to new data from the target population.

Second, we demonstrate how expert judgment modeling (EJM) may be used for calibration. EJM ^13^ is also termed judgmental bootstrap ^14^, but is entirely unrelated to the standard statistical resampling technique. Synonymous terms include expert elicitation ^15,16^ and policy capturing ^14^. The fundamental idea of EJM is that experts evaluate observations or representative cases using their domain knowledge to make informed guesses of outcomes, and their judgments form a calibration data set.

Third, we compare SimCal with three further alternative approaches for out-of-population predictions. One alternative approach is similarity binning averaging (SBA) ^7^ (Figure 1). Its key idea is that, for a new data point, the predictions of its k nearest neighbors in the training data are averaged, and this average is taken as the calibrated probability. SBA uses both the information from features, synonymously termed covariates, and the estimated outcome probability for determining the k nearest neighbors. This is different from the second calibration method, termed adaptive calibration of predictions (ACP) which only uses the outcome information for identifying similar observations from the training data ^17^.

The calibration approach by Elkan ^18^ does not require re-estimation from a regression model. It relies only on the availability of a prevalence estimate of the outcome in the target population, which comes with an important assumption (Figure 1). Elkan calibration is guaranteed to be valid if the covariate distributions of both populations 1 and 2, i.e, those for the training data and the target population are identical ^18^.

Fourth, we make these calibrations easy to apply in practice by providing R code ^19^ for estimating the learning machines.

The outline of this work is as follows. In the next Section 2, we introduce SimCal, the use of EJM, and the three additional calibration approaches in detail. Section 3 describes the design and results of simulation studies for SimCal, SBA, ACP, and Elkan calibration in various settings. A specific part of the simulations is to investigate the effect of model misspecification for SimCal and for Elkan calibration. In our opinion, it is impossible to provide realistic simulation scenarios for EJM, and we therefore refrained from incorporating this method in the simulation study. In Section 3.5, we show the application of all 5 calibration approaches, including EJM, for predicting functional outcome and survival at 100 days within the first 6 hours after onset of acute cerebral ischemia ^20^. A data set on coronary heart disease (CHD) is freely available from a data repository. This data set allows the application of SBA, ACP, Elkan calibration, and SimCal. All analysis code for estimating the learning machines for this data set plus the code for the calibration methods is provided in the Supplementary Material. The R code for the simulation studies is available as Supplementary Material. The simulated data used in this work is available on Zenodo ^21^.

## 2 CALIBRATION METHODS

A brief summary of the different calibration approaches is provided in Figure 1. In all cases, it is assumed that training data from population 1 have been used to develop a risk prediction model, which may be, e.g., a prognostic or a diagnostic model. To apply the model to population 2, a standard calibration approach can be used if validation data is available (Figure 1 and Algorithm 1). If validation data is not available, one of the 5 approaches described below can be used.

### 2.1 Simulation-based calibration (SimCal)

Simulation-based calibration (SimCal) relies on the idea that one might be able to provide marginal statistics from population 2, i.e., the validation population, for the variables included in the prediction model. It is assumed that for all categorical variables proportions are specified for the validation data. These proportions may be estimated from an external data source, such as a hospital information system or a clinical or public health database. If such data are unavailable from an external data source, the estimates may be individual guesses from a treating clinician or from a group of clinical experts. For all quantitative variables, means and variances also need to be specified. Moreover, pairwise correlation coefficients between variables need to be available for the simulation process. In cases when no information is available, a natural choice is to estimate the variances and correlations from the training data from population 1.

The concept of SimCal is depicted in Figure 2, and the standard SimCal algorithm is described in Algorithm 2. We assume that a prediction model has been developed using the training data from population 1 (Step 1). The training data are also used to estimate the parameters required for the simulation of a cohort in Step 2. Specifically, Step 2 consists in transforming the original training data by the rank inverse normal transformation. Technical details for this transformation have been described elsewhere ^24^.

**FIGURE 2.**
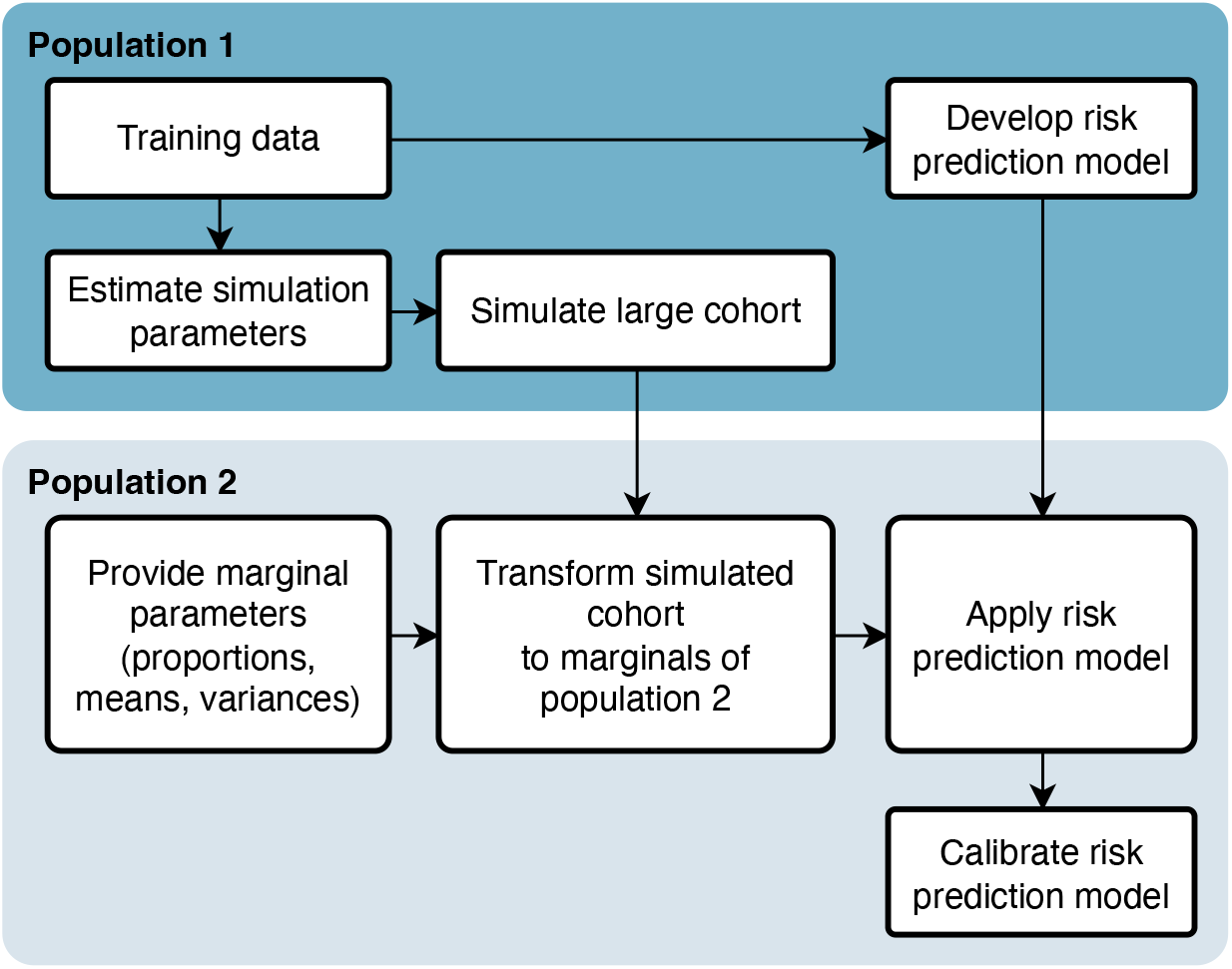
Concept of simulated calibration (SimCal), a calibration approach relying on synthetic data.

In brief, the rank inverse normal transformation is a data transformation technique that maps observed values to standard normal quantiles by first ranking the observations and then applying the inverse normal distribution function to the resulting ranks. It is used when the normality assumption of a statistical method may be violated, as it guarantees that the transformed variable follows a normal distribution, irrespective of the shape of the original distribution. However, while the marginal distribution of a variable is normalized, it does not necessarily preserve the underlying correlation structure or the interpretability of the original scale. This aspect is generally addressed by using the rank-transformed variables to estimate the correlation/covariance matrix for all variables. Tetrachoric/polychoric and biserial/polyserial correlation coefficients are the default for combinations of dichotomous/multi-categorical variables ^24^. These correlation coefficients are specifically designed for categorical and mixed-type variable combinations, providing more accurate estimates of the underlying latent continuous normal distributions when compared with the standard Pearson correlation. For estimating the correlation/covariance matrix, we use the `R` package `modgo` ^22^. Alternative R packages are available, see, e.g., `sbgcop` ^26^.

In Step 3, a large cohort is simulated with the aim to reflect a large sample from population 1. Since simulation is done only once, we recommend using a very large sample size, and we have been working with at least 100, 000 observations throughout. For the simulations, the correlation/covariance matrix obtained in step 2 is used as input, and the samples are generated with the `mrvnorm` function from the `MASS` package ^25^. After simulation on the normal distribution scale, the data need to be transferred back to the original scale of the covariates. We recommend applying the cumulative distribution function (CDF) of the standard normal followed by the inverse of the empirical CDF of each variable because this is more precise than the back transformation with the quantile function from the generalized lambda distribution (GLD) ^22^. The rank inverse normal transformation requires, however, the availability of the quantiles from the training data. In contrast, the GLD only needs the estimates of the first four moments. The GLD thus has the advantage that no individual information is necessary from the training data, and only the marginal parameter estimates from the GLD need to be available.

Marginal statistics for the target population, i.e., population 2 need to be provided (Step 4) so that the simulated data are transferred from population 1 to the target population 2 (Step 5). To this end, let *µ*_*pop*_ and 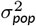 denote the mean and variance of a quantitative variable for population *pop*, then a simulated observation *x*_*i*1_ for the simulated individual *i* from population 1 is transferred to population 2 using

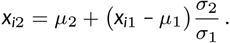

For a dichotomous or a multicategorical variable, proportions can be changed on the scale of the simulated normal distribution. For example, if the proportion in population 1 was 0.8, the threshold for dichotomization was 0.84. If the proportion in population 2 is 0.9, the corresponding threshold is 1.28.

The next step after the transformation is to estimate the calibration function on the simulated data (Step 6). To this end, let **x**_*i*_ denote the features, i.e., covariates, also termed independent variables and apply prediction model 1 to the simulated data *i* = 1, …, *n*_*sim*_. The linear predictor 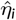 or the estimated probability are stored. We denote the simulated outcome of individual *i* by 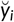. In Step 7, the simulated outcome 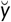 is regressed against the estimated linear predictor 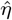 from Step 6, and the calibration model 2 is ^5^

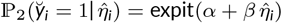

for logistic calibration ^5,11^, where expit(*x*) = exp(*x*)/(1 + exp(*x*)).

SimCal can be applied to a new observation from the target population by first applying prediction model 1 and storing the estimated linear predictor/the estimated probability (Step 8.a). Second, model 2 is applied to the estimated linear predictor/the estimated probability obtained in Step 8.a. The calibrated probability is the probability estimate obtained after applying model 2.

### 2.2 Expert judgment modeling

EJM is a prediction approach that integrates expert judgment with statistical models to improve predictive accuracy. It is commonly applied in domains such as finance, business, and marketing, where sufficient data for statistical analysis are difficult to obtain, yet predictions are required for decision-making ^27^. Further areas of application include sports, sociology, meteorology, environmental science, and public health ^27^.

The core principle of EJM is that domain experts assess observations and provide guesses of outcomes based on their knowledge. The expert guesses serve as the response variable in a regression or machine learning model, which learns to replicate the expert judgments from available data. Once trained, the model instead of the expert is used to make predictions for new observations. To illustrate the process, assume that a group of experienced cardiologists is given data on *n* patients, comprising variables such as gender, age, cholesterol levels, blood pressure, smoking, diabetes, and chest pain symptoms. For each patient, the cardiologists provide a subjective estimate of whether the patient has heart disease. In the second step, a statistical model, such as LogReg, or a machine learning model, is trained to approximate the expert guesses from the patient characteristics. The model thus learns patterns from the expert judgments. The resulting model is subsequently applied to predict the probability of heart disease in new patients.

#### Algorithm 2

Simulated calibration (SimCal) approach.

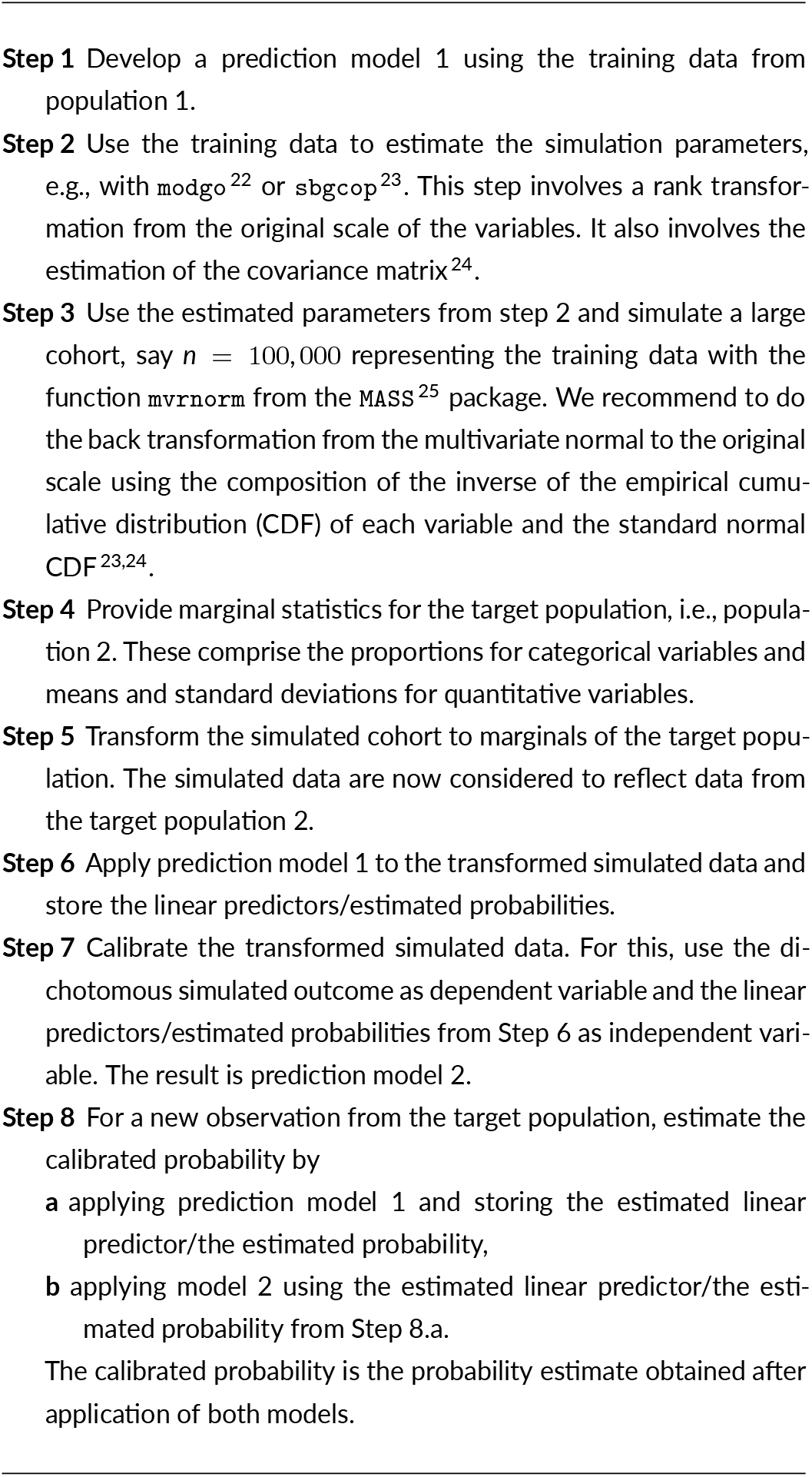

EJM may be used to calibrate a prediction model if baseline data are available for a cohort from the validation population 2, together with expert predictions for the outcome. The difference from standard calibration is that no true outcome data are required; only expert predictions are.

For expert judgment modeling (EJM), the following situation is considered. A prediction model 1 has been developed using training data from population 1. Furthermore, an additional dataset is available from population 2. This dataset includes information on all features ***x*** included in prediction model 1 so that the linear predictor 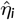 or the estimated probability can be obtained. The dataset does, however, not include information on the outcome variable *y*_*i*_. Instead, at least one prediction 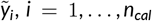, of the outcome is available which has been made by an expert for every individual *i*. The logistic calibration can then be estimated using the data pairs 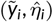:

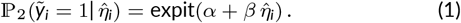

As an example, Section 3.5 considers the prediction of functional outcome 100 days after acute ischemic stroke. Clinical information is assumed to be available for all admitted patients, whereas the true 100day outcome is treated as unobserved at the time of prediction. Instead of the outcome, the treating physician’s assessment of the patient’s 100day prognosis, recorded within the first hours after stroke onset, serves as the basis for expert judgmental modeling.

In several applications, even baseline information ***x***_*i*_ might not be available. In such cases, one could expand the classical EJM approach and construct a set of artificial but representative cases 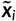 for population 2. These representative cases could then be rated by one or more experts ^15,16^. The expert-rated cases could, in turn, be used for calibration.

### 2.3 Elkan calibration

The three calibration methods described in this and the next two sections were proposed in the literature before. They are presented in their order of publication.

Elkan calibration ^18^ captures the difference in base rates between the two study populations. Let *b*_1_ = ℙ_1_(*y* = 1) and *b*_2_ = ℙ_2_(*y* = 1) denote the base rates, i.e., the unconditional event probability in the two populations in which the machine has been developed and to which it is to be applied, respectively. For a new observation *i* with features **x**_*i*_ from population 2, the probability ℙ_1_(*y*_*i*_ = 1|**x**_*i*_) is estimated by applying the learning machine for probability estimation developed on the training data. The calibrated probability ℙ_2_(*y*_*i*_ = 1|**x**_*i*_) can then be expressed as a function of ℙ_1_(*y*_*i*_ = 1|**x**_*i*_), *b*_1_, and *b*_2_

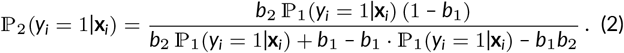

The validity of Elkan calibration relies on the assumption that the change in the base rate is the only difference between the two populations. The covariate distribution between both populations thus needs to be identical: ℙ_1_(**x**|*y* = 0) = ℙ_2_(**x**|*y* = 0) and ℙ_1_(**x**|*y* = 1) = ℙ_2_(**x**|*y* = 1). The same Bayes formula has been independently derived by Baker and colleagues ^28^.

The base probability *b*_1_ is generally estimated from the training data: 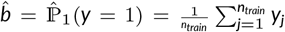. If validation data is not available, the event rate *b*_2_ may be estimated from an external data source, such as a hospital information system, a clinical or public health database or may be based on the group estimate or individual guesses from clinical experts.

In summary, Elkan calibration can be used for updating probability estimates in the absence of calibration data, and the transfer of information about the training data is not required. However, its applicability is limited by the assumption of equal covariate distributions in both populations ^5^.

### 2.4 Similarity binning averaging (SBA)

For Similarity Binning averaging (SBA) ^7^, consider a new observation *i* from population 2 with feature vector **x**_*i*_ and dichotomous outcome *y*_*i*_, for which a calibrated probability is to be obtained. It is assumed that the training data from population 1, used to develop the model, are available. The *k* nearest neighbors of observation *i* are identified within these training data based on the features **x** and the predicted probabilities 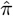 obtained from the model fitted on the training data. The calibrated probability 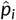 is then defined as the average of predicted probabilities 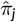 across the *k* nearest neighbors (see Algorithm 3).

#### Algorithm 3

Similarity-binning averaging (SBA) algorithm. Based on Bella and colleagues ^7^ and modified from Ojeda and colleagues ^5^.

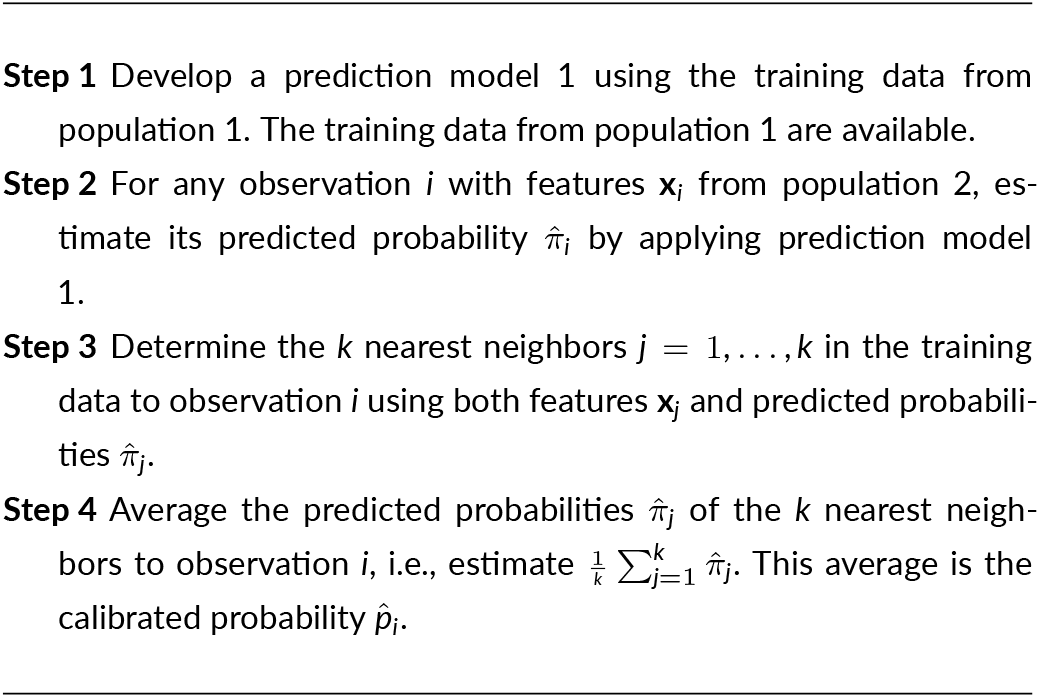

SBA is related to binning methods ^4,5,7^ in that the calibrated probability is derived from the *k* training observations most similar to the target observation from the population 2. A practical limitation of SBA is its computational burden for large datasets. Further, it requires the availability of all observations and the estimated probabilities from the training data. Furthermore, the choice of a metric for determining the *k* nearest neighbors was not discussed in detail by Bella and colleagues ^7^, who naturally used the Euclidean distance. Finally, the selection of *k* is a further tuning decision. Bella and colleagues ^7^ proposed *k* = 10 nearest neighbors, giving rise to SBA-10.

### 2.5 Adaptive calibration of predictions (ACP)

Adaptive Calibration of Predictions (ACP) is another binning method ^17^. It differs from SBA in the way bins are created. Suppose there is a new observation *i* with features **x**_*i*_ and estimated probability 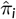, when the prediction model developed on the training data is applied. A confidence interval can be estimated for *π*_*i*_ by assuming that the new observation *i* is from population 1. Next, the *b* = 1, …, *n*_*b*_ individuals from the training data are identified that have probability estimates in the confidence interval. The calibrated probability is the proportion of 1s among the *n*_*b*_ individuals. ACP thus requires the individual predictions 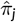 paired with the outcome information *y*_*j*_ for all *j* individuals from the training data. Algorithm 4 describes the ACP approach in detail. A shortcoming of ACP is that it has only been developed for LogReg for which the estimation of confidence intervals is straightforward. It has therefore not been used in performance reviews, see, e.g., Pakdaman Naeini and Cooper ^29^.

#### Algorithm 4

Adaptive calibration of predictions algorithm. Based on Jiang and colleagues ^17^.

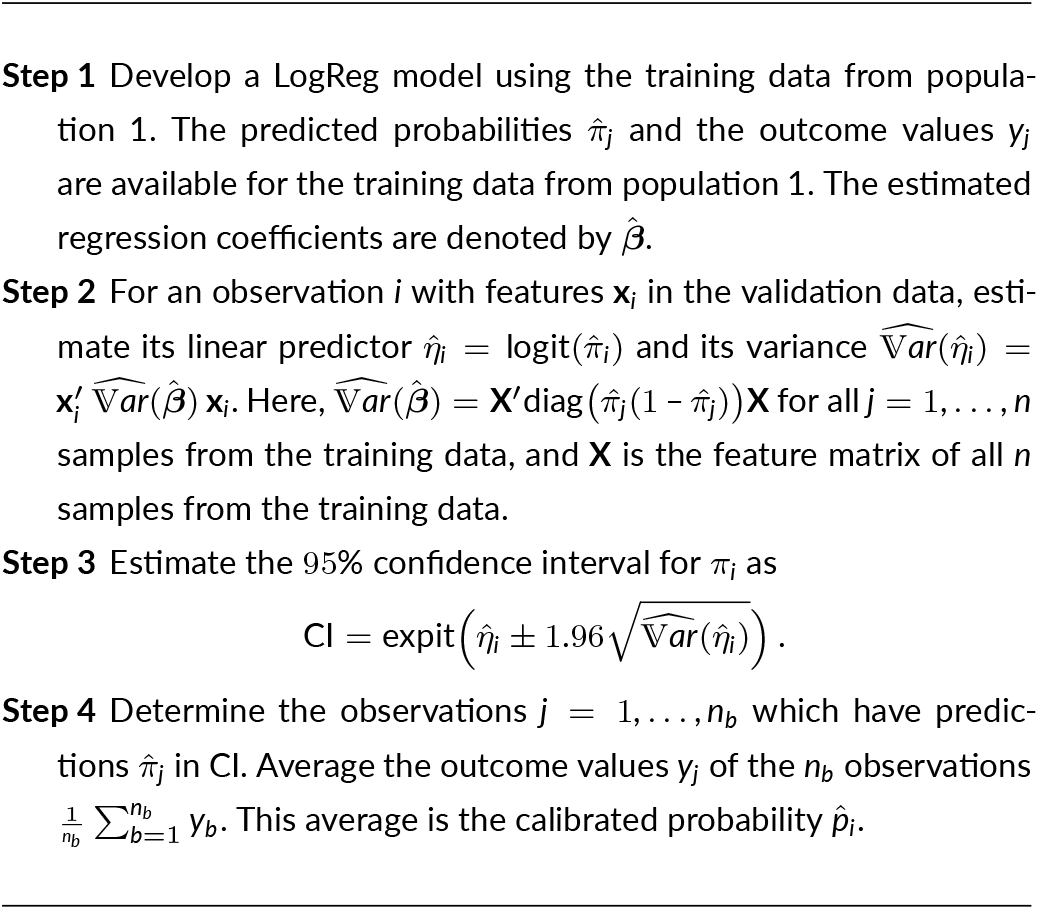

## 3 SIMULATION STUDIES AND REAL DATA ANALYSES

### 3.1 Simulation study 1: performance of SimCal compared with published alternative approaches

The aim of simulation study 1 is to compare the performance of SimCal with the previously published approaches Elkan calibration, SBA, and ACP. The design of the simulation study is based on Ojeda and colleagues ^5^, where detailed descriptions can be found. The code for the simulation study is provided in the Supplementary Material. Details are provided at the end of this section. We generated data from two populations with different disease prevalences. The data from the first population was used for model building. For a fair comparison with the reference methods, data from the second population were split into training (cal-training) data and test (cal-test) data.

The different simulation settings are summarized in Table 1. The numbering of the simulation scenarios follows the numbering of Ojeda and colleagues ^5^. Scenario 1 is a basic scenario with two continuous predictors and without noise variables. Scenario 5 is comparable to scenario 1, except that the disease prevalence in the model building and in the calibration data differs. Scenario 9 is again similar to scenarios 1 and 5, but it has 10 predictors, 5 being continuous and 5 being categorical. Scenario 14 has a smaller cal-training data set for estimating a calibration model with reference methods. Models 16 and 19 to 21 all investigate the effect of differences in the feature distribution or differences in the regression coefficients between training and calibration data. All 8 simulation scenarios were generated using a sample size of 1000 for model building, cal-training, and cal-test. The exception was scenario 14, where the cal-training data set was smaller and comprised only 100 observations. One-hundred replications were generated for each simulation scenario.

**TABLE 1.** Settings in the simulation scenarios: Number of continuous features (Cont), number of categorical features (Cat), outcome prevalence in model building data (Prev MB), outcome prevalence in cal-training data (Prev Cal) and, if applicable, additional distinctive feature (Distinctive feature). The sample size was 1000 for model building, 1000 for cal-training and 1000 for cal-test datasets, except for scenario 14, where the cal-training data had a sample size of 100.

| Scenario | Cont | Cat | Prev MB | Prev Cal | Distinctive feature |
| --- | --- | --- | --- | --- | --- |
| 1 | 2 | 0 | 0.5 | 0.69 | — |
| 5 | 2 | 0 | 0.2 | 0.41 | Lower disease prevalence |
| 9 | 5 | 5 | 0.57 | 0.77 | Additional covariates |
| 14 | 2 | 0 | 0.5 | 0.69 | Smaller calibration data set for training |
| 16 | 1 | 0 | 0.7 | 0.84 | Covariate distribution: MB $\mathcal{N}(1, 1)$, Cal $\mathcal{N}(1, 1)$ |
| 19 | 1 | 0 | 0.7 | 0.5 | Unequal covariate distribution: MB $\mathcal{N}(1, 1)$, Cal $\mathcal{N}(-1, 2)$ |
| 20 | 1 | 0 | 0.5 | 0.61 | Unequal coefficients: MB $\beta = 1$, Cal $\beta = 3$ |
| 21 | 1 | 0 | 0.5 | 0.7 | Unequal coefficients: MB $\beta = 1$, Cal $\beta = -1$ |

MB: Model building data; Cal: Calibration data.

LogReg was used to generate the data sets. For all models, features **x** = (*x*_1_, …, *x*_*q*_) were simulated according to predefined probability distributions, and regression coefficients *β*_1_, …, *β*_*q*_, and the intercepts *α* were set to specific values. The linear predictor *η* = *α*+*β*_1_*x*_1_ +…+*β*_*q*_*x*_*q*_ was expit-transformed *p* = ℙ(*y* = 1|**x**) = expit(*η*). The class of the outcome *y* was obtained from a Bernoulli distribution with event probability *p*. For simulation scenario 5, *p*^3^ was used as event probability instead of *p* to reduce the outcome prevalence, when sampling was done from the Bernoulli distribution. In all simulation scenarios, the intercept was set to 0 for the model building data, and it was set to 1 for the calibration datasets.

Continuous variables were generated according to a standard normal distribution, except for simulation scenario 16, where the *N* (1, 1) distribution was used for the model building and calibration data and scenario 19, where the distribution of the continuous variables differed between the model building data and the calibration data. The 5 additional categorical variables in simulation scenario 9 had respectively 2, 2, 3, 4, and 5 equally probable categories. The regression coefficients were identical in the model building data and the calibration data sets for all features, except for scenarios 20 and 21. In simulation scenario 20, the regression coefficient was 1 in the model building data and 3 in the calibration data. In simulation scenario 21, the regression coefficient was 1 in the model building data and −1 in the calibration data. This is the most challenging scenario for calibration because of the sign change of the regression coefficient.

The R code for generating the data for all simulation studies is available as Supplementary Material 2. The generated datasets are available for download on Zenodo ^21^. Supplementary Material 3.a provides the R Markdown code for generating the figures and tables for simulation study 1, and Supplementary Material 3.b contains the output of the first simulation study.

### 3.2 Simulation study 2: performance of SimCal with misspecified mean

The aim of simulation study 2 is to investigate the effect of misspecifying the mean of the marginal parameters for SimCal. Simulation Study 2 was based on simulation scenario 16 (Table 1), where the feature distribution in the model building data, the cal-training, and cal-test data was *N* (1, 1). For SimCal, normal distributions were assumed with variance 1 and varying means from 1.0 to 2.0 in steps of 0.1.

Supplementary Material 2 provides the R code for generating the datasets. The simulated datasets are available for download ^21^. Supplementary Material 4.a provides the R Markdown code for generating the figures and tables for simulation study 2, and Supplementary Material 4.b contains the output of this simulation study.

### 3.3 Simulation study 3: performance of SimCal with misspecified outcome probability

The aim of simulation study 3 is to investigate the effect of misspecifying the outcome probability, i.e., the prevalence ℙ_2_(*y* = 1) in the calibration data for SimCal. Simulation Study 3 was based on simulation scenario 16 (Table 1). The true prevalence in the calibration was 0.84, and we varied the assumed prevalence for SimCal from 0.76 to 0.92 in steps of 0.02.

Supplementary Material 2 provides the R code for generating the datasets. The simulated datasets are available for download ^21^. Supplementary Material 5.a provides the R Markdown code for generating the figures and tables for simulation study 3, and Supplementary Material

### 3.4 Simulation study 4: performance of SimCal with misspecified mean and misspecified outcome probability

The aim of simulation study 4 is to investigate the effect of misspecifying both the mean and the outcome probability in the calibration data for SimCal. Simulation Study 4 was again based on simulation scenario 16 (Table 1). The normally distributed independent variable was assumed to have variance 1. Misspecification of means was varied from −1.0 over 0.0 (no misspecification) to 1.0 in steps of 0.25. The true prevalence in the calibration data was 0.84, and the assumed prevalence for SimCal varied from 0.56, i.e., a misspecification of −0.28, to 0.96, i.e., a misspecification of 0.12, in steps of 0.04.

Supplementary Material 2 provides the R code for generating the datasets. The simulated datasets are available for download ^21^. Supplementary Material 6.a provides the R Markdown code for generating the figures and tables for simulation study 4, and Supplementary Material 6.b contains the output of the this simulation study.

### 3.5 Application 1: prediction of functional outcome after stroke—German Stroke Study Collaboration data

The German Stroke Study Collaboration investigated prognosis 100 days after acute ischemic stroke. The study has been described in detail elsewhere ^20,30,31^. For important information to comply with the Methods part of the TRIPOD statement ^32^, we refer to Supplementary Material 1. In brief, the training data comprised 1079 patients with ischemic stroke who had a Rankin Scale score < 3 before the stroke, who were admitted within 6 hours after the onset of the stroke symptoms, and who survived during the first 6 hours ^20^.

Training data were prospectively collected from 23 neurology departments with acute stroke units in 1998 and 1999, and 7 of those centers met the specified quality criteria. Before patients were enrolled in the validation study during 2001 and 2002, the required sample size was determined using simulations ^30^. Nine hospitals participated in the validation study, but not in the model building study. Seven of the 9 centers met the quality criteria and were thus used for external validation. Four hospitals meeting the quality criteria participated in both the model building and the calibration study, thus providing data for temporal validation. Patients were informed about study participation, and all patients provided informed consent for the transfer of their personal data to the data management center. The study was approved by the Ethics Committee of the Essen University Hospital, Germany. Patient characteristics are provided in Table 2.

**TABLE 2.** Patient characteristics in the stroke data. *n* (%) are displayed for dichotomous variables, mean and standard deviation (SD) for continuous variables.

| Variable | Training data (n = 1079) |  | Temporal validation (n = 976) |  | External validation (n = 1156) |  |
| --- | --- | --- | --- | --- | --- | --- |
| Barthel Index after 100 days: $\geq 95^*$ | 645 | (59.8%) | 505 | (51.7%) | 595 | (51.5%) |
| Mortality after 100 days: Yes | 123 | (11.4%) | 183 | (18.7%) | 240 | (20.8%) |
| Prior stroke: Yes | 210 | (19.5%) | 220 | (22.5%) | 244 | (21.1%) |
| Prior myocardial infarction: Yes | 160 | (14.8%) | 247 | (25.3%) | 244 | (21.1%) |
| Hypertension: Yes | 670 | (62.1%) | 293 | (30.0%) | 394 | (34.1%) |
| Atrial fibrillation on 1st ECG: Yes | 238 | (22.1%) | 223 | (22.9%) | 269 | (23.3%) |
| Diabetes: Yes | 215 | (19.9%) | 194 | (19.9%) | 225 | (19.5%) |
| Gender: Female | 426 | (39.5%) | 438 | (44.9%) | 481 | (41.6%) |
| Age at event (in years) | 67.0 | (12.3) | 67.9 | (13.0) | 66.8 | (12.7) |
| NIHSS left arm | 0.7 | (1.3) | 0.7 | (1.3) | 0.7 | (1.4) |
| NIHSS right arm | 0.7 | (1.2) | 0.7 | (1.3) | 0.7 | (1.2) |
| NIHSS level of consciousness | 0.2 | (0.6) | 0.2 | (0.5) | 0.3 | (0.6) |
| NIHSS gaze | 0.3 | (0.6) | 0.3 | (0.6) | 0.3 | (0.6) |
| NIHSS right leg | 0.6 | (1.1) | 0.6 | (1.2) | 0.6 | (1.2) |
| NIHSS left leg | 0.5 | (1.0) | 0.6 | (1.2) | 0.6 | (1.2) |
| NIHSS dysarthria | 0.6 | (0.7) | 0.6 | (0.7) | 0.5 | (0.8) |
| NIHSS extinction and inattention | 0.2 | (0.6) | 0.2 | (0.6) | 0.2 | (0.6) |
| NIHSS total score | 7.5 | (7.8) | 6.9 | (7.0) | 7.0 | (6.9) |

NIHSS: National Institutes of Health Stroke Scale.
\* Complete restitution.

The Barthel Index (BI) ^33^ is a widely used rating scale and measures a person’s daily functioning on a scale from 0 to 100 in steps of 5 points. The first aim was to investigate a model for complete restitution, i.e., functional independence, versus incomplete restitution or mortality. Functional independence was assumed for individuals with BI ≥ 95. The second aim was to investigate a model for mortality versus survival.

We identified possibly predictive variables in a systematic search of the literature, which is available upon request from the corresponding author. To allow for a very early prediction, we used the variables that can be routinely assessed on admission. The following 16 features were employed for model development: age at event (in years), gender (men/women), atrial fibrillation on first electrocardiogram (ECG) at admission, history of stroke, history of myocardial infarction, arterial hypertension, and diabetes mellitus as well as baseline neurologic impairments at admission as rated on the National Institutes of Health Stroke Scale (NIHSS) with single items level of consciousness, gaze, motor right arm, motor left arm, motor right leg, motor left leg, dysarthria, extinction and inattention, plus the NIHSS total score ^20^. Sample sizes were 1156 and 976 for the external and the temporal validation data, respectively. The distribution differs between the model building data and the temporal and external validation data for some features, especially for hypertension and atrial fibrillation.

Missing values were imputed for pragmatic reasons before the analysis in the training data. In detail, for each continuous variable, imputed values were drawn from a normal distribution with the mean and standard deviation calculated from non-missing values. For each categorical variable, imputed values were drawn from a random sample with the probability of each category calculated from non-missing values. There was only one patient with missing data in the each of the validation data, and we dropped these subjects. Information on missing data patterns is provided in Supplementary Material 1, Figures S1 to S3.

For LogReg, we used the regression coefficients from the previously developed regression model ^20^. More information on the regression models and regression coefficients for the final LogReg models are provided in Supplementary Material 1.

### 3.6 Application 2: diagnosis of coronary artery disease—Cleveland Clinic data

In the second application, the objective was to predict coronary artery disease (CAD) using data from the Cleveland Clinic project. While invasive coronary angiography is the reference standard for diagnosing CAD, the study explored non-invasive methods to identify patients who might benefit from undergoing angiography. The dataset included 303 consecutively enrolled patients referred for coronary angiography at the Cleveland Clinic between May 1981 and September 1984. These individuals also underwent non-invasive tests such as exercise electrocardiography, thallium scintigraphy, and cardiac fluoroscopy; for details, see Detrano and colleagues ^34^ (Table 3).

**TABLE 3.**
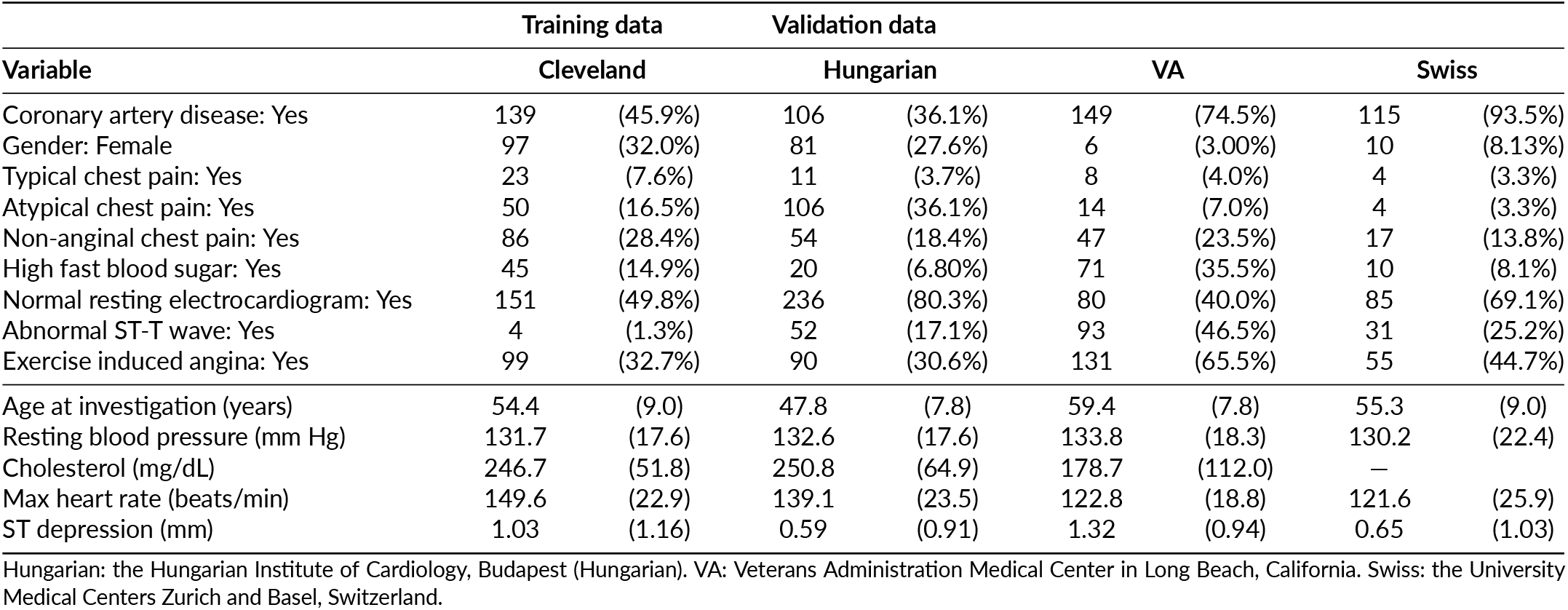
Patient characteristics in the coronary artery disease data. *n* (%) are displayed for dichotomous variables, mean and standard deviation (SD) for continuous variables.

For external validation, data from 617 patients across three other centers were used: the Veterans Administration Medical Center in Long Beach, California (VA); the Hungarian Institute of Cardiology in Budapest (Hungarian); and the University Medical Centers in Zurich and Basel, Switzerland (Swiss). Descriptive statistics for these cohorts are presented in Table 3. We used the same data pre-processing strategy as described in Ojeda and colleagues ^5^.

The outcome of interest was the presence of CAD ^34^, and the features are listed in Supplementary Material 1. Missing values were imputed using the same method as in Application 1. Information on missing data patterns is provided in Supplementary Material 1, Figures S4 to S13. For the LogReg analysis, backward variable selection was performed with a significance threshold of *p* < 0.001 for inclusion. Further details can be found in the R code provided in Supplementary Material 7. The estimated regression coefficients are reported in Supplementary Material 1.

### 3.7 Performance evaluation

Several measures may be used for evaluating the performance of machine learning approaches for probability estimation ^5^. The mean squared error (MSE) was used as performance measure in the simulation studies and estimated by

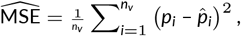

where *p*_*i*_ denotes the true probability and 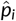 is its calibrated estimator for individual *i*. The theoretical probabilities are only available in simulations, and the Brier score (BS) is its equivalent for real data analysis ^35^. The BS estimator is given by

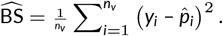

The log loss (LogLoss) was also estimated ^36^. The expected (exp) and observed LogLoss estimators used for the simulated and the real data are given by

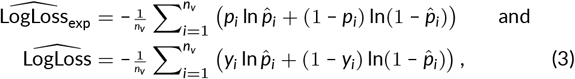

respectively. The BS and LogLoss scores are strictly proper ^36,37^, which means that the scores are minimal if and only if the true probabilities are inserted as estimates. The LogLoss is preferable over the BS because a prediction algorithm that is optimal under the LogLoss function will also be optimal under the BS loss ^36^. The reverse is not necessarily true.

To depict the relationship between the observed and predicted probabilities, we generated calibration plots ^38^. Calibration plots are obtained by regressing the binary outcome on the predicted probabilities using smoothing techniques and plotting the resulting curve. A line showing the angle bisector is displayed for orientation. In the context of a simulation study, where the true probabilities are known, the binary outcome is replaced by the true probabilities. In this case, the underlying pairs of predicted and true probabilities can also be presented in the calibration plot. We used generalized additive model (GAM) estimates as smoother for the calibration plots in the simulation study and locally weighted scatterplot smoothing (LOESS) for the real data. For the simulation study, the results of all replications considered are displayed in a single calibration plot.

To compare calibration approaches across multiple datasets, we follow the non-parametric framework of Demšar ^39^, which may be used for comparing different learning machines by evaluating homogeneous subgroups non-parametrically based on the Nemenyi post-hoc test approach ^39^. Suppose *C* calibration approaches are evaluated on *M* data sets, where each data set may correspond to the simulated data sets for a specific simulation scenario. Let 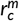 denote the rank of calibration approach *c* on data set *m*. The average rank of calibration approach *c* is then

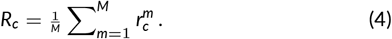

Two calibration approaches are considered to perform significantly differently if their average ranks differ by at least the critical difference (CD)

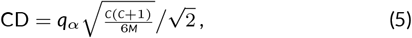

where *qα* is the upper 1 – *α* quantile of the studentized range distribution. Results are visualized using CD plots, which display the average ranks of the machines and/or calibration approaches, with lower average ranks showing better performance ^5^. Homogeneous groups of calibration approaches are represented by bars, and their bar lengths are shorter than the CD. A more detailed description of how to interpret calibration curves, LogLoss boxplots, and CD plots is provided in Figure 1 of Ojeda and colleagues ^5^.

### 3.8 Software packages for estimation, tuning, and calibration

For the simulation studies, we decided to only estimate LogReg models for two reasons. First, LogReg allows a comparison with ACP, which has only been derived for LogReg ^17^ so far. LogReg was fitted using the `glm` function in the `stats` package in the simulation studies and the applications. Second, in our previous work ^5,6^, we comprehensively evaluated the combination of five different machine learning approaches with 10 different calibration functions and observed that the main driver for the performance of the calibration was the performance of the learning machine.

LogReg, gradient boosting (GB), and probability forests, i.e., random forests for probability estimation (RF) were used for the applications. The software packages used for the analysis were identical to those of our previous work ^5^. For GB, the `xgboost` package ^40^ was used to fit boosted trees using the `logloss` loss function. Tuning was done for the hyperparameters depth of the trees, number of trees, learning rate, reduction in the loss function required to split further, proportion of candidate variables sampled at each split, proportion of observations used to fit each tree, and minimum number of observations in a terminal node. The `ranger` ^41^ package was used to grow probability forests. The number of trees was set to 500, and the hyperparameters tuned were the number of candidate predictors sampled at each split and the minimum number of observations in a terminal node.

In the real data analyses, no replicates were available. LogReg coefficients were taken from Weimar and colleagues ^20^ for the stroke data. Bootstrapping with 500 bootstrap replicates was done in all modeling steps of GB and RF for both the training and the validation data. These modeling steps included tuning and calibration. Within each bootstrap step, we reduced cross-validation to two times repeated 2-fold cross-validation for selecting the hyperparameters because of high computational demands. For the estimation of the performance measures of the different calibration approaches in the validation data, 10-fold cross-validation was used to obtain honest performance estimates, and this was within each bootstrap step.

For all learning machines for probability estimation we used the `tidymodels` packages including `parsnip`, `dials`, and `tune`. GB and RF were tuned using a space-filling grid of hyperparameters using the function `grid_latin_hypercube` of size 50.

The `glm` function from the `stats` package was used to implement the LogReg calibration. Critical differences and related plots for comparing the calibration approaches were generated with the help of the `performanceEstimation` package ^42^. The code for ACP was based on the R code provided in the Supplement to Jiang and colleagues ^17^. The code for SBA and EJM was developed in house. The SBA code is provided, e.g., in Supplement 2, file `03-calibration-functions.R`, section `19) SBA-10 calibration`. We do not provide the code for the EJM because EJM is a simple logistic calibration with the expert judgment as the outcome variable and the estimated linear predictor as independent variable, see Eq. (1). All analyses were performed in `Rstudio` with `R` version 4.5.2.

## 4 RESULTS

### 4.1 Simulation study 1: performance of SimCal compared with published alternative approaches

The complete results of the simulation study are provided in Supplementary Material 3b. Figure 3 shows the CD plots for all 8 simulation scenarios. Simulation scenario 5 demonstrates a clear order of the different calibration approaches. The standard calibration approach logistic calibration (Logistic), which requires a calibration data set, outperforms NoCal and the four calibration methods, which do not require a calibration data set. All 6 approaches were significantly different from each other. SimCal performed best among the calibration methods which do not require calibration data. Unexpectedly, ACP performed worse than NoCal. This order of the calibration approaches was identical for simulation scenarios 1, 5, 9, and 16.

**FIGURE 3.**
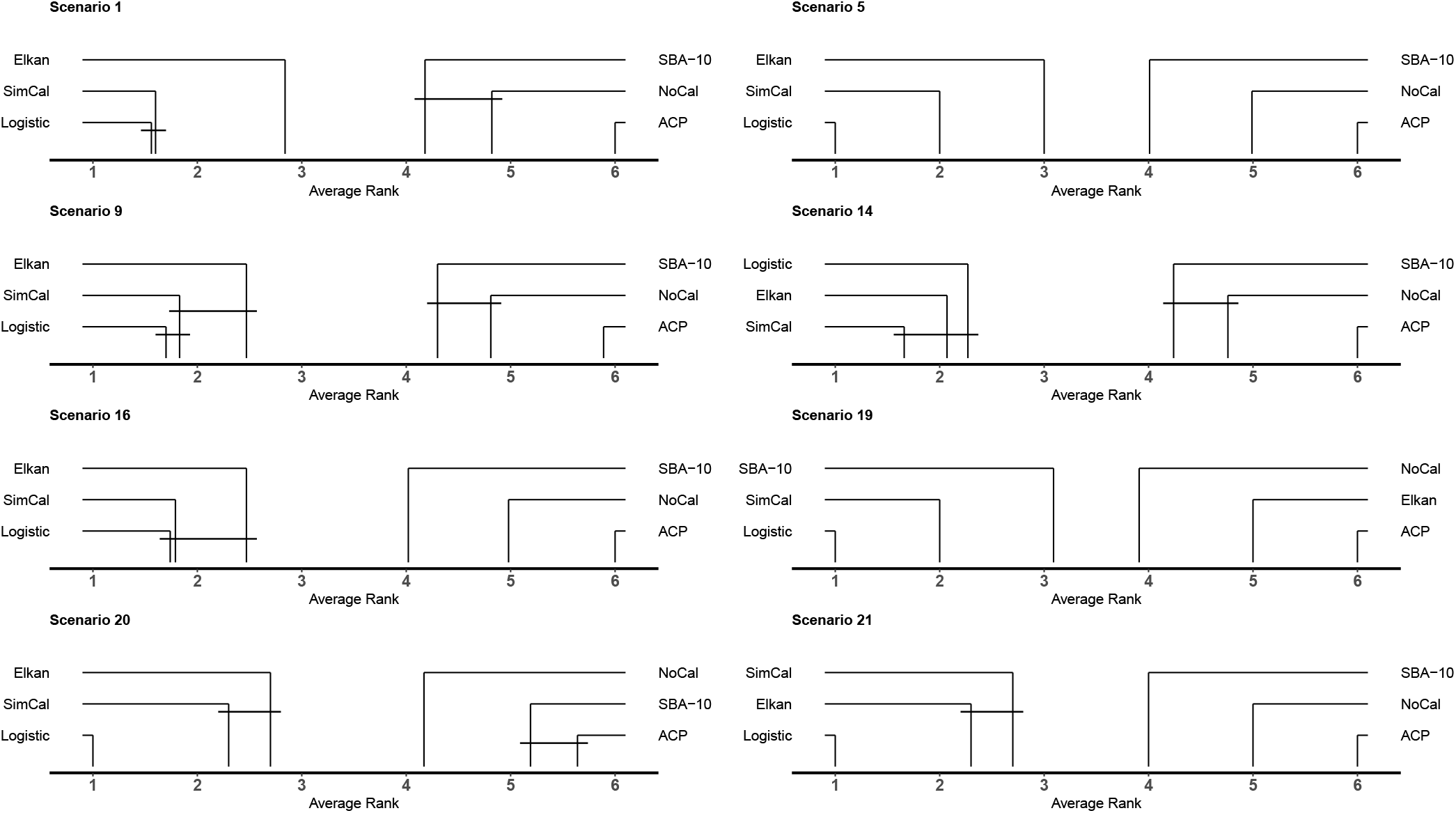
Critical difference (CD) plots for all 8 simulation scenarios. The CD plots are based on the LogLoss. The lower the average rank of the calibration approach, the better the performance of the calibration approach. Groups of calibration approaches connected by a horizontal line segment is lower than the critical difference (CD), which was 0.8 for all models, and they form a homogeneous subgroup. They could thus not be shown to perform significantly differently. The calibration approaches are no calibration (NoCal), logistic calibration (Logistic), similarity binning averaging with 10 nearest neighbors (SBA-10), Elkan calibration, simulation calibration (SimCal), and adaptive calibration of predictions (ACP).

In simulation scenario 14, the cal-training data were smaller, and both SimCal and Elkan calibration performed better than logistic calibration. However, the three approaches were not statistically distinguishable with the performed 100 simulations. Simulation scenario 19 had an unequal covariate distribution with a change from an *N* (1, 1) feature distribution in the training data to an *N* (–1, 2) covariate distribution in the calibration data. This model violates the assumption of identical covariate distribution between training and calibration data underlying Elkan calibration. In consequence, Elkan calibration performed worse than NoCal in this simulation scenario, but Elkan calibration surprisingly was still better than ACP. In simulation scenario 20, where the magnitude of the effect changed for the regression coefficient from *β* = 1 to *β* = 3, SBA-10 performed significantly worse than NoCal.

The final simulation scenario 21 is the most challenging one. Here, the regression coefficient changed from a *β* = 1 in the training data to *β* = –1 in the calibration. Only logistic calibration was able to capture this change from a risk factor to a protective factor in the association between the feature and the outcome. All calibration approaches that operated without calibration data performed poorly (Figure 4).

**FIGURE 4.**
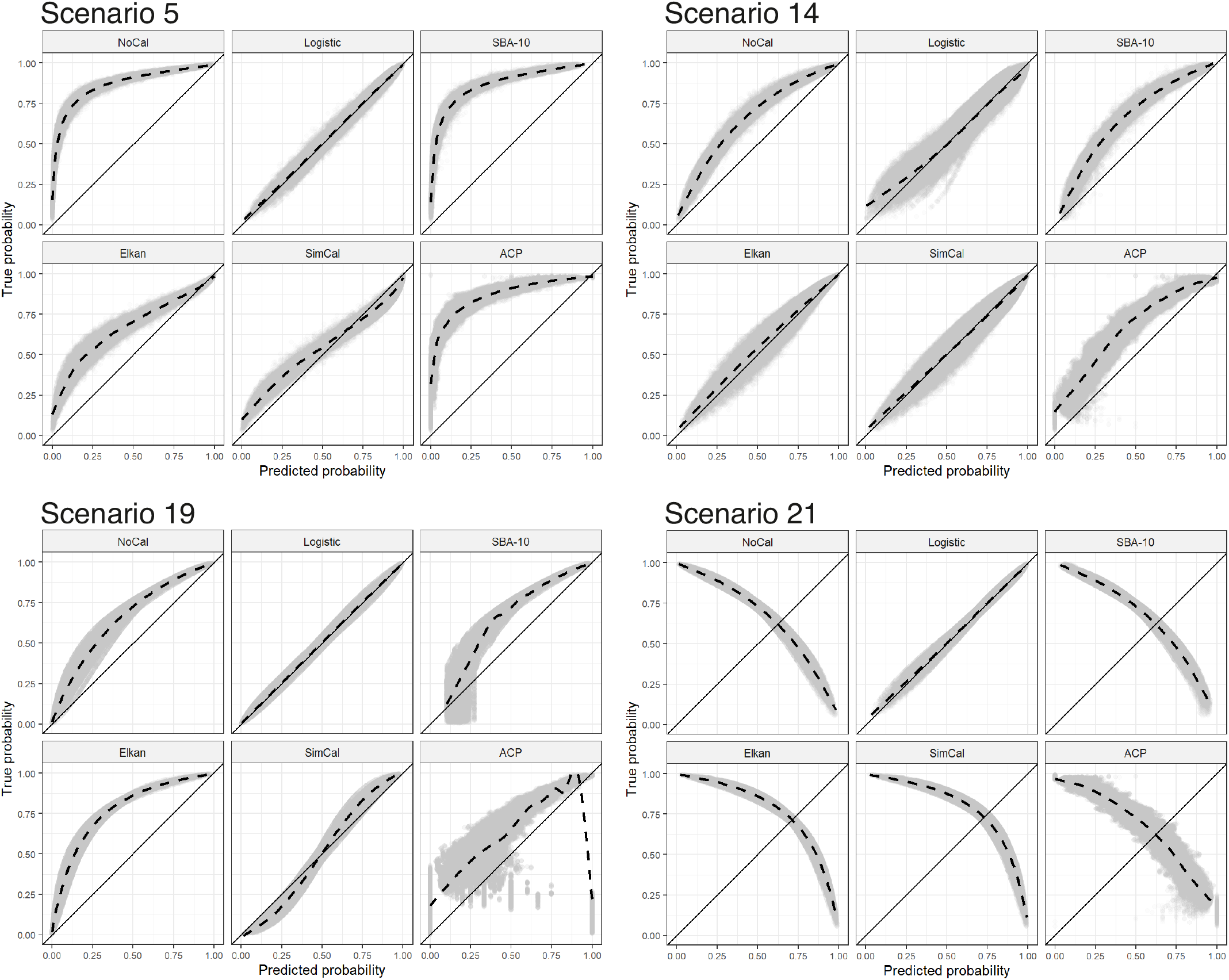
Calibration plots for simulation scenarios 5, 14, 19, and 21. The calibration plots show true versus predicted probabilities. A solid line showing the angle bisector is displayed for orientation, and a dashed line provides the generalized additive model (GAM) estimates as smoother in the scatterplot. The grey shaded areas are the scatterplots of all 100 simulation cases. In all scenarios, the following calibration approaches are displayed: no calibration (NoCal), logistic calibration (Logistic), similarity binning averaging with 10 nearest neighbors (SBA-10), Elkan calibration (Elkan), simulation calibration (SimCal), and adaptive calibration of predictions (ACP). Only logistic calibration uses the cal-training data. The other approaches do not utilize the cal-training data and are just based on the cal-test data.

Figure 4 shows the calibration plots for simulation scenarios 5, 14, 19, and 21. The calibration plots for simulation scenario 5 reveal that logistic calibration was almost identical to the angle bisector. The calibration curve for SimCal was slightly curved, thus SimCal was inferior to logistic calibration in this simulation scenario. The Elkan calibration curve appeared to be closer to the angle bisector in simulation scenario 5 than NoCal, SBA-10, and ACP.

Simulation scenario 14 shows excellent fits of the calibration curves to the angle bisector for SimCal and Elkan calibration. This is the simulation scenario with a smaller cal-training data set, where the curve for logistic calibration exhibits some deviation from the ideal line for lower predicted probabilities. NoCal, SBA-10, and ACP showed a large deviation from the angle bisector. In simulation scenarios 19 and 21, logistic calibration showed a perfect fit. ACP revealed an unusually bad fit in simulation scenario 19.

### 4.2 Simulation study 2: performance of SimCal with misspecified mean

Figure 5 displays the boxplot of the LogLoss for simulation study 2. Basis for this simulation study was simulation model 16, which has a single feature distributed as *N* (1, 1). The result for NoCal is shown before the correctly specified SimCal model, denoted as “true mean” in the figure. The mean misspecification started at 1.1, representing a misspecification of 0.1, and ended at 2.0, corresponding to a misspecification of 1.0, i.e., a coefficient of variation of 1.0. As expected, the LogLoss increased with an increasing level of misspecification, and the increase had an almost linear pattern. Detailed results from simulation study 2 are provided in Supplementary Material 4.b.

**FIGURE 5.**
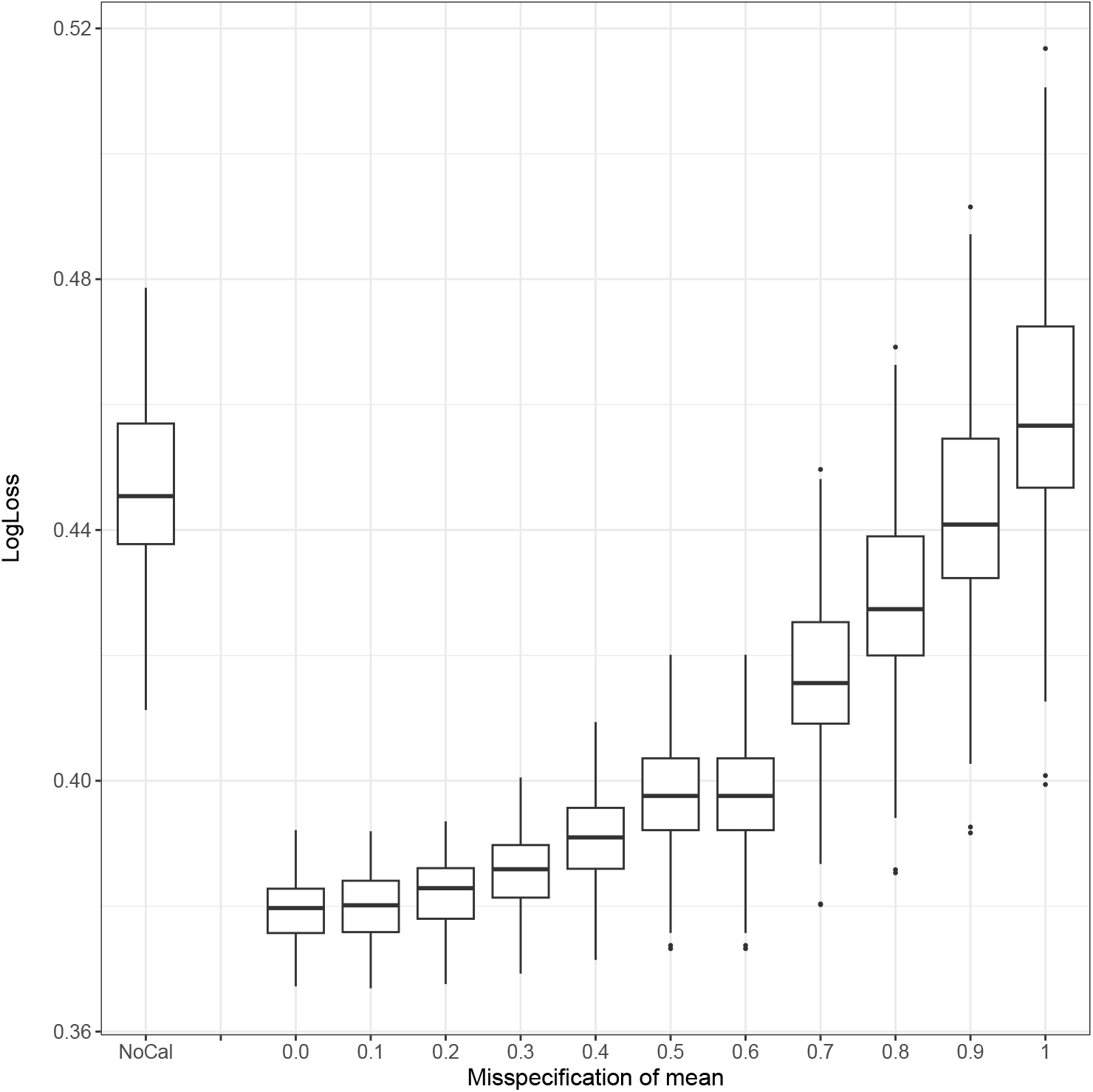
Boxplots for simulation study 2, which investigated the effect of mean misspecification for SimCal. The basis for the simulation study was simulation model 16, which had a true distribution of the feature of *N* (1, 1). The first boxplot shows the LogLoss for no calibration (NoCal). All subsequent boxplots display the Logloss when the mean of 1 was correctly specified (true mean), i.e., when there was 0.0 misspecification. The misspecification of the mean was increased to 1 in steps of 0.1, i.e., the assumed mean ranged from 1 to 2.

### 4.3 Simulation study 3: performance of SimCal with misspecified outcome probability

Figure 6 displays the boxplots of the LogLoss for simulation study 3, where the effect of a misspecified outcome probability was investigated. Results for NoCal are shown before the misspecified SimCal model. The misspecification of 0.00 corresponds to the correctly specified model, i.e., the true outcome probability of 0.84. The performance of the calibrated model deteriorated with the increase of the misspecification. The figure also shows that the effect of the misspecification was not symmetric. Specifically, misspecification had a more serious effect when it was closer to the tail of the probability distribution. Detailed results from simulation study 3 are provided in Supplementary Material 5.b.

**FIGURE 6.**
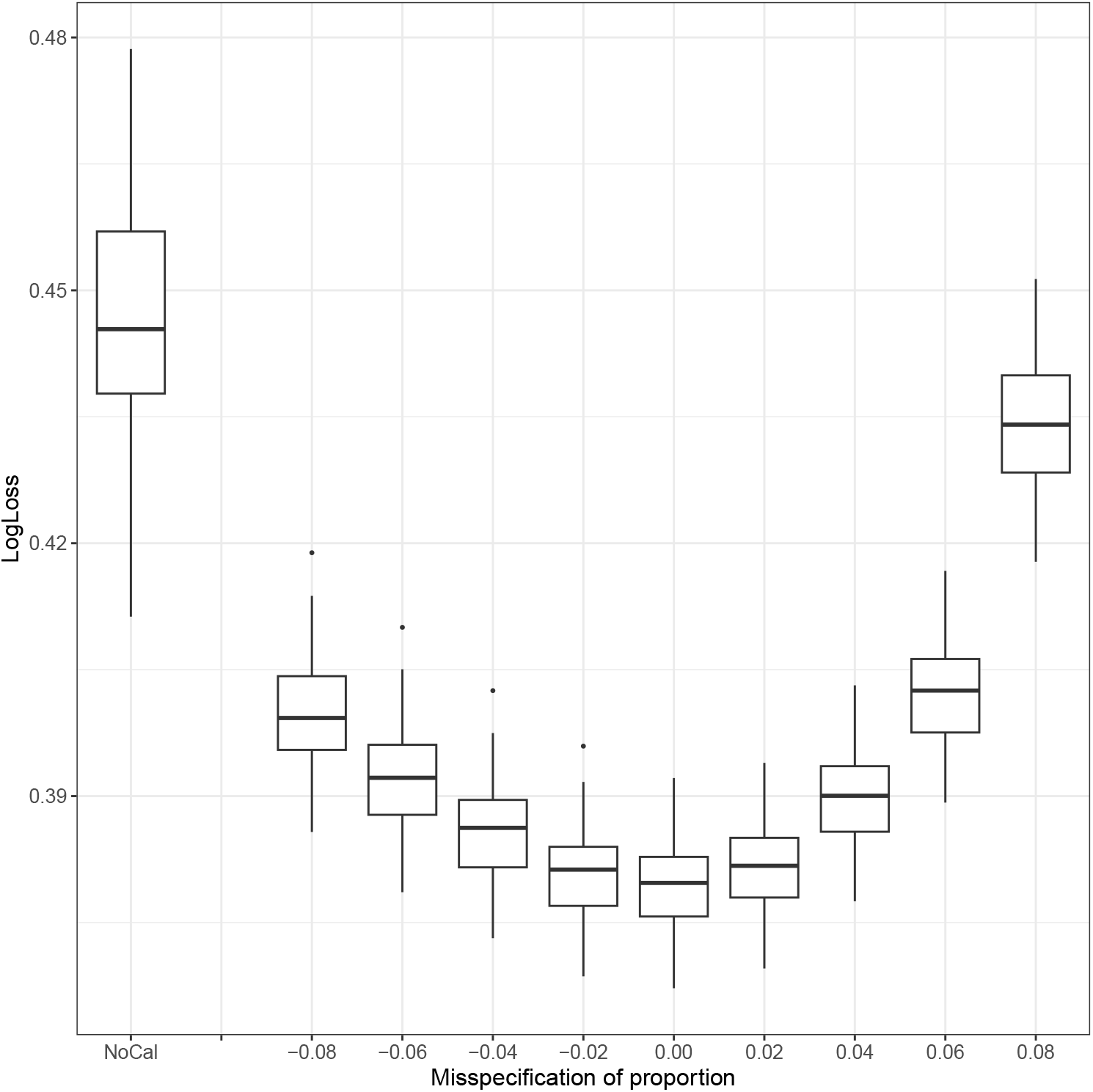
Boxplots for simulation study 3, which investigated the effect of outcome probability misspecification for SimCal. The basis for the simulation study was simulation model 16, which had a true outcome probability of 0.84. When the assumed outcome probability was 0.84, the misspecification was 0. For sake of comparison, the first boxplot shows the calibration plot for no calibration (NoCal). The next boxplots show the calibration plots for varying outcome probabilities. Outcome probabilities were varied from 0.76 to 0.92 in steps of 0.02. This corresponds to misspecifications from −0.08 to 0.08. The correctly specified model had an outcome probability of 0.84.

### 4.4 Simulation study 4: performance of SimCal with misspecified mean and misspecified outcome probability

Figure 7 shows the boxplots of the LogLoss for simulation study 4, where the effect of jointly misspecifying the mean and the outcome probability were investigated. The results were surprising. One would expect an increase of the LogLoss when the misspecification of both the mean and the outcome probability increase. This expectation was confirmed in one case, namely when the mean was overestimated and the outcome probability was underestimated. However, if both the mean and the outcome probability were overestimated or when both were underestimated, there was a compensation effect. In fact, the LogLoss could be as low as if there was no misspecification, i.e., neither a mis-specification of the mean nor of the outcome probability. This effect is most pronounced in the two right most figures, which correspond to mean misspecifications of +1 (upper part) and −1 (lower part). In the upper part, the boxplot with the lowest LogLoss was at an outcome mis-specification of 0.08. In the lower part, the LogLoss was lowest at an outcome misspecification of −0.16.

**FIGURE 7.**
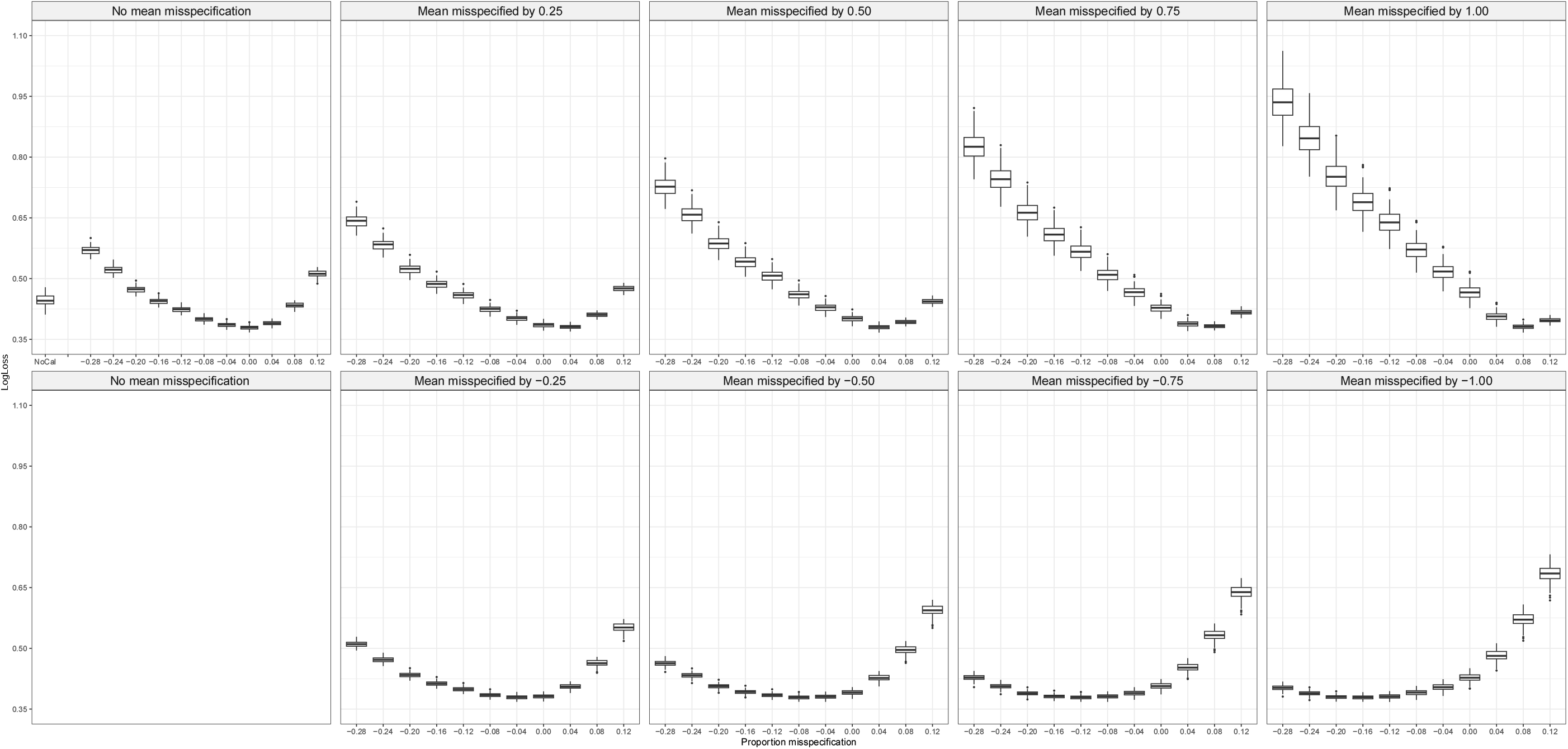
Boxplots for simulation study 4, which investigated the effect of misspecifying both the mean and the outcome probability for SimCal. The basis for the simulation study was simulation model 16, which had a true distribution of the feature of *N* (1, 1) and an outcome proportion of 0.84 in the true calibration data. The first boxplot shows the LogLoss for no calibration (NoCal). All subsequent boxplots display the Logloss for varying assumed means, with a variation of the misspecification from −1.00 to +1.00 in steps of 0.25, and assumed outcome proportions, specifically with a variation of the misspecification from −0.28 to 0.12 in steps of 0.04. The model with a misspecified mean of 0.00 and a misspecified outcome probability of 0.00 corresponds to the correctly specified model (true model).

The question is how this compensation effect may be explained. Equation (3) provides the answer, in our opinion. The LogLoss is lowest when 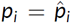. If *p*_*i*_ increases due to a misspecification of the outcome probability, the estimated probability 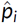 should also increase to minimize the LogLoss. This naturally occurs if the mean is overestimated. Similarly, if the outcome probability is underestimated, the mean should be under-estimated as well to minimize the LogLoss. However, we are not aware of the specific functional relationship between the misspecification of both parameters which lead to the minimum of the LogLoss.

Detailed results from simulation study 4 are provided in Supplementary Material 6.b.

### 4.5 Application 1: prediction of functional outcome after stroke

Boxplots for the calibrated learning machines using the LogLoss as performance measure are shown in Figure 8 for both the complete restitution model (CRM; upper panel) and the mortality model (MM; lower panel) of the stroke data for the temporal validation data (red) and the external validation data (yellow). The refit and the logistic calibration are displayed as comparators to the calibration models that do not require calibration data. Since clinical judgments by the treating physician were available for both the temporal and the external validation data, EJM could be performed and are displayed. Critical difference plots are shown in Figure 9 for the average across the machines LogReg, RF, and GB. Detailed figures are provided in Supplementary Material 1.

**FIGURE 8.**
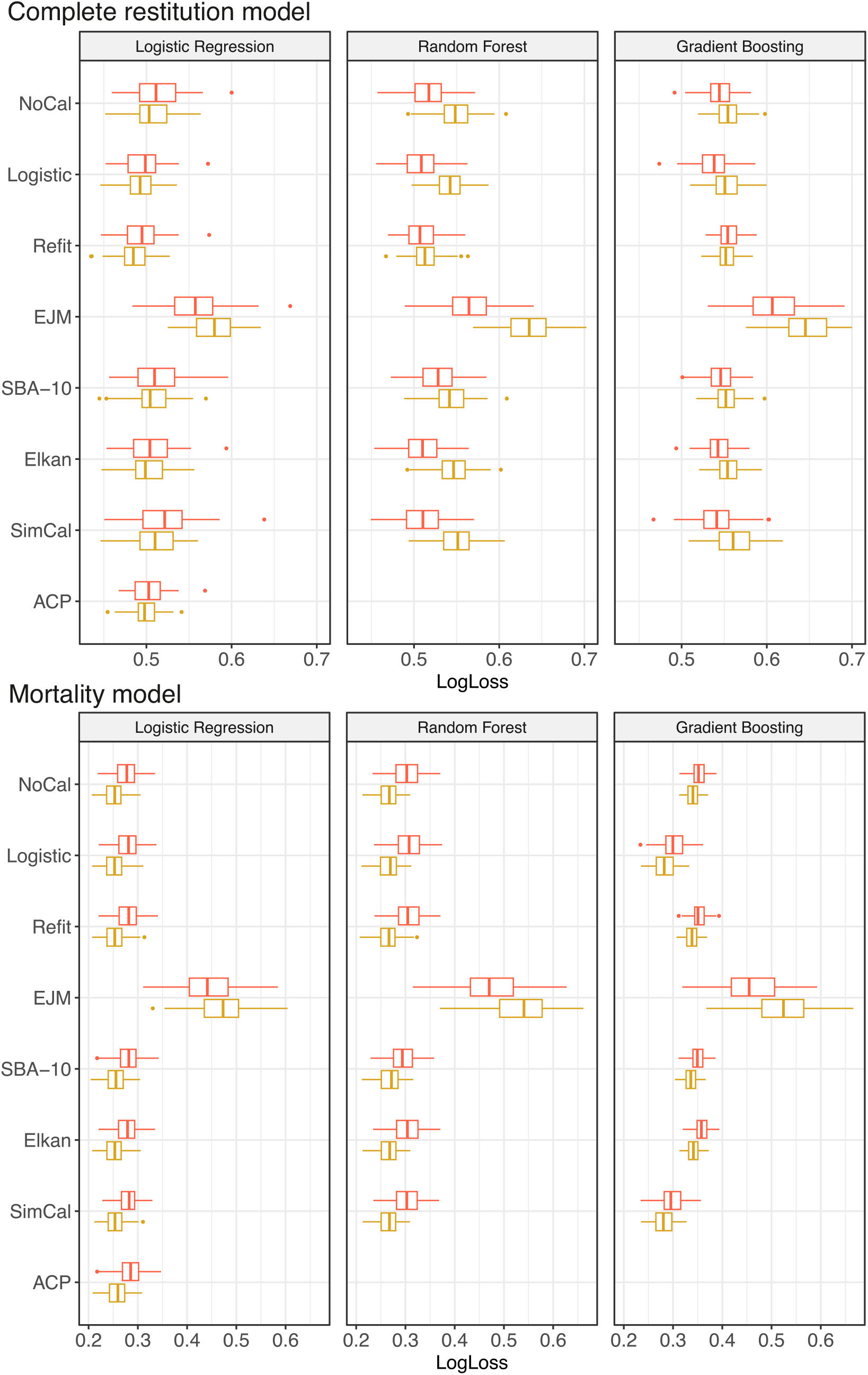
Boxplots for calibrated learning machines for probability estimation in the complete restitution model (upper panel) and the mortality model (lower panel) of the stroke data using the LogLoss (*x*-axis) as performance measure for logistic regression, random forest, and gradient boosting for both the temporal validation data (red) and the external validation data (yellow). Adaptive calibration of predictions (ACP) can only be applied to LogReg. EJM: expert judgment modeling. NoCal: no calibration. SBA-10: similarity binning averaging with 10 nearest neighbors. SimCal: simulated calibration.

**FIGURE 9.**
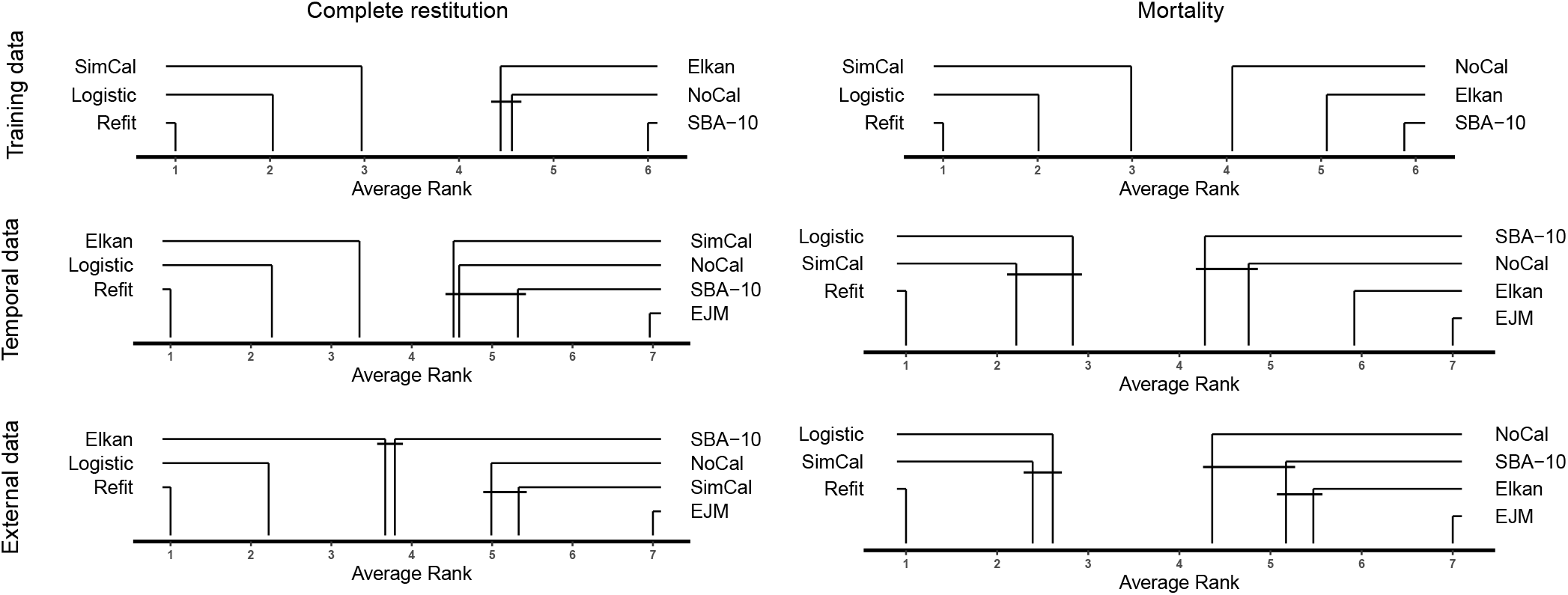
Critical difference plot for the stroke data averaged over all learning machines for probability estimation for the training, temporal validation, and external validation data. Lower average ranks are considered better. Groups of calibration approaches connected by a horizontal line segment could not be shown to have a significantly different performance. The critical difference was 0.9.

Over all models (Figure 9), EJM performed worst, and refit performed best. The averaged CD plots show that Elkan calibration performed well in the complete restitution model. SimCal was the best calibration approach not requiring calibration data for the mortality model. SBA-10 did not perform well on these imbalanced data. A closer look at the performance of the individual machines (Figure 8) shows that SBA-10 performed especially well for GB (Supplementary Material 1, Figure S16). ACP showed a low LogLoss for the complete restitution model, but a worse performance on the mortality model (Supplementary Material 1, Figure S14).

Results differed for the calibration plots (Figures 10 and 11). The largest deviation from the angle bisector in the complete restitution model was observed for EJM. Furthermore, the CIs were unusually shaped for EJM in the mortality model. The reason for these unusually shaped CIs may lie in the few experts who predicted that the patients would die during the observational period. In fact, the experts predicted that only 16 of 892 patients and 10 of 1031 patients across the temporal and external validation data sets, respectively, would die within the follow-up period. One reviewer noted that NoCal was ranked best for the mortality data with LogReg (Supplementary Figure S14, panels C) and D)). Similarly, it was intriguing for the reviewer to see that refit ranked worse than any other approach for the mortality data. We stress that the mortality model is difficult to fit given the imbalance of the outcome variable; the ratio is approximately 1:9 for mortality:survival. Moreover, the histogram of LogLoss difference for the mortality stroke data based on the LogReg model shows that the difference in LogLoss between Refit and NoCal is negligible (Supplementary Figure S30), and they varied between −0.001 and 0.009 across the 100 bootstrap replicates for these models. Finally, the calibration plots for the training data from the LogReg model appear to deviate substantially from the angle bisector for predicted probabilities larger than 50% (blue dotted line and blue shaded area). The data are scarce in this area as can be seen from the wide CIs.

**FIGURE 10.**
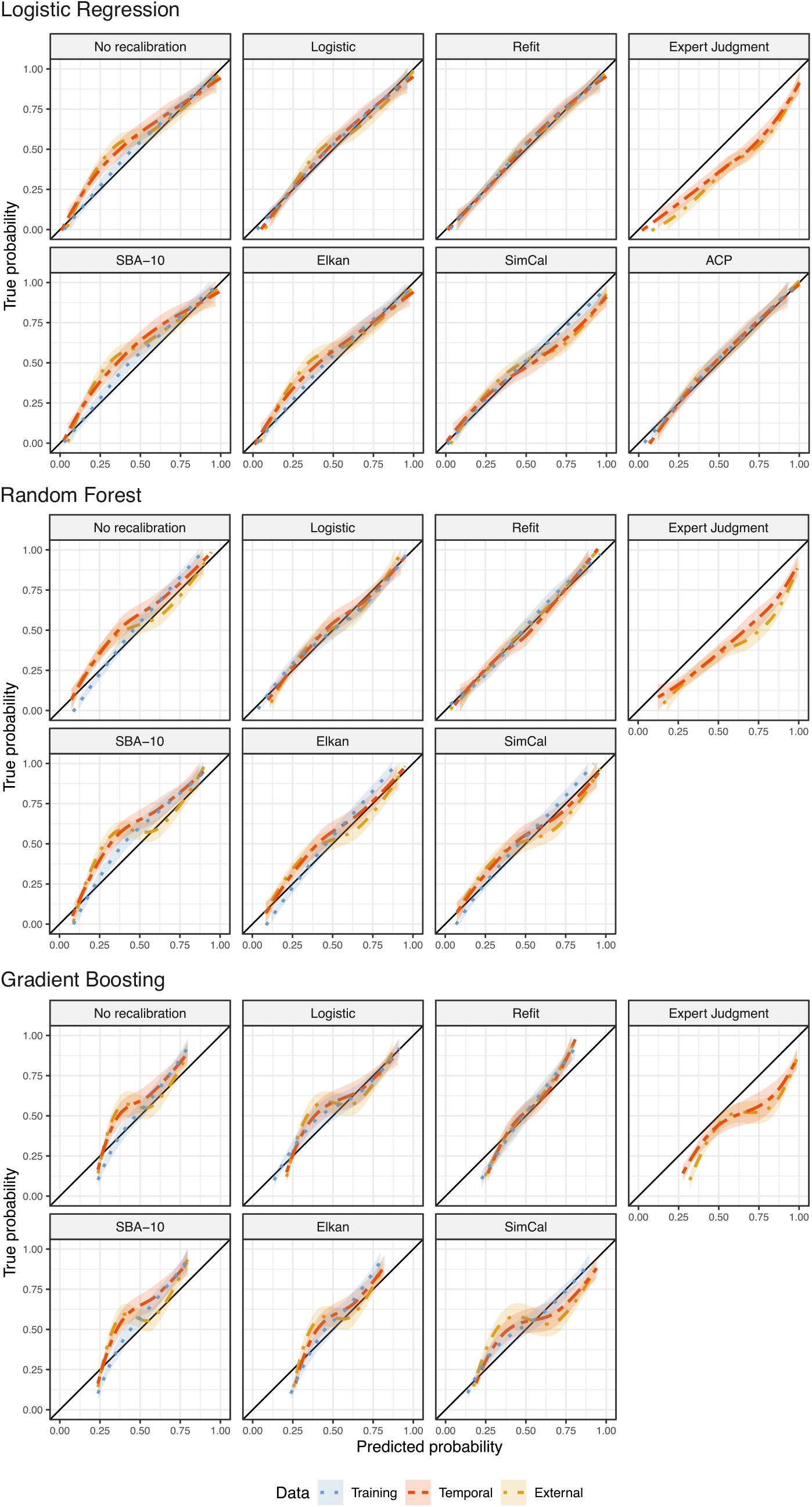
Calibration plots with confidence intervals for calibrated logistic regression analysis, random forests, and gradient boosting in the complete restitution model of the stroke data.

**FIGURE 11.**
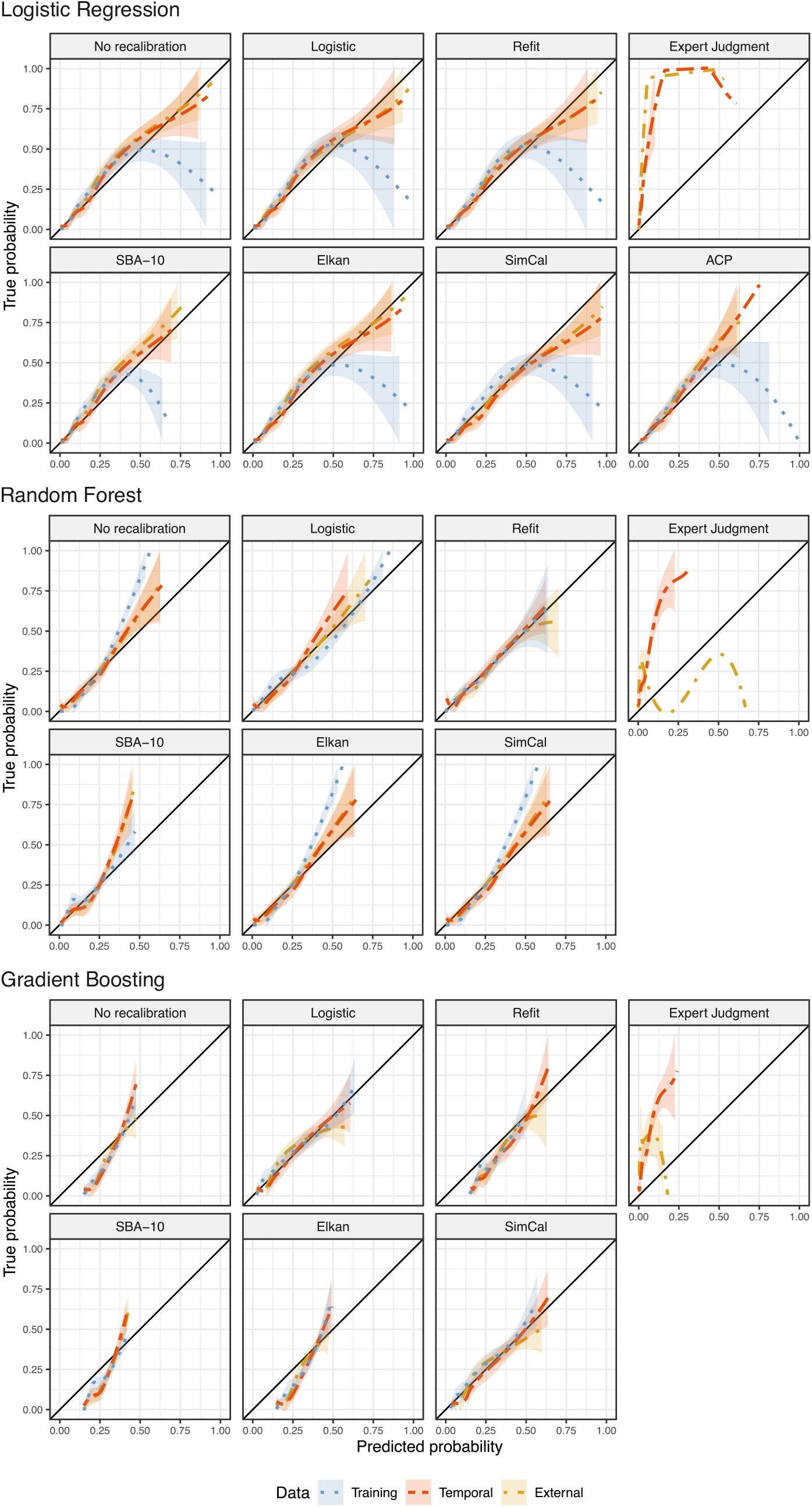
Calibration plots with confidence intervals (CIs) for calibrated logistic regression analysis, random forests, and gradient boosting in the mortality model of the stroke data. Single observations lead to the failure of CIs for expert judgment modeling and adaptive calibration of predictions.

Both Elkan and SBA-10 revealed major deviations from the angle bisector for the complete restitution data, especially for the LogReg model. In contrast, SimCal appeared to be close to the angle bisector. According to the calibration plots, we expected a better performance of SimCal compared to SBA-10 (Figure 10). The difference between SimCal and Elkan and between SimCal and SBA-10 for the LogLoss results and the calibration plots in the complete restitution model can be explained by the scatterplot with frequency weighting (Figures 12.a and 12.b). They illustrate the different performance of the machines in combination with the calibration methods, where the predicted probabilities for SimCal (y-axis) are plotted against predicted probabilities in panel A) for Elkan calibration (x-axis), in panel B) for SBA-10, in panel C) for logistic calibration, and in panel D) for the refit. The left plot shows the results for LogReg, the right plot for GB; the upper row provides the results for the complete restitution model, and the lower row the results for the mortality model. Both Elkan and SBA-10 showed good agreement with the angle bisector in the calibration plot for low predicted probabilities of approximately 0.25. However, most observations from the stroke data had calibrated predicted probabilities in the range between 0.18 and 0.30 (Figure 12.a).

**FIGURE 12.**
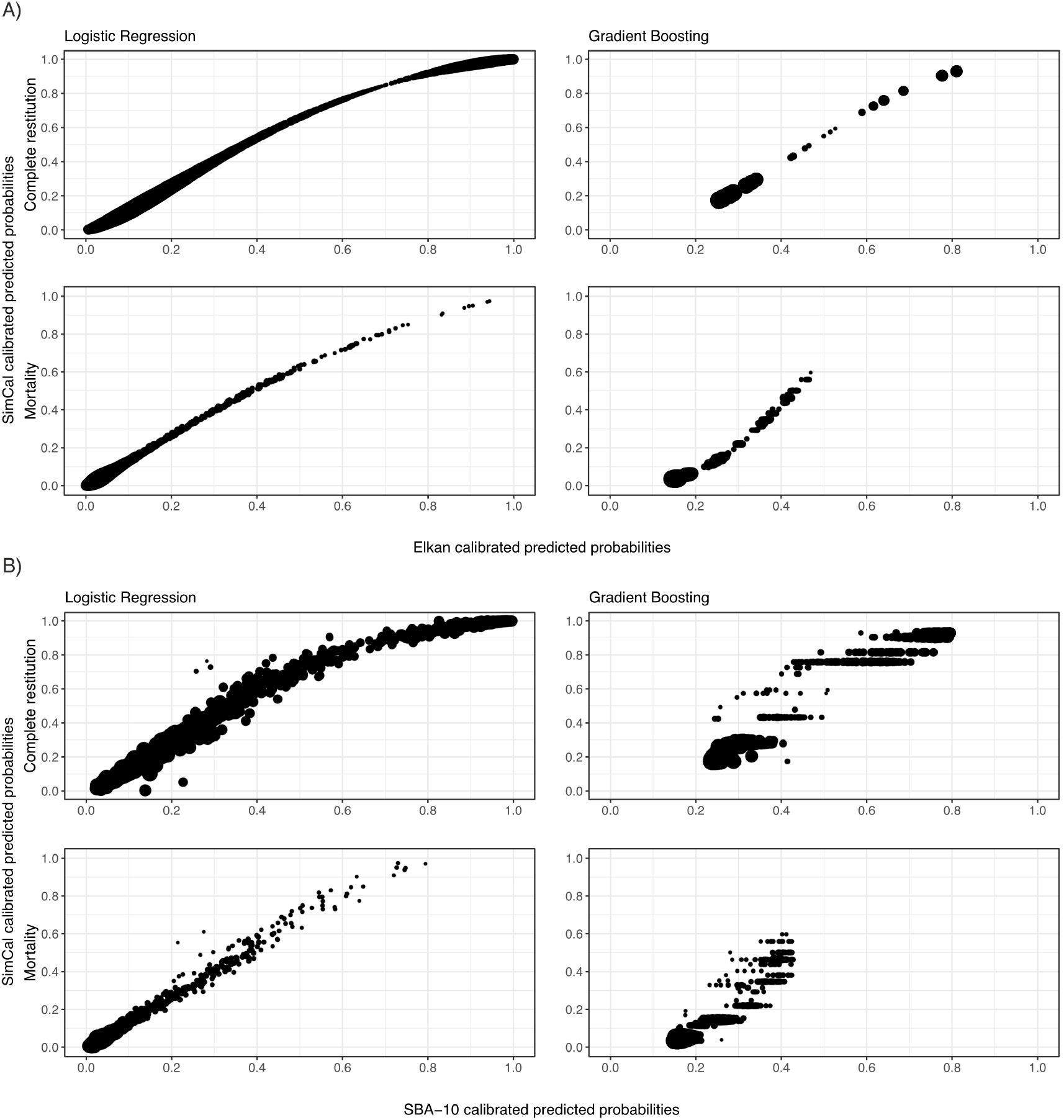
**a** Scatterplot for calibrated probabilities from logistic regression and gradient boosting for the complete restitution model and the mortality model in the external validation data for pairs of calibration methods. Panel A): simulated calibration (SimCal) versus Elkan calibration, panel B): SimCal versus SBA-10.

**FIGURE 12.b.**
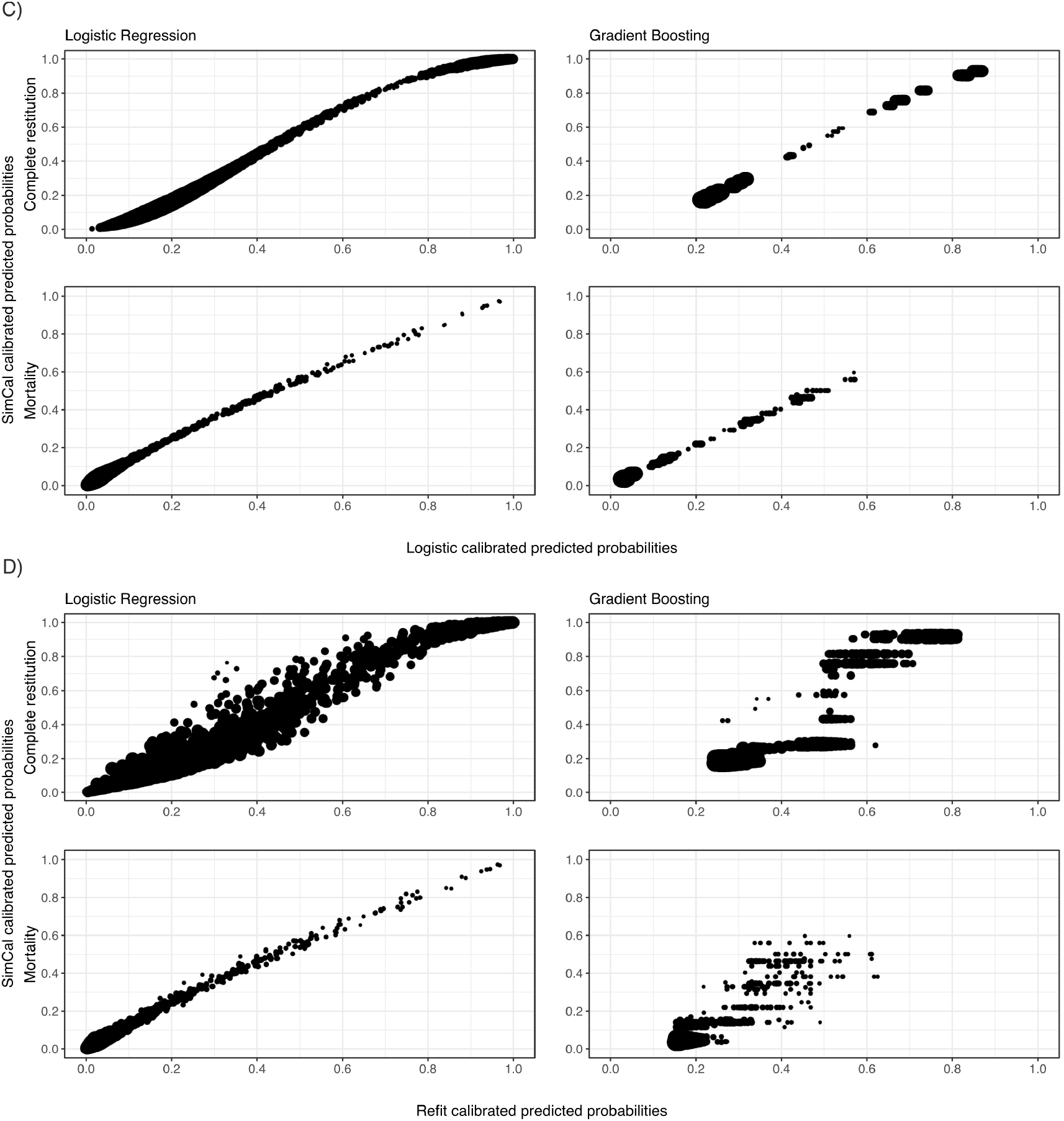
Scatterplot for calibrated probabilities from logistic regression and gradient boosting for the complete restitution model and the mortality model in the external validation data for pairs of calibration methods. Panel C): simulated calibration (SimCal) versus logistic regression, panel D): SimCal versus refit.

The most pronounced difference is observed between LogReg and GB, not between the calibration methods. While LogReg yielded estimates across almost the entire probability range, GB was restricted to a small spectrum of probabilities. Specifically, probability estimates varied only between > 10% and < 50% for Elkan calibrated predicted probabilities in the GB mortality model. Calibration methods yielded more homogeneous results. For example, SimCal varied between 0% and 60% in the GB mortality model. These probabilities indicate that the calibration methods still have an effect on the estimated probabilities, which is also recognized by a clear deviation of the plots from the angle bisector, but that is not displayed in this figure. Since the outcome data are unbalanced in the mortality model with an approximate 1:9 ratio, calibration is more challenging on these data. This is probably due to the larger variability in the mortality model, thus reduced model stability. The calibration plots reflect this observation (Figure 11), where single observations lead to the failure of CIs for EJM and ACP.

### 4.6 Application 2: diagnosis of coronary artery disease

Results varied substantially between the different machines for the Cleveland Clinic data. For example, the refit had a substantially lower LogLoss for the RF than for GB (Figure 13). Moreover, logistic calibration performed slightly better than the refit for GB, but refit substantially outperformed logistic calibration for the RF.

**FIGURE 13.**
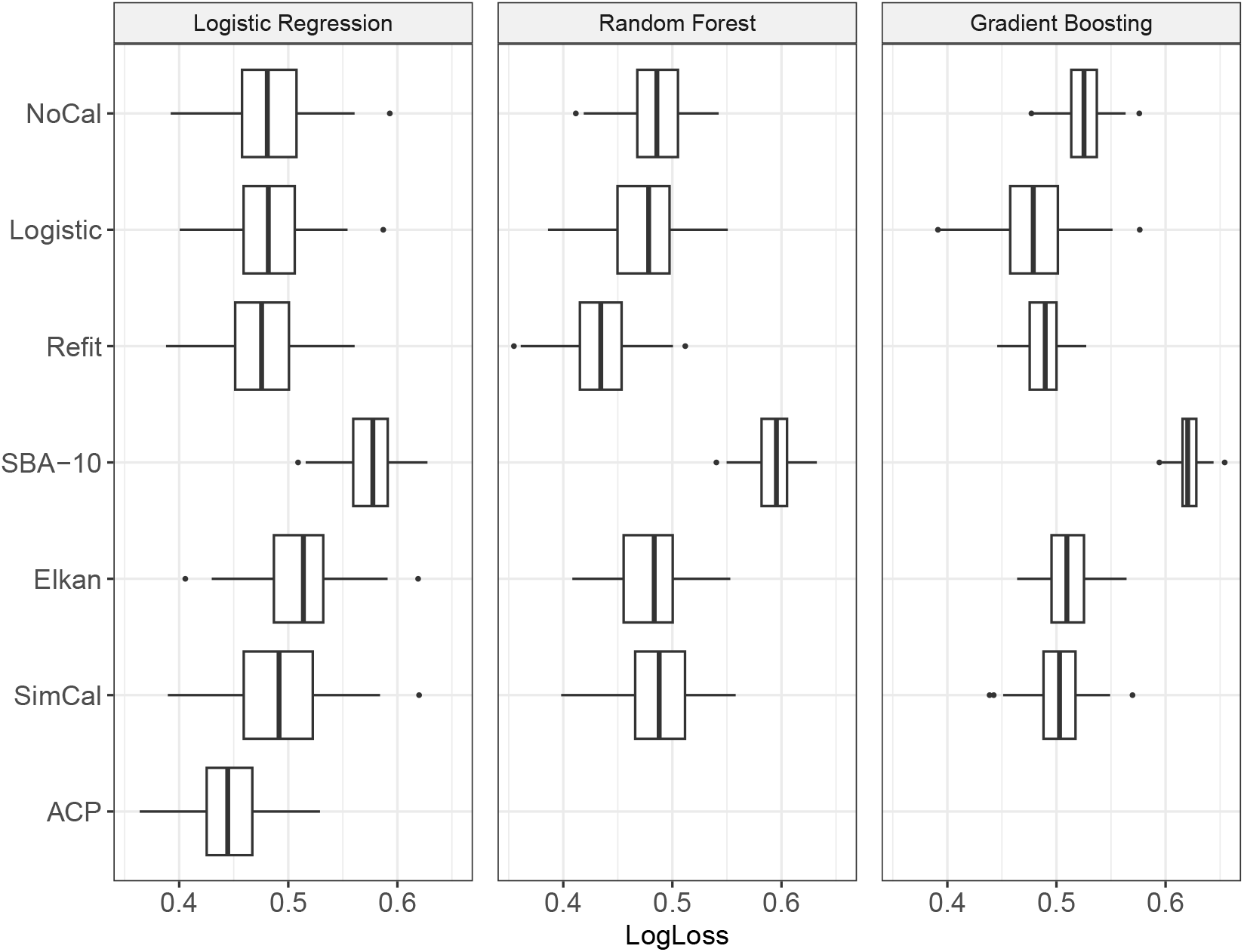
Boxplots for calibrated learning machines for probability estimation in the Cleveland Clinic validation data using the LogLoss (*x*axis) as performance measure.

Over all three machines, refit performed best on the Cleveland Clinic data, followed by logistic calibration (Figure 14). SimCal showed the best performance among all calibration approaches without calibration data. However, SimCal was not significantly better than NoCal or Elkan calibration (Figure 14). SBA-10 performed worst on this dataset for all three machines.

**FIGURE 14.**
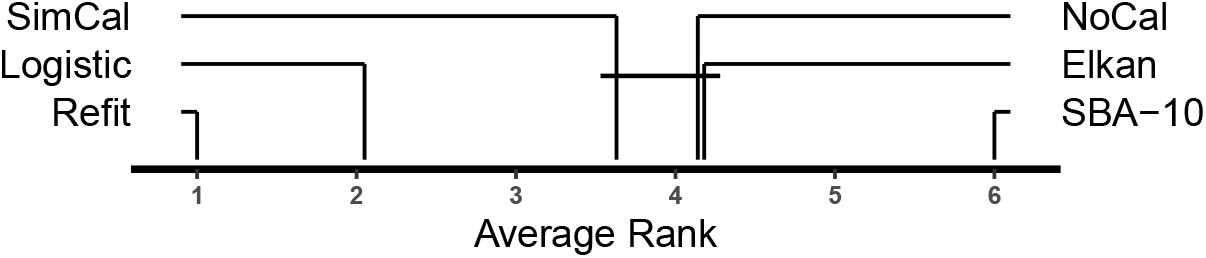
Critical difference plot for the LogLoss in the Cleveland Clinic validation data for each learning machine for probability estimation, averaged over all three machines. Lower average ranks are considered better. Groups of calibration approaches connected by a horizontal line segment could not be shown to have a significantly different performance. The critical difference was 0.8.

The boxplots of the LogLoss show that ACP performed best on the Cleveland Clinic data for the LogReg model (Figure 13). In the LogReg model, NoCal had a slightly lower LogLoss than logistic calibration and SimCal. Elkan calibration and SBA-10 showed the highest LogLoss for LogReg on the Cleveland Clinic data. In contrast, Elkan calibration performed slightly better than SimCal for the RF, but worse than SimCal for GB.

## 5 DISCUSSION

Calibration of probability estimates is possible even in the absence of calibration data. Several calibration approaches outperformed NoCal in the limited set of simulation studies used in this work and in the analysis of two real data sets. As expected, logistic calibration, which requires the availability of calibration data outperformed all the calibration approaches that can be used in the absence of calibration data. The largest difference between logistic calibration and calibration approaches that do not require calibration data was observed in simulation scenario 21, where the sign of the regression coefficients changed from the training population to the calibration population. In this model, a risk factor became a protective factor and all calibration approaches without calibration data failed. In contrast, SimCal and Elkan calibration outperformed logistic calibration when only a small calibration dataset was available (Figure 3, simulation scenario 14).

Overall, SimCal and Elkan calibration performed well with SimCal performing better than Elkan calibration in both the simulations and the real data analysis. The most striking difference between SimCal and Elkan calibration could be observed in simulation scenario 19, where the covariate distribution differed between the two populations. This simulation scenario explicitly violates the assumption of Elkan calibration of equal covariate distributions for both populations ^5,18^, and Elkan calibration performed even worse than NoCal in this setting.

ACP ^17^ performed worse than NoCal in all simulation scenarios, and SBA-10 performed worse than NoCal in simulation scenario 20. The poor performance of both methods can be attributed to their reliance on training data from population 1 to calibrate new observations from population 2: when structural differences exist between populations, this leads to biased probability estimates. The same conceptual weakness applies to Platt’s (2000) approach, which proposed cross-validation or sample splitting to estimate the calibration function. ACP carries the additional drawback of requiring a confidence interval around each predicted probability, restricting it to specific machines and, so far, limiting its applicability to LogReg. However, the general goal is to develop calibration methods applicable to all learning machines ^29^. As noted by a reviewer, however, approaches such as ACP could in principle be combined with Elkan calibration to form a novel calibration method. We also note that SBA-10 can be used when calibration data are available ^7^, in which case the nearest neighbors are drawn from the calibration data rather than the training data.

SimCal outperformed the other calibration approaches in simulation study 1, where the SimCal parameters were assumed to be correctly specified. Simulation studies 2 to 4 showed that SimCal is robust against some level of misspecification of the features and of the outcome probabilities. When the misspecification was moderate, SimCal still performed better than NoCal. In simulation study 2, NoCal was superior only in case of an extreme misspecification of *µ*/*σ* ≥ 0.8, and in simulation study 3, SimCal performed better than NoCal across all considered scenarios.

Simulation study 4 investigated the simplest effect of multiple simultaneous misspecifications. When the misspecifications of the mean and the outcome probability were in opposing directions, i.e., a lower mean was assumed than the true one, while the assumed outcome probability was higher than the true outcome probability, the performance of SimCal deteriorated rapidly. In contrast, misspecification in the same direction yielded a compensation effect. This is in line with the property of the LogLoss, which is minimal when the estimated and the true outcome probabilities coincide. A limitation of the performed simulation studies is that a misspecification of the covariance matrix was not systematically evaluated; however, a real data analysis using previously published data ^43^ demonstrated good robustness (data not shown).

In this work, the simulation of the large population for SimCal was based on the R package `modgo` ^24^ for sake of convenience. Other packages, such as `sbgcop` ^23^ or `SimMultiCorrData` ^44^ could have been chosen as well. The simulation approach implemented in `modgo` has two limitations. First, `modgo` cannot model non-linearities in continuous covariates, and it needs special configurations for handling continuous covariates with heterogeneous variances. Specifically, we have not examined how non-linearities or heteroscedasticity affects the performance of SimCal. We speculate this effect to be less relevant than in the development of machine learning models, and this aspect should be investigated in future studies. Moreover, we would like to stress that the main aim of our work was to describe the different approaches that may be used for calibration in the absence of calibration data. Specifically, we wanted to illustrate the novel SimCal approach and the use of EJM in the context of calibration. Future work should evaluate the performance of the learning machines in conjunction with additional, substantially more complex simulation scenarios, in which multiple misspecifications of SimCal are addressed.

Another limitation of our work is that we did not fully explore SBA. The availability of the number of nearest neighbors as a tuning parameter makes a fair comparison of this machine learning approach with other methods challenging ^45^. We therefore chose to work with the original algorithm suggested by Bella and colleagues ^7^, which fixed the number of nearest neighbors to 10. These authors did not evaluate the performance of other numbers of nearest neighbors, and we are not aware of any other work that has investigated this question. Future work should evaluate how to select the tuning parameter, and the optimal number might be determined as described by Kruppa and colleagues ^46^.

Findings were surprisingly heterogeneous in both real data applications. For example, Elkan calibration performed best for the complete restitution model of the stroke data when averaged over all machines. However, it performed worse than NoCal for the mortality model. The most striking finding for the Cleveland Clinic data was that the choice of the machine learning model showed a substantially larger effect than the choice of calibration method (Figure 13). It is obvious that the calibration cannot perform well if the learning machine itself performs poorly.

The application of EJM to the stroke data led to a bad performance. The calibration curves for the complete restitution model were far away from the angle bisector (Figure 10), and the shape of the calibration curves was far from a smooth function in the mortality model (Figure 11). Our results are in contrast with the benchmark study by Armstrong ^14^ who applied EJM to 11 studies from various application areas. He showed using studies from psychology, education, personnel, marketing, and finance that bootstrapping forecasts were more accurate than forecasts made by experts using unaided judgment. Specifically, higher accuracy of EJM over unaided judgment was obtained for 8 of the studies, and EJM was less accurate in only one study. However, our previous analyses ^31^ showed that the originally developed LogReg models performed substantially better than the treating physicians’ predictions made within 6 hours after admission.

A disadvantage of EJM is that its performance, unfortunately, cannot be studied by simulations. Further, we are not aware of additional freely available real datasets that allow to expand the benchmark of Armstrong on the performance of EJM. The usefulness of EJM for calibration, therefore, remains unclear. In our opinion, the performance of EJM will be driven primarily by the quality of expert judgments, which may vary across applications. As pointed out by one reviewer, the performance of EJM might be improved by applying updates within a Bayesian framework and by incorporating uncertainty in expert judgments into the model.

One reviewer asked whether predictions are required at all. This question may be expanded to ask whether diagnoses or treatment decisions are required at all. In our opinion, the answer to all these questions is “Yes”. Clinicians need to make decisions for patients, and these decisions should be based on the best evidence available. These decisions primarily concern rendering a diagnosis or making a treatment decision. However, patients want to get information about their prognosis. In our opinion, it is thus of great clinical importance to provide reliable predictions.

A final aspect is whether online calibration or self-learning calibration would be an option to overcome the issue of calibration in the absence of calibration data. Indeed, outcome data could, in principle, be used as they come in, e.g., in blocks, and then used to successively calibrate the model. In our opinion, this approach is unlikely to be feasible in medical applications. In Europe, both a diagnostic test and a prediction model are subject to the Medical Device Regulation (MDR), under which their performance must be described in detail in the technical documentation (TD). This performance may not change instantaneously, but it has to remain constant. Otherwise, it would constitute a change of the medical device.

## 6 CONCLUSIONS

Machine learning models for probability estimation can be calibrated even in the absence of a calibration dataset. SimCal and Elkan performed well in the simulation studies and, in general, in the real data applications. ACP and SBA-10 conceptually do not appear to be reasonable for calibration if there are structural differences between study populations. EJM performed poorly on the real data. SimCal performed well when model parameters were correctly specified. We stress that none of the approaches performed convincingly well in all situations. Further studies to improve calibration when no calibration data are available are urgently needed.

## Data Availability

The code for and the results from all simulations is available as supplementary material. The simulated data are available on Zenodo (DiCarluccio 2026). The Cleveland Clinic data used for illustration are freely available from the machine learning data repository from the University of California in Irvine; details are provided in the supplementary material.

https://zenodo.org/records/21099064

https://archive.ics.uci.edu/ml/machine-learning-databases/heart-disease/processed

## AUTHOR CONTRIBUTIONS

E.D.C. analyzed the data and performed the simulation studies. A.Z. and E.D.C. drafted the manuscript. G.K. developed the modgo software and contributed in getting modgo run for this study. C.W. collected the stroke data and participated in the interpretation of the clinical results. F.M.O. contributed to the R code and the design of the study. A.Z. conceived, designed, and supervised the study. All authors revised the manuscript and approved its submission.

## ACKNOWLEDGMENTS

The German Stroke Study Collaboration was financed by the German Ministry of Education and Research (BMBF) as part of the Competence Net Stroke. We are grateful to the members of the German Stroke Study Collaboration for data collection, including the following departments and responsible study investigators (in alphabetical order): St. Katharinen-Hospital Frechen (R. Adams), Charité Berlin (N. Amberger), Städtisches Krankenhaus München-Harlaching (K. Aulich, M. J. L. Wimmer), Klinikum Minden (J. Glahn), University of Magdeburg (M. Goertler), Krankenanstalten Gilead Bielefeld (C. Hagemeister), Klinikum MünchenGroßhadern (G. F. Hamann, A. Müllner), Rheinische Kliniken Bonn (C. Kley), University of Rostock (A. Kloth), Benjamin Franklin University of Berlin (C. Koennecke), University of Saarland (P. Kostopoulos), Bürgerhospital Stuttgart (T. Mieck), Universities of Essen (G. Mörger-Kiefer, C. Weimar), Ulm (M. Riepe), Leipzig (D. S. Schneider), and Jena (V. Willig). We thank K. Kraywinkel, MD, MSc, and P. Dommes, PhD, for central data collection and management.

## DATA AVAILABILITY

The code for and the results from all simulations is available as supplementary material. The simulated data are available on Zenodo ^21^. The Cleveland Clinic data used for illustration are freely available from the machine learning data repository from the University of California in Irvine; details are provided in the supplementary material.

## USE OF ARTIFICIAL INTELLIGENCE

The authors used Claude (Sonnet 4.6) to assist with language editing and improving clarity of this manuscript. All content has been reviewed and revised by the authors. The authors take full responsibility for the accuracy and originality of the work.

## SUPPORTING INFORMATION

Additional supporting information may be found in the online version of the article at the publisher’s website.

- Supplementary Material 1: TRIPOD checklist for stroke data, LogReg regression coefficients for stroke data and Cleveland Clinic data, graphics for real data application (stroke and Cleveland Clinic data)
- Supplementary Material 2: Code for simulation studies
- Supplementary Material 3: a) R Markdown code for graphics and tables for Simulation Study 1, b) detailed results for Simulation Study 1
- Supplementary Material 4: a) R Markdown code for graphics and tables for Simulation Study 2, b) detailed results for Simulation Study 2
- Supplementary Material 5: a) R Markdown code for graphics and tables for Simulation Study 3, b) detailed results for Simulation Study 3
- Supplementary Material 6: a) R Markdown code for graphics and tables for Simulation Study 4, b) detailed results for Simulation Study 4
- Supplementary Material 7: R Quarto code for real data application (Cleveland Clinic data)
- All simulation data has been uploaded to and can be dowloaded from Zenodo ^21^

