## Supplementary material for "Calibrating machine learning approaches for probability estimation without calibration data": Supplementary_material_1.pdf

### 1 TRIPOD checklist

The 'TRIPOD Checklist: Prediction Model Development and Validation' was used to enhance the quality and transparency of our work. Detailed information corresponding to each item of the methods section is presented below. The study has been described in detail by Weimar et al. [7, 8] and König et al. [4]. In this work, we only considered features, i.e., covariates that were available within the first 6 hours after hospital admission.

#### 1.1 Source of data/4a: Describe the study design or source of data (e.g., randomized trial, cohort, or registry data), separately for the development and validation data sets, if applicable

For the development data set, registry data were collected prospectively within the German Stroke Database of the Stiftung Deutsche Schlaganfall-Hilfe in 23 neurology departments in 1998 and 1999. All participating hospitals had an acute stroke unit and, in most cases, also a neurological intensive care unit. They served catchment areas of 100,000 to 1 million inhabitants and were the main care providers for stroke patients in these regions.

In order to assure high data quality, only patients from those centers were considered that had included more than 90% and followed up more than 80% of their patients. Of these, all patients were included who additionally met the following criteria: No serious functional impairment (Rankin Scale $< 4$) before the event to ensure that patients were functionally independent to a certain degree, admission within 24 hours after stroke, not intubated during the first 6 hours to allow for a valid assessment of all relevant variables, survival in the first 6 hours, and follow up between 80 and 150 days after admission or death until follow up.

For the validation data sets, consecutive patients were registered during 2001 and 2002. Four neurology departments participating in the initial study met the required quality criteria and also took part in the validation study, allowing for temporal validation. Seven neurology departments met the required quality criteria and participated in the validation study only, thus contributed data for external validation. As before, all participating centers operated an acute stroke unit.

#### 1.2 Source of data/4b: Specify the key study dates, including start of accrual; end of accrual; and, if applicable, end of follow-up.

For the training data, 6412 patients with ischemic stroke were included in the database from January 1, 1998 until December 31, 1999. A follow-up at 80 to 150 (median 96) days after admission assessed mortality and functional outcomes on the Barthel Index.

For the validation data, 1834 patients with complete follow-up were included from January 1, 2001 until December 31, 2002. Follow-up after 100 days assessed mortality and functional outcome.

#### 1.3 Participants/5a: Specify key elements of the study setting (e.g., primary care, secondary care, general population) including number and location of centers.

For the training data set, data were collected in 7 hospitals that met the specified quality criteria (see above): Minden, München-Harlaching (only 1998), Essen, Benjamin Franklin Berlin, München-Großhadern (only 1998), Frechen and Leipzig.

For the validation data sets, data were collected in 13 centers: Charité Berlin, Krankenanstalten Gilead Bielefeld, Rheinische Kliniken Bonn, University of Saarland, University of Jena, University of Magdeburg, Klinikum Minden, Städtisches Krankenhaus München Harlaching, Klinikum München Großhadern, University of Rostock, Bürgerhospital Stuttgart, University of Ulm, University of Essen. Eleven centers met the specified quality criteria. As before, all participating hospitals operated an acute stroke unit and thus provide both primary and secondary care for acute stroke patients.

#### 1.4 Participants/5b: Describe eligibility criteria for participants.

The logistic regression model was taken from Weimar et al. [8]. Detailed descriptions about the participants for model development and internal validation can be found there. For developing the random forest and the boosting models, we used data from the stroke data bank of the German Stroke Foundation. In 1998 and 1999, 7238 patients with acute cerebral ischemic symptoms at admission were included in the database. Seven centers (Minden, München-Harlaching [only 1998], Essen, Benjamin Franklin Berlin, München-Großhadern [only 1998], Frechen, and Leipzig) met the specified quality criteria, registering a total of 3575 patients. Of the 3575 registered patients, only patients with a Rankin Scale score $< 3$ before the event ($n = 3281$), patients admitted within 6 hours after the onset of the stroke symptoms ($n = 1346$), and those who survived during the first 6 hours ($n = 1344$) were included. Of the remaining patients, 1079 patients were interviewed between 80 and 150 days after admission or were found to have died by the time of the interview. Mean age of these patients was 67.0 years (standard deviation (SD) 12.3), and 39.5% were women. After 100 days, 645 patients (59.8%) were completely restituted (BI $\geq 95$), 311 patients (28.8%) were incompletely restituted (BI $< 95$), and 123 patients (11.4%) had died.

For the validation data, patients were enrolled during 2001 and 2002. Four departments participating in the initial study also took part in the validation study, allowing for temporal validation. Nine departments participated in the validation study and thus contributed data for a combined temporal and external validation. As before, all participating centres operated an acute stroke unit. According to the study protocol, we excluded all patients from centers with less than 75% follow-up or a drop-out rate of $> 10\%$. Of the 1934 patients with complete follow-up after 100 days, 1100 patients (56.9%) were completely restituted, 609 patients (31.5%) incompletely restituted, and 225 patients (11.6%) had died.

#### 1.5 Participants/5c: Give details of treatments received, if relevant.

All participants were treated by neurologists according to best medical practice in each center.

#### 1.6 Outcome/6a: Clearly define the outcome that is predicted by the prediction model, including how and when assessed.

The Barthel Index (BI), approximately three months after stroke, is commonly used as a primary endpoint in randomized clinical trials [1], and mortality after three months is often used as a secondary endpoint in such randomized controlled trials [3]. The BI 100 days after an acute ischemic stroke was thus chosen as the primary outcome. This scale evaluates individual abilities in feeding, dressing, mobility (walking on a level surface and ascending/descending stairs), and personal hygiene (grooming, toileting, bathing, and control of bodily functions). It therefore adequately reflects functional consequences for daily activities that are immediately important to the patient. To identify patients with complete recovery, as advocated in clinical trials, a cutoff of BI $\geq 95$ versus $< 95$ was used. Mortality after 100 days was chosen as the second outcome variable, representing the worst possible outcome.

#### 1.7 Outcome/6b: Report any actions to blind assessment of the outcome to be predicted.

Functional outcome was assessed via central telephone interview with the patient or caregiver. The interviewer was not aware of baseline parameters or predicted outcome. Mortality was determined from death certificates.

#### 1.8 Predictors/7a: Clearly define all predictors used in developing or validating the multivariable prediction model, including how and when they were measured.

We identified possibly predictive variables in a systematic search of the literature, which is available upon request from the corresponding author. To allow for a very early prediction, we used the 16 variables age at event (in years), gender, history of stroke, myocardial infarction, arterial hypertension, and diabetes mellitus, as well as atrial fibrillation on first ECG at admission, and baseline neurologic impairments at admission as rated on the National Institutes of Health Stroke Scale (NIHSS) with single items (level of consciousness, gaze, motor right arm, motor left arm, motor right leg, motor left leg, dysarthria, extinction, and inattention) and the total score [8]. These variables can be routinely assessed on admission.

#### 1.9 Predictors/7b: Report any actions to blind assessment of predictors for the outcome and other predictors.

Data on the predictors were routinely collected on the stroke or intensive care unit and were not known as designated predictors.

#### 1.10 Sample size/8: Explain how the study size was arrived at.

On the basis of results from models developed on the training data, the required sample size for validation was determined using Monte Carlo simulations. For details, see König et al. [4].

#### 1.11 Missing data/9: Describe how missing data were handled (e.g., complete-case analysis, single imputation, multiple imputation) with details of any imputation method.

Missing values were imputed before analysis for sake of convenience in the training data. In detail, for each continuous variable imputed values were drawn from a normal distribution with the mean and standard deviation calculated from non-missing values. For each categorical variable, imputed values were drawn from a random sample with the probability of each category calculated from non-missing values. The imputation approach is valid if data are missing completely at random. Illustrative code for the imputation approach taken is provided in `supplementary_material_7.qmd`. There was only one individual with missing data in the temporal and the external validation data, and these two individuals were dropped from the analyses.

#### 1.12 Statistical analysis methods/10a: Describe how predictors were handled in the analyses.

The procedure has been described in detail by Weimar et al. [8]. Descriptive statistics were obtained for all 16 variables and the recruiting center. To model the relationship between the dichotomous outcome variables "complete restitution" yes/no and "mortality" yes/no using logistic regression, fractional polynomials were used on a randomly selected 25% of the total sample for the continuous variables. The best fit was obtained with inclusion of only the linear term. Because of substantive correlations with other variables and less predictive value or reliability than the respective correlated variable, 2 single variables were eliminated (NIHSS motor left arm and NIHSS motor right arm). The remaining 14 variables were fitted into the logistic regression models via forward, backward, and stepwise selection. For the complete restitution model, the number of events per predictor was $> 30$. Nevertheless, variables were retained only if their resulting probability value was $< 0.005$. For the mortality model, because of the lower events per predictor of 9, all variables with probability values $< 0.001$ were excluded. From models with all variables that resulted from any of the selection procedures, any variable with $p < 0.005$ (complete restitution model) or $p < 0.001$ (mortality model) was eliminated stepwise. To the remaining set of variables, every previously eliminated variable was again added and kept in the model if it fulfilled the same criteria. Finally, all 2-way interactions of the resulting variables were investigated and kept if $p < 0.005$ (complete restitution model) or $p < 0.001$ (mortality model). In addition, the proportion of explained variance $R^2$ was calculated for each model according to McKelvey-Zavoina. Leave-one-out cross-validation was used to estimate the shrinkage factor in both models. The threshold for classification with the use of the logistic distribution function was set so that the predicted proportion of events was equal to the observed.

##### 1.13 Statistical analysis methods/10b: Specify type of model, all model-building procedures (including any predictor selection), and method for internal validation.

For the logistic regression models, all 16 variables were fitted into logistic regression models as described in detail in 10b.

For random forests (RF) and gradient boosting (GB), all 16 variables were included in the model-building procedure. Different parameters were used to build the model and the LogLoss was used to assess the model performance. Parameters with the lowest LogLoss value for each machine were selected as the final model.

##### 1.14 Statistical analysis methods/10c: For validation, describe how the predictions were calculated.

For all three machines, the final models were used to calculate the predictions in the temporal and the external validation data set, respectively. The predictions were then calibrated using the different approaches detailed in the main text.

##### 1.15 Statistical analysis methods/10d: Specify all measures used to assess model performance and, if relevant, to compare multiple models.

The measures used to assess model performance were detailed in the Section 'Performance measures' of the main text.

##### 1.16 Statistical analysis methods/10e: Describe any model updating (e.g., recalibration) arising from the validation, if done.

The calibration approaches were detailed in the main text.

##### 1.17 Risk groups/11: Provide details on how risk groups were created, if done.

Not applicable.

##### 1.18 Development vs. validation/12: For validation, identify any differences from the development data in setting, eligibility criteria, outcome, and predictors.

To be included in the validation/calibration part, hospitals had to meet specific quality criteria. Four such hospitals participating in the development study took part in the validation study, contributing data for a temporal validation. Seven such hospitals participated in the validation study only and thus contributing data for an external validation.

#### 2 Logistic regression

##### 2.1 Application 1: Prediction of functional outcome after stroke — German Stroke Study Collaboration data

The development of the logistic regression model is fully described by Weimar et al. [8]. Also see the previous section. Tables S1 and S2 display the results of the logistic regression model reported by Weimar et al. [8].

##### 2.2 Application 2: Diagnosis of coronary artery disease — Cleveland Clinic data

The development of the logistic regression model for the Cleveland Clinic data has been described in detail by Kruppa et al. [6]. The R code for the analysis of the Cleveland Clinic data is provided in Supplementary

Table S1: Model I to predict complete restitution ($BI \geq 95$) versus incomplete restitution ($BI < 95$) or mortality. Parameter estimates (log Odds ratios), standard errors, Odds ratios, and 95% confidence intervals are presented. Table from Weimar et al. [8], Table 2.

| **Variable** | **log odds ratio** | **Standard error** | **Odds ratio** | **95% CI** |
|---|---|---|---|---|
| Intercept | -5.782 | 0.537 | | |
| Age (difference of 1 year) | 0.049 | 0.007 | 1.051 | 1.036 – 1.065 |
| NIHSS total score at admission (difference of 1 scale score) | 0.272 | 0.018 | 1.313 | 1.268 – 1.360 |

Table S2: Model II to predict mortality versus survival. Parameter estimates (log Odds ratios), standard errors, Odds ratios, and 95% confidence intervals are presented. Table from Weimar et al. [8], Table 3.

| **Variable** | **log odds ratio** | **Standard error** | **Odds ratio** | **95% CI** |
|---|---|---|---|---|
| Intercept | -7.040 | 0.778 | | |
| Age (difference of 1 year) | 0.049 | 0.010 | 1.050 | 1.029 – 1.071 |
| NIHSS total score at admission (difference of 1 scale score) | 0.155 | 0.015 | 1.168 | 1.134 – 1.203 |

Table S3: Logistic regression model in the training data of the Cleveland Clinical data. Parameter estimates (log Odds ratios), standard errors, Odds ratios, and 95% confidence intervals (95% CI) are presented.

| Variable | **log odds ratio** | **Standard error** | **Odds ratio** | 95**% CI** |
|---|---|---|---|---|
| Intercept | −0.303 | 1.346 | 0.739 | 0.053 – 10.331 |
| Gender (men) | 1.556 | 0.340 | 4.740 | 2.434 – 9.229 |
| Chest pain type | 0.871 | 0.169 | 2.389 | 1.716 – 3.328 |
| Maximum heart rate | −0.030 | 0.008 | 0.970 | 0.955 – 0.986 |
| ST depression | 0.699 | 0.152 | 2.012 | 1.493 – 2.710 |

Material 7. In brief, backward selection for logistic regression was done using 10 independent variables, all entering the model linearly, and the final model included four variables (Table S3).

Independent variables were

- age (in years),
- gender (1 = men, 0 = women),
- chest pain type (cp with values 1 = typical angina, 2 = atypical angina, 3 = non-anginal pain, and 4 = asymptomatic),
- resting blood pressure (trestbps in mm Hg) on admission to the hospital,
- serum cholesterol (chol in mg/dl),
- fasting blood sugar > 120 mg/dl (fbs with values 1 = true, and 0 = otherwise),
- resting electrocardiographic results (restecg with values 0 = normal, 1 = having ST-T wave abnormality (T wave inversions and/or ST elevation or depression of > 0.05 mV), and 2 = showing probable or definite left ventricular hypertrophy by Estes' criteria),
- maximum heart rate achieved (thalach),
- exercise induced angina (exang with values 1 = yes, 0 = no),
- ST depression induced by exercise relative to rest (oldpeak),
- the slope of the peak exercise ST segment (slope with values 1 = upsloping, 2 = flat, and 3 = downsloping),
- number of major vessels (ca with values 0–3) colored by fluoroscopy,

- exercise thallium scintigraphic defects (thall with values 3 = normal, 6 = fixed, and 7 = reversible).

Three categorical variables (slope of the peak exercise ST segment, number of major vessels, exercise thallium scintigraphic defects) had too many missing values in the calibration data and were therefore removed from the analysis.

### 3 Missing data plots

One reviewer asked to provide detailed information about missingness in both real data applications.

#### 3.1 Stroke data

The three figures below describe the missingness and the missingness pattern in the stroke training data. We refrained from creating missingness figures for the temporal and external validation data because there was just one individual with missing data per dataset.

The UpSet plot shows that fourteen patients out of $n = 1097$ had missing information only for atrial fibrillation (AF), hypertension only was missing in 9 individuals, information solely on stroke and myocardial infarction was missing in 8 patients. All other variables and combinations of variables had missing information in less than 5 individuals.

Figure S1: UpSet plot for missing data in the stroke training data.

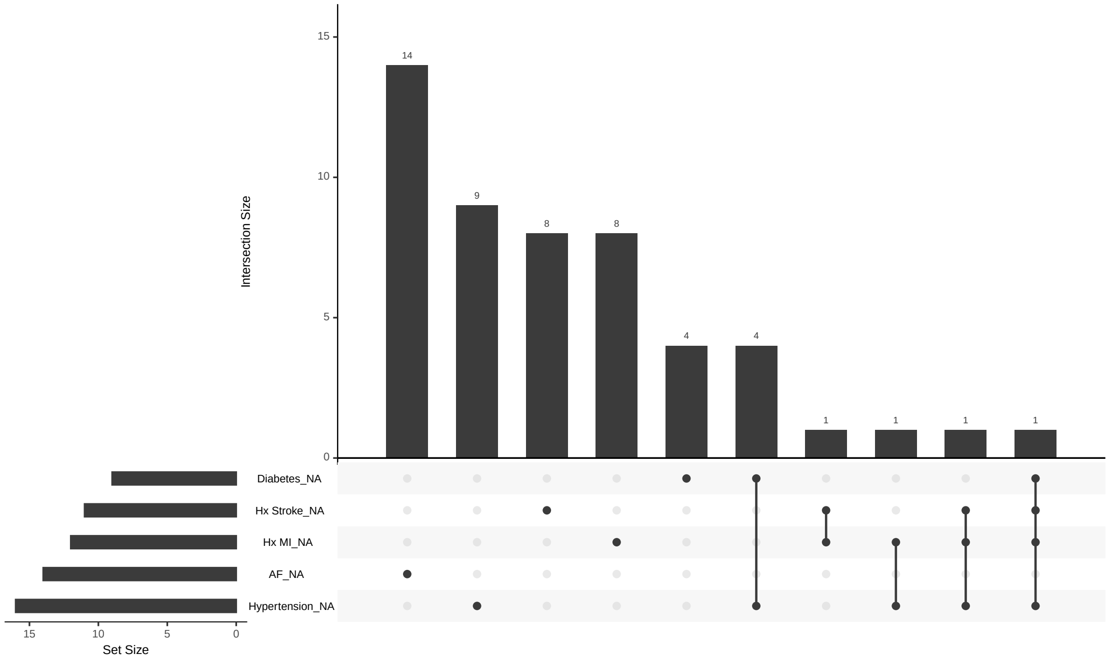


#### 3.2 Cleveland Clinic data

There were no missing data for the Cleveland Clinic data (Supplementary Figure S4). All three centers from the validation studies had missing data. Most importantly, the variable serum cholesterol was not coded as missing (-9) but as 0 for all individuals from the Swiss validation cohort. We accepted this physiologically impossible value as true values in the analyses.

Figure S2: Missingness lollipop plot for the stroke training data.

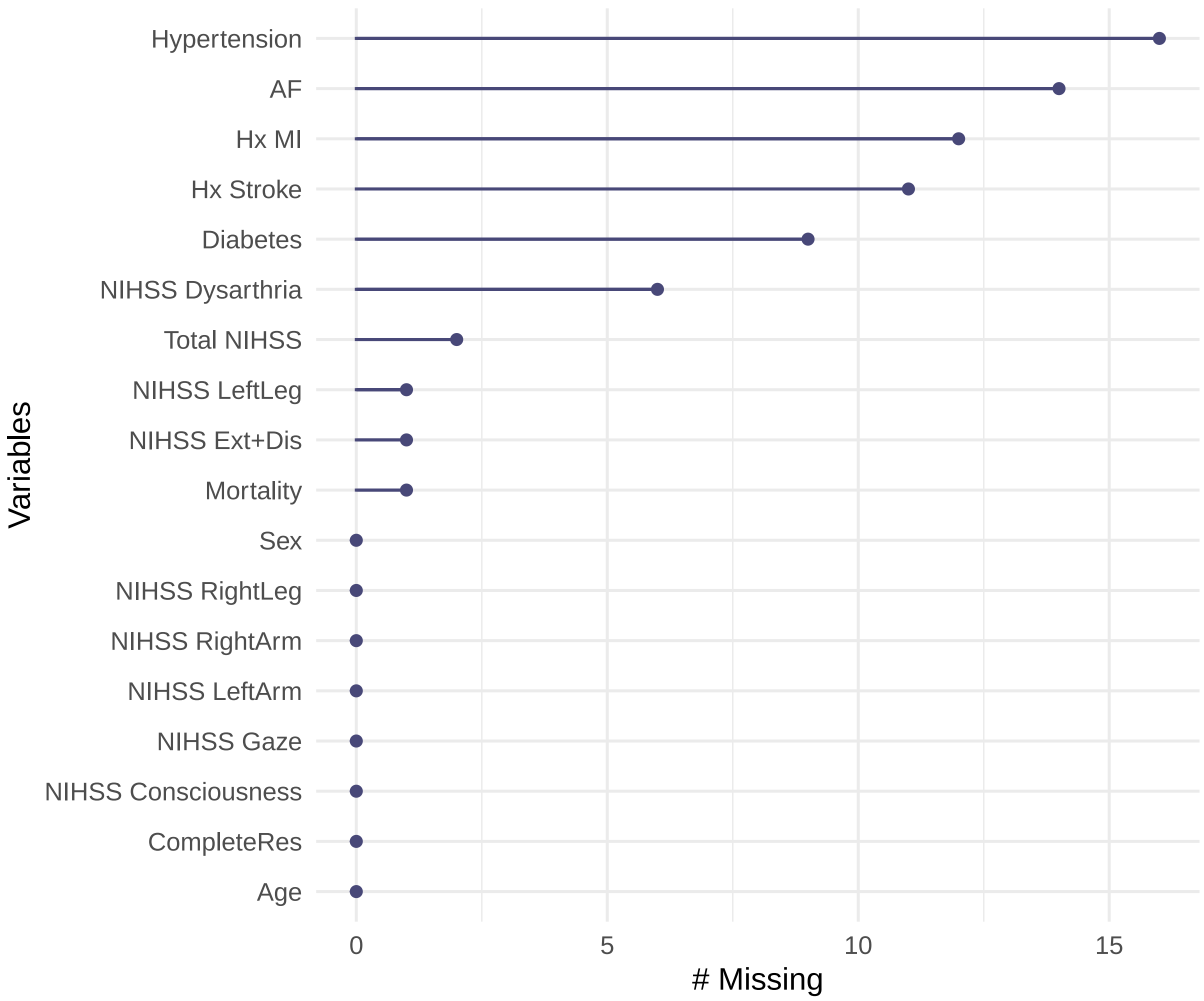

Figure S3: Missingness map for the stroke training data.

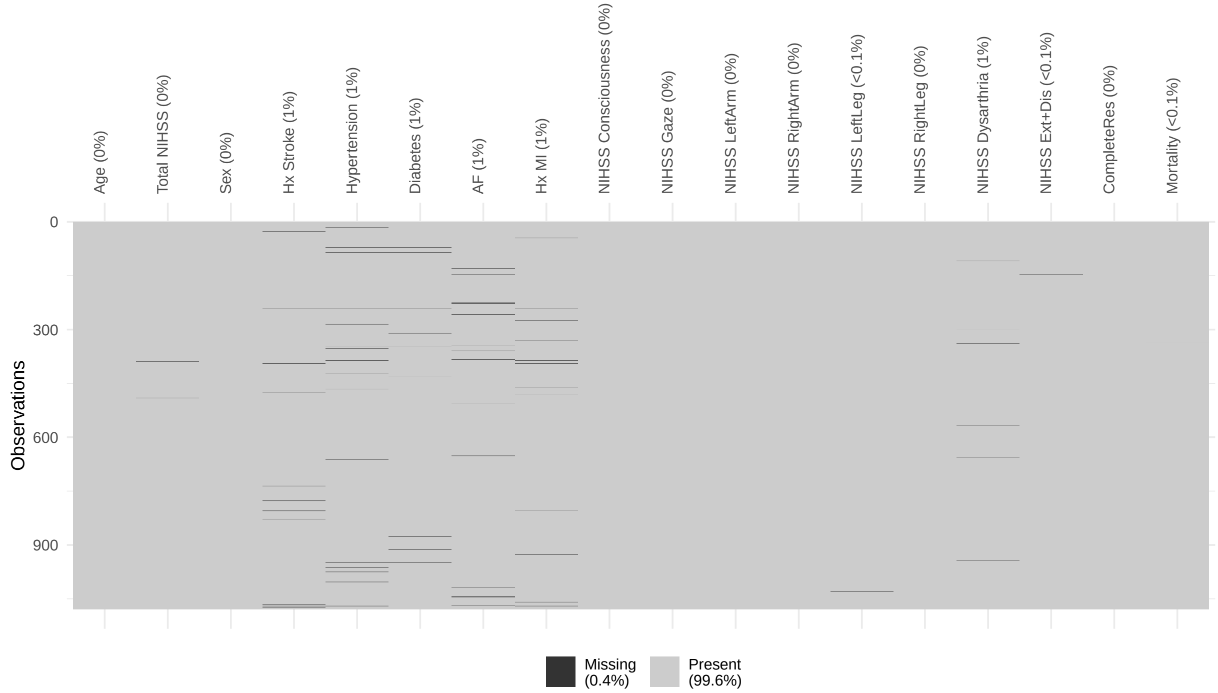

Figure S4: Missingness map for the Cleveland Clinic data for all centers.

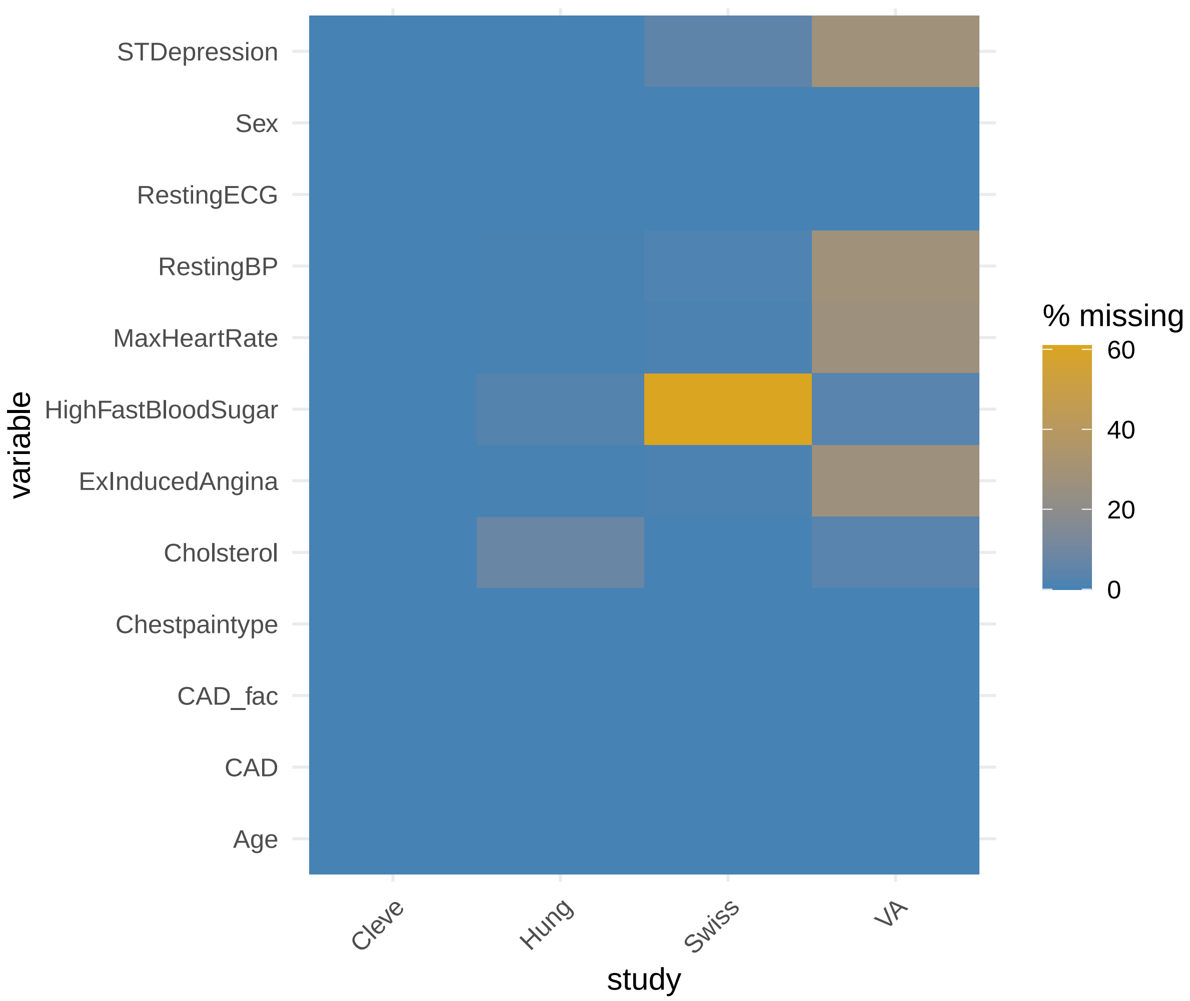

Figure S5: UpSet plot for missing data in the Swiss cohort from the Cleveland Clinic data.

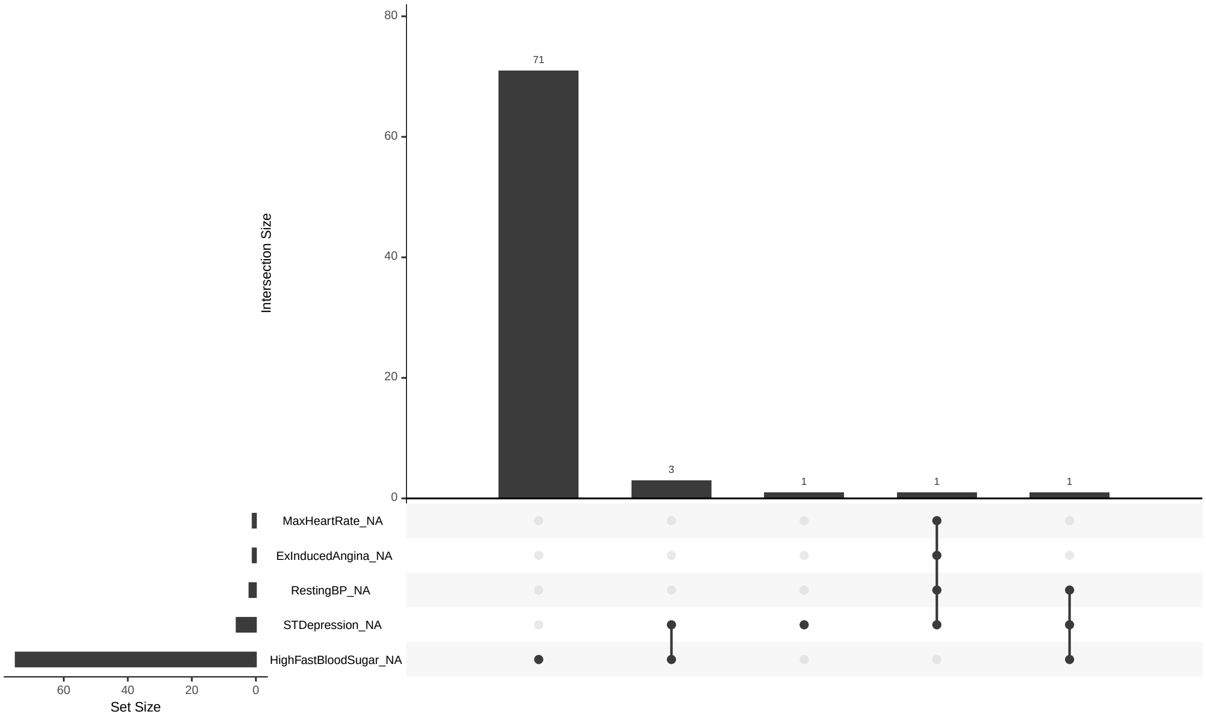

Figure S6: Missingness lollipop plot in the Swiss cohort from the Cleveland Clinic data.

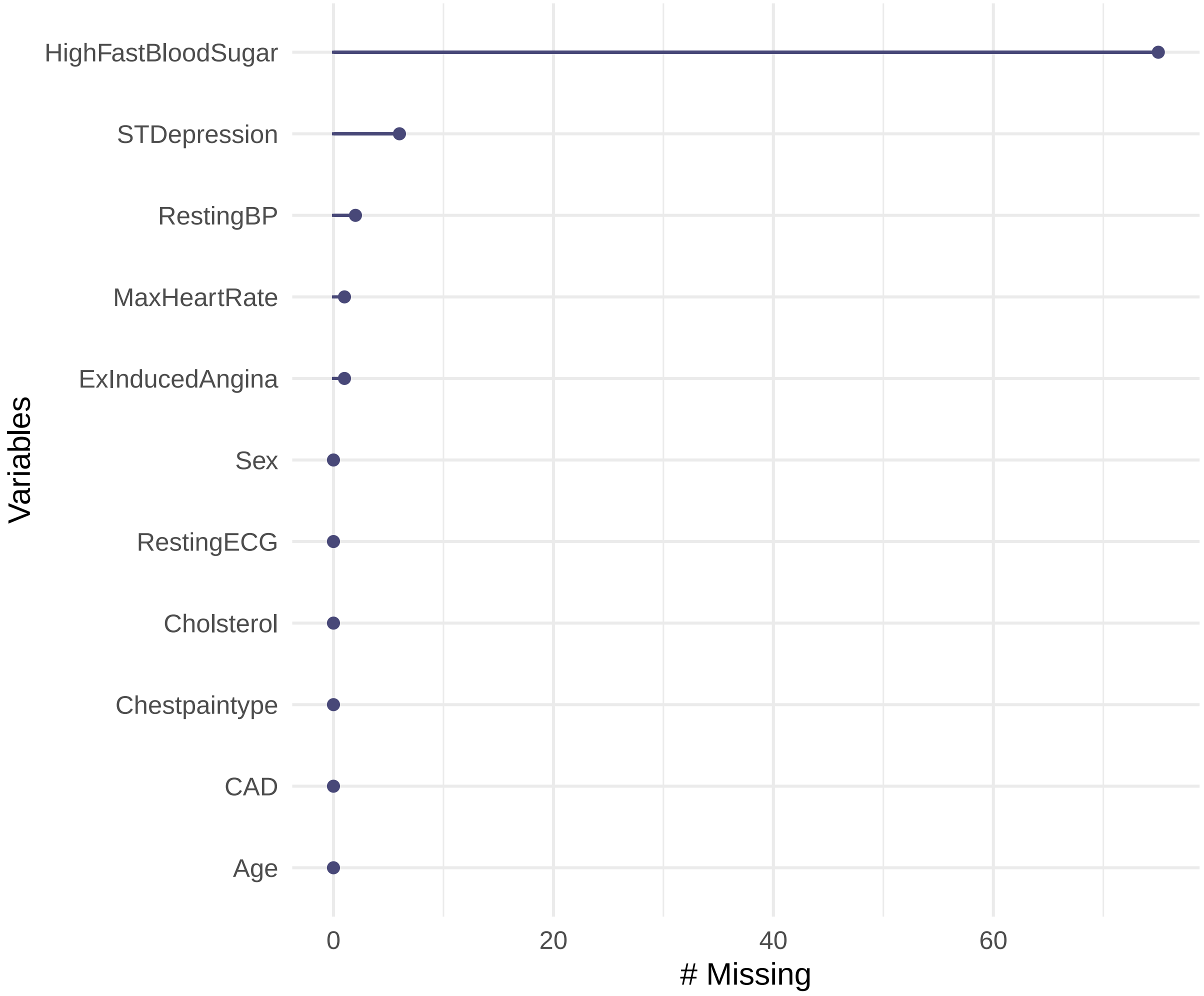

Figure S7: Missingness map in the Swiss cohort from the Cleveland Clinic data.

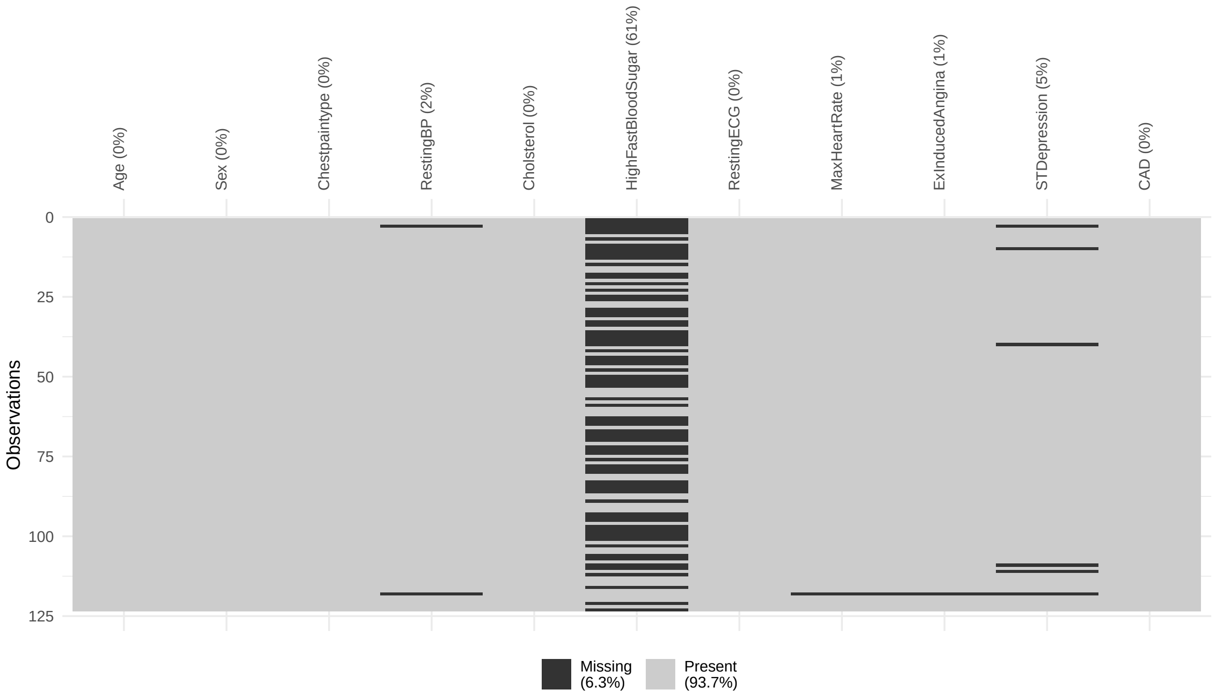


Figure S8: UpSet plot for missing data in the Veterans cohort from the Cleveland Clinic data.

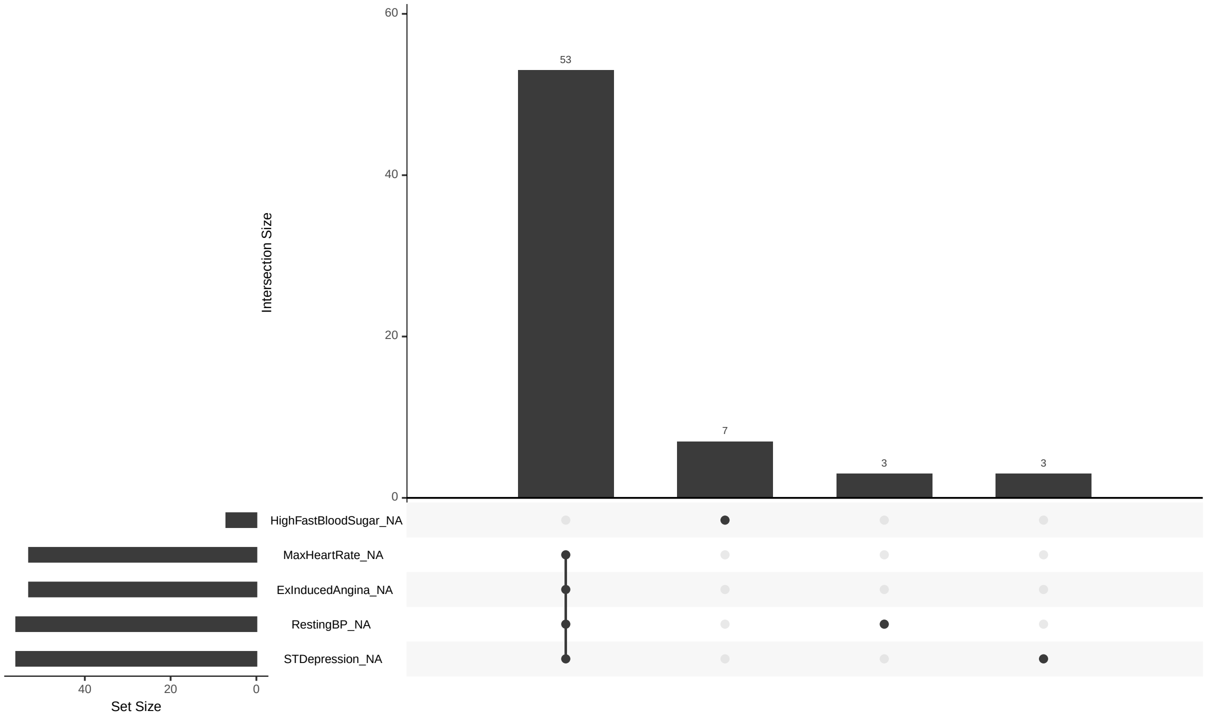

Figure S9: Missingness lollipop plot in the Veterans cohort from the Cleveland Clinic data.

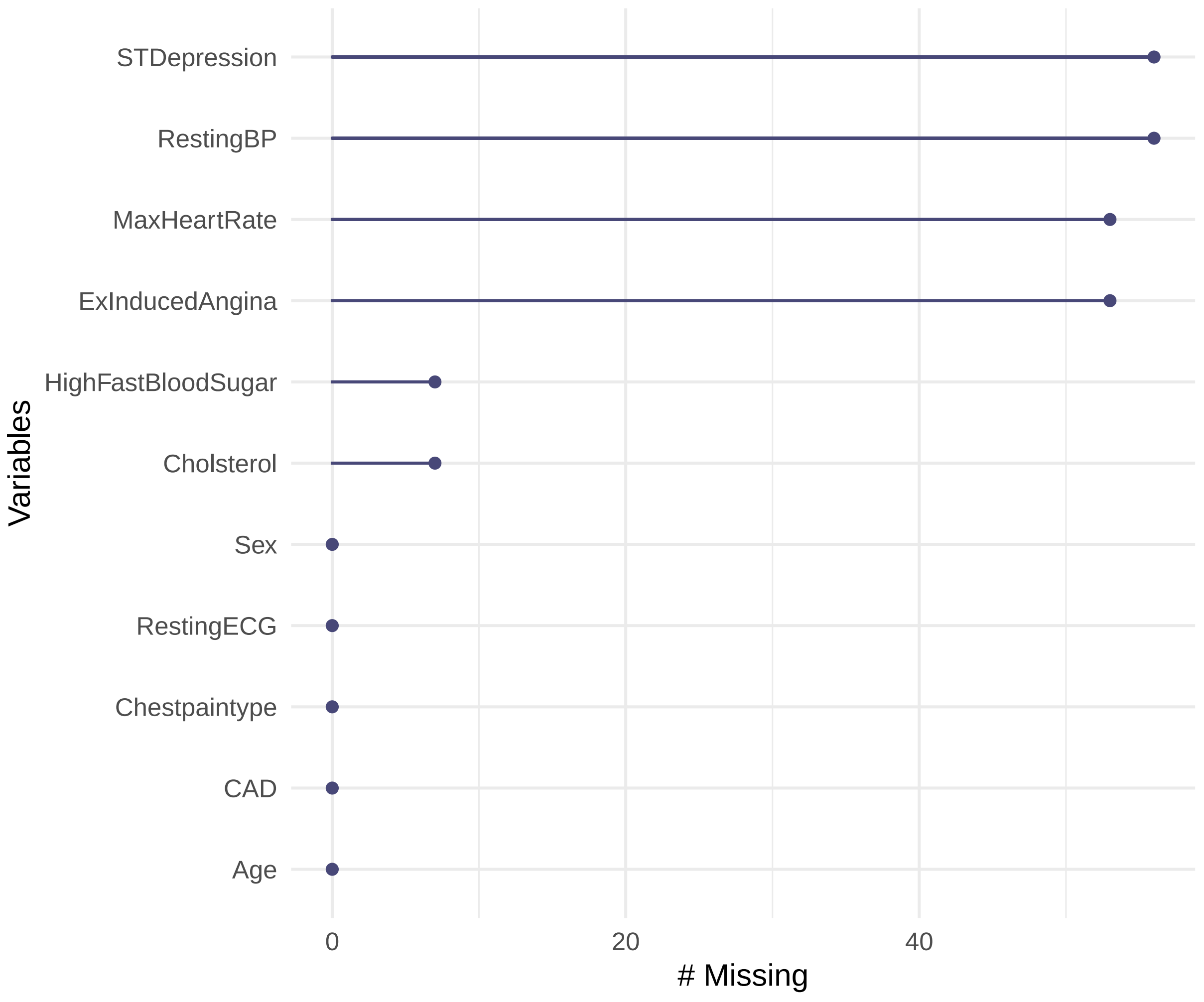

Figure S10: Missingness map in the Veterans cohort from the Cleveland Clinic data.

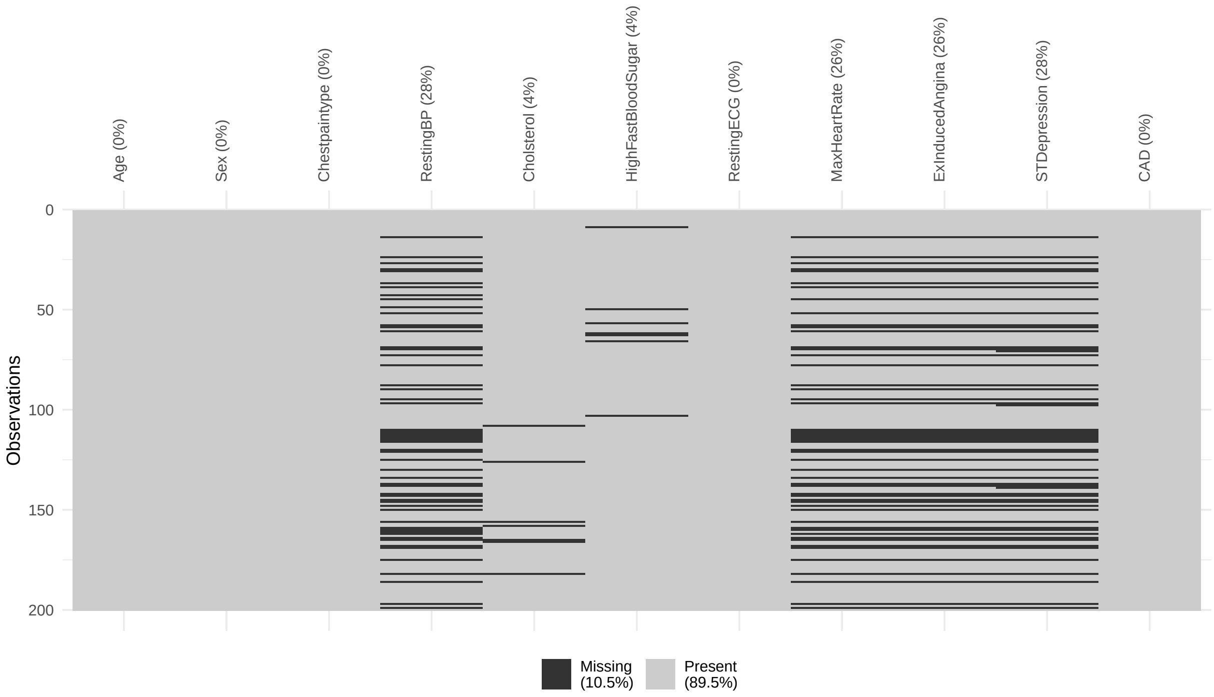


Figure S11: UpSet plot for missing data in the Hungarian cohort from the Cleveland Clinic data.

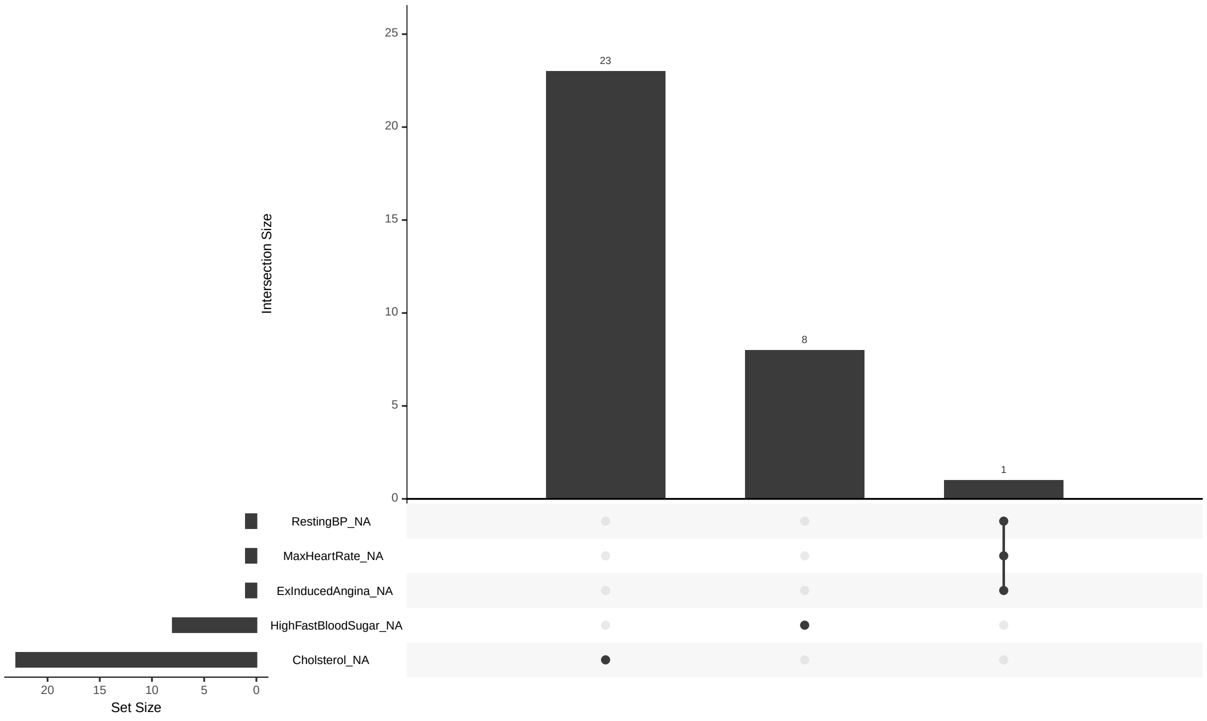

Figure S12: Missingness lollipop plot in the Hungarian cohort from the Cleveland Clinic data.

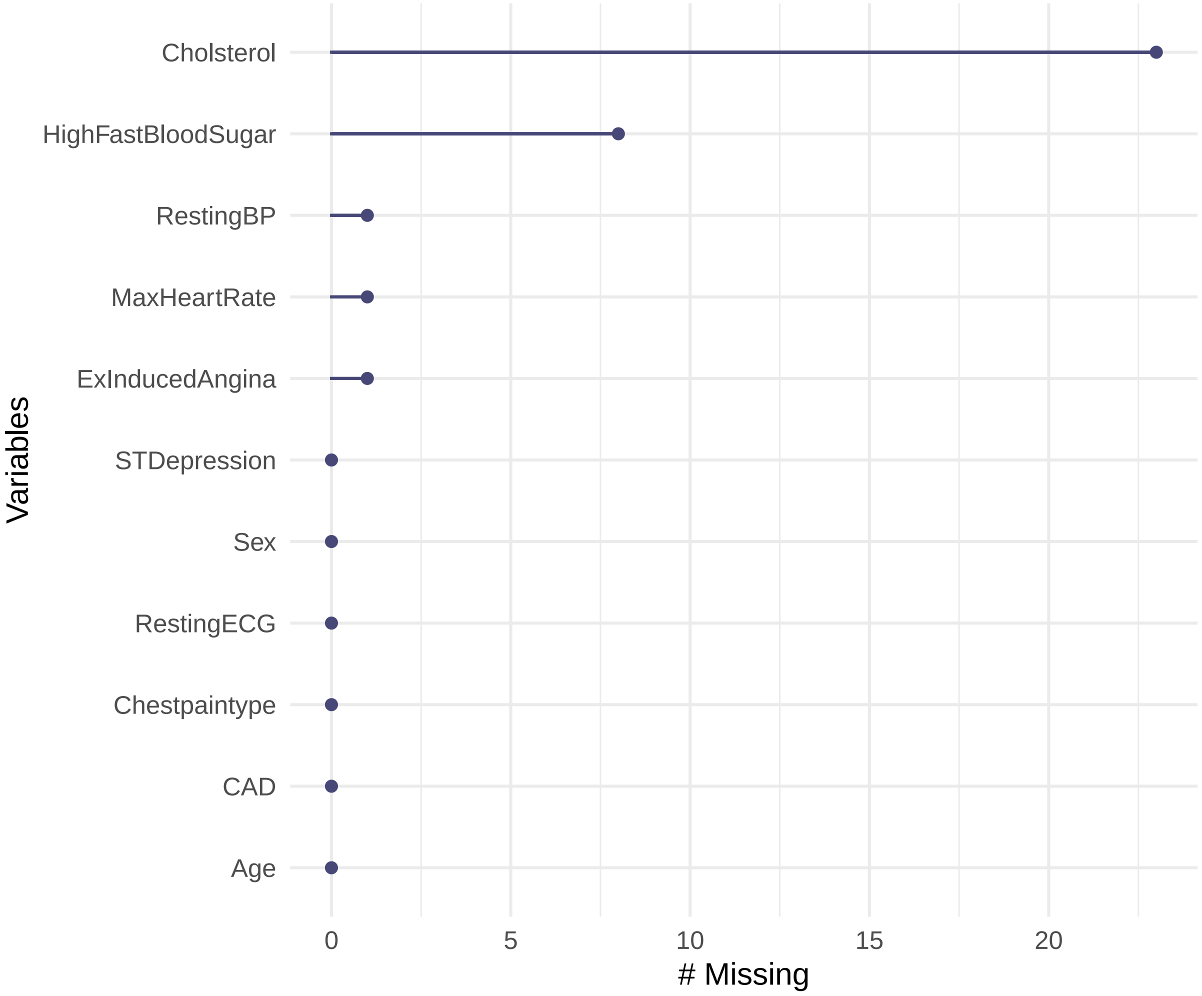

Figure S13: Missingness map in the Hungarian cohort from the Cleveland Clinic data.

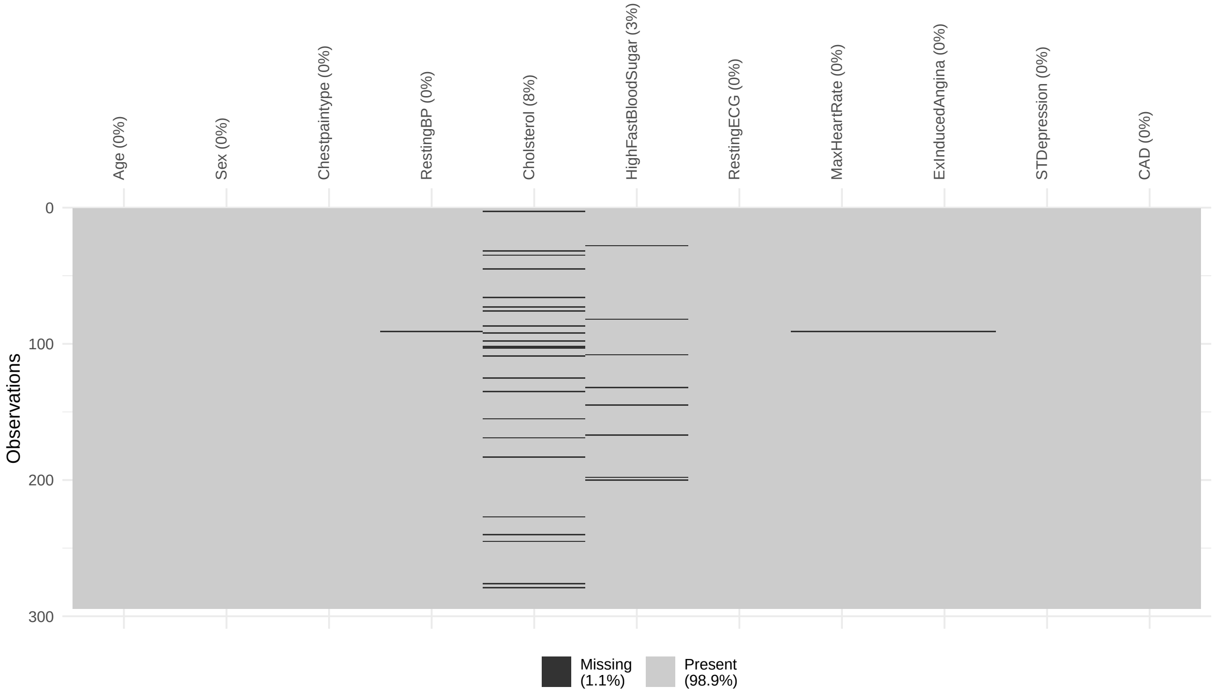

### 4 Additional plots for both real data applications

Figure S14: Critical difference plot for the LogLoss for the stroke data for probability estimation with logistic regression. Lower average ranks are considered better. Groups of calibration approaches connected by a horizontal line segment could not be shown to have a significantly different performance. The critical difference was 0.9. Panel A: complete restitution model in the temporal validation data; panel B: complete restitution model in the external validation data; panel C: mortality model in temporal validation data; panel D: mortality model in external validation data.

A)

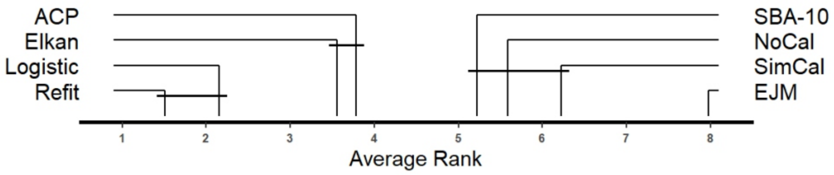


B)

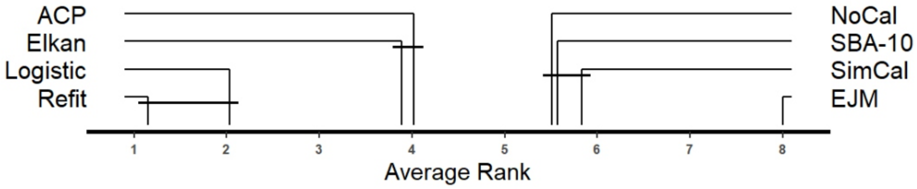


C)

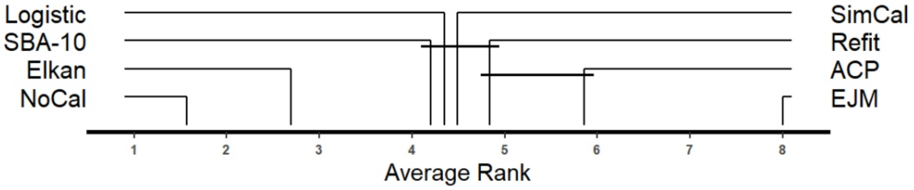


D)

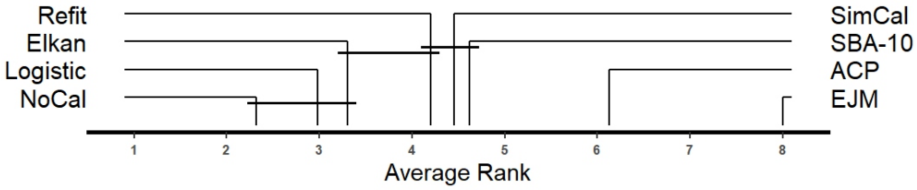

Figure S15: Critical difference plot for the LogLoss for the stroke data for probability estimation with random forest. Lower average ranks are considered better. Groups of calibration approaches connected by a horizontal line segment could not be shown to have a significantly different performance. The critical difference was 0.9. Panel A: complete restitution model in the temporal validation data; panel B: complete restitution model in the external validation data; panel C: mortality model in temporal validation data; panel D: mortality model in external validation data.

A)

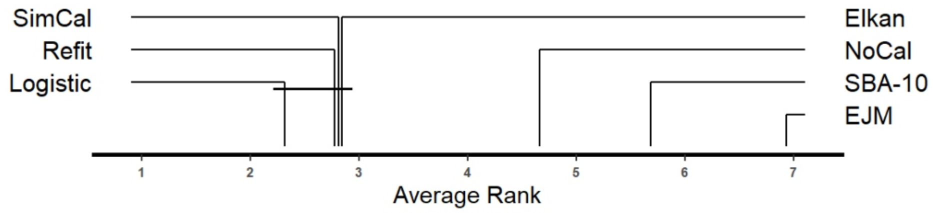


B)

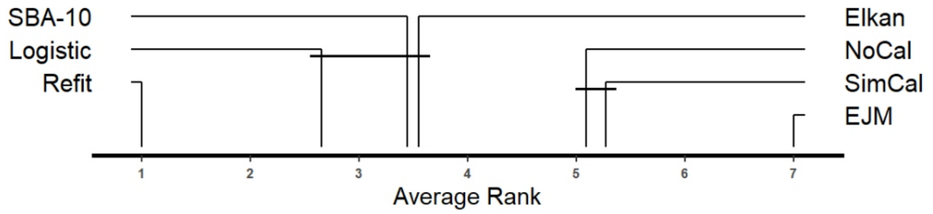


C)

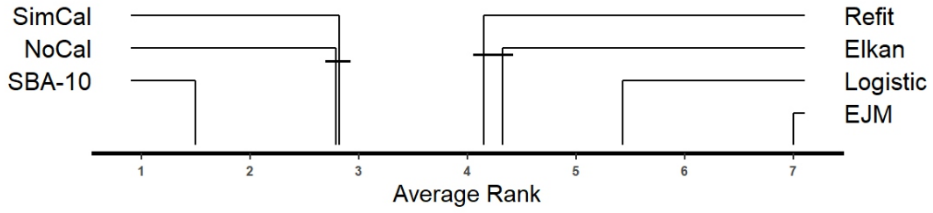


D)

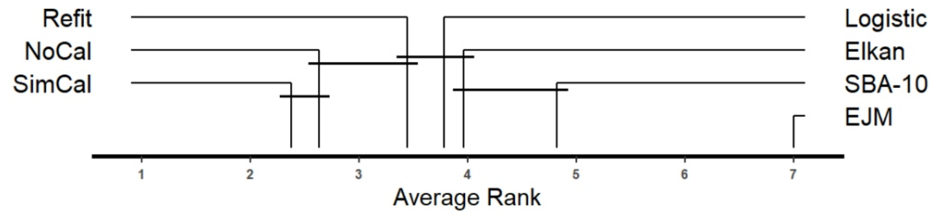

Figure S16: Critical difference plot for the LogLoss for the stroke data for probability estimation with gradient boosting. Lower average ranks are considered better. Groups of calibration approaches connected by a horizontal line segment could not be shown to have a significantly different performance. The critical difference was 0.9. Panel A: complete restitution model in the temporal validation data; panel B: complete restitution model in the external validation data; panel C: mortality model in temporal validation data; panel D: mortality model in external validation data.

A)

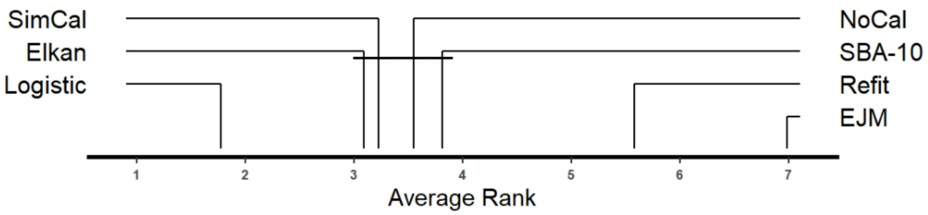


B)

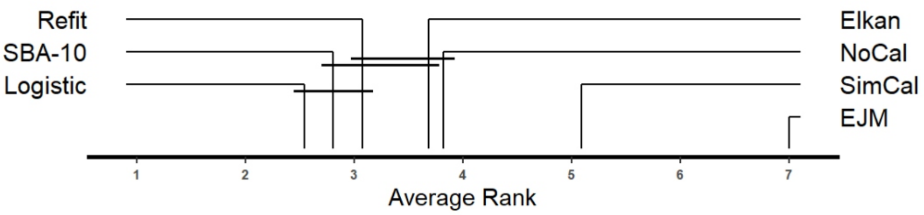


C)

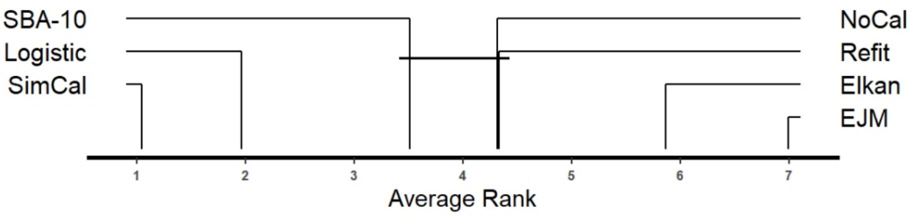


D)

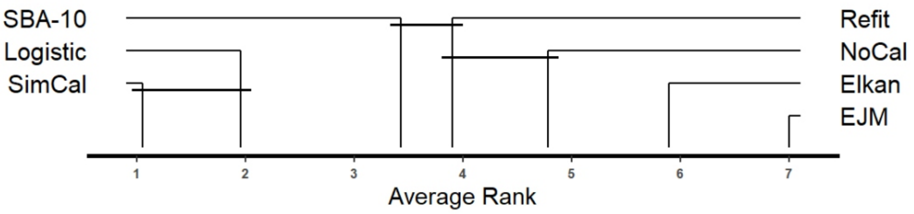

Figure S17: Boxplots for calibrated learning machines for probability estimation in the complete restitution model of the stroke data using the LogLoss (x-axis) as performance measure for logistic regression, random forest, and gradient boosting for the temporal data.

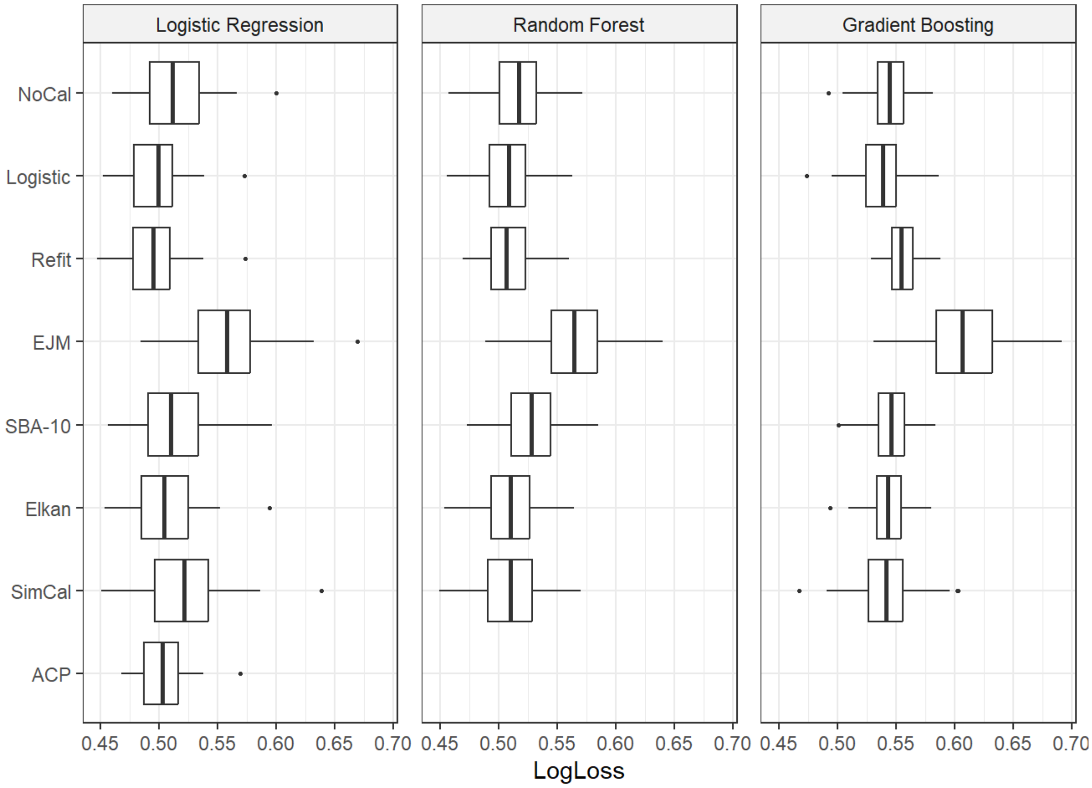

Figure S18: Boxplots for calibrated learning machines for probability estimation in the complete restitution model of the stroke data using the LogLoss (x-axis) as performance measure for logistic regression, random forest, and gradient boosting for the external data.

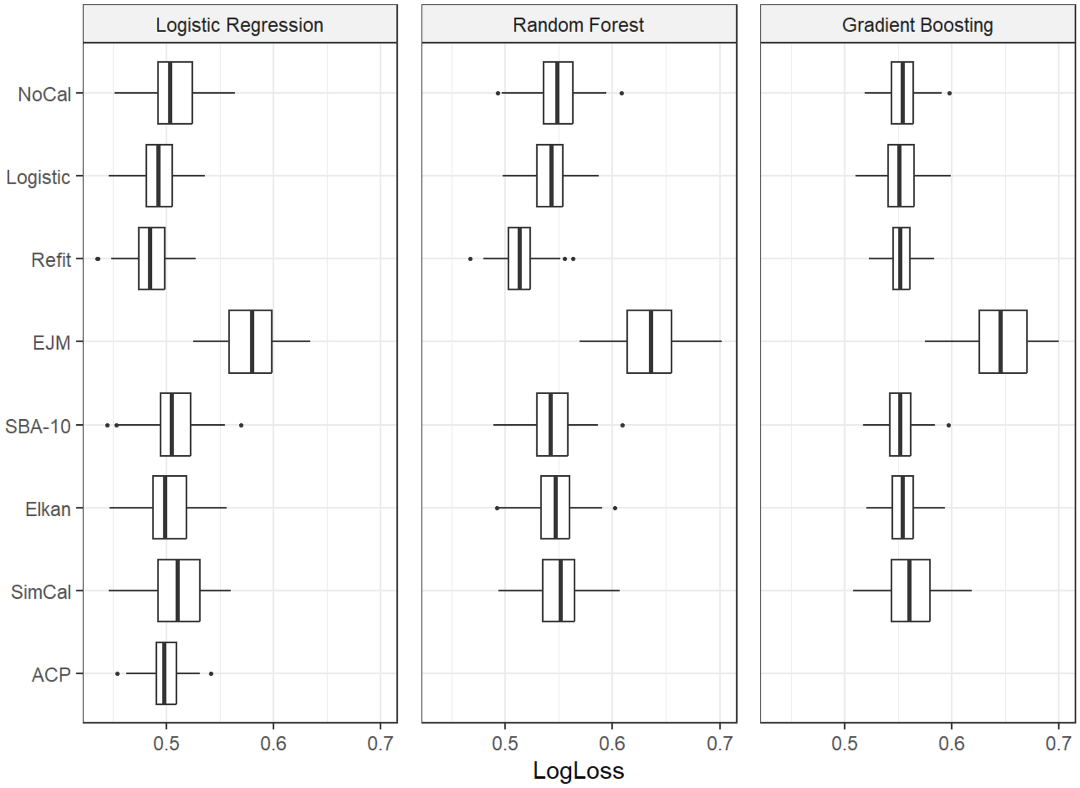

Figure S19: Boxplots for calibrated learning machines for probability estimation in the mortality model of the stroke data using the LogLoss (x-axis) as performance measure for logistic regression, random forest, and gradient boosting for the temporal data.

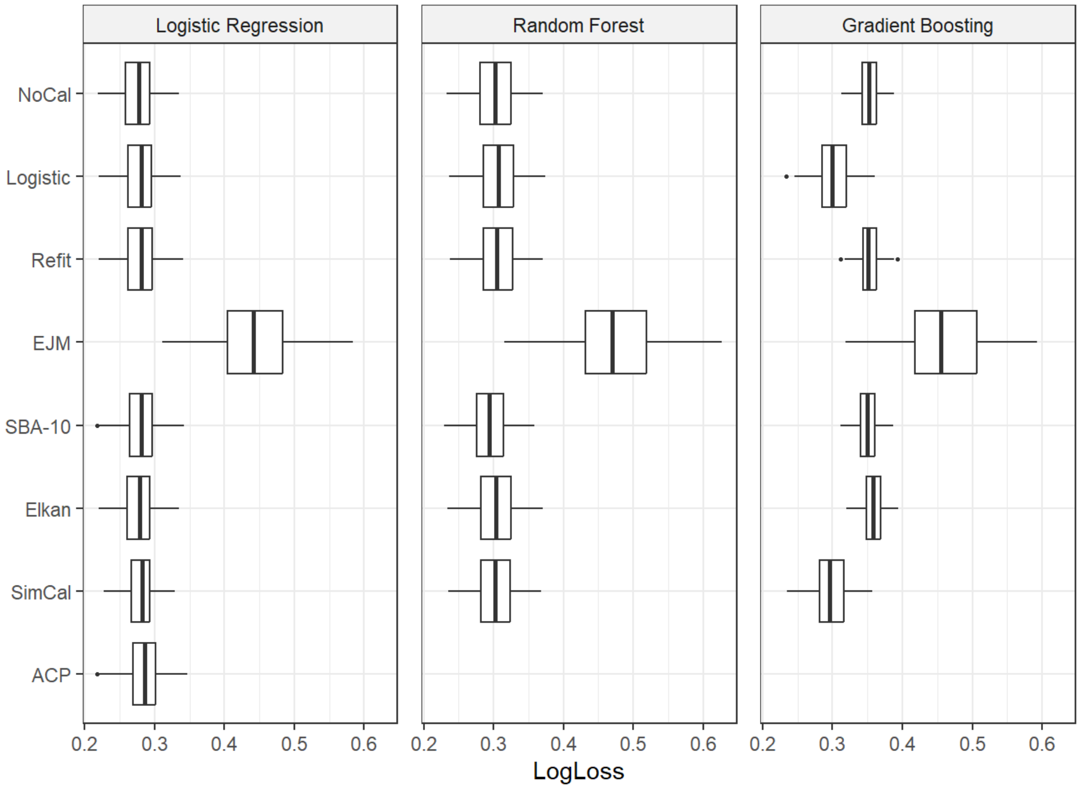

Figure S20: Boxplots for calibrated learning machines for probability estimation in the mortality model of the stroke data using the LogLoss (x-axis) as performance measure for logistic regression, random forest, and gradient boosting for the external data.

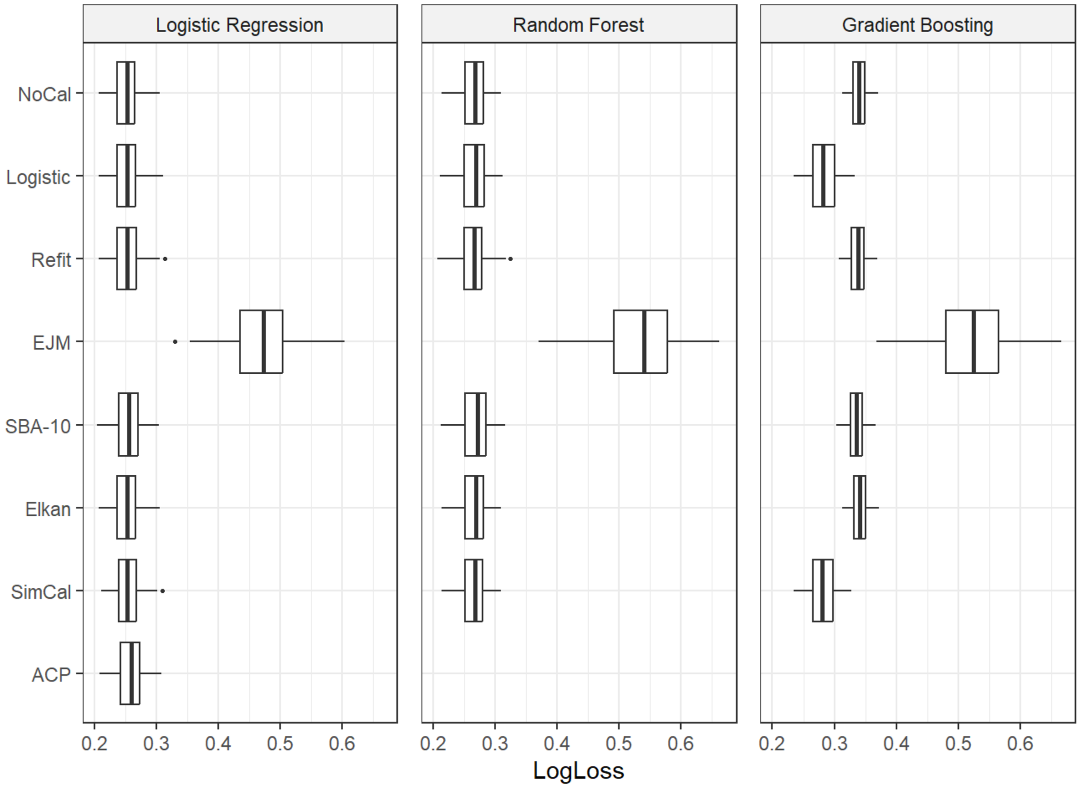


Figure S21: Center-specific and average (dashed black) true calibration plots for the validation cohorts from the Cleveland Clinic data for the logistic regression model. Blue: Hungarian; green: Swiss; yellow: Veterans

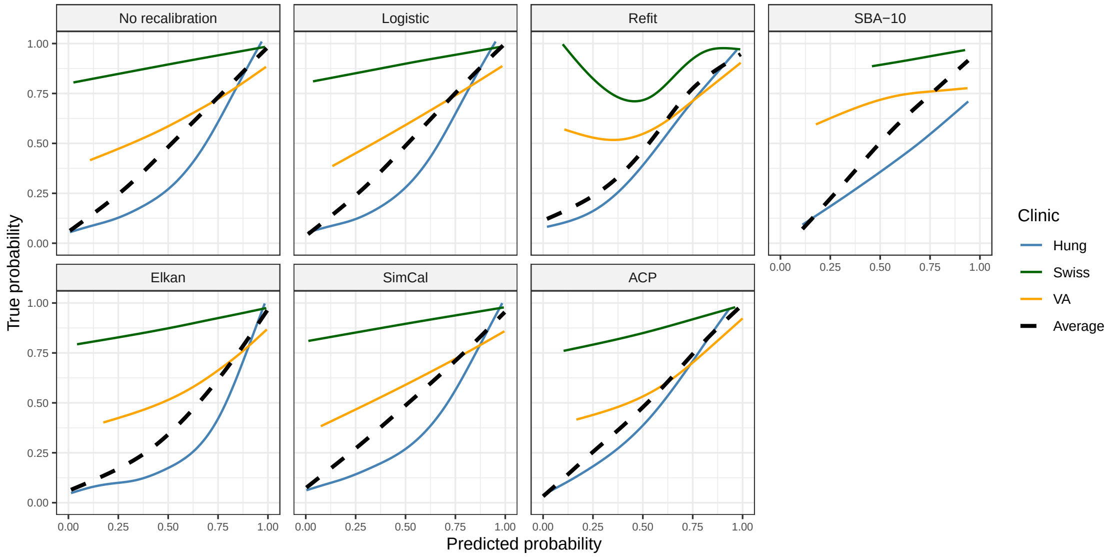

Figure S22: Center-specific and average (dashed black) true calibration plots for the validation cohorts from the Cleveland Clinic data for the gradient boosting model. Blue: Hungarian; green: Swiss; yellow: Veterans

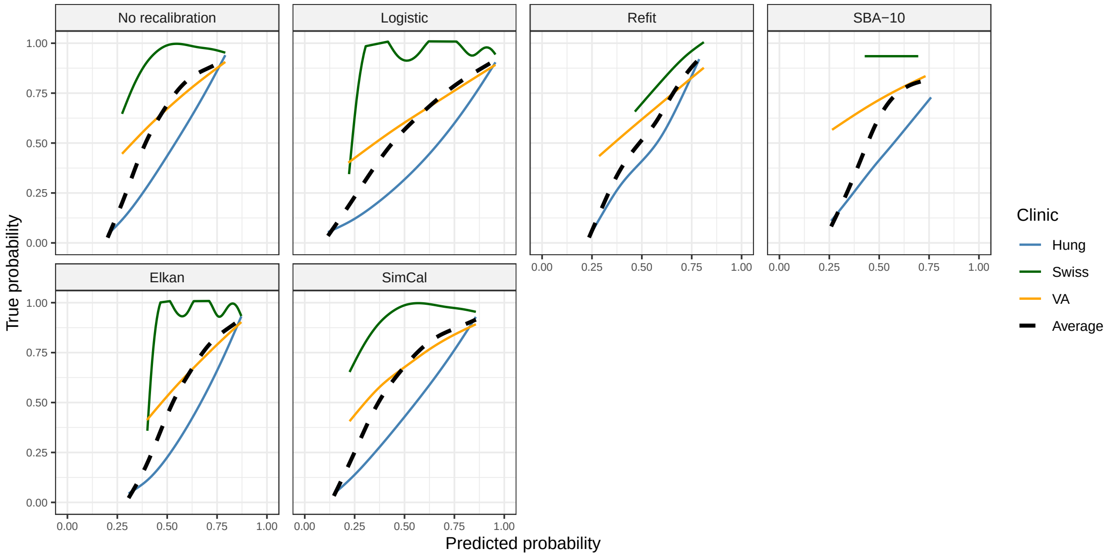


Figure S23: Center-specific and average (dashed black) true calibration plots for the validation cohorts from the Cleveland Clinic data for the random forest model. Blue: Hungarian; green: Swiss; yellow: Veterans

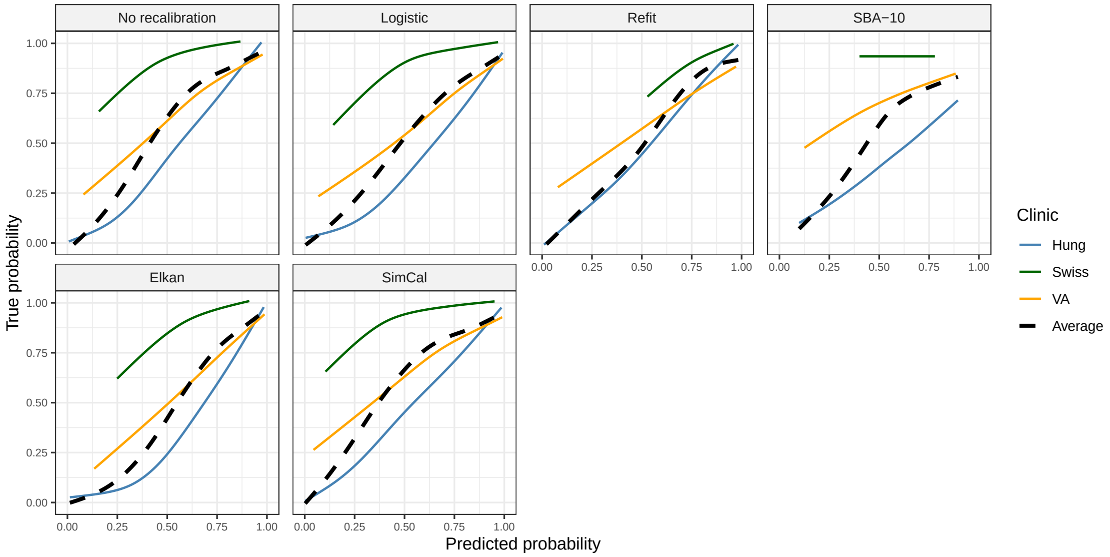

Figure S24: Calibration plots for calibrated logistic regression in the Cleveland Clinic validation data.

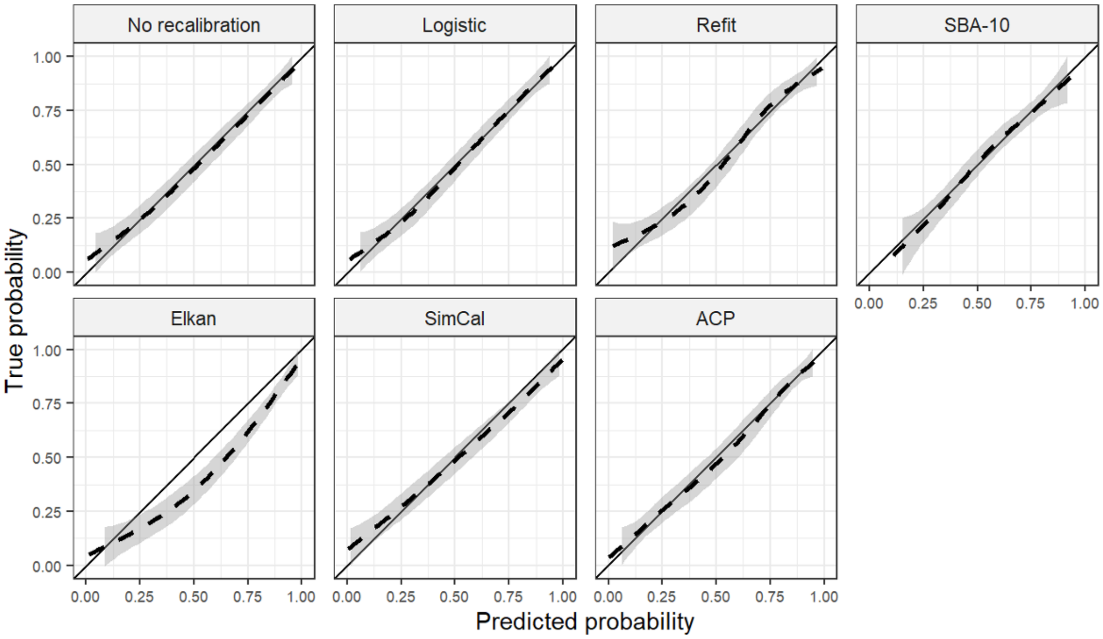

Figure S25: Calibration plots for calibrated random forests in the Cleveland Clinic validation data.

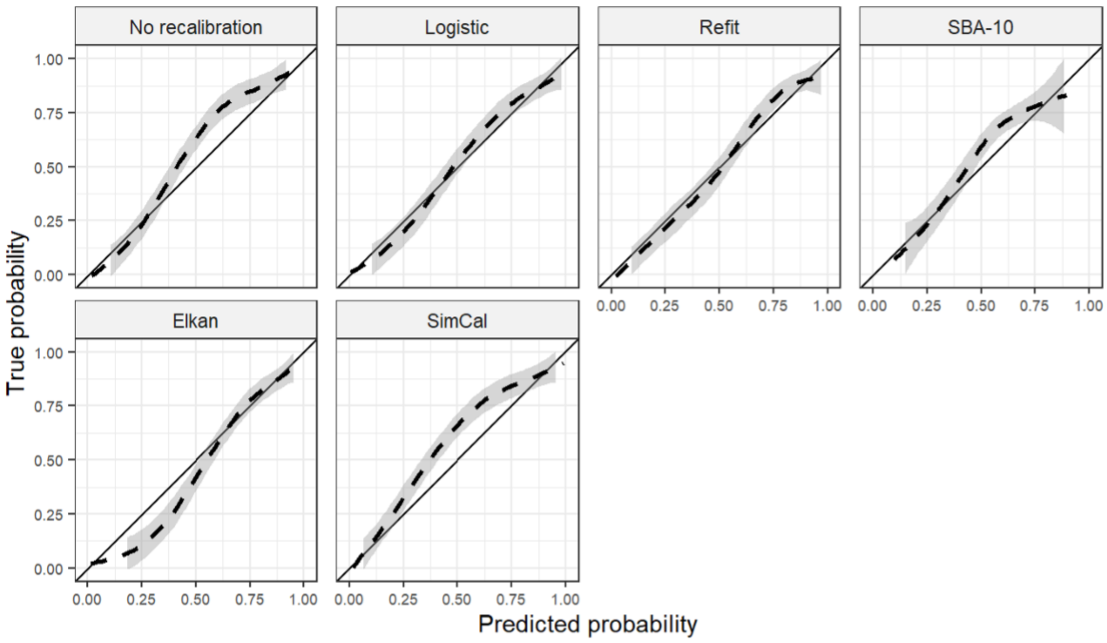

Figure S26: Calibration plots for calibrated gradient boosting in the Cleveland Clinic validation data.

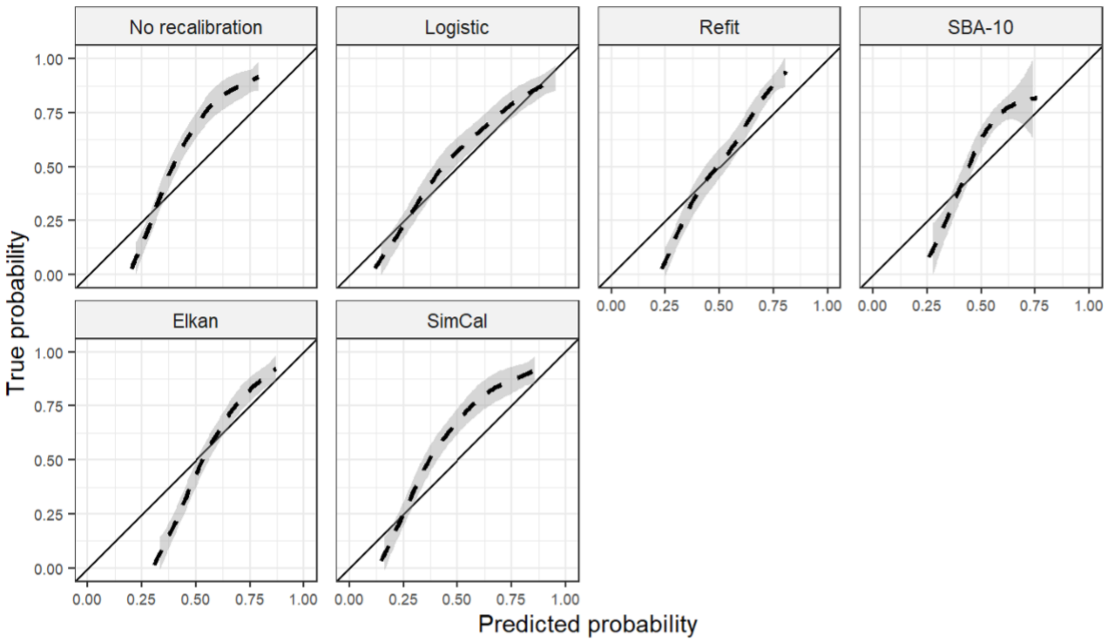

Figure S27: Critical difference plot for the LogLoss in the Cleveland Clinic validation data for logistic regression. Lower average ranks are considered better. Groups of calibration approaches connected by a horizontal line segment could not be shown to have a significantly different performance. The critical difference was 0.8.

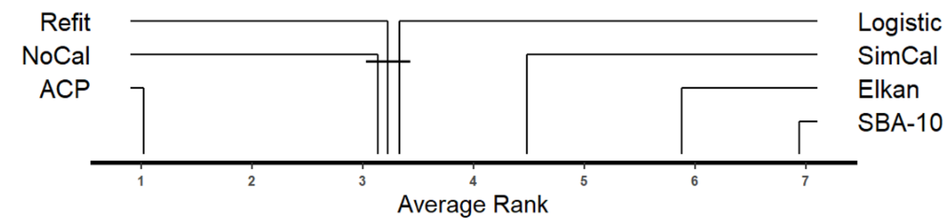


Figure S28: Critical difference plot for the LogLoss in the Cleveland Clinic validation data for random forest. Lower average ranks are considered better. Groups of calibration approaches connected by a horizontal line segment could not be shown to have a significantly different performance. The critical difference was 0.8.

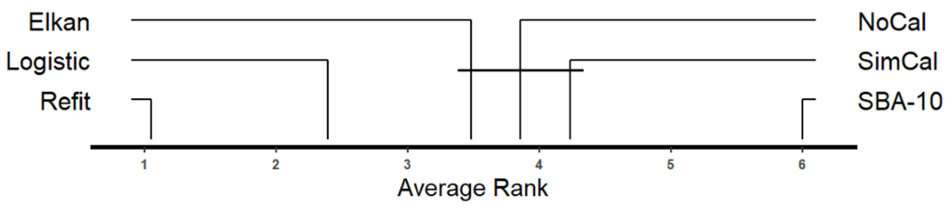


Figure S29: Critical difference plot for the LogLoss in the Cleveland Clinic validation data for gradient boosting. Lower average ranks are considered better. Groups of calibration approaches connected by a horizontal line segment could not be shown to have a significantly different performance. The critical difference was 0.8.

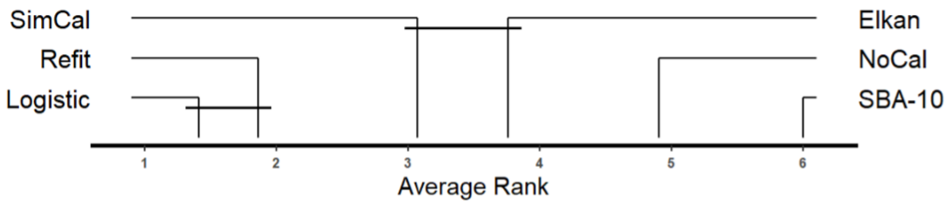

### 5 Histogram of LogLoss difference for the mortality stroke data

Figure S30: Histogram of LogLoss difference for the mortality stroke data based on the LogReg model. Displayed are the frequencies of the LogLoss differences between Refit and NoCal for the 100 bootstrap replicates. Counterintuitively, the LogLosses of NoCal were generally smaller than the LogLosses of Refit. However, the difference was small for all replicates and varied between -0.001 and 0.009.

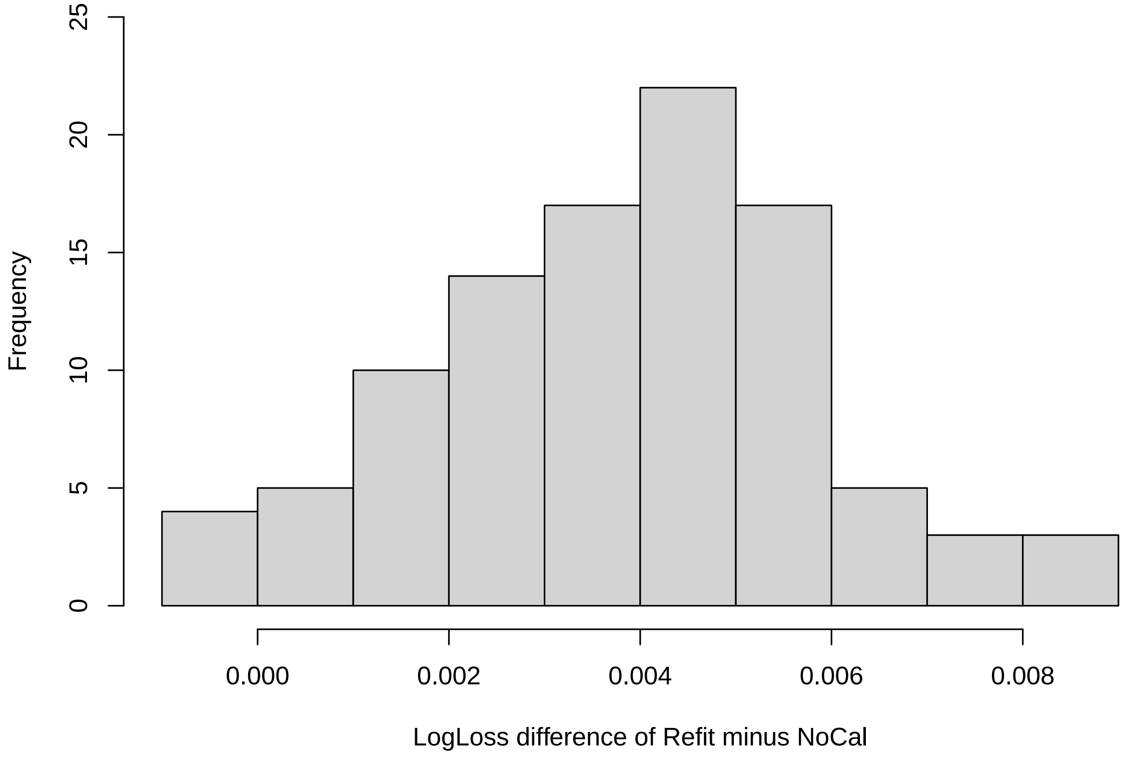
