## Supplementary material for "Calibrating machine learning approaches for probability estimation without calibration data": 01-data-generation-functions.R

```
# Supplementary Material 2 for 
# "Calibrating machine learning approaches for probability
# estimation in case of the absence of calibration data"


# Functions used for data generation of simulated data. 
# ==============================================================================
#
# Contents
# ==============================================================================

# 01) simulate_data
# 02) simulate_data_wrapper

################
#              #
#      ##      #
#      ##      #
#      ##      #
#   ########   #
#    ######    #
#     ###      #
#      #       #
#              #
################

# ==============================================================================
# 01) Simulates data for a given scenario.
#     
#     This function is a modified version of the function 
#     'simulateData' in the R script 
#     'functions-for-calibrating-random-forests.R' provided in 
#     the supplementary material of 
#     Dankowski, T., & Ziegler, A. (2016). Calibrating random forests for 
#     probability estimation. Statistics in medicine, 35(22), 3949-3960.
#
#     This function has to be called twice for each simulation scenario; once 
#     for the model building step and once for the calibration step. The 
#     returned objects data_train and data_test are generated using the 
#     same data generating mechanism.
#      
#     ARGUMENTS
#
#     intercept: intercept of logistic model used for data generation. Ignored
#               for those simulations scenarios using the Mease model.
#     scenario:  simulation scenario number.
#     mean_norm1, sd_norm1, 
#     mean_norm2, sd_norm2: mean and standard deviation for normal distributions 
#                           used to generate the continuous 
#                           predictors for those scenarios based on a logistic 
#                           model using two continuous predictors.
#     modify_mease: if TRUE the base rate of the data generated under the Mease 
#                   model is changed to 1.7 the original rate via Elkan’s 
#                   formula.
#     coeff: covariate coefficients used for data generation under logistic 
#            model (the intercept is specified separately). Ignored for those 
#            simulation scenarios using the Mease model.
#     size_train, size_test: sample size for training and test data. Observe 
#                            that the two datasets are generated using the same
#                            data generating mechanism. 
#     number_noise_variables: number of noise variables to be added to the 
#                             generated data. If greater than zero and the 
#                             scenario is not one of 2, 3, 4, 6, 7, 8, 10, 11, 
#                             12, 22, 23 an error is produced.
#     seed: random seed.
#
#     VALUE
#     
#     A list with the following components:
#     data_train: data generated under logistic or Mease model
#     data_test: data generated under same data generating mechanism as 
#                data_train
#     probs_test: true probabilities for dichotomous endpoint in data_test.
#     
#      
# ==============================================================================

simulate_data <- function(intercept, scenario, mean_norm1 = 0, sd_norm1 = 1,
                          mean_norm2 = mean_norm1, sd_norm2 = sd_norm1,
                          modify_mease = FALSE,
                          coeff, size_train, size_test = 0, 
                          number_noise_variables = 0,  
                          transform_probs = FALSE, seed = NULL){  
  
  if(!is.null(seed)) {
    set.seed(seed)
    print("Setting seed in simulate_data")  
  }
  
  # Mease model scenarios
  scenarios_mease <- c(13, 15)
  # Scenarios where y is generating using two predictors (except Mease model)
  scenarios_two_predictors <- c(1:8, 14, 22, 23)
  # Scenarios where y is generated using ten predictors (5 cont.,  5 cat.) 
  # editing out non binary cat. predictors
  scenarios_ten_predictors <- 9:12
  # Scenarios where y is generated using one predictor
  scenarios_one_predictor <- 16:21
  
  n <- size_train + size_test
  
  if(size_train < 0 | size_test < 0) stop("At least one sample size < 0")
  
  ## initialize independent variables for different simulation scenarios
  if(scenario %in% scenarios_two_predictors) {
    
    if(length(coeff) != 2) stop(paste("For scenario", scenario, 
                                      "coeff should have length 2.")) 
    
    x1 <- rnorm(n, mean = mean_norm1, sd = sd_norm1)
    x2 <- rnorm(n, mean = mean_norm2, sd = sd_norm2)
    
    data_eff <- cbind(x1, x2)
  }
  if(scenario %in% scenarios_mease) { 
    # Mease model covariates
    x1 <- runif(n, min = 0, max = 50)
    x2 <- runif(n, min = 0, max = 50)
    
    data_eff <- cbind(x1, x2)
  }
  if(scenario %in% scenarios_ten_predictors) {
    x1 <- rnorm(n, mean = 0, sd = 1)
    x2 <- rnorm(n, mean = 0, sd = 1)
    x3 <- rnorm(n, mean = 0, sd = 1)
    x4 <- rnorm(n, mean = 0, sd = 1)
    x5 <- rnorm(n, mean = 0, sd = 1)
    x6 <- base::sample(c(0, 1), n, replace = TRUE)
    x7 <- base::sample(c(0, 1), n, replace = TRUE)
    # x8 <- base::sample(c(1, 2, 3), n, replace = TRUE)
    # x9 <- base::sample(c(1, 2, 3, 4), n, replace = TRUE)
    # x10 <- base::sample(c(1, 2, 3, 4, 5), n, replace = TRUE)
    
    data_eff <- cbind(x1, x2, x3, x4, x5, x6, x7)#, x8, x9, x10)
  }
  if(scenario %in% scenarios_one_predictor) {
    x1 <- rnorm(n, mean = mean_norm1, sd = sd_norm1)
    
    data_eff <- cbind(x1)
  }
  
  # calculate probabilities for y=1 and assign values for y according to these 
  # probabilities
  
  if(scenario %in% scenarios_mease) {
    # Mease model
    r <- function(x) sqrt(sum((x - c(25, 25))^2))
    
    r_vector <- apply(data_eff, 1, r)
    
    prob <- rep(0, n)
    
    v <- which(r_vector < 8)
    if(length(v) > 0) prob[v] <- 1
    v <- which(r_vector >= 8 & r_vector <= 28)
    if(length(v) > 0) prob[v] <- (28 - r_vector[v]) / 20
    rm(v)
    
    if(modify_mease) {
      br1 <- 0.4442        # base rate Mease model 
      br2 <- br1 * 1.7     # new data base rate
      
      # due to numerical errors Elkan's formula can produce calibrated 
      # probabilities slightly larger than 1 when the original probability is 1
      v1 <- which(prob == 1); v0 <- which(prob == 0)
      
      # Using Elkan formula
      prob <- br2 * (prob - prob * br1) / (br1 - prob * br1 + br2 * prob - 
                                             br1 * br2)
      if(length(v1) > 0) prob[v1] <- 1
      if(length(v0) > 0) prob[v0] <- 0
      rm(v1, v0)
    }
    
  } else {
    # calculate linear predictor L
    
    if(!ncol(data_eff) == length(coeff)) {
      stop("Dimension of true predictors and coefficients do not match")
    }
    L <- intercept + data_eff %*% coeff  
    prob <- 1 / (1 + exp(-L))
    
    if(transform_probs & scenario %in% 5:8) {
      prob <- prob^3
    }
  } 
  y <- rbinom(n, size = 1, prob = prob)
  
  if(number_noise_variables > 0) {
    
    if(!scenario %in% c(2:4, 6:8, 10:12, 22:23)) {
      stop(paste("Noise variables are not needed in scenario", scenario))
    }
    
    sd_noise <- 1
    if(scenario %in% scenarios_mease) sd_noise <- sqrt(210)
    
    noise <- matrix(rnorm(number_noise_variables * n, mean = 0, sd = sd_noise), 
                    ncol = number_noise_variables, nrow = n, 
                    byrow = FALSE)
    df <- data.frame(y, data_eff, noise)
  } else {
    df <- data.frame(y, data_eff)
  }
  
  # split data into training and test data
  df_train <- df[1:size_train, ]
  
  if(size_test == 0){
    new_list <- list(train = df_train, test = NULL, probs_test = NULL)
  } else{
    indices_test <- (size_train + 1):(size_test + size_train)
    new_list <- list(train = df_train, test = df[indices_test, ],
                     probs_test = prob[indices_test])
  }
  new_list    
}


# ==============================================================================
# 02) Generates data for a given simulation scenario.
#
#     Wrapper of 'simulate_data' function that generates the data for a given 
#     simulation scenario. This function calls 'simulate_data' twice: once 
#     for the model building step and once for the calibration step.
#
#     ARGUMENTS
#
#     scenario:  simulation scenario number.
#     params_df: data frame containing the parameters to be passed to the 
#                'simulate_data' for each simulation scenario. 
#     repn:      Number of current replication to be generated for the given 
#                simulation scenario. 
#
#     VALUE
#     
#     A list with the following components:
#     scenario: simulation scenario number.
#     train: a list with containing two data frames generated under the data 
#            generating mechanism for population 1 (which is used for model 
#            building). 
#     test: a list with containing two data frames generated under the data 
#            generating mechanism for population 2.The first one 'data_train' 
#            will be used to recalibrate the machines and the second one 
#            'data_test' to test the recalibration. 
#     params_df: the parameters used for the data generation for the given 
#                scenario.
#     repn: Number of current replication that was generated for the given 
#                simulation scenario. 
# ==============================================================================

simulate_data_wrapper <- function(scenario, params_df, repn) {
  
  params <- params_df[params_df[["scenario"]] == scenario, ]
  if(nrow(params) != 1) stop("More than one row from params_df chosen")
  
  
  args <- list(intercept = params[["intercept_train"]],
               scenario = params[["scenario"]],
               mean_norm1 = params[["mean_norm1_train"]],
               sd_norm1 = params[["sd_norm1_train"]],
               mean_norm2 = params[["mean_norm2_train"]],
               sd_norm2 = params[["sd_norm2_train"]],
               modify_mease = FALSE,
               coeff = params[["coeff_train"]][[1]],
               size_train = params[["num_train"]],
               size_test = params[["num_train"]],
               number_noise_variables = params[["number_noise_variables"]],
               transform_probs = TRUE,
               seed = NULL)
  
  
  sim_data_fun <- function(args, stage = c("build", "cal", "cal-alt")) {
    sim_data <- do.call("simulate_data", args)
    for(data_name in c("train", "test")) {
      sim_data[[data_name]][["y_fac"]] <- factor(sim_data[[data_name]][["y"]])
      sim_data[[data_name]][["repn"]] <- repn
      data_desc <- paste0(stage, "-", data_name)
      sim_data[[data_name]][["data_desc"]] <- data_desc
      rm(data_desc)
    }
    sim_data[["data_test"]][["prob_true"]] <- sim_data[["probs_test"]]
    sim_data[which(names(sim_data) == "probs_test")] <- NULL 
    sim_data
  }
  
  sim_build <- sim_data_fun(args, "build")
  
  args[["coeff"]] <- params[["coeff_test"]][[1]]
  args[["intercept"]] <- params[["intercept_test"]]
  args[["mean_norm1"]] <- params[["mean_norm1_test"]]
  args[["sd_norm1"]] <- params[["sd_norm1_test"]]
  args[["mean_norm2"]] <- params[["mean_norm2_test"]]
  args[["sd_norm2"]] <- params[["sd_norm2_test"]]  
  args[["modify_mease"]] <- TRUE
  args[["size_train"]] <- params[["num_test1"]]
  args[["size_test"]] <- params[["num_test2"]]
  args[["transform_probs"]] <- FALSE
  
  sim_cal <- sim_data_fun(args, "cal")
  
  # Alternative sample size for cal-train . Sample size is doubled.
  # Cal-test data is not changed (to allow for comparisons)
  args[["size_train"]] <- 2 * args[["size_train"]]
  sim_cal_alt <- sim_data_fun(args, "cal-alt")
  sim_cal_alt[["test"]] <- sim_cal[["test"]]
  sim_cal_alt[["test"]][["data_desc"]] <- "cal-alt-test"
  
  sim_cal_alt[["data_test"]] <- sim_cal[["data_test"]]
  
  list(scenario = scenario, build = sim_build, cal = sim_cal,
       cal_alt = sim_cal_alt, params_df = params, repn = repn)
  
}
```
