## Supplementary material for "Calibrating machine learning approaches for probability estimation without calibration data": 02-machine-fitting-functions.R

```
# Supplementary Material 2 for 
# "Calibrating machine learning approaches for probability
# estimation in case of the absence of calibration data"


# Functions used for machine fitting and computation of predicted probabilities
# (without recalibration) during simulation.
# ==============================================================================

# Contents
# ==============================================================================

# 01) add_empty_prob_columns
# 02) create_formula
# 03) create_grids_and_workflows
# 04) extract_train_test_dfs
# 05) tuning_machines
# 06) fitting_machines
# 07) add_predictions
# 08) update_train_test_dfs

################
#              #
#      ##      #
#      ##      #
#      ##      #
#   ########   #
#    ######    #
#     ###      #
#      #       #
#              #
################

# 01) add_empty_prob_columns
# 02) create_formula
# 03) create_grids_and_workflows
# 04) extract_train_test_dfs
# 05) tuning_machines
# 06) fitting_machines
# 07) add_predictions
# 08) update_train_test_dfs

################
#              #
#      ##      #
#      ##      #
#      ##      #
#   ########   #
#    ######    #
#     ###      #
#      #       #
#              #
################


# ==============================================================================
# 01) Adds (empty) probability columns to simulated datasets.
#
#     This function has no arguments and returns NULL. It initializes the 
#     probability columns in the simulated data frames (with NAs). These empty 
#     columns will contain the predicted probabilities from the 
#     different machines (but not the ones that will be re-calibrated, only 
#     those that are obtained directly from the machines). 
#     The following objects are taken from the environment where the function
#     is called: 'simu_data' and 'machine_abbreviations'.
#     The function updates (overwrites) 'simu_data' in the environment
#     where the function is called.
# ==============================================================================

add_empty_prob_columns <- function() {
  
  objects_to_get <- c("simu_data", "machine_abbreviations")
  for(i in objects_to_get) assign(i, get(i, envir = parent.frame())) 
  
  for(machine_abbr in machine_abbreviations) {
    prob_col <- paste0("prob_", machine_abbr)
    prob_col_refit <- paste0(prob_col, "_refit")
    
    simu_data[["build"]][["test"]][[prob_col]] <- NA_real_
    
    for(cal_name in c("cal", "cal_alt")) {
      simu_data[[cal_name]][["train"]][[prob_col]] <- NA_real_
      simu_data[[cal_name]][["test"]][[prob_col]] <- NA_real_
      simu_data[[cal_name]][["test"]][[prob_col_refit]] <- NA_real_      
    }
    
    rm(prob_col, prob_col_refit)
  }
  
  assign("simu_data", simu_data, envir = parent.frame())
  
  invisible(NULL)
}


# ==============================================================================
# 02) Creates formula for machines.
#     
#     This function has no arguments and returns NULL. It creates the formula 
#     that will be used by the different machines. 'simu_data' is taken from 
#     the environment where the function is called. The function creates a 
#     object termed 'Formula' in the environment where the function is called.
#
#     The predictors use to generate the data are denoted by x1, x2, x3, ....
#     Noise is denoted by X1, X2, X3, ....
# ==============================================================================

create_formula <- function() {
  simu_data <- get("simu_data", envir = parent.frame())
  predictors <- names(simu_data[["build"]][["train"]])
  predictors <- predictors[str_detect(predictors, "^(x|X)")]
  
  Formula <- paste0("y_fac ~ ", paste(predictors, collapse = "+")) %>%
    as.formula 
  assign("Formula", Formula, envir = parent.frame())
  invisible(NULL)
}

# ==============================================================================
# 03) Creates tidymodels tuning grids and workflows.
#
#     This function has no arguments and returns NULL. It gets the following
#     objects from the environment the function was called: 'simu_data', 
#     'Formula', 'machine_abbreviations_tune', 'grid_size'.
#     All created grids and workflows are added to the environment where the
#     function was called.
# ==============================================================================

create_grids_and_workflows <- function() {
  
  objects_to_get <- c("simu_data", "Formula", 
                      "machine_abbreviations_tune", "grid_size")
  for(i in objects_to_get) assign(i, get(i, envir = parent.frame()))
  objects_to_get <- paste0("spec_", machine_abbreviations_tune)
  for(i in objects_to_get) assign(i, get(i, envir = parent.frame()))
  rm(i)
  
  vars <- all.vars(Formula)[-1] # these are the predictors
  
  for(i in c("build", "cal", "cal_alt")) {
    
    df_current <- simu_data[[i]][["train"]]
    
    v <- which(names(df_current) == "y_fac")
    if(length(v) != 1) stop("y_fac not found")
    rm(v)
    
    # elastic net grid
    
    if("en" %in% machine_abbreviations_tune){
      grid_en <- dials::grid_latin_hypercube(
        dials::penalty(), mixture(),
        size = grid_size)  
      assign(paste0("grid_en_", i), grid_en, envir = parent.frame())
      rm(grid_en)
    }
    # random forest grid. Adding the parameters used in the 
    # Dankowski, T., & Ziegler, A. (2016) paper to the grid (if not already 
    # included).
    
    grid_rf <- dials::grid_latin_hypercube(
      finalize(dials::mtry(), 
               x = df_current[, vars, drop = FALSE]),
      dials::min_n(),
      size = grid_size
    )
    grid_aux <- grid_rf[1, ]
    grid_aux[["mtry"]] <- floor(sqrt(ncol(df_current[, vars, 
                                                     drop = FALSE])))
    grid_aux[["min_n"]] <- floor(nrow(df_current) * 0.1)
    grid_rf <- rbind(grid_rf, grid_aux)
    grid_rf <- grid_rf[!duplicated(grid_rf), ]
    rm(grid_aux)
    
    assign(paste0("grid_rf_", i), grid_rf, envir = parent.frame())
    
    # xgboost grid
    
    grid_boost <- dials::grid_latin_hypercube(
      tree_depth(), trees(), 
      learn_rate(), loss_reduction(),
      finalize(dials::mtry(), 
               x = df_current[, vars, drop = FALSE]), 
      sample_size = sample_prop(),
      min_n(),
      size = grid_size)
    
    assign(paste0("grid_boost_", i), grid_boost, envir = parent.frame())   
    
    # svm grid
    
    grid_svm <- 
      dials::grid_latin_hypercube(
        cost(),
        rbf_sigma(),
        size = grid_size
      )
    assign(paste0("grid_svm_", i), grid_svm, envir = parent.frame())
    
    # Creating workflows
    
    for(j in machine_abbreviations_tune) {
      machine_spec <- get(paste0("spec_", j))
      wf <- workflows::workflow() %>%
        add_model(machine_spec) %>% 
        add_formula(Formula)
      
      assign(paste0("wf_", j,  "_", i), wf, envir = parent.frame())
    }
    rm(j, grid_rf, grid_boost, grid_svm)
  }
  
  invisible(NULL)  
}  


# ==============================================================================
# 04) Extracts training and test data frames.
#
#     This function has no arguments and returns NULL. It gets the following
#     objects from the environment the function was called: 'simu_data'.
#     The function extracts from 'simu_data' the different data 
#     frames used for model building and calibration. The data frames are then 
#     added to the environment where the function was called so that they
#     can be passed later to the machine fitting routines.
# ==============================================================================

extract_train_test_dfs <- function() {
  simu_data <- get("simu_data", envir = parent.frame())
  
  for(i in c("build", "cal", "cal_alt")) {
    for(j in c("train", "test")) {
      df_current <- simu_data[[i]][[j]]
      assign(paste0("df_", i, "_", j), df_current, envir = parent.frame())
      rm(df_current)
    }
  }
  invisible(NULL)
}

# ==============================================================================
# 05) Tunes machines. 
#
#     This function has no arguments and returns NULL. It gets the tuning grids,
#     workflows, grids and list of machines to tune from the environment the 
#     function is called. The best machine is chosen according 
#     to the LogLoss and the workflow is finalized. The finalized workflow is 
#     is added to the environment where the function is called.
# ==============================================================================

tuning_machines <- function() {
  
  objects_to_get <- c("machine_abbreviations_tune")
  for(i in objects_to_get) assign(i, get(i, envir = parent.frame()))  
  rm(i)
  
  for(i in c("build", "cal", "cal_alt")) {
    
    folds_current <- get(paste0("folds_", i), envir = parent.frame())
    for(machine_abbr in machine_abbreviations_tune) {
      wf_current <- get(paste0("wf_", machine_abbr, "_", i), 
                        envir = parent.frame())
      grid_current <- get(paste0("grid_", machine_abbr, "_", i), 
                          envir = parent.frame())
      machine_tuned <- tune::tune_grid(
        object = wf_current,
        resamples = folds_current,
        grid = grid_current,
        metrics = metric_set(mn_log_loss),
        control = tune::control_grid(verbose = TRUE, allow_par = FALSE))
      
      best_log_loss <- select_best(machine_tuned, "mn_log_loss")
      machine_tuned_final <- finalize_workflow(wf_current, best_log_loss)
      
      
      assign(paste0("tuned_", machine_abbr, "_", i), machine_tuned, 
             envir = parent.frame())
      assign(paste0("tuned_final_", machine_abbr, "_", i), machine_tuned_final, 
             envir = parent.frame())
      
    }
  }
  
  invisible(NULL) 
}

# ==============================================================================
# 06) Fits machines. 
#
#     This function has no arguments and returns NULL. This function gets the 
#     list of machines (including those that require tuning), finalized 
#     workflows, formula, and simulated datasets and fits the machines. No 
#     tuning is performed here (this was taken care by 'tuning_machines' above).
#     The fits are added to the environment where the function is called.
#
#     During code testing an error was encountered once, which suggests that the 
#     error does not surface that often, when trying to make 
#     probability predictions from an svm (package kernlab). The error could be
#     traced to kernlab:::.SigmoidPredict. Upon closer examination it was 
#     observed that the fit could predict classes but not probabilities. 
#
#     This seems to be a known issue:
#
#     https://github.com/topepo/caret/issues/809
#
#     Refitting the svm model (without repeating the tuning) solved the issue
#     in our example. For that reason a loop was added to the code to test
#     if probabilities can be computed with the fitted svm. If this fails
#     the fit is repeated. 
#
# ==============================================================================

fitting_machines <- function() {
  
  objects_to_get <- c("machine_abbr", "machine_abbreviations_tune", 
                      "df_build_train", "df_cal_train", 
                      "df_cal_alt_train", "Formula")
  for(i in objects_to_get) assign(i, get(i, envir = parent.frame()))  
  rm(i)
  
  for(i in c("build", "cal", "cal_alt")) {
    df_current <- get(paste0("df_", i, "_train"))
    if(machine_abbr %in% machine_abbreviations_tune) {
      machine_specification <- get(paste0("tuned_final_", machine_abbr, "_", i), 
                                   envir = parent.frame())
      fit <- machine_specification %>% fit(data = df_current)
      
      if(machine_abbr == "svm") {
        
        svm_fit_index <- 1
        pred_test <- tryCatch(predict(fit, new_data = df_current[1:5, ], 
                                      type = "prob")[[".pred_1"]], 
                              error = identity) 
        
        while(svm_fit_index <= 20 & "simpleError" %in% class(pred_test)) {
          fit <- machine_specification %>% fit(data = df_current)
          pred_test <- tryCatch(predict(fit, new_data = df_current[1:5, ], 
                                        type = "prob")[[".pred_1"]], 
                                error = identity)
          svm_fit_index <- svm_fit_index + 1
        }
        rm(pred_test)
      }
      
    } else {
      machine_specification <- get(paste0("spec_", machine_abbr))
      fit <- machine_specification %>% fit(Formula, data = df_current)
    }
    assign(paste0("fit_", i), fit, envir = parent.frame())
    
    rm(fit, df_current)
  }
  
  invisible(NULL) 
}

# ==============================================================================
# 07) Populates probability columns.
# 
#     This function has no arguments and returns NULL. For each machine it adds
#     the estimated predicted probabilities to the data frames used during the
#     simulation. It gets the list of machines, fits and data frames from the 
#     environment where the function is called and updates the data frames in
#     that environment. Only those predicted probabilities that can be obtained
#     directly from the machines (i.e., without re-calibration) are added.
# ==============================================================================

add_predictions <- function() {
  
  objects_to_get <- c("machine_abbr", "fit_build", "fit_cal", "fit_cal_alt")
  for(i in objects_to_get) assign(i, get(i, envir = parent.frame()))  
  
  prob_col <- paste0("prob_", machine_abbr)
  
  for(data_name in c("df_build_test",  "df_cal_train", "df_cal_test",
                     "df_cal_alt_train", "df_cal_alt_test")) {
    df_current <- get(data_name, envir = parent.frame())
    
    aux <- tryCatch(predict(fit_build, 
                            new_data = df_current, 
                            type = "prob")[[".pred_1"]], error = identity)
    
    if("simpleError" %in% class(aux)) aux <- rep(NA_real_, nrow(df_current))
    
    df_current[[prob_col]] <- aux; rm(aux)
    
    if(data_name == "df_cal_test") {
      aux <- tryCatch(predict(fit_cal, new_data = df_current, 
                              type = "prob")[[".pred_1"]], error = identity)
      
      if("simpleError" %in% class(aux)) aux <- rep(NA_real_, nrow(df_current))
      
      df_current[[paste0(prob_col, "_refit")]] <- aux; rm(aux)      
    }
    
    if(data_name == "df_cal_alt_test") {
      aux <-tryCatch(predict(fit_cal_alt, new_data = df_current, 
                             type = "prob")[[".pred_1"]], error = identity)
      
      if("simpleError" %in% class(aux)) aux <- rep(NA_real_, nrow(df_current))
      
      df_current[[paste0(prob_col, "_refit")]] <- aux; rm(aux)      
    }    
    
    assign(data_name, df_current, envir = parent.frame())
    rm(df_current)
  }
  
  invisible(NULL)  
  
}

# ==============================================================================
# 08) Updates simulated data frames.
#
#     This function has no arguments and returns NULL. This function gets
#     the 'simu_data' object from the environment where the function is called
#     and the different data frames containing the probabilities from the fitted
#     machines and updates this information in the 'simu_data' object.
# ==============================================================================

update_train_test_dfs <- function() {
  
  simu_data <- get("simu_data", envir = parent.frame())
  
  for(i in c("build", "cal", "cal_alt")) {
    for(j in c("train", "test")) {
      df_current <- get(paste0("df_", i, "_", j), 
                        envir = parent.frame())
      simu_data[[i]][[j]] <- df_current
      rm(df_current)
    }
  }
  assign("simu_data", simu_data, envir = parent.frame())
  
  invisible(NULL)
}
```
