## Supplementary material for "Calibrating machine learning approaches for probability estimation without calibration data": 03-calibration-functions.R

```
# Supplementary Material 2 for 
# "Calibrating machine learning approaches for probability
# estimation in case of the absence of calibration data"
#
# This script is a modified version of the one found in supplementary material 
# 2 of Ojeda FM, et al., Calibrating machine learning approaches for probability 
# estimation: A comprehensive comparison. Stat Med. 2023 Dec 20;42(29):5451-5478

# Functions used for calibration of predictions of simulated data. 
# ==============================================================================
#
# Contents
# ==============================================================================

# 01) add_empty_cal_prob_columns
# 02) calibrate_preds
# 03) update_cal_prob_columns
# 04) winsorize_probs
# 05) calibrate_elkan
# 06) calibrate_logistic
# 07) calibrate_intercept
# 08) calibrate_slope
# 09) calibrate_beta 
# 10) calibrate_isotonic
# 11) calibrate_platt
# 12) calibrate_chen
# 13) calibrate_rcs
# 14) calibrate_simcal
# 15) create_prev_epv_rcs_matrices
# 16) compute_prev_epv_and_update_matrix
# 17) calibrate_simcal_mispec
# 18) calibrate_elkan_mispec
# 19) calibrate_sba10
# 20) calibrate_acp


################
#              #
#      ##      #
#      ##      #
#      ##      #
#   ########   #
#    ######    #
#     ###      #
#      #       #
#              #
################

# ==============================================================================
# 01) Adds (empty) probability columns for calibrated probabilities and true 
#     probabilities column.
#
#     This function has no arguments and returns NULL. This function gets the 
#     following objects from the environment it is called: 'results', 
#     'machine_abbreviations', 'calibration_names', 'repns'.
#     'results' contains the simulation results before re-calibration. This 
#     function adds empty (i.e., filled with NA) columns for each combination of
#     re-calibration method and machine to 'results' (this columns will contain 
#     later the re-calibrated probabilities). Then this object is 
#     updated in the environment where the function is called.
# ==============================================================================

add_empty_cal_prob_columns <- function() {
  
  objects_to_get <- c("results", "machine_abbreviations", "calibration_names",
                      "repns")
  for(i in objects_to_get) assign(i, get(i, envir = parent.frame())) 
  
  FUN <- function(x) paste0(x, collapse = "")
  newcols <- expand.grid("prob_", machine_abbreviations, "_", 
                         calibration_names, "_cal",
                         stringsAsFactors = FALSE) %>% as.matrix() %>% 
    apply(1, FUN) 
  
  
  for(i in repns) {
    results[["data_list"]][[i]][["cal"]][["test"]][, newcols] <- NA_real_
    results[["data_list"]][[i]][["cal_alt"]][["test"]][, newcols] <- NA_real_
    
    p_true <- results[["data_list"]][[i]][["cal"]][["data_test"]][["prob_true"]]
    results[["data_list"]][[i]][["cal"]][["test"]][, "prob_true"] <- p_true
    rm(p_true)
    
    p_true <- results[["data_list"]][[i]][["cal_alt"]][["data_test"]][["prob_true"]]
    results[["data_list"]][[i]][["cal_alt"]][["test"]][, "prob_true"] <- p_true
    rm(p_true)
    
    p_true <- results[["data_list"]][[i]][["build"]][["data_test"]][["prob_true"]]
    results[["data_list"]][[i]][["build"]][["test"]][, "prob_true"] <- p_true
    rm(p_true)    
    
  }
  assign("results", results, envir = parent.frame())
  
  invisible(NULL)
}


# ==============================================================================
# 02) Re-calibrates predicted probabilities.
#
#     This function has no arguments and returns NULL. This function gets
#     the different data frames produced during the simulation (model 
#     building, cal-training and cal-test) and loops
#     over the different re-calibration methods for a given machine 
#     ('machine_abbr') adding the re-calibrated probabilities to the cal-test 
#     data frame. The function gets the required objects from the environment 
#     where it is called and updates the cal-test and alternative cal-test 
#     data frames there.
# ==============================================================================

calibrate_preds <- function() {
  objects_to_get <- c("machine_abbr", "calibration_names", 
                      "df_build_train", "df_cal_train", "df_cal_test",
                      "df_cal_alt_train", "df_cal_alt_test", "repn_number")
  for(i in objects_to_get) assign(i, get(i, envir = parent.frame()))
  
  assign("rcs_info_cal", get(paste0("rcs_info_cal_", machine_abbr), 
                             envir = parent.frame()))
  assign("rcs_info_cal_alt", get(paste0("rcs_info_cal_alt_", machine_abbr), 
                                 envir = parent.frame()))
  
  assign("chen_info_cal", get(paste0("chen_info_cal_", machine_abbr), 
                              envir = parent.frame()))
  assign("chen_info_cal_alt", get(paste0("chen_info_cal_alt_", machine_abbr), 
                                  envir = parent.frame()))  
  
  for(calibration_name in calibration_names) {
    
    calibrate_fun <- get(paste0("calibrate_", calibration_name), 
                         envir = parent.frame())
    prob <-  paste0("prob_", machine_abbr)
    newcol <- paste0(prob, "_", calibration_name, "_cal")
    
    cal_version <- "cal"
    df_cal_test[[newcol]] <- calibrate_fun(response = "y", 
                                           data = df_build_train, 
                                           newdata1 = df_cal_train, 
                                           newdata2 = df_cal_test, 
                                           prob = prob,
                                           mispec = mispec)
    
    cal_version <- "cal_alt"
    df_cal_alt_test[[newcol]] <- calibrate_fun(response = "y", 
                                               data = df_build_train, 
                                               newdata1 = df_cal_alt_train, 
                                               newdata2 = df_cal_alt_test, 
                                               prob = prob,
                                               mispec = mispec)    
    
  }
  
  assign(paste0("rcs_info_cal_", machine_abbr), rcs_info_cal, 
         envir = parent.frame())
  assign(paste0("rcs_info_cal_alt_", machine_abbr), rcs_info_cal_alt, 
         envir = parent.frame())  
  
  assign(paste0("chen_info_cal_", machine_abbr), chen_info_cal, 
         envir = parent.frame())
  assign(paste0("chen_info_cal_alt_", machine_abbr), chen_info_cal_alt, 
         envir = parent.frame())    
  
  assign("df_cal_test", df_cal_test, envir = parent.frame())
  assign("df_cal_alt_test", df_cal_alt_test, envir = parent.frame())  
  
  invisible(NULL)  
  
}

# ==============================================================================
# 03) Updates simulation results object with re-calibrated probabilities.
#      
#     This function has no arguments and returns NULL. After probabilities are 
#     re-calibrated updates 'results' object (containing the simulation results)
#     in the environment the function is called. Removes data frames with 
#     simulated data used during the re-calibration from environment where the
#     function was called.
# ==============================================================================

update_cal_prob_columns <- function() {
  
  objects_to_get <- c("results", "df_cal_test", "df_cal_alt_test", 
                      "repn_number")
  
  for(i in objects_to_get) assign(i, get(i, envir = parent.frame()))
  
  
  results[["data_list"]][[repn_number]][["cal"]][["test"]] <- df_cal_test
  results[["data_list"]][[repn_number]][["cal_alt"]][["test"]] <- df_cal_alt_test
  
  assign("results", results, envir = parent.frame())
  rm(list = c("df_build_train", "df_build_test", 
              "df_cal_train", "df_cal_test", 
              "df_cal_alt_train", "df_cal_alt_test"), 
     envir = parent.frame())
  
  invisible(NULL)  
}

# ==============================================================================
# 04) Winsorizes a vector of probabilities. 
#
#     The approach used to winsorize a vector of probabilities is taken from
#     yardstick:::mn_log_loss_multiclass. Which is how that package deals with
#     probabilities close to 0 or 1 when working with the LogLoss.
#
#     ARGUMENTS
#     
#     x: a numeric vector (of probabilities)
#
#     VALUE
#
#     The numeric vector x after winsorization.
#
# ==============================================================================

winsorize_probs <- function(x) {
  eps <- .Machine$double.eps
  pmax(pmin(x, 1 - eps), eps)
}

# ==============================================================================
# 05) Calibrates probabilities using Elkan approach.
#     
#     Calibrates a vector of probabilities using Elkan general updating 
#     approach: 
#     Elkan C. The foundations of cost-sensitive learning. In: Nebel B., ed. 
#     Proceedings of the Seventeenth International Joint Conference on 
#     Artificial Intelligence. Vol. 2. San Francisco: Morgan Kaufmann. 2001 
#     (pp. 973–978).
#
#     ARGUMENTS
#     
#     response: character string giving the name of the column containing the 
#               binary response in 'data' and 'newdata1'.
#     data: data frame from population 1 used for model building.
#     newdata1: data frame from population 2 used for model recalibration.
#     newdata2: data frame from population 2 used for testing of recalibrated
#               probabilities.
#     prob:     character string column indicating the name of the column in 
#               data frames 'newdata1' and 'newdata2' that will be contain
#               the probabilities that will be recalibrated.
#
#     VALUE
#
#     A numeric vector containing the recalibrated probabilities for 
#     'newdata2'.  
# ==============================================================================
calibrate_elkan <- function(response, data, 
                            newdata1, newdata2, prob, mispec) {
  
  br1 <- mean(data[, response])
  br2 <- mean(newdata1[, response]) # misspecify; replace with misspecified prop or mean
  
  prob2 <- newdata2[, prob]
  
  br2 * (prob2 - prob2 * br1) / (br1 - prob2 * br1 + br2 * prob2 - br1 * br2)
  
}

# ==============================================================================
# 06) Logistic calibration.
#
#     Calibrates a vector of probabilities using logistic calibration (intercept 
#     and slope are estimated).
#
#     ARGUMENTS
#     
#     response: character string giving the name of the column containing the 
#               binary response in 'data' and 'newdata1'.
#     data: data frame from population 1 used for model building.
#     newdata1: data frame from population 2 used for model recalibration.
#     newdata2: data frame from population 2 used for testing of recalibrated
#               probabilities.
#     prob:     character string column indicating the name of the column in 
#               data frames 'newdata1' and 'newdata2' that will be contain
#               the probabilities that will be recalibrated.
#
#     VALUE
#
#     A numeric vector containing the recalibrated probabilities for 
#     'newdata2'.  
# ==============================================================================

calibrate_logistic <- function(response, data, 
                               newdata1, newdata2, prob, mispec) {
  
  prob1 <- newdata1[, prob] %>% winsorize_probs()
  prob2 <- newdata2[, prob] %>% winsorize_probs()
  
  newdata1[["lp"]] <- qlogis(prob1)
  newdata2[["lp"]] <- qlogis(prob2)
  
  Formula <- paste0(response, " ~ lp") %>% as.formula()
  fit <- glm(Formula, data = newdata1, family = binomial())
  
  predict(fit, newdata = newdata2, type = "response")
}


# ==============================================================================
# 07) Intercept calibration.
#
#     Calibrates a vector of probabilities using intercept calibration (only an 
#     intercept is estimated).
#
#     ARGUMENTS
#     
#     response: character string giving the name of the column containing the 
#               binary response in 'data' and 'newdata1'.
#     data: data frame from population 1 used for model building.
#     newdata1: data frame from population 2 used for model recalibration.
#     newdata2: data frame from population 2 used for testing of recalibrated
#               probabilities.
#     prob:     character string column indicating the name of the column in 
#               data frames 'newdata1' and 'newdata2' that will contain
#               the probabilities that will be recalibrated.
#
#     VALUE
#
#     A numeric vector containing the recalibrated probabilities for 
#     'newdata2'.  
# ==============================================================================    

calibrate_intercept <- function(response, data, 
                                newdata1, newdata2, prob) {
  
  prob1 <- newdata1[, prob] %>% winsorize_probs()
  prob2 <- newdata2[, prob] %>% winsorize_probs()
  
  newdata1[["lp"]] <- qlogis(prob1)
  newdata2[["lp"]] <- qlogis(prob2)
  
  Formula <- paste0(response, " ~ offset(lp)") %>% as.formula()
  fit <- glm(Formula, data = newdata1, family = binomial())
  
  predict(fit, newdata = newdata2, type = "response")
}

# ==============================================================================
# 08) Slope calibration. 
#
#     Calibrates a vector of probabilities using slope calibration.
#
#     ARGUMENTS
#     
#     response: character string giving the name of the column containing the 
#               binary response in 'data' and 'newdata1'.
#     data: data frame from population 1 used for model building.
#     newdata1: data frame from population 2 used for model recalibration.
#     newdata2: data frame from population 2 used for testing of recalibrated
#               probabilities.
#     prob:     character string column indicating the name of the column in 
#               data frames 'newdata1' and 'newdata2' that contain
#               the probabilities that will be recalibrated.
#
#     VALUE
#
#     A numeric vector containing the recalibrated probabilities for 
#     'newdata2'.  
# ==============================================================================

calibrate_slope <- function(response, data, 
                            newdata1, newdata2, prob) {
  
  prob1 <- newdata1[, prob] %>% winsorize_probs()
  prob2 <- newdata2[, prob] %>% winsorize_probs()
  
  newdata1[["lp"]] <- qlogis(prob1)
  newdata2[["lp"]] <- qlogis(prob2)
  Formula <- paste0(response, " ~ lp -1 ") %>% as.formula()
  fit <- glm(Formula, data = newdata1, family = binomial())
  
  predict(fit, newdata = newdata2, type = "response")
}

# ==============================================================================
# 09) Beta calibration.
#
#     Calibrates a vector of probabilities using Beta calibration.
#
#     ARGUMENTS
#     
#     response: character string giving the name of the column containing the 
#               binary response in 'data' and 'newdata1'.
#     data: data frame from population 1 used for model building.
#     newdata1: data frame from population 2 used for model recalibration.
#     newdata2: data frame from population 2 used for testing of recalibrated
#               probabilities.
#     prob:     character string column indicating the name of the column in 
#               data frames 'newdata1' and 'newdata2' that contain
#               the probabilities that will be recalibrated.
#
#     VALUE
#
#     A numeric vector containing the recalibrated probabilities for 
#     'newdata2'.  
# ==============================================================================

calibrate_beta <- function(response, data, 
                           newdata1, newdata2, prob, mispec) {
  
  prob1 <- newdata1[, prob] %>% winsorize_probs()
  prob2 <- newdata2[, prob] %>% winsorize_probs() 
  
  newdata1[["lp1"]] <- log(prob1)
  newdata1[["lp2"]] <- -log(1 - prob1)
  
  newdata2[["lp1"]] <- log(prob2)
  newdata2[["lp2"]] <- -log(1 - prob2)
  
  Formula <- paste0(response, " ~ lp1 + lp2") %>% as.formula()
  fit <- glm(Formula, data = newdata1, family = binomial())
  
  predict(fit, newdata = newdata2, type = "response")
  
} 

# ==============================================================================
# 10) Isotonic calibration.
#
#     Calibrates a vector of probabilities using isotonic calibration.
#
#     ARGUMENTS
#     
#     response: character string giving the name of the column containing the 
#               binary response in 'data' and 'newdata1'.
#     data: data frame from population 1 used for model building.
#     newdata1: data frame from population 2 used for model recalibration.
#     newdata2: data frame from population 2 used for testing of recalibrated
#               probabilities.
#     prob:     character string column indicating the name of the column in 
#               data frames 'newdata1' and 'newdata2' that contain
#               the probabilities that will be recalibrated.
#
#     VALUE
#
#     A numeric vector containing the recalibrated probabilities for 
#     'newdata2'.  
# ==============================================================================

calibrate_isotonic <- function(response, data, 
                               newdata1, newdata2, prob) {
  
  fit <- isoreg(x = newdata1[, prob], y = newdata1[, response])
  
  stfun <- as.stepfun(fit)
  
  stfun(newdata2[, prob])
}

# ==============================================================================
# 11) Platt calibration.
#
#     Calibrates a vector of probabilities using Platt calibration.
#
#     ARGUMENTS
#     
#     response: character string giving the name of the column containing the 
#               binary response in 'data' and 'newdata1'.
#     data: data frame from population 1 used for model building.
#     newdata1: data frame from population 2 used for model recalibration.
#     newdata2: data frame from population 2 used for testing of recalibrated
#               probabilities.
#     prob:     character string column indicating the name of the column in 
#               data frames 'newdata1' and 'newdata2' that contain
#               the probabilities that will be recalibrated.
#
#     VALUE
#
#     A numeric vector containing the recalibrated probabilities for 
#     'newdata2'.  
# ==============================================================================

calibrate_platt <- function(response, data, 
                            newdata1, newdata2, prob) {
  
  Formula <- paste0(response, " ~ ", prob) %>% as.formula()
  fit <- glm(Formula, data = newdata1, family = binomial())
  
  predict(fit, newdata = newdata2, type = "response")
  
}  


# ==============================================================================
# 12) Chen calibration.
#
#     Calibrates a vector of probabilities using Chen's general updating 
#     approach. Kernel density estimation is used.
#
#     ARGUMENTS
#     
#     response: character string giving the name of the column containing the 
#               binary response in 'data' and 'newdata1'.
#     data: data frame from population 1 used for model building.
#     newdata1: data frame from population 2 used for model recalibration.
#     newdata2: data frame from population 2 used for testing of recalibrated
#               probabilities.
#     prob:     character string column indicating the name of the column in 
#               data frames 'newdata1' and 'newdata2' that contain
#               the probabilities that will be recalibrated.
#
#     VALUE
#
#     A numeric vector containing the recalibrated probabilities for 
#     'newdata2'. 
#
#     If the estimated densities are both equal to zero at the same point
#     in newdata2 the formula used to computed the calibrated probabilities 
#     returns 0 / 0. In this cases the calibrated probabilities are set to 1.
#
#     This function gets the following objects from the environment it is 
#     called: 'cal_version', 'repn_number' and 'chen_info_[cal_version]'. It 
#     adds information about the number of times 0 / 0 encountered during 
#     calibration for the current 'repn_number' and updates 
#     'chen_info_[cal_version]' in the calling environment.
#
#     Remark: If one wants to use this function outside the simulation to 
#             calibrate predicted probabilities, the lines of code that are used
#             to keep track of the number 0 / 0 encountered should be disabled.
# ==============================================================================

calibrate_chen <- function(response, data, 
                           newdata1, newdata2, prob) {
  
  
  objects_to_get <- c("cal_version", "repn_number")
  for(i in objects_to_get) assign(i, get(i, envir = parent.frame()))  
  rm(i)
  
  assign("chen_info", get(paste0("chen_info_", cal_version), 
                         envir = parent.frame())) 
  
  br2 <- mean(newdata1[, response])
  
  prob1 <- newdata1[, prob] %>% winsorize_probs()
  prob2 <- newdata2[, prob] %>% winsorize_probs()
  
  newdata1[["lp"]] <- qlogis(prob1)
  newdata2[["lp"]] <- qlogis(prob2)  
  
  for(current_value in 0:1) {
    v <- which(newdata1[[response]] == current_value)
    if(length(v) == 0) stop("Response only takes one value")
    fit <- density(newdata1[["lp"]][v], bw = "nrd0")
    
    assign(paste0("dens_fun_", current_value), approxfun(fit, rule = 2))
    rm(fit, v)
  }
  
  lp2 <- newdata2[["lp"]]
  
  f1 <- dens_fun_1(lp2)
  f0 <- dens_fun_0(lp2)
  
  numerator <- f1 * br2 
  denominator <- (numerator + f0 * (1 - br2))
  
  output <- numerator / denominator
  
  pos_zero <- which(f1 == 0 & f0 == 0)
  if(length(pos_zero) > 0) {
    output[pos_zero] <- 1
  }
  
  chen_info_row <- data.frame(n_cal_train = nrow(newdata1), 
                              n_events_cal_train = sum(newdata1[["y"]]),
                              n_cal_test = nrow(newdata2), 
                              n_events_cal_test = sum(newdata2[["y"]]), 
                              n_zero_over_zero_cal_test = length(pos_zero))  
  
  chen_info[repn_number, ] <- chen_info_row
  assign(paste0("chen_info_", cal_version), chen_info, envir = parent.frame())
  
  output
} 


# ==============================================================================
# 13) Rcs calibration.
#
#     Calibrates a vector of probabilities using a logistic regression with
#     with the logit of the probabilities as the only predictor using 
#     restricted cubic splines.
#
#     ARGUMENTS
#     
#     response: character string giving the name of the column containing the 
#               binary response in 'data' and 'newdata1'.
#     data: data frame from population 1 used for model building.
#     newdata1: data frame from population 2 used for model re-calibration.
#     newdata2: data frame from population 2 used for testing of re-calibrated
#               probabilities.
#     prob:     character string column indicating the name of the column in 
#               data frames 'newdata1' and 'newdata2' that contain
#               the probabilities that will be re-calibrated.
#
#     VALUE
#
#     A numeric vector containing the re-calibrated probabilities for 
#     'newdata2'.
#
#
#     The following text is taken from Harrell "Regression modelling strategies"
#     'When the sample size is large (e.g., n ≥ 100 with a continuous uncensored 
#      response variable), k = 5 is a good choice. Small samples (< 30, say) may 
#      require the use of k = 3.'
#
#     The number of knots is chosen using the recommendations above using 
#     n = limiting sample size = min(number of events, number of non-events).
#
#     If no model can be fit with the current number of knots (nk > 3), the 
#     number of is decreased by 1 and a new fit is attempted. If the fit for 
#     nk = 3 does not work, the logit of the linear predictor is modeled 
#     linearly. This is just logistic calibration. 
#
#     In the help of rms::rcspline.eval the following is mentioned about the 
#     knot locations:
#     'If not given, knots will be estimated using default quantiles of x. For 
#      3 knots, the outer quantiles used are 0.10 and 0.90. For 4-6 knots, the
#      outer quantiles used are 0.05 and 0.95. For \code{nk}>6, the outer 
#      quantiles are 0.025 and 0.975. The knots are equally spaced between 
#      these on the quantile scale. For fewer than 100 non-missing values of x, 
#      the outer knots are the 5th smallest and largest x.
#
#     This function gets the following objects from the environment it is 
#     called: 'cal_version', 'repn_number' and 'rcs_info_[cal_version]'. It adds 
#     information about the number of knots used by RCS during the current 
#     'repn_number' and updates 'rcs_info_[cal_version]' in the calling 
#     environment.
#
#     Remark: If one wants to use this function outside the simulation to 
#             calibrate predicted probabilities, the lines of code that are used
#             to keep track of the number of knots used should be disabled.
# ==============================================================================

calibrate_rcs <- function(response, data, 
                          newdata1, newdata2, prob) {
  
  objects_to_get <- c("cal_version", "repn_number")
  for(i in objects_to_get) assign(i, get(i, envir = parent.frame()))  
  rm(i)
  
  assign("rcs_info", get(paste0("rcs_info_", cal_version), 
                         envir = parent.frame()))
  
  n <- nrow(newdata1)
  n_events <- sum(newdata1[[response]])
  limiting_sample_size <- min(n_events, n - n_events)
  
  rcs_info_row <- data.frame(n_cal_train = n, n_events_cal_train = n_events, 
                             limiting_sample_size_cal_train = 
                               limiting_sample_size,
                             nk_5 = FALSE, nk_4 = FALSE, nk_3 = FALSE,
                             nk_0 = FALSE)
  
  nk <- 5
  if(limiting_sample_size >= 30 & limiting_sample_size < 100) nk <- 4
  if(limiting_sample_size < 30) nk <- 3
  
  for(nk_used in 3:5) {
    if(nk == nk_used) rcs_info_row[[paste0("nk_", nk_used)]] <- TRUE
  }

  prob1 <- newdata1[, prob] %>% winsorize_probs()
  prob2 <- newdata2[, prob] %>% winsorize_probs()
  
  newdata1[["lp"]] <- qlogis(prob1)
  newdata2[["lp"]] <- qlogis(prob2)
  
  Formula <- paste0(response, " ~ rcs(lp, ", nk, ")") %>% as.formula
  fit <- tryCatch(lrm(Formula, data = newdata1, maxit = 100), error = identity)
  
  while(("simpleError" %in% class(fit) | 
         ("lrm" %in% class(fit) & length(names(fit)) == 1)) & nk >= 3) {
    nk_old <- nk
    if(nk > 3) {
      nk <- nk - 1
      
      rcs_info_row[[paste0("nk_", nk)]] <- TRUE
      
      Formula <- paste0(response, " ~ rcs(lp, ", nk, ")") %>% as.formula()
      mssg <- paste("Reducing nk in rcs from", nk_old, "to", nk, 
                    "when recalibrating", prob)
      print(mssg)
      fit <- tryCatch(lrm(Formula, data = newdata1, maxit = 100), 
                      error = identity)    
      
    } else {
      nk <- 0
      rcs_info_row[[paste0("nk_0")]] <- TRUE
      # prediction failed when using nk = 0 , which is forcing a linear fit 
      # (different errors were observed). 
      # For that reason slightly different code is used here:
      mssg <- paste("Reducing nk in rcs from", nk_old, "to", nk,
                    "when recalibrating", prob)
      print(mssg)
      
      Formula <- paste0(response, " ~ lp") %>% as.formula
      fit <- glm(Formula, data = newdata1, family = binomial())
      
      rcs_info[repn_number, ] <- rcs_info_row
      assign("rcs_info", rcs_info, envir = parent.frame())
      
      return(predict(fit, newdata = newdata2, type = "response"))     
    }
  }
  
  rcs_info[repn_number, ] <- rcs_info_row
  assign(paste0("rcs_info_", cal_version), rcs_info, envir = parent.frame())
  
  as.numeric(predict(fit, newdata = newdata2, type = "fitted"))
  
}


# ==============================================================================
# 14) Simcal calibration.
#
#     Calibrates a vector of probabilities using the novel simcal approach. 
#
#     ARGUMENTS
#     
#     response: character string giving the name of the column containing the 
#               binary response in 'data' and 'newdata1'.
#     data: data frame from population 1 used for model building.
#     newdata1: data frame from population 2 used for its correlation matrix.
#     newdata2: data frame from population 2 used for testing of recalibrated
#               probabilities.
#     prob:     character string column indicating the name of the column in 
#               data frames 'newdata1' and 'newdata2' that will be contain
#               the probabilities that will be recalibrated.
# 
#     VALUE
#
#     A numeric vector containing the recalibrated probabilities for 
#     'newdata2'.  
# ==============================================================================

is.bin <- function(x) { all(x %in% 0:1) }

calibrate_simcal <- function(response, data, 
                             newdata1, newdata2, prob, mispec) {
  
  data_app <<- data
  vars_modgo <<- setdiff(colnames(data_app), c("y_fac", "repn", "data_desc"))
  
  vars_modgo_bin <<- c()
  for (var_name in vars_modgo) {
    if (is.bin(newdata1[[var_name]])) {
      vars_modgo_bin <<- c(vars_modgo_bin, var_name)
    }
  }
  
  vars_modgo_cont <<- setdiff(vars_modgo, vars_modgo_bin)
  
  # simulate data
  res_modgo <- modgo(data = data_app[,setdiff(colnames(data_app), c("y_fac", "repn", "data_desc"))], 
                     variables = setdiff(colnames(data_app), c("y_fac", "repn", "data_desc")), 
                     bin_variables = vars_modgo_bin, nrep = 1)
  norm_cor_matrix <- res_modgo[["covariance_matrix"]]
  
  n_samples <- 100000L
  set.seed(10034)
  df_sim <- MASS::mvrnorm(n = n_samples, mu = rep(0, ncol(norm_cor_matrix)), 
                          Sigma = norm_cor_matrix) %>% data.frame() 
  
  
  for (j in vars_modgo_cont) { 
    df_sim[[j]] <-
      modgo::rbi_normal_transform_inv(df_sim[[j]], data_app[[j]])
    
    # updating mean and sd of simulated data for continuous variables
    # using new study information
    
    mean_new <- mean(data_app[[j]])
    mean_study <- mean(newdata1[[j]])
    
    
    sd_new <- sd(data_app[[j]])
    sd_study <- sd(newdata1[[j]])
    
    df_sim[[j]] <-
      ((df_sim[[j]] - mean_new) / sd_new) * sd_study +
      mean_study 
    
  }
  
  # processing binary variables using the cutoffs of the new study
  
  for (j in c(vars_modgo_bin)) {
    df_sim[[j]] <-
      modgo::rbi_normal_transform_inv(df_sim[[j]], newdata1[[j]])
  }
  
  machine_specification <- logistic_reg() %>% 
    set_engine("glm") %>% 
    set_mode("classification")
  
  predictors <- names(data_app)
  predictors <- predictors[str_detect(predictors, "^(x|X)")]
  
  Formula <- paste0("y_fac ~ ", paste(predictors, collapse = "+")) %>%
    as.formula 
  
  fit_build <- machine_specification %>% fit(Formula, data = data_app)
  
  aux <- tryCatch(predict(fit_build, 
                          new_data = df_sim, 
                          type = "prob")[[".pred_1"]], error = identity)
  
  if("simpleError" %in% class(aux)) aux <- rep(NA_real_, nrow(df_sim))
  
  df_sim[[prob]] <- aux; rm(aux)
  
  Formula <-
    paste0(response, " ~ I( log(", prob , "/(1-", prob, ")))") %>%
    as.formula()
  
  # if (length(df_sim[[prob]][df_sim[[prob]] == 0]) > 0 ) {
  #   df_sim[[prob]][df_sim[[prob]] == 0] <- 10^(-8)
  # }
  # 
  
  fit <- glm(Formula, data = df_sim, family = binomial())
  
  stats::predict.glm(fit, newdata = newdata2, type = "response")
  
}


# ==============================================================================
# 15) Creates a matrix where prevalences and EPVs per simulation repetition will 
#     be stored. For each machine creates a data frame where the information on 
#     the number of knots used during the RCS calibration is stored. For each
#     machine creates a data frame where the information about the number of 
#     observations where Chen calibration produces a 0 / 0 is encountered.
#
#     This function has no arguments and returns NULL. This function gets the 
#     following objects from the environment it is called: 'repns', 
#     'machine_abbreviations'.
#     This function creates a matrix 'matrix_prev_epv'. For each machine an
#     object named 'rcs_info_[cal_train_version]_[machine name]' and and 
#     object named 'chen_info_[cal_train_version]_[machine name]'in the 
#     environment where the function is called.

# ==============================================================================

create_prev_epv_rcs_matrices <- function() {
  
  objects_to_get <- c("repns", "machine_abbreviations")
  for(i in objects_to_get) assign(i, get(i, envir = parent.frame()))

  mat <- matrix(NA_real_, nrow = max(repns), ncol = 6)
  colnames(mat) <- c("prev_build_train", "prev_cal_train", "prev_cal_alt_train",  
                     "epv_build_train", "epv_cal_train", "epv_cal_alt_train")
  
  assign("matrix_prev_epv", mat, envir = parent.frame())
  
  aux <- rep(NA_integer_, max(repns))
  rcs_info <- data.frame(n_cal_train = aux, n_events_cal_train = aux, 
                         limiting_sample_size_cal_train = aux,
                         nk_5 = FALSE, nk_4 = FALSE, nk_3 = FALSE,
                         nk_0 = FALSE)
  
  chen_info <- data.frame(n_cal_train = aux, n_events_cal_train = aux,
                          n_cal_test = aux, n_events_cal_test = aux,
                          n_zero_over_zero_cal_test = aux)
  
  for(i in machine_abbreviations) {
    assign(paste0("rcs_info_cal_", i), rcs_info, envir = parent.frame()) 
    assign(paste0("rcs_info_cal_alt_", i), rcs_info, envir = parent.frame()) 
    
    assign(paste0("chen_info_cal_", i), chen_info, envir = parent.frame()) 
    assign(paste0("chen_info_cal_alt_", i), chen_info, envir = parent.frame())     
  }
  
  invisible(NULL)
  
}

# ==============================================================================
# 16) Computes prevalences and number of event per variables in simulated data
#     and updates the matrix where this information is being stored.
#
#     This function has no arguments and returns NULL. This function gets the 
#     following objects from the environment it is called: 'df_build_train', 
#     'df_cal_train', 'df_cal_alt_train', 'matrix_prev_epv', 'repn_number'. 
#     'df1_train' is the original machines were trained. 'df2_train' is the data
#     where the machines are recalibrated. 'matrix_prev_epv' is the matrix where
#     the prevalences and events per variable per replication will be stored.
#     'repn_number' is the replication number.
#     The function computes the prevalences ('prev_build', 'prev_cal_train')
#     of the simulated outcome and the number of events per variable 
#     ('epv_build', 'epv_cal_train') in 'df1_train' and 'df2_train', and stores 
#     these objects 'matrix_prev_epv'. The latter is updated in the environment 
#     where the function is called.
# ==============================================================================

compute_prev_epv_and_update_matrix <- function() {
  objects_to_get <- c("df_build_train", "df_cal_train", "df_cal_alt_train",
                      "matrix_prev_epv", "repn_number")
  for(i in objects_to_get) assign(i, get(i, envir = parent.frame()))  
  rm(i)
  
  n_predictors <- names(df_build_train)[str_detect(names(df_build_train), 
                                                   "^(x|X)")] %>% length
  
  for(df_name in c("build_train", "cal_train", "cal_alt_train")) {
    df <- get(paste0("df_", df_name))
    prev <- mean(df[["y"]])
    epv <- min(sum(df[["y"]]), sum(1 - df[["y"]])) / n_predictors
    matrix_prev_epv[repn_number, paste0("prev_", df_name)] <- prev 
    matrix_prev_epv[repn_number, paste0("epv_", df_name)] <- epv
    rm(epv)
  }
  
  
  assign("matrix_prev_epv", get("matrix_prev_epv"), envir = parent.frame())
  
  invisible(NULL)
}

# ==============================================================================
# 17) Misspecified simcal calibration.
#
#     Calibrates a vector of probabilities using the novel simcal approach with
#     wrongly specified parameters to investigate their effects.
#
#     ARGUMENTS
#     
#     response: character string giving the name of the column containing the 
#               binary response in 'data' and 'newdata1'.
#     data: data frame from population 1 used for model building.
#     newdata1: data frame from population 2 used for its correlation matrix.
#     newdata2: data frame from population 2 used for testing of recalibrated
#               probabilities.
#     prob:     character string column indicating the name of the column in 
#               data frames 'newdata1' and 'newdata2' that will be contain
#               the probabilities that will be recalibrated.
#    mispec:    character string composed of three elements (mean, variance, 
#               proportion) that indicate misspecified parameter values.
# 
#     VALUE
#
#     A numeric vector containing the recalibrated probabilities for 
#     'newdata2'.  
# ==============================================================================

format_mean_sd <- function(Mean, Sd, round_dec) 
{
  paste0(epade::round_n.ade(Mean, round_dec), " ± ", epade::round_n.ade(Sd, 
                                                                        round_dec))
}

is.bin <- function(x) { all(x %in% 0:1) }

calibrate_simcal_mispec <- function(response, data, 
                             newdata1, newdata2, prob, mispec) {
  
  mispec_mean <- mispec[1]
  mispec_var <- mispec[2]
  mispec_prop <- mispec[3]
  
  vars_modgo <<- setdiff(colnames(data), c("y_fac", "repn", "data_desc"))
  
  vars_modgo_bin <<- c()
  for (var_name in vars_modgo) {
    if (is.bin(newdata1[[var_name]])) {
      vars_modgo_bin <<- c(vars_modgo_bin, var_name)
    }
  }
  
  vars_modgo_cont <<- setdiff(vars_modgo, vars_modgo_bin)#, vars_modgo_cat)
  
  # simulate data
  res_modgo <- modgo(data = data[,vars_modgo], 
                     variables = vars_modgo, 
                     bin_variables = vars_modgo_bin, 
                     #categ_variables = vars_modgo_cat,
                     nrep = 1)
  norm_cor_matrix <- res_modgo[["covariance_matrix"]]
  
  n_samples <- 100000L 
  set.seed(10034) 
  # Simulating data using that correlation matrix estimated by modgo
  df_sim <- MASS::mvrnorm(n = n_samples, mu = rep(0, ncol(norm_cor_matrix)), 
                          Sigma = norm_cor_matrix) %>% data.frame() 
  
  df_sim[[ vars_modgo_cont[1] ]] <-
    modgo::rbi_normal_transform_inv(df_sim[[vars_modgo_cont[1]]], data[[vars_modgo_cont[1]]])
  
  # updating mean and sd of simulated data for continuous variables
  # using new study information and misspecification
  
  mean_new <- mean(data[[vars_modgo_cont[1]]])
  if (mispec_mean != "none") {
    mean_study <- as.numeric(mispec_mean)
  } else {mean_study <- mean(newdata1[[vars_modgo_cont[1]]])}
  
  sd_new <- sd(data[[vars_modgo_cont[1]]])
  if (mispec_var != "none") {
    sd_study <- sqrt(as.numeric(mispec_var)) 
  } else {sd_study <- sd(newdata1[[vars_modgo_cont[1]]])}
  
  df_sim[[vars_modgo_cont[1]]] <-
    ((df_sim[[vars_modgo_cont[1]]] - mean_new) / sd_new) * sd_study + mean_study 
  
  for (j in setdiff(vars_modgo_cont, vars_modgo_cont[1])) {
    df_sim[[j]] <-
      modgo::rbi_normal_transform_inv(df_sim[[j]], data[[j]])

    # updating mean and sd of simulated data for continuous variables
    # using new study information

    mean_new <- mean(data[[j]])
    mean_study <- mean(newdata1[[j]])


    sd_new <- sd(data[[j]])
    sd_study <- sd(newdata1[[j]])

    df_sim[[j]] <-
      ((df_sim[[j]] - mean_new) / sd_new) * sd_study +
      mean_study

  }
  

  # processing binary variables using the cutoffs of the new study 
  # and misspecification
  
  
  if (length(vars_modgo_bin) == 1 & mispec_prop != "none") {
    
    df_sim[[response]] <-
      modgo::rbi_normal_transform_inv(df_sim[[response]], 
                                      rbinom(length(newdata1[[response]]), 
                                             size = 1, 
                                             prob = as.numeric(mispec_prop)))
  }
  
  if (length(vars_modgo_bin) > 1 & mispec_prop != "none") {
  
      df_sim[[response]] <-
        modgo::rbi_normal_transform_inv(df_sim[[response]], 
                                        rbinom(length(newdata1[[response]]), 
                                               size = 1, 
                                               prob = as.numeric(mispec_prop)))
      
      for (j in setdiff(vars_modgo_bin, response)) {
        df_sim[[j]] <-
          modgo::rbi_normal_transform_inv(df_sim[[j]], newdata1[[j]])
      }
      
  } else if (mispec_prop == "none") {
    for (j in (vars_modgo_bin)) {
      df_sim[[j]] <-
        modgo::rbi_normal_transform_inv(df_sim[[j]], newdata1[[j]])
    }
  }
  
  
  machine_specification <- logistic_reg() %>% 
    set_engine("glm") %>% 
    set_mode("classification")
  
  predictors <- names(data)
  predictors <- predictors[str_detect(predictors, "^(x|X)")]
  
  Formula <- paste0("y_fac ~ ", paste(predictors, collapse = "+")) %>%
    as.formula 
  
  fit_build <- machine_specification %>% fit(Formula, data = data)
  
  aux <- tryCatch(predict(fit_build, 
                          new_data = df_sim, 
                          type = "prob")[[".pred_1"]], error = identity)
  
  if("simpleError" %in% class(aux)) aux <- rep(NA_real_, nrow(df_sim))
  
  df_sim[[prob]] <- aux; rm(aux)
  
  Formula <-
    paste0(response, " ~ I( log(", prob , "/(1-", prob, ")))") %>%
    as.formula()
  
  # if (length(df_sim[[prob]][df_sim[[prob]] == 0]) > 0 ) {
  #   df_sim[[prob]][df_sim[[prob]] == 0] <- 10^(-8)
  # }
  # 
  
  fit <- glm(Formula, data = df_sim, family = binomial())
  
  stats::predict.glm(fit, newdata = newdata2, type = "response")
  
}

# ==============================================================================
# 18) Calibrates probabilities using Elkan approach.
#     
#     Calibrates a vector of probabilities using Elkan general updating 
#     approach: 
#     Elkan C. The foundations of cost-sensitive learning. In: Nebel B., ed. 
#     Proceedings of the Seventeenth International Joint Conference on 
#     Artificial Intelligence. Vol. 2. San Francisco: Morgan Kaufmann. 2001 
#     (pp. 973–978).
#     with wrongly specified parameters to investigate their effects.
#
#     ARGUMENTS
#     
#     response: character string giving the name of the column containing the 
#               binary response in 'data' and 'newdata1'.
#     data: data frame from population 1 used for model building.
#     newdata1: data frame from population 2 used for model recalibration.
#     newdata2: data frame from population 2 used for testing of recalibrated
#               probabilities.
#     prob:     character string column indicating the name of the column in 
#               data frames 'newdata1' and 'newdata2' that will be contain
#               the probabilities that will be recalibrated.
#     mispec:    character string composed of three elements (mean, variance, 
#               proportion) that indicate misspecified parameter values.
#
#     VALUE
#
#     A numeric vector containing the recalibrated probabilities for 
#     'newdata2'.  
# ==============================================================================
calibrate_elkan_mispec <- function(response, data, 
                            newdata1, newdata2, prob, mispec) {
  
  mispec_mean <- mispec[1]
  mispec_var <- mispec[2]
  mispec_prop <- mispec[3]
  
  br1 <- mean(data[, response])
  
  if (mispec_prop != "none") {
    br2 <- as.numeric(mispec_prop) 
  } else {
    br2 <- mean(newdata1[, response]) 
  }

  prob2 <- newdata2[, prob]
  
  br2 * (prob2 - prob2 * br1) / (br1 - prob2 * br1 + br2 * prob2 - br1 * br2)
}


# ==============================================================================
# 19) SBA-10 calibration.
#
#     Calibrates a vector of probabilities using SBA-10 calibration, as 
#     described in Bella et al., Similarity-binning averaging: a generalisation 
#     of binning calibration (2009). doi:10.1007/978-3-642-04394-9_42
#
#     ARGUMENTS
#     
#     response: character string giving the name of the column containing the 
#               binary response in 'data' and 'newdata1'.
#     data: data frame from population 1 used for model building.
#     newdata1: data frame from population 2 used for model recalibration.
#     newdata2: data frame from population 2 used for testing of recalibrated
#               probabilities.
#     prob:     character string column indicating the name of the column in 
#               data frames 'newdata1' and 'newdata2' that contain
#               the probabilities that will be recalibrated.
#
#     VALUE
#
#     A numeric vector containing the recalibrated probabilities for 
#     'newdata2'.  
# ==============================================================================

calibrate_sba10 <- function(response, data, 
                            newdata1, newdata2, prob, mispec) {
  predictors <- names(data)
  predictors <- predictors[str_detect(predictors, "^(x|X)")]
  Formula <- paste0("y_fac ~ ", paste(predictors, collapse = "+")) %>%
    as.formula 
  machine_abbr <- substr(prob, nchar(prob)-2+1, nchar(prob))
  if(machine_abbr == "lr") {
    machine_specification <- logistic_reg() %>% 
      set_engine("glm") %>% 
      set_mode("classification")
  }
  fit <- machine_specification %>% fit(Formula, data = data)
  aux <- tryCatch(predict(fit, 
                          new_data = data, 
                          type = "prob")[[".pred_1"]], error = identity)
  
  if("simpleError" %in% class(aux)) aux <- rep(NA_real_, nrow(df_current))
  data_aux <- data
  data_aux[[prob]] <- aux; rm(aux)
  
  p_cal <- NULL
  for (i in 1:nrow(newdata2)) {
    query_point <- newdata2[i, ]
    query_point <- query_point %>%
      select(all_of(predictors), all_of(prob))
    neighbors <- get.knnx(data = data_aux %>%
                            select(all_of(predictors), all_of(prob)), 
                          query = query_point, k = 10)  
    neighbor_indices <- neighbors$nn.index
    neighbor_train <- data_aux[neighbor_indices,]
    p_cal[i] <- sum(neighbor_train[prob])/10
  }
  
  
  return(p_cal)
  
}  

# ==============================================================================
# 20) ACP calibration.
#
#     Calibrates a vector of probabilities using ACP calibration, as decribed in 
#     Jiang et al., Calibrating predictive model estimates to support 
#     personalized medicine (2012). doi:10.1136/amiajnl-2011-000291
#     This method has only been developed for logistic regression and is not 
#     applicable to other predictor machines.
#
#     ARGUMENTS
#     
#     response: character string giving the name of the column containing the 
#               binary response in 'data' and 'newdata1'.
#     data: data frame from population 1 used for model building.
#     newdata1: data frame from population 2 used for model recalibration.
#     newdata2: data frame from population 2 used for testing of recalibrated
#               probabilities.
#     prob:     character string column indicating the name of the column in 
#               data frames 'newdata1' and 'newdata2' that contain
#               the probabilities that will be recalibrated.
#
#     VALUE
#
#     A numeric vector containing the recalibrated probabilities for 
#     'newdata2'.  
# ==============================================================================

invlogit <- function(x)
{
  return( exp(x) / (1 + exp(x) ) )
}

calibrate_acp <- function(response, data, 
                          newdata1, newdata2, prob, mispec)
{
  train.x = data
  train.y = response
  
  # (1-alpha)% critical term
  crit.term = qnorm(1 - 0.05/2)
  
  predictors <- names(data)
  predictors <- predictors[str_detect(predictors, "^(x|X)")]
  Formula <- paste0("y_fac ~ ", paste(predictors, collapse = "+")) %>%
    as.formula 
  machine_abbr <- substr(prob, nchar(prob)-2+1, nchar(prob))
  if(machine_abbr == "lr") {
    machine_specification <- logistic_reg() %>% 
      set_engine("glm") %>% 
      set_mode("classification")
  }
  fit <- machine_specification %>% fit(Formula, data = data)
  #aux <- predict(fit, new_data = data, type = "prob")[[".pred_1"]]
  
  test.x = newdata2 %>% select(all_of(predictors))
  
  train = list()
  train$pred = stats::predict(fit[["fit"]], new_data=train.x, se.fit=TRUE)
  train$yhat = invlogit(train$pred$fit)
  
  # apply the trained model to a test data
  test = list()
  test$pred = predict(fit[["fit"]], newdata=test.x, se.fit=TRUE)
  n.test = nrow(test.x)
  # prediction interval for the test set
  test$yhat = invlogit( test$pred$fit )
  test$lower = invlogit( test$pred$fit - crit.term * test$pred$se)
  test$upper = invlogit( test$pred$fit + crit.term * test$pred$se)
  test$ublb = cbind(lower=test$lower, upper=test$upper)
  # prediction range
  r = max(train$yhat) - min(train$yhat)
  # covariance matrix for coefficients
  beta.covmat = as.matrix( summary(fit[["fit"]])$cov.unscaled )
  # Equation 5
  X.mat = as.matrix( cbind(1, test.x) )
  logit.covmat = X.mat %*% beta.covmat %*% t(X.mat)
  vxb = diag(logit.covmat)
  idxSE = sqrt(vxb)
  w = as.numeric( fit[["fit"]]$coef )
  idxLower = X.mat %*% w - crit.term * idxSE * r
  idxUpper = X.mat %*% w + crit.term * idxSE * r
  # Step 2
  lb = invlogit( idxLower )
  ub = invlogit( idxUpper )
  # Step 3 & 4
  p.calib = rep(0, nrow(test.x))
  for(i in 1:n.test) {
    idx.WithinCI = ( lb[i] < train$yhat) & (train$yhat < ub[i])
    n.trainedPredWithinCI = sum(idx.WithinCI)
    if(n.trainedPredWithinCI == 0 ) {
      dist.testpoint = (train$yhat - test$yhat[i])^2
      dist.minimum = min(dist.testpoint)
      isMinimizer = dist.testpoint == dist.minimum
      idx.WithinCI = c(1:length(dist.testpoint))[isMinimizer]
      n.trainedPredWithinCI = length(idx.WithinCI)
    }
    p.calib[i] = sum(fit[["fit"]]$y[idx.WithinCI]) / n.trainedPredWithinCI
  }
  return(p.calib)
}
```
