## Supplementary material for "Calibrating machine learning approaches for probability estimation without calibration data": 04-rmd-functions.R

```
# Supplementary Material 2 for 
# "Calibrating machine learning approaches for probability
# estimation in case of the absence of calibration data"

# Functions used for presenting simulation results in .Rmd file 
# ==============================================================================
#
# Contents
# ==============================================================================

# 01) calibration_names, machine_abbreviations, machine_full_names, cols_lr
# 02) kable_wrapper
# 03) prepare_plot_df
# 04) beautify_recal_labels
# 05) log_loss, mse_brier
# 06) summarise_perf_measure
# 07) iman_davenport_test
# 08) CD_diagram_Nemenyi
# 09) prepare_data_average_logloss_demsar
# 10) format_p.ade
# 11) round_n.ade
# 12) prepare_plot_df_mispec

################
#              #
#      ##      #
#      ##      #
#      ##      #
#   ########   #
#    ######    #
#     ###      #
#      #       #
#              #
################

# ==============================================================================
# 01) Selected objects containing information (like machine, calibration 
#     approaches names or abbreviations) that will be used during the generation 
#     of figures and tables produced to examine the simulation results.
# ==============================================================================

calibration_names <- c("elkan", "rcs", "slope", "logistic", 
                       "beta", "isotonic", "platt", "chen")
machine_abbreviations <- c("lr", "rf", "boost", "svm", "en")
machine_full_names <- c("Logistic regression", "Random forest", 
                        "Gradient boosting", "Support vector machine", 
                        "Elastic net")
names(machine_full_names) <- machine_abbreviations

cols_lr <- c("prob_lr", "prob_lr_refit", 
             paste0("prob_lr_", calibration_names, "_cal")) 

# ==============================================================================
# 02) Wrapper for knitr::kable and kableExtra::kable_styling.
#     
#     ARGUMENTS
#
#     x: matrix or data frame.
#     caption: table caption 
#     position: this is the position argument of kableExtra::kable_styling.
#     ...: further arguments passed to knitr::kable.
#
#     VALUE
#
#     A character string with 'x' formatted as html table.
#
# ==============================================================================

kable_wrapper <- function (x, caption = NULL, position = "center", ...) {

  knitr::kable(x, caption = caption, ...) %>% 
    kableExtra::kable_styling(bootstrap_options = c("striped", "hover"), 
                              position = position)
}

# ==============================================================================
# 03) Formats results of a single simulation scenario. 
#
#     The Rds file containing the results of a simulation scenario is 
#     read and the results are processed into a single data frame to facilitate 
#     the production of figures and tables. 
#     
#     ARGUMENTS
#
#     scenario: the simulation scenario.
#     cols: names of columns in simulation results containing the estimated
#           probabilities that should be kept.
#     build_or_cal: 'build' refers to the model-building test data. 'cal'
#                    and 'cal_alt' refer to the two versions of the calibration 
#                    data (the 'cal_alt' train dataset has twice the number 
#                    of observations as the corresponding train dataset in 
#                    'cal').
#     reduce_repn: keeps only 'N_REPN' out of the 100 replications used in the 
#                  simulation scenario.
#     N_REPN: number of replications to to use. Used only if reduce_repn is set 
#             to TRUE.
#
#     VALUE
#
#     A data frame containing the results of a simulation scenario.
#
# ==============================================================================

prepare_plot_df <- function(scenario, cols, 
                            build_or_cal = c("build", "cal", "cal_alt"),
                            reduce_repn = FALSE, N_REPN = 5) {
  
  build_or_cal <- match.arg(build_or_cal)

  # Reading results file 
  File <- here("rds-files/after-recalibration", 
               paste0("data-with-cal-probs--scen-", scenario, ".Rds"))
  results <- readRDS(file = File)
  
  
  data_list <- results[["data_list"]]
  repn <- length(data_list)
  dsub <- lapply(data_list, `[[`, build_or_cal) %>% 
    lapply(`[[`, "test") %>% do.call("rbind", .)
  
  if(reduce_repn) {
    dsub <- subset(dsub, repn <= N_REPN)
  }  
  dsub[["id"]] <- 1:nrow(dsub)
  dsub <- dsub[, c("id", "repn", "prob_true", cols)]
  dsub$scenario <- scenario
  
  pivot_longer(dsub, cols = {{cols}}, 
               names_to = "machine_and_recal_method",
               names_prefix = "prob_", 
               values_to = "Probability")
  
}


# ==============================================================================
# 04) Beautifies calibration method names
#
#     This function is used to 'beautify' the column 'machine_and_recal_method'
#     from the output returned by the function 'prepare_plot_df' (see above). 
#     This function should be used only the the data frame used as input 
#     contains the results for a single machine (e.g., lr). It removes the 
#     machine abbreviation (e.g. 'svm') and underscores, leaving only the 
#     calibration method name.
#     
#     ARGUMENTS
# 
#     x: The column 'machine_and_recal_method' by a data frame produced by 
#        'prepare_plot_df'.
#     machine: the machine abbreviation (see object 'machine_abbreviations' at 
#              the beginning of this script).
#
#     VALUE
# 
#     A factor with the names used for the calibration methods in tables and 
#     figures.
#     
# ==============================================================================

beautify_recal_labels <- function(x, machine) {
  Levels <- c("NoCal", "Logistic", "Beta", "RCS", "Slope", 
              "Refit", "Platt", "Isotonic", "Elkan", "Chen")
  
  x <- gsub("_", " ", x) %>% 
    sub(paste0("^", machine, " "), "", .) %>%
    sub(" cal$", "", .) %>% 
    sub(paste0("^", machine), "NoCal", .) %>%
    sub("rcs", "RCS", .) %>%
    capitalize() %>% factor(levels = Levels)
  if(sum(is.na(x)) > 0) stop("NAs introduced")
  x
}


# ==============================================================================
# 05) Computes LogLoss or MSE.
#
#     Functions to compute LogLoss or mean squared error (MSE).
#     
#     ARGUMENTS
# 
#     p_true: vector with true probabilities.
#     p_hat: vector with estimated probabilities.
#     winsorize: should 'p_hat' be winsorized? It relies on function 
#                'winsorize_probs' found in '03-calibration-functions.R'
#
#     VALUE
# 
#     The computed LogLoss or MSE.
#
# ==============================================================================

log_loss <- function(p_true, p_hat, winsorize = TRUE) {
  
  if(winsorize) {
    p_hat <- winsorize_probs(p_hat)
  } 
  
  (-1) * mean(p_true * log(p_hat) + (1 - p_true) * log(1 - p_hat))
}

mse_brier <- function(p_true, p_hat, winsorize = TRUE) {
  
  if(winsorize) {
    p_hat <- winsorize_probs(p_hat)
  } 
  mean((p_true - p_hat)^2)
}


# ==============================================================================
# 06) Produces data frame with summary statistics of performance measure. 
#     
#     This function takes the output of 'prepare_plot_df' to which a performance
#     measure has been added (like LogLoss or MSE) for each simulation 
#     replications and computes summary statistics for that summary measure.
#     
#     ARGUMENTS
# 
#     df: a data frame
#     perf_measure: charactring string giving the name in 'df' of the column
#                   containing the performance measure to be summarized.
#     
#     VALUE
#     
#     A data frame with summary statistics for the performance measure.
#
# ==============================================================================

summarise_perf_measure <- function(df, perf_measure) {
  names(df)[names(df) == "recal_method"] <- "Approach"
  out <- group_by({{df}}, scenario, Approach) %>% 
    summarise(`Mean` = mean({{perf_measure}}), 
              `Standard deviation` = sd({{perf_measure}}), 
              `Median` = median({{perf_measure}}), 
              `25th percentile` = quantile({{perf_measure}}, probs = 0.25),
              `75th percentile` = quantile({{perf_measure}}, probs = 0.75)) 
  names(out)[names(out) == "scenario"] <- "Scenario"
  out
}


# ==============================================================================
# 07) Performs Iman & Davenport version of Friedman test and computes
#     Nemenyi post-hoc test.
#
#     The code is taken from the function pairedComparisons in the package
#     performanceEstimation.
#     
#     The function 'rankMatrix' defined 'inside iman_davenport_test' is taken 
#     from scmamp package (with minor changes).
#     
#     ARGUMENTS
#
#     input_matrix: numeric matrix. Rows correspond to the different 
#                   replications for a simulation scenario and columns to the
#                   different calibration methods. Each cell contains a 
#                   performance measure (we assume it is the  LogLoss) 
#                   comparing the true probabilities with the calibrated 
#                   predicted probabilities.
#
#     p.value: significance level.
#     small_better: Are smaller average ranks better?
#     
#     VALUE
#
#     A list with the test results, including the critical difference.
#
# ==============================================================================

iman_davenport_test <- function(input_matrix, p.value = 0.05, 
                                small_better = TRUE) {
  
  rankMatrix <- function(data, small_better) {
    if (!small_better){
      f <- function(x){
        rank(-x, ties.method="average")
      }
    } else {
      f <- function(x){
        rank(x, ties.method="average")
      }
    }
    rankings <- t(apply(data, MARGIN = 1, FUN = f))
    colnames(rankings) <- colnames(data)
    rownames(rankings) <- rownames(data)
    rankings
  }
  
  nts <- nrow(input_matrix)
  nws <- ncol(input_matrix)
  mr <- colMeans(rankMatrix(input_matrix, small_better = small_better))
  
  chi <- 12 * nts/(nws * (nws + 1)) * (sum(mr^2) - (nws * (nws + 1)^2)/4)
  FF <- (nts - 1) * chi/(nts * (nws - 1) - chi)
  critVal <- df(1 - p.value, nws - 1, (nws - 1) * (nts - 1))
  rejNull <- FF > critVal
  pval_id <- 1 - pf(FF, df1 = nws - 1, df2 = (nws - 1) * (nts - 1))
  
  output <- list(F.test = list(chi = chi, FF = FF, 
                               critVal = critVal, rejNull = rejNull, 
                               pval_iman_davenport = pval_id),
                 Nemenyi.test = NULL,
                 avgRksWFs = mr
  )
  
  if (rejNull) {
    CD.n <- qtukey(1 - p.value, nws, df = Inf)/sqrt(2) * 
      sqrt(nws * (nws + 1)/(6 * nts))
    allRkDifs <- outer(mr, mr, function(x, y) abs(x - y))
    signifDifs <- allRkDifs >= CD.n
    output[["Nemenyi.test"]] <- list(critDif = CD.n, rkDifs = allRkDifs, 
                                     signifDifs = signifDifs)
  }
  
  list(logloss = output)
}

# ==============================================================================
# 08) Plots results of Nemenyi post hoc test. 
#
#     Modification of CDdiagram.Nemenyi from package performanceEstimation.
#     This modification returns the graphic produced by the function and 
#     implements a minor correction suggested in
#     https://github.com/ltorgo/performanceEstimation/pull/7/commits/b086879f2de916edb7d76ff3a0ba0c30f82fe350
#     
#     Some arguments or changes in the code were added to modify aspects of 
#     the plot and to return the ggplot object created inside the function.
#     In the original plot the the code to draw the figure was part of the 
#     function (neither the gtable object gt nor the ggplot object g were 
#     returned in original code):
#
#         grid::grid.newpage()
#         gt <- ggplot2::ggplot_gtable(ggplot2::ggplot_build(g))
#         gt$layout$clip[gt$layout$name == "panel"] <- "off"
#         grid::grid.draw(gt)
#
#     clip in the code above is modified, because the (calibration) approach 
#     labels and the CD text are drawn outside of the plot panel. We no longer 
#     need this since we set clip = "off inside ggplot2::coord_fixed when 
#     generating the ggplot object inside the function.
#     
#     The original arguments of the function are r and metrics. All other 
#     arguments were added.
#
#     The argument coord_fixed_ratio was added to control the aspect ratio of 
#     the plot (default is 0.5).
#
#     The argument bw was added to control if the graphic should be black and 
#     white or use colors (default FALSE).
#
#     The argument x_axis_1_right was added to have the axis-labels starting
#     with 1 on the right of the plot and then they increase from left to right.
#     Default is TRUE.
#
#     The argument size_cd_text was added to control the size of the text
#     above the CD segment.
#     
#     The argument size_cd_segment was added to control the thickness of the
#     CD segment. Default is 1.5. This argument is passed to the size argument
#     of ggplot2::annotate.
#
#     The argument size_segments_joining_methods was added to control
#     the thickness of the line segments joining the methods compared.
#
#     The argument show_cd_segment_val was added to be able to omit 
#     the CD segment.
#
#     The argument dist_cd_segment was added to be able to shift vertically
#     the CD segment.
#
#     The argument base_size was added to control the size of the text in the
#     graphics. This does not affect the text written outside the plot panel.
#
#     The argument size_methods_text was added to control the size of the 
#     text giving the names of the (calibration) methods compared.
#
#     The arguments axis_x_text_face and axis_x_text_size were introduced
#     to have more control over the appearance of the tick marks labels
#     on the x-axis.
#
#     ARGUMENTS
#
#     r: the function CDdiagram.Nemenyi from which this function is a 
#        modification expects a 
#        "list resulting from a call to pairedComparisons". The function 
#        'iman_davenport_test' defined above produces such a list.
#     metric: a performance metric
#     coord_fixed_ratio: argument to be passed to 'ratio' argument of 
#                        ggplot2::coord_fixed.
#     bw: should the plot use colors or be in Black and White? Default is FALSE.
#     x_axis_1_right: Should the 1 in the x-axis tick labels be on the right
#                     of the graphic (e.g. 3, 2, 1, if FALSE 1, 2, 3). Default
#     is TRUE.
#     size_cd_text: size of text giving the magnitude of the CD. Default is 3.
#     size_cd_segment: size of CD segment. Default is 1.5.
#     size_segments_joining_methods: size of line segment joining different 
#                                    methods. Default is 1.2.
#     show_cd_segment_val: should the CD segment be shown? FALSE will also 
#                          disable the text printed above the CD segment.
#     dist_cd_segment: adjusts vertical position of CD segment. Distance above 
#                      the highest horizontal line segment connecting the 
#                      average rank with the method label. Default is 1.
#     base_size: argument to be passed to the argument of the same name in
#                ggplot2::theme_grey. This argument does not affect the size of
#                labels of the calibration methods nor the size of the 
#                text giving the magnitude of the critical difference. 
#                Default is 11.
#     size_methods_text: Size of the text giving the names of the (calibration)
#                        methods. Default is 4.
#     axis_x_text_face:  font face of x-axis tick mark labels. Default is 
#                        "plain".
#     axis_x_text_size: size of text for tick mark labels.
#     
#     VALUE
#
#     A ggplot2 object.
# ==============================================================================

CD_diagram_Nemenyi <- function(r, metric = names(r)[1], coord_fixed_ratio = 0.5, 
                               bw = FALSE, x_axis_1_right = TRUE,
                               size_cd_text = 3,
                               size_cd_segment = 1.5, 
                               size_segments_joining_methods = 1.2, 
                               show_cd_segment_val = TRUE, dist_cd_segment = 1,
                               base_size = 11, 
                               size_methods_text = 4,
                               axis_x_text_face = "plain",
                               axis_x_text_size = 5) {
  
  if (is.null(r[[metric]]$F.test) | 
      is.null(r[[metric]]$Nemenyi.test)) stop(paste0("Results of both the F ",
                                                     "and Nemenyi tests are ",
                                                     "required for these ",
                                                     "diagrams."))
  
  o <- rank(r[[metric]]$avgRksWFs, ties.method = "first")
  mxl <- ceiling(length(r[[metric]]$avgRksWFs)/2)
  data <- data.frame(avgRk = r[[metric]]$avgRksWFs, 
                     invRk = length(r[[metric]]$avgRksWFs) + 
                       1 - r[[metric]]$avgRksWFs, 
                     sys = names(r[[metric]]$avgRksWFs), 
                     line = o%%mxl + ifelse(o%%mxl == 0, mxl, 0), 
                     side = ifelse(o <= mxl, -1, 1))
  data$line <- ifelse(data$side == 1, mxl + 1 - data$line, 
                      data$line)
  len <- length(r[[metric]]$avgRksWFs)
  cd <- r[[metric]]$Nemenyi.test$critDif
  
  col_arg <- "sys"; if(bw){col_arg <- NULL}
  
  g <- ggplot(data, aes_string(x = "invRk", y = "line")) + 
    geom_segment(aes_string(x = "invRk", y = 0, xend = "invRk", yend = "line", 
                            col = col_arg)) + 
    geom_text(data = data[data$side == -1, ], 
              aes_string(label = "sys", x = +Inf, y = "line", col = col_arg), 
              hjust = 0, size = size_methods_text) + 
    geom_text(data = data[data$side == 1, ], 
              aes_string(label = "sys", x = -Inf, y = "line", col = col_arg), 
              hjust = 1, 
              size = size_methods_text) + 
    geom_segment(data = data[data$side == -1, ], 
                 aes_string(x = max(o) + 0.1, y = "line", xend = "invRk", 
                            yend = "line", col = col_arg)) + 
    geom_segment(data = data[data$side == 1, ], 
                 aes_string(x = "invRk", y = "line", 
                            xend = 1 - 0.1, yend = "line", col = col_arg)) 
  
  labels_scale_x <- paste(1:len)
  if(x_axis_1_right) {
    labels_scale_x <- paste(len:1)
  }
  g <- g +  scale_x_continuous(limits = c(1 - 0.15, len + 0.15), 
                               breaks = 1:len, 
                               labels = labels_scale_x) 
  
  g <-  g +  scale_y_continuous(limits = c(0, mxl + 1)) + 
    xlab("Average Rank") + ylab("") + theme_grey(base_size = base_size) +
    theme(axis.text.y = element_blank(), 
          axis.ticks.y = element_blank(), axis.title.y = element_blank(),
          axis.text.x = element_text(size = axis_x_text_size, 
                                     face = axis_x_text_face),
          #axis.title.x = element_text(size = 5),
          legend.position = "none", panel.background = element_blank(), 
          panel.border = element_blank(), 
          axis.line = element_line(size = 1), 
          axis.line.y = element_blank(), panel.grid.major = element_blank(), 
          plot.background = element_blank(), 
          plot.margin = grid::unit(c(3, 6, 1, 5), "lines")) + 
    coord_fixed(ratio = coord_fixed_ratio, clip = "off")
  
  if(show_cd_segment_val) {
    g <- g + annotate("segment", x = 1, xend = cd + 1, 
                      y = mxl + dist_cd_segment, 
                      yend = mxl + dist_cd_segment, 
                      size = size_cd_segment) + 
      annotate("text", x = 1, y = mxl + dist_cd_segment, 
               label = paste("Critical Difference", round(cd, 1), sep = " = "), 
               vjust = -0.5, hjust = 0, size = size_cd_text)
  }  
  wfsOrd <- names(r[[metric]]$avgRksWFs[order(r[[metric]]$avgRksWFs)])
  ss <- r[[metric]]$Nemenyi.test$signifDifs[wfsOrd, wfsOrd]
  mx <- ncol(ss)
  pos <- rep(1:mxl, each = mxl) - seq(0, 1, by = 1/(mxl + 1))[-c(1, mxl + 2)]
  frees <- rep(1, mxl)
  from <- 1
  currTill <- 0
  for (i in 1:nrow(ss)) {
    
    # add i-1 to which(ss[i,i:mx]) to get the correct which-values (since the 
    # column selection removes i-1 values)
    till <- which(ss[i,i:mx]) + (i-1) 
    till <- if (length(till)>0) till[1]-1 else mx
    if (till > i && till > currTill) {
      theLine <- min(data[wfsOrd[i], "line"], data[wfsOrd[till], 
                                                   "line"])
      ypos <- pos[(theLine - 1) * mxl + frees[theLine]]
      frees[theLine] <- frees[theLine] + 1
      g <- g + annotate("segment", x = data[wfsOrd[till], "invRk"] - 0.1, 
                        xend = data[wfsOrd[i], "invRk"] + 0.1, y = ypos, 
                        yend = ypos, size = size_segments_joining_methods)
      currTill <- till
    }
    if (till == mx) break
  }
  
  g
}

# ==============================================================================
# 09) Prepares data for Iman Davenport test.
#
#     This function to prepares the results of a single simulation 
#     scenario, averaging the LogLoss over all machines, so that they can be 
#     passed to the function 'iman_davenport_test' 
#     
#     ARGUMENTS
# 
#     scenario: simulation scenario.
#     reduce_repn: logical. Argument to be passed to prepare_plot_df (in case a 
#                  smaller number of simulation replications should be used in 
#                  computation).
#     N_REPN: number of replications to to use. Used only if reduce_repn is set 
#             to TRUE.
#     
#     VALUE
#    
#     A matrix that can be passed to 'iman_davenport_test'.
#     
# ==============================================================================

prepare_data_average_logloss_demsar <- function(scenario, reduce_repn, N_REPN,
                                                cal_version = c("cal", 
                                                                "cal_alt")) {
  cols <- NULL
  
  if(scenario %in% 16:21) {
    machine_abbreviations <- setdiff(machine_abbreviations, "en")
  }
  
  for(machine in machine_abbreviations){
    cols <- c(cols, sub("prob_lr", paste0("prob_", machine), cols_lr))
  }
  df <- prepare_plot_df(scenario = scenario, cols = cols,
                        build_or_cal = cal_version,
                        reduce_repn = reduce_repn, N_REPN = N_REPN)
  
  df[["Method"]] <- df[["machine_and_recal_method"]]
  
  df[["Machine"]] <- (strsplit(df[["machine_and_recal_method"]], "_") %>% 
                        sapply(`[`, 1))
  
  y <- "Method"
  for(machine in machine_abbreviations){
    df[[y]] <- sub(paste0(machine, "$"), "NoCal", df[[y]])
  }
  df[[y]] <- sub("_cal$", "", df[[y]])
  v <- which(str_detect(df[[y]], "_"))
  df[[y]][v] <- (strsplit(df[[y]][v], "_") %>% sapply(`[`, 2) %>% 
                   sub("rcs", "RCS", .)  %>% capitalize()) 
  Levels <- c("NoCal", "Logistic", "Beta", "RCS", "Slope", 
              "Refit", "Platt", "Isotonic", "Elkan", "Chen")
  
  df[[y]] <- factor(df[[y]], levels = Levels)
  
  df <- group_by(df, Machine, Method, repn) %>% 
    summarise(LogLoss = log_loss(prob_true, Probability)) %>%
    group_by(Method, repn) %>% summarise(`AverageLogLoss` = mean(LogLoss))
  
  df_wide <- pivot_wider(df, names_from = Method, 
                         values_from = AverageLogLoss, id_cols = repn)
  
  df_wide[["repn"]] <- NULL
  
  as.matrix(df_wide)
  
}

# ==============================================================================
# 10) Formats a p-value for printing. 
#
#     This function is taken from R package 'epade' by Andreas Schulz, which at 
#     the time of writing was no longer available on CRAN repository. Minor 
#     cosmetic changes were performed.
#
#     ARGUMENTS
#     
#     x:	a single p-value, a vector of numeric p-values or a matrix of 
#          numeric p-values
#     pgits: Number of digits after decimal for cutoff of the p-value, 4 means, 
#            all under 0.0001 will be shown like <0.0001
#     digits: how many significant digits are to be used. (before the cut with 
#             pgits)
# 
#     VALUE
#     
#     An object of similar structure to x containing character representations 
#     of the elements of x in a common format.
# ==============================================================================


format_p.ade <- function (x, pgits = 4, digits = 2) {
  x <- unlist(x)
  if (!is.vector(x) & !is.matrix(x)) 
    stop("x must be vector or matrix")
  if (length(x) > 1) {
    if (is.vector(x)) {
      y <- x
      for (i in 1:length(x)) {
        y[i] <- format_p.ade(x[i], pgits, digits)
      }
      return(y)
    }
    if (is.matrix(x)) {
      M <- x
      for (i in 1:dim(x)[1]) {
        for (j in 1:dim(x)[2]) {
          M[i, j] <- format_p.ade(x[i, j], pgits, digits)
        }
      }
      return(M)
    }
  }
  if (length(x) == 1) {
    if (is.na(x)) 
      return(x)
    x <- as.numeric(x)
    if (digits%%1 == 0) {
      if (pgits%%1 == 0) {
        y <- base::format(x, digits = digits, scientific = FALSE)
        if (y != 1) {
          for (i in 1:(digits - 1)) {
            yy <- sub("0.", "", y)
            sdigs <- nchar(as.numeric(yy))
            if (sdigs < digits) 
              y <- paste(y, "0", sep = "")
          }
          aux <- paste0("<", base::format((1/(10^pgits)), scientific = FALSE))
          y[as.numeric(y) < (1/(10^pgits))] <- aux
          rm(aux)
        }
        if (y == 1) {
          y <- "1."
          for (i in 1:digits) {
            y <- paste(y, "0", sep = "")
          }
        }
      }
      if (pgits%%1 == 0.5) {
        y <- base::format(x, digits = digits, scientific = FALSE)
        if (y != 1) {
          for (i in 1:(digits - 1)) {
            yy <- sub("0.", "", y)
            sdigs <- nchar(as.numeric(yy))
            if (sdigs < digits) 
              y <- paste(y, "0", sep = "")
          }
          aux <- paste0("<", base::format((1/(10^trunc(pgits)))/2, 
                                          scientific = FALSE))
          y[as.numeric(y) < (1/(10^trunc(pgits)))/2] <- aux
          rm(aux)
        }
        if (y == 1) {
          y <- "1."
          for (i in 1:digits) {
            y <- paste0(y, "0")
          }
        }
      }
    }
    if (digits == 1.5) {
      if (pgits%%1 == 0) {
        if (as.numeric(x) > 0.01) 
          y <- base::format(x, digits = 2, scientific = FALSE)
        if (as.numeric(x) <= 0.01) 
          y <- base::format(x, digits = 1, scientific = FALSE)
        if (y != 1) {
          if (y > 0.01) {
            yy <- sub("0.", "", y)
            sdigs <- nchar(as.numeric(yy))
            if (sdigs < 2) 
              y <- paste(y, "0", sep = "")
          }
          aux <- paste0("<", base::format((1/(10^pgits)), scientific = FALSE))
          y[as.numeric(y) < (1/(10^pgits))] <- aux
          rm(aux)
        }
        if (y == 1) 
          y <- "1.0"
      }
    }
    return(y)
  }
}

# ==============================================================================
# 11) Rounds a numeric value for pretty printing
#
#     This function is taken from R package 'epade' by Andreas Schulz, which at 
#     the time of writing was no longer available on CRAN repository.
#
#     ARGUMENTS
#     
#     x:	a numeric vector or a numeric matrix
#     digits: how many digits are to be shown after decimal?
# 
#     VALUE
#     
#     An object of similar structure to x containing character representations 
#     of the elements of x in a rounded format.
# ==============================================================================


round_n.ade <- function (x, digits = 0) {
  x <- unlist(x)
  if (!is.vector(x) & !is.matrix(x)) stop("x must be vector or matrix")
  if (length(x) > 1) {
    if (is.vector(x)) {
      y <- x
      for (i in 1:length(x)) {
        y[i] <- round_n.ade(x[i], digits)
      }
      return(y)
    }
    if (is.matrix(x)) {
      M <- x
      for (i in 1:dim(x)[1]) {
        for (j in 1:dim(x)[2]) {
          M[i, j] <- round_n.ade(x[i, j], digits)
        }
      }
      return(M)
    }
  }
  if (length(x) == 1) {
    if (is.na(x)) 
      return(x)
    if (x == 0) return(as.character(x))
    y <- base::format(base::round(x, digits = digits), nsmall = digits, 
                      scientific = FALSE)
    return(y)
  }
}

# ==============================================================================
# 12) Formats results of a single simulation scenario with misspecification. 
#
#     The Rds file containing the results of a simulation scenario is 
#     read and the results are processed into a single data frame to facilitate 
#     the production of figures and tables. 
#     
#     ARGUMENTS
#
#     scenario: the simulation scenario.
#     cols: names of columns in simulation results containing the estimated
#           probabilities that should be kept.
#     build_or_cal: 'build' refers to the model-building test data. 'cal'
#                    and 'cal_alt' refer to the two versions of the calibration 
#                    data (the 'cal_alt' train dataset has twice the number 
#                    of observations as the corresponding train dataset in 
#                    'cal').
#     reduce_repn: keeps only 'N_REPN' out of the 100 replications used in the 
#                  simulation scenario.
#     N_REPN: number of replications to to use. Used only if reduce_repn is set 
#             to TRUE.
#     mispec_param: type of parameter misspecification introduced. 
#
#     VALUE
#
#     A data frame containing the results of a simulation scenario.
#
# ==============================================================================

prepare_plot_df_mispec <- function(scenario, cols,
                                   build_or_cal = c("build", "cal", "cal_alt"),
                                   reduce_repn = FALSE, N_REPN = 5,
                                   mispec_param = c("mean", "mean-prop", "prop")) {
  
  build_or_cal <- match.arg(build_or_cal)
  mispec_param <- match.arg(mispec_param)
  # Reading results file 
  File <- here(paste0("rds-files/after-recalibration-misspec/", 
              "data-with-cal-probs--scen-", scenario, mispec_param,
                      "-mispec",".Rds"))
  results <- readRDS(file = File)
  
  
  data_list <- results[["data_list"]]
  repn <- length(data_list)
  dsub <- lapply(data_list, `[[`, build_or_cal) %>% 
    lapply(`[[`, "test") %>% do.call("rbind", .)
  
  if(reduce_repn) {
    dsub <- subset(dsub, repn <= N_REPN)
  }  
  dsub[["id"]] <- 1:nrow(dsub)
  dsub <- dsub[, c("id", "repn", "prob_true", cols)]
  dsub$scenario <- scenario
  
  pivot_longer(dsub, cols = {{cols}}, 
               names_to = "machine_and_recal_method",
               names_prefix = "prob_", 
               values_to = "Probability")
  
}
```
