## Supplementary material for "Calibrating machine learning approaches for probability estimation without calibration data": 05-fitting-machines-estimating-probs.R

```
# Supplementary Material 2 for 
# "Calibrating machine learning approaches for probability
# estimation in case of the absence of calibration data"

# This script is a modified version of the one found in supplementary material 
# 2 of Ojeda FM, et al., Calibrating machine learning approaches for probability 
# estimation: A comprehensive comparison. Stat Med. 2023 Dec 20;42(29):5451-5478
#
# The script does the following for all simulation scenarios considered:
#
# 1) Data is simulated
# 2) Machines are fitted (including tuning) and probabilities are estimated.
#
# Observe that no recalibration is performed at this point (with the exception 
# of refitting the machines). Only those probabilities that can be directly 
# estimated from the machines are computed.
# For the simulations presented in the manuscript 'repns_vector' below was
# set to '1:100' but for testing purposes, this may be set to a smaller sequence
# like '1:10'.
#
# While code and descriptions are available for all original 23 scenarios, 
# the analysis is performed on scenarios 1, 5, 9, 14, 16, 19, 20, 21
#
# To run this code, please open the supplementary material Rproject file.
# ==============================================================================

# Selected parameters to set when running analyses -----------------------------
repns_vector <- 1:100  # vector with indices of replications to use in simulation
cpus <- 10            # number of cpus for parallelization over repns_vector
grid_size <- 10 #50        # size of grid for tuning
folds_number <- 5     # number of partitions of data set (folds)
folds_repeats <- 5    # number of times to repeat the x-fold partitioning

# Attaching R packages ---------------------------------------------------------
# The following packages should be installed, but do not need to be attached:
# ranger, xgboost, kernlab, glmnet

library(here)
library(tidymodels)
library(dplyr)
library(stringr)
library(parallel)

# Loading functions ------------------------------------------------------------

source(here("Calibration_Sup2/01-data-generation-functions.R"))
source(here("Calibration_Sup2/02-machine-fitting-functions.R"))


################
#              #
#      ##      #
#      ##      #
#      ##      #
#   ########   #
#    ######    #
#     ###      #
#      #       #
#              #
################

# Parameters for simulations for each scenario.
# ==============================================================================

coeff_train <- list(c(0.5, 1),  # scenario 1
                    #c(0.5, 1),  # scenario 2
                    #c(0.5, 1),  # scenario 3
                    #c(0.5, 1),  # scenario 4                    
                    c(0.5, 1),  # scenario 5 (lower disease prevalence)
                    #c(0.5, 1),  # scenario 6 (lower disease prevalence)
                    #c(0.5, 1),  # scenario 7 (lower disease prevalence)
                    #c(0.5, 1),  # scenario 8 (lower disease prevalence)
                    c(0.1, 0.2, 0.3, 0.4, 0.5, 0.01, 0.02),#, 0.03, 0.04, 0.05), #9
                    #c(0.1, 0.2, 0.3, 0.4, 0.5, 0.01, 0.02, 0.03, 0.04, 0.05),
                    #c(0.1, 0.2, 0.3, 0.4, 0.5, 0.01, 0.02, 0.03, 0.04, 0.05),
                    #c(0.1, 0.2, 0.3, 0.4, 0.5, 0.01, 0.02, 0.03, 0.04, 0.05),
                    #NULL,       # scenario 13 (Mease model)
                    c(0.5, 1),  # scenario 14
                    #NULL,       # scenario 15 (Mease model)
                    1,          # scenario 16
                    #1,          # scenario 17
                    #1,          # scenario 18
                    1,          # scenario 19
                    1,          # scenario 20
                    1)          # scenario 21
                    #c(0.5, 1),  # scenario 22
                    #c(0.5, 1)  # scenario 23                    


coeff_test <- coeff_train
coeff_test[[7]] <- 3
coeff_test[[8]] <- (-1)

params_df <- data.frame(scenario = c(1,5,9,14,16,19,20,21), stringsAsFactors = FALSE)
params_df[["coeff_train"]] <- coeff_train
params_df[["coeff_test"]] <- coeff_test
desc <- c("-",                                                        # scen. 1
          #"10 noise variables",                                       # scen. 2
          #"50 noise variables",                                       # scen. 3
          #"100 noise variables",                                      # scen. 4
          "Lower disease prevalence",                                 # scen. 5
          #"Lower disease prevalence",                                 # scen. 6
          #"Lower disease prevalence",                                 # scen. 7
          #"Lower disease prevalence",                                 # scen. 8
          "Additional covariates",                                    # scen. 9
          #"Additional covariates plus noise variables",               # scen. 10
          #"Additional covariates plus noise variables",               # scen. 11
          #"Additional covariates plus noise variables",               # scen. 12
          #"Data generation using Mease model",                        # scen. 13
          "Smaller calibration data set for training",                # scen. 14
          #paste0("Smaller calibration data set for ",                 # scen. 15
          #       "training, data generation using Mease",
          #       "model"),
          "Covariate distribution: MB N(1, 1), Cal N(1, 1)",          # scen. 16
          #"Unequal covariate distribution: MB N(1, 1), Cal N(1, 2)",  # scen. 17
          #"Unequal covariate distribution: MB N(1, 1), Cal N(-1, 1)", # scen. 18
          "Unequal covariate distribution: MB N(1, 1), Cal N(-1, 2)", # scen. 19
          "Unequal coefficients: MB beta = 1, Cal beta = 3",          # scen. 20
          "Unequal coefficients: MB beta = 1, Cal beta = -1")         # scen. 21
          #paste0("Unequal covariate distribution for both continuous",
          #       " variables: MB N(1, 1), Cal N(-1, 1)"),
          #paste0("Unequal covariate distribution for one ",           # scen. 23
          #       "continuous variable: MB N(1, 1), Cal N(3, 1), ",
          #       "for the other continuous variable: MB N(1, 1), Cal N(-1, 1)")          
  


params_df[["description"]] <- desc; rm(desc)


params_df[["repn"]] <- repns_vector[length(repns_vector)]
params_df[["num_train"]] <- 1000
params_df[["num_test1"]] <- 1000
params_df[["num_test2"]] <- 1000
params_df[["num_test1"]][params_df$scenario %in% 14] <- 100

params_df[["intercept_train"]] <- 0
params_df[["intercept_test"]] <- 1

params_df[["mean_norm1_train"]] <- 0
params_df[["sd_norm1_train"]] <- 1
params_df[["mean_norm2_train"]] <- 0
params_df[["sd_norm2_train"]] <- 1

params_df[["mean_norm1_test"]] <- 0
params_df[["sd_norm1_test"]] <- 1
params_df[["mean_norm2_test"]] <- 0
params_df[["sd_norm2_test"]] <- 1

v <- which(params_df$scenario == 16)
params_df[["mean_norm1_train"]][v] <- 1
params_df[["mean_norm1_test"]][v] <- 1

# v <- which(params_df$scenario == 17)
# params_df[["mean_norm1_train"]][v] <- 1
# params_df[["mean_norm1_test"]][v] <- 1
# params_df[["sd_norm1_test"]][v] <- sqrt(2)

# v <- which(params_df$scenario == 18)
# params_df[["mean_norm1_train"]][v] <- 1
# params_df[["mean_norm1_test"]][v] <- (-1)

v <- which(params_df$scenario == 19)
params_df[["mean_norm1_train"]][v] <- 1
params_df[["mean_norm1_test"]][v] <- (-1)
params_df[["sd_norm1_test"]][v] <- sqrt(2)

# v <- which(params_df$scenario == 22)
# params_df[["mean_norm1_train"]][v] <- 1
# params_df[["mean_norm2_train"]][v] <- 1
# params_df[["mean_norm1_test"]][v] <- (-1)
# params_df[["mean_norm2_test"]][v] <- (-1)

# v <- which(params_df$scenario == 23)
# params_df[["mean_norm1_train"]][v] <- 1
# params_df[["mean_norm2_train"]][v] <- 1
# params_df[["mean_norm1_test"]][v] <- 3
# params_df[["mean_norm2_test"]][v] <- (-1)

params_df[["number_noise_variables"]] <- 0

# No noise variables:
# v <- which(params_df$scenario %in% c(2, 6, 10, 22, 23))
# params_df[["number_noise_variables"]][v] <- 10
# 
# v <- which(params_df$scenario %in% c(3, 7, 11))
# params_df[["number_noise_variables"]][v] <- 50
# 
# v <- which(params_df$scenario %in% c(4, 8, 12))
# params_df[["number_noise_variables"]][v] <- 100

params_df[["seed"]] <- 202108

rm(coeff_test, coeff_train, v)

################
#              #
#      ##      #
#      ##      #
#      ##      #
#   ########   #
#    ######    #
#     ###      #
#      #       #
#              #
################

# Machine specifications.
# Tuning grids are specified later since they may depend on the particular 
# dataset used. 
# ==============================================================================
spec_lr <- logistic_reg() %>% 
  set_engine("glm") %>% 
  set_mode("classification")

# spec_en <- logistic_reg(penalty = tune(),
#                         mixture = tune()) %>%
#   set_engine("glmnet", family = "binomial") %>%
#   set_mode("classification")
# 
# spec_rf <- rand_forest(mtry = tune(), trees = 500,
#                        min_n = tune()) %>%
#   set_engine("ranger") %>%
#   set_mode("classification") %>% set_args(seed = NULL)
# 
# spec_boost <- boost_tree(tree_depth = tune(), trees = tune(),
#                          learn_rate = tune(), loss_reduction = tune(),
#                          mtry = tune(), sample_size = tune(),
#                          min_n = tune()) %>%
#   set_engine("xgboost") %>%
#   set_mode("classification")
# 
# spec_svm <- svm_rbf(cost = tune(), rbf_sigma = tune()) %>%
#   set_engine("kernlab", scaled = TRUE) %>%
#   set_mode("classification")

machine_abbreviations <- c("lr")
machine_abbreviations_tune <- c()


machines_spec_info <- list(session_info = sessionInfo(),
                           date_saved = Sys.time(), 
                           r_version = R.Version(),
                           system_info = Sys.info())

################
#              #
#      ##      #
#      ##      #
#      ##      #
#   ########   #
#    ######    #
#     ###      #
#      #       #
#              #
################


# Altering functions to add a predicted probability column to df_build_train for
# future use of calibration methods such as sba and acp.

add_empty_prob_columns <- function() {
  
  objects_to_get <- c("simu_data", "machine_abbreviations")
  for(i in objects_to_get) assign(i, get(i, envir = parent.frame())) 
  
  for(machine_abbr in machine_abbreviations) {
    prob_col <- paste0("prob_", machine_abbr)
    prob_col_refit <- paste0(prob_col, "_refit")
    
    simu_data[["build"]][["train"]][[prob_col]] <- NA_real_
    simu_data[["build"]][["test"]][[prob_col]] <- NA_real_
    
    for(cal_name in c("cal", "cal_alt")) {
      simu_data[[cal_name]][["train"]][[prob_col]] <- NA_real_
      simu_data[[cal_name]][["test"]][[prob_col]] <- NA_real_
      simu_data[[cal_name]][["test"]][[prob_col_refit]] <- NA_real_      
    }
    
    rm(prob_col, prob_col_refit)
  }
  
  assign("simu_data", simu_data, envir = parent.frame())
  
  invisible(NULL)
}


add_predictions <- function() {
  
  objects_to_get <- c("machine_abbr", "fit_build", "fit_cal", "fit_cal_alt")
  for(i in objects_to_get) assign(i, get(i, envir = parent.frame()))  
  
  prob_col <- paste0("prob_", machine_abbr)
  
  for(data_name in c("df_build_train", "df_build_test",  "df_cal_train", "df_cal_test",
                     "df_cal_alt_train", "df_cal_alt_test")) {
    df_current <- get(data_name, envir = parent.frame())
    
    aux <- tryCatch(predict(fit_build, 
                            new_data = df_current, 
                            type = "prob")[[".pred_1"]], error = identity)
    
    if("simpleError" %in% class(aux)) aux <- rep(NA_real_, nrow(df_current))
    
    df_current[[prob_col]] <- aux; rm(aux)
    
    if(data_name == "df_cal_test") {
      aux <- tryCatch(predict(fit_cal, new_data = df_current, 
                              type = "prob")[[".pred_1"]], error = identity)
      
      if("simpleError" %in% class(aux)) aux <- rep(NA_real_, nrow(df_current))
      
      df_current[[paste0(prob_col, "_refit")]] <- aux; rm(aux)      
    }
    
    if(data_name == "df_cal_alt_test") {
      aux <-tryCatch(predict(fit_cal_alt, new_data = df_current, 
                             type = "prob")[[".pred_1"]], error = identity)
      
      if("simpleError" %in% class(aux)) aux <- rep(NA_real_, nrow(df_current))
      
      df_current[[paste0(prob_col, "_refit")]] <- aux; rm(aux)      
    }    
    
    assign(data_name, df_current, envir = parent.frame())
    rm(df_current)
  }
  
  invisible(NULL)  
  
}

# Fitting machines.
# Computations are parallelized within each scenario over the 
# replications (repns_vector).
#
# The function 'simu_fit_predict' does the following for a given scenario and 
# replication:
#
# a) generates the required data frames
# b) fits the machines
# c) computes the predicted probabilities (from the machines)
# 
# The results are saved in the folder 'here("rds-files/before-recalibration")'.
# ==============================================================================
 
simu_fit_predict <- function(repn) {
  
  
  simu_data <- simulate_data_wrapper(scenario = scenario, params_df, 
                                     repn = repn)
  
  
  # if(scenario %in% 16:21) {
  #   # glmnet does not support a single predictor. For that reason the elastic
  #   # net is not computed for those scenarios using only one predictor 
  #   # (including noise variables)
  #   machine_abbreviations <- setdiff(machine_abbreviations, "en") 
  #   machine_abbreviations_tune <- setdiff(machine_abbreviations_tune, "en")
  #   
  # }
  
  add_empty_prob_columns()
  create_formula()
  
  #create_grids_and_workflows()
  extract_train_test_dfs()
  
  folds_build_test <- vfold_cv(df_build_test, v = folds_number, 
                          repeats = folds_repeats)
  folds_build <- vfold_cv(df_build_train, v = folds_number, 
                          repeats = folds_repeats)
  folds_cal <- vfold_cv(df_cal_train, v = folds_number, repeats = folds_repeats)
  folds_cal_alt <- vfold_cv(df_cal_alt_train, v = folds_number, 
                            repeats = folds_repeats)
  #tuning_machines()
  
  for(machine_abbr in machine_abbreviations) {
    fitting_machines()
    add_predictions()
    rm(fit_build, fit_cal, fit_cal_alt)
  }
  
  update_train_test_dfs()
  
  list(data = simu_data, warn = warnings())
  
}  


for(scenario in c(1,5,9,14,16,19,20,21)) {
  
  begin_time <- Sys.time()
  
  cl <- makeCluster(cpus)
  clusterEvalQ(cl, {
    library(tidymodels)
    library(dplyr)
    library(stringr)
  })
  clusterSetRNGStream(cl, iseed = params_df[["seed"]][1])
  clusterExport(cl, varlist = c("scenario", "params_df", "grid_size",
                                "simulate_data", "simulate_data_wrapper",
                                paste0("spec_", machine_abbreviations), 
                                "machines_spec_info",
                                "machine_abbreviations", 
                                "machine_abbreviations_tune",
                                "folds_number", "folds_repeats",
                                "add_empty_prob_columns", "create_formula",
                                "create_grids_and_workflows",
                                "tuning_machines",
                                "fitting_machines", "extract_train_test_dfs",
                                "add_predictions", "update_train_test_dfs")) 
  results <- parLapply(cl, repns_vector, simu_fit_predict)
  stopCluster(cl)
  
  end_time <- Sys.time()
  run_time <- end_time - begin_time
  
  results <- list(data_list = lapply(results, `[[`, "data"), 
                  folds_number = folds_number,
                  folds_repeats = folds_repeats,
                  run_time = run_time, 
                  session_info = sessionInfo(),
                  date_saved = Sys.time(), 
                  r_version = R.Version(),
                  system_info = Sys.info(),
                  warning_list = lapply(results, `[[`, "warn"))
  
  File <- here("rds-files/before-recalibration", 
               paste0("data-with-probs--scen-", scenario, ".Rds"))
  saveRDS(object = results, file = File) 
  
  print("#################################")
  print(Sys.time())
  mssg <- paste("Done with scenario", scenario)
  print(mssg)
  
  rm(results, run_time, begin_time, end_time, mssg)
  
}
```
