## Supplementary material for "Calibrating machine learning approaches for probability estimation without calibration data": 06-calibrating-predictions.R

```
# Supplementary Material 2 for 
# "Calibrating machine learning approaches for probability
# estimation in case of the absence of calibration data"

# This script is a modified version of the one found in supplementary material 
# 2 of Ojeda FM, et al., Calibrating machine learning approaches for probability 
# estimation: A comprehensive comparison. Stat Med. 2023 Dec 20;42(29):5451-5478

# This script does the following for all simulation scenarios considered:
#
# 1) For the simulated data the estimated probabilities are calibrated using
#    several calibration methods.
#
#    Refitting of the machines is not done by this script. This was already done
#    by '05-fitting-machines-estimating-probs.R'.
#
# While code and descriptions are available for all original 23 scenarios, 
# the analysis is performed on scenarios 1, 5, 9, 14, 16, 19, 20, 21
#
# To run this code, please open the supplementary material Rproject file.
#
# ==============================================================================
# Attaching R packages ---------------------------------------------------------
library(here)
library(FNN)
library(dplyr)
library(stringr)
library(rms)
library(modgo)
library(dqrng)
library(parsnip)
library(epade)
# Loading functions ------------------------------------------------------------
source(here("Calibration_Sup2/03-calibration-functions.R"))
source(here("Calibration_Sup2/02-machine-fitting-functions.R"))

################
#              #
#      ##      #
#      ##      #
#      ##      #
#   ########   #
#    ######    #
#     ###      #
#      #       #
#              #
################

# Recalibrating probabilities in all simulation scenarios.
# ==============================================================================

# Leaving BOTH parameters set to FALSE generates the data necessary for 
# supplementary material 3a

# Set to TRUE to generate calibrated probabilities with misspecified means. 
# Setting ONLY this parameter to TRUE generates the data necessary
# for supplementary material 4a
MISSPEC_MEAN <- FALSE 

# Set to TRUE to generate calibrated probabilities with misspecified proportions. 
# Setting ONLY this parameter to TRUE generates the data necessary 
# for supplementary material 5a
MISSPEC_PROP <- FALSE

# Setting BOTH parameters to TRUE generates the data necessary for 
# supplementary material 6a

machine_abbreviations <- c("lr")

if (MISSPEC_MEAN | MISSPEC_PROP) {
  calibration_names <- c("elkan","simcal", "elkan_mispec", "simcal_mispec")
  scenarios <- c(16)
} else {
  # Roughly 10 minutes per scenario, 8 scenarios
  calibration_names <- c("logistic", "elkan", "simcal", "sba10", "acp")
  scenarios <- c(1, 5, 9, 14, 16, 19, 20, 21)
}

if (MISSPEC_MEAN == TRUE & MISSPEC_PROP == FALSE) {
  # Roughly 3-4 minutes per scenario, 10 scenarios
  mean_misspec_params <- c(1.1, 1.2, 1.3, 1.4, 1.5, 1.6, 1.7, 1.8, 1.9, 2)
  naming_index <- paste0("16.", 1:10)
  length_params <- length(mean_misspec_params)
}

if (MISSPEC_PROP == TRUE & MISSPEC_MEAN == FALSE) {
  # Roughly 3-4 minutes per scenario, 8 scenarios
  prop_misspec_params <- c(0.76, 0.78, 0.80, 0.82, 0.86, 0.88, 0.90, 0.92)
  naming_index <- c(16.1, 16.2, 16.3, 16.4, 16.5, 16.6, 16.7, 16.8)
  length_params <- length(prop_misspec_params)
}

if (MISSPEC_PROP == TRUE & MISSPEC_MEAN == TRUE) {
  # Roughly 3-4 minutes per scenario, 99 scenarios
  prop_misspec_params <- c(0.56, 0.60, 0.64, 0.68, 0.72, 0.76, 0.80, 0.84, 
                           0.88, 0.92, 0.96)
  mean_misspec_params <- c(0, 0.25, 0.5, 0.75, 1, 1.25, 1.5, 1.75, 2)
  naming_index <- paste0("16.", 1:99)
  length_params <- length(naming_index)
}


wrapper_misspec <- function(scenario) {
  

  for(i in 1:length_params) {
  
  begin_time <- Sys.time()
  
  # if(scenario %in% 16:21) {
  #   machine_abbreviations <- setdiff(machine_abbreviations, "en")
  # }
  File <- here(paste0("rds-files/before-recalibration/", 
               "data-with-probs--scen-", scenario, ".Rds"))
  
  results <- readRDS(file = File)
  
  # Specifies misspecification of c(mean, proportion)
  if(MISSPEC_MEAN == TRUE & MISSPEC_PROP == FALSE) {
    mispec <<- c(mean_misspec_params[i], "none", "none")
  } else if (MISSPEC_PROP == TRUE & MISSPEC_MEAN == FALSE) {
    mispec <<- c("none", "none", prop_misspec_params[i])
  } else if (MISSPEC_MEAN == TRUE & MISSPEC_PROP == TRUE) {
    
    grid <- expand.grid(mean = mean_misspec_params,
                        prop = prop_misspec_params)
    
    mispec_list <- lapply(seq_len(nrow(grid)), function(j) {
      c(grid$mean[j], "none", grid$prop[j])
    })
    
    mispec <<- mispec_list[[i]]
  }
  
  repns <- 1:length(results[["data_list"]])
  
  add_empty_cal_prob_columns()
  create_prev_epv_rcs_matrices()
  
  for(repn_number in repns) {
    print("# ========================")
    print(paste("Calibrating rep",repn_number,"in scenario", naming_index[i]))
    
    simu_data <- results[["data_list"]][[repn_number]]
    extract_train_test_dfs()
    compute_prev_epv_and_update_matrix()
    for(machine_abbr in machine_abbreviations) {  
      calibrate_preds()
    }
    update_cal_prob_columns()
  }  
  
  end_time <- Sys.time()
  session_info <- sessionInfo()
  run_time <- end_time - begin_time
  date_saved <- end_time
  r_version <- R.Version()
  system_info <- Sys.info()
  
  
  calibration_info <- list(run_time = run_time, 
                           session_info = session_info,
                           date_saved = date_saved, 
                           r_version = r_version,
                           system_info = system_info)
  objects_results <- c("calibration_info", "matrix_prev_epv", 
                       paste0("rcs_info_cal_", machine_abbreviations),
                       paste0("rcs_info_cal_alt_", machine_abbreviations),
                       paste0("chen_info_cal_", machine_abbreviations),
                       paste0("chen_info_cal_alt_", machine_abbreviations))
  for(j in objects_results) {
    results[[j]] <- get(j)    
  }
  
  suffix_app <- paste0(
    ifelse(MISSPEC_MEAN & !MISSPEC_PROP, "mean",
           ifelse(MISSPEC_PROP & !MISSPEC_MEAN, "prop",
                  ifelse(MISSPEC_MEAN & MISSPEC_PROP, "mean-prop", "")))
  )
  
  File <- here(paste0("rds-files/after-recalibration", if_else(MISSPEC_MEAN | MISSPEC_PROP, "-misspec/","/"), 
               "data-with-cal-probs--scen-", naming_index[i], 
               suffix_app,"-mispec.Rds")
               )

  saveRDS(object = results, file = File)
  
  print("# ========================")
  print(paste("Done re-calibrating probabilities for scenario", naming_index[i]))
  print(Sys.time())  
  
  
  invisible(NULL)  
  }
}


wrapper <- function(scenario) {
  
    
    begin_time <- Sys.time()
    
    # if(scenario %in% 16:21) {
    #   machine_abbreviations <- setdiff(machine_abbreviations, "en")
    # }
    File <- here(paste0("rds-files/before-recalibration/", 
                        "data-with-probs--scen-", scenario, ".Rds"))
    
    results <- readRDS(file = File)
    
    repns <- 1:length(results[["data_list"]])
    
    add_empty_cal_prob_columns()
    create_prev_epv_rcs_matrices()
    
    for(repn_number in repns) {
      print("# ========================")
      print(paste("Calibrating rep",repn_number,"in scenario", scenario))
      
      simu_data <- results[["data_list"]][[repn_number]]
      extract_train_test_dfs()
      compute_prev_epv_and_update_matrix()
      for(machine_abbr in machine_abbreviations) {  
        calibrate_preds()
      }
      update_cal_prob_columns()
    }  
    
    end_time <- Sys.time()
    session_info <- sessionInfo()
    run_time <- end_time - begin_time
    date_saved <- end_time
    r_version <- R.Version()
    system_info <- Sys.info()
    
    
    calibration_info <- list(run_time = run_time, 
                             session_info = session_info,
                             date_saved = date_saved, 
                             r_version = r_version,
                             system_info = system_info)
    objects_results <- c("calibration_info", "matrix_prev_epv", 
                         paste0("rcs_info_cal_", machine_abbreviations),
                         paste0("rcs_info_cal_alt_", machine_abbreviations),
                         paste0("chen_info_cal_", machine_abbreviations),
                         paste0("chen_info_cal_alt_", machine_abbreviations))
    
    for(i in objects_results) {
      results[[i]] <- get(i)    
    }
  
    
    File <- here(paste0("rds-files/after-recalibration", if_else(MISSPEC_MEAN | MISSPEC_PROP, "-misspec/","/"), 
                        "data-with-cal-probs--scen-", scenario,".Rds")
    )
    
    saveRDS(object = results, file = File)
    
    print("# ========================")
    print(paste("Done re-calibrating probabilities for scenario", scenario))
    print(Sys.time())  
    
    
    invisible(NULL)  
  
}


# Running wrapper function -----------------------------------------------------
if (MISSPEC_MEAN | MISSPEC_PROP) {
  lapply(scenarios, wrapper_misspec)
} else {
  lapply(scenarios, wrapper)
}
```
