## Supplementary material for "Calibrating machine learning approaches for probability estimation without calibration data": Supplementary_material_3b.html

Simulation study 1: performance and comparison of new calibrating machine learning approach (SimCal) for probability estimation.


### Simulation study 1: performance and comparison of new calibrating machine learning approach (SimCal) for probability estimation.

##### Supplementary material 3b

###### Eleonora Di Carluccio, Francisco M. Ojeda

#### 2026-06-24

### 1 About this file

This html file contains tables and figures presenting results of the
simulation study conducted to compare the performance of different
calibration approaches.

Data from two populations with different disease prevalences were
generated. The data from the first population were used for model
building. Data from the second population were split into training
(cal-training) data and test (cal-test) data.

In all simulation scenarios the sample size of the model building data
and cal-test data was set to 1000 observations.

In (this file) results are
presented where the number of observations in the cal-training data was
set to 1000 for all scenarios, except simulation scenarios 14 where only
100 observations were used.

### 2 Simulation scenarios

On this section a table summarizing all simulation scenarios is
presented.

Table 2.1: Table 2.2: Main settings in the simulation scenarios: Number of continuous covariates (Continuous), number of categorical covariates (Categorical), number of noise variables (Noise), model building data set sample size (MB N), calibration training data set sample size (Cal-training N), calibration test data set sample size (Cal-test N), model building data set outcome prevalence (MB prevalence), calibration training data set outcome prevalence (Cal-training prevalence), model building data set number of events per variable (MB EVP), calibration training data set number of events per variable (Cal-training EVP) and, if applicable, additional distinctive feature (Distinctive feature).

| Scenario | Continuous | Categorical | Noise | MB N | Cal-training N | Cal-test N | MB prevalence | Cal-training prevalence | MB EPV | Cal-training EPV | Distinctive feature |
| --- | --- | --- | --- | --- | --- | --- | --- | --- | --- | --- | --- |
| 1 | 2 | 0 | 0 | 1000 | 1000 | 1000 | 0.5 | 0.69 | 243 | 156.2 |  |
| 5 | 2 | 0 | 0 | 1000 | 1000 | 1000 | 0.2 | 0.69 | 100.3 | 156.2 | Lower disease prevalence |
| 9 | 5 | 2 | 0 | 1000 | 1000 | 1000 | 0.51 | 0.71 | 69.6 | 41 | Additional covariates |
| 14 | 2 | 0 | 0 | 1000 | 100 | 1000 | 0.5 | 0.7 | 243.4 | 15.2 | Smaller calibration data set for training |
| 16 | 1 | 0 | 0 | 1000 | 1000 | 1000 | 0.7 | 0.84 | 303.1 | 156.4 | Covariate distribution: MB N(1, 1), Cal N(1, 1) |
| 19 | 1 | 0 | 0 | 1000 | 1000 | 1000 | 0.7 | 0.5 | 303.1 | 485.6 | Unequal covariate distribution: MB N(1, 1), Cal N(-1, 2) |
| 20 | 1 | 0 | 0 | 1000 | 1000 | 1000 | 0.5 | 0.61 | 488.4 | 385.8 | Unequal coefficients: MB beta = 1, Cal beta = 3 |
| 21 | 1 | 0 | 0 | 1000 | 1000 | 1000 | 0.5 | 0.7 | 488.4 | 304.6 | Unequal coefficients: MB beta = 1, Cal beta = -1 |
|  |  |  |  |  |  |  |  |  |  |  |  |
| --- | --- | --- | --- | --- | --- | --- | --- | --- | --- | --- | --- |
| MB: Model building data; EPV: events per variable. |  |  |  |  |  |  |  |  |  |  |  |

### 3 Calibration plots

#### 3.1 Logistic regression

##### 3.1.1 Logistic regression scenario 1

Figure 3.1: Calibration plot for logistic regression for simulation scenario 1. GAM is used as scatterplot smoother over all 100 replications to estimate the calibration curve.

##### 3.1.2 Logistic regression scenario 5

Figure 3.2: Calibration plot for logistic regression for simulation scenario 5. GAM is used as scatterplot smoother over all 100 replications to estimate the calibration curve.

##### 3.1.3 Logistic regression scenario 9

Figure 3.3: Calibration plot for logistic regression for simulation scenario 9. GAM is used as scatterplot smoother over all 100 replications to estimate the calibration curve.

##### 3.1.4 Logistic regression scenario 14

Figure 3.4: Calibration plot for logistic regression for simulation scenario 14. GAM is used as scatterplot smoother over all 100 replications to estimate the calibration curve.

##### 3.1.5 Logistic regression scenario 16

Figure 3.5: Calibration plot for logistic regression for simulation scenario 16. GAM is used as scatterplot smoother over all 100 replications to estimate the calibration curve.

##### 3.1.6 Logistic regression scenario 19

Figure 3.6: Calibration plot for logistic regression for simulation scenario 19. GAM is used as scatterplot smoother over all 100 replications to estimate the calibration curve.

##### 3.1.7 Logistic regression scenario 20

Figure 3.7: Calibration plot for logistic regression for simulation scenario 20. GAM is used as scatterplot smoother over all 100 replications to estimate the calibration curve.

##### 3.1.8 Logistic regression scenario 21

Figure 3.8: Calibration plot for logistic regression for simulation scenario 21. GAM is used as scatterplot smoother over all 100 replications to estimate the calibration curve.

#### 3.2 Logistic regression single repetitions

##### 3.2.1 lr repetition number 1

Figure 3.9: Calibration plot for logistic regression for simulation scenario 5. GAM is used as scatterplot smoother over repetition number 1 to estimate the calibration curve.

##### 3.2.2 lr repetition number 2

Figure 3.10: Calibration plot for logistic regression for simulation scenario 5. GAM is used as scatterplot smoother over repetition number 2 to estimate the calibration curve.

##### 3.2.3 lr repetition number 3

Figure 3.11: Calibration plot for logistic regression for simulation scenario 5. GAM is used as scatterplot smoother over repetition number 3 to estimate the calibration curve.

##### 3.2.4 lr repetition number 4

Figure 3.12: Calibration plot for logistic regression for simulation scenario 5. GAM is used as scatterplot smoother over repetition number 4 to estimate the calibration curve.

##### 3.2.5 lr repetition number 5

Figure 3.13: Calibration plot for logistic regression for simulation scenario 5. GAM is used as scatterplot smoother over repetition number 5 to estimate the calibration curve.

##### 3.2.6 lr repetition number 6

Figure 3.14: Calibration plot for logistic regression for simulation scenario 5. GAM is used as scatterplot smoother over repetition number 6 to estimate the calibration curve.

##### 3.2.7 lr repetition number 7

Figure 3.15: Calibration plot for logistic regression for simulation scenario 5. GAM is used as scatterplot smoother over repetition number 7 to estimate the calibration curve.

##### 3.2.8 lr repetition number 8

Figure 3.16: Calibration plot for logistic regression for simulation scenario 5. GAM is used as scatterplot smoother over repetition number 8 to estimate the calibration curve.

##### 3.2.9 lr repetition number 9

Figure 3.17: Calibration plot for logistic regression for simulation scenario 5. GAM is used as scatterplot smoother over repetition number 9 to estimate the calibration curve.

##### 3.2.10 lr repetition number 10

Figure 3.18: Calibration plot for logistic regression for simulation scenario 5. GAM is used as scatterplot smoother over repetition number 10 to estimate the calibration curve.

##### 3.2.11 lr repetition number 11

Figure 3.19: Calibration plot for logistic regression for simulation scenario 5. GAM is used as scatterplot smoother over repetition number 11 to estimate the calibration curve.

##### 3.2.12 lr repetition number 12

Figure 3.20: Calibration plot for logistic regression for simulation scenario 5. GAM is used as scatterplot smoother over repetition number 12 to estimate the calibration curve.

##### 3.2.13 lr repetition number 13

Figure 3.21: Calibration plot for logistic regression for simulation scenario 5. GAM is used as scatterplot smoother over repetition number 13 to estimate the calibration curve.

##### 3.2.14 lr repetition number 14

Figure 3.22: Calibration plot for logistic regression for simulation scenario 5. GAM is used as scatterplot smoother over repetition number 14 to estimate the calibration curve.

##### 3.2.15 lr repetition number 15

Figure 3.23: Calibration plot for logistic regression for simulation scenario 5. GAM is used as scatterplot smoother over repetition number 15 to estimate the calibration curve.

##### 3.2.16 lr repetition number 16

Figure 3.24: Calibration plot for logistic regression for simulation scenario 5. GAM is used as scatterplot smoother over repetition number 16 to estimate the calibration curve.

##### 3.2.17 lr repetition number 17

Figure 3.25: Calibration plot for logistic regression for simulation scenario 5. GAM is used as scatterplot smoother over repetition number 17 to estimate the calibration curve.

##### 3.2.18 lr repetition number 18

Figure 3.26: Calibration plot for logistic regression for simulation scenario 5. GAM is used as scatterplot smoother over repetition number 18 to estimate the calibration curve.

##### 3.2.19 lr repetition number 19

Figure 3.27: Calibration plot for logistic regression for simulation scenario 5. GAM is used as scatterplot smoother over repetition number 19 to estimate the calibration curve.

##### 3.2.20 lr repetition number 20

Figure 3.28: Calibration plot for logistic regression for simulation scenario 5. GAM is used as scatterplot smoother over repetition number 20 to estimate the calibration curve.

##### 3.2.21 lr repetition number 21

Figure 3.29: Calibration plot for logistic regression for simulation scenario 5. GAM is used as scatterplot smoother over repetition number 21 to estimate the calibration curve.

##### 3.2.22 lr repetition number 22

Figure 3.30: Calibration plot for logistic regression for simulation scenario 5. GAM is used as scatterplot smoother over repetition number 22 to estimate the calibration curve.

##### 3.2.23 lr repetition number 23

Figure 3.31: Calibration plot for logistic regression for simulation scenario 5. GAM is used as scatterplot smoother over repetition number 23 to estimate the calibration curve.

##### 3.2.24 lr repetition number 24

Figure 3.32: Calibration plot for logistic regression for simulation scenario 5. GAM is used as scatterplot smoother over repetition number 24 to estimate the calibration curve.

##### 3.2.25 lr repetition number 25

Figure 3.33: Calibration plot for logistic regression for simulation scenario 5. GAM is used as scatterplot smoother over repetition number 25 to estimate the calibration curve.

##### 3.2.26 lr repetition number 26

Figure 3.34: Calibration plot for logistic regression for simulation scenario 5. GAM is used as scatterplot smoother over repetition number 26 to estimate the calibration curve.

##### 3.2.27 lr repetition number 27

Figure 3.35: Calibration plot for logistic regression for simulation scenario 5. GAM is used as scatterplot smoother over repetition number 27 to estimate the calibration curve.

##### 3.2.28 lr repetition number 28

Figure 3.36: Calibration plot for logistic regression for simulation scenario 5. GAM is used as scatterplot smoother over repetition number 28 to estimate the calibration curve.

##### 3.2.29 lr repetition number 29

Figure 3.37: Calibration plot for logistic regression for simulation scenario 5. GAM is used as scatterplot smoother over repetition number 29 to estimate the calibration curve.

##### 3.2.30 lr repetition number 30

Figure 3.38: Calibration plot for logistic regression for simulation scenario 5. GAM is used as scatterplot smoother over repetition number 30 to estimate the calibration curve.

##### 3.2.31 lr repetition number 31

Figure 3.39: Calibration plot for logistic regression for simulation scenario 5. GAM is used as scatterplot smoother over repetition number 31 to estimate the calibration curve.

##### 3.2.32 lr repetition number 32

Figure 3.40: Calibration plot for logistic regression for simulation scenario 5. GAM is used as scatterplot smoother over repetition number 32 to estimate the calibration curve.

##### 3.2.33 lr repetition number 33

Figure 3.41: Calibration plot for logistic regression for simulation scenario 5. GAM is used as scatterplot smoother over repetition number 33 to estimate the calibration curve.

##### 3.2.34 lr repetition number 34

Figure 3.42: Calibration plot for logistic regression for simulation scenario 5. GAM is used as scatterplot smoother over repetition number 34 to estimate the calibration curve.

##### 3.2.35 lr repetition number 35

Figure 3.43: Calibration plot for logistic regression for simulation scenario 5. GAM is used as scatterplot smoother over repetition number 35 to estimate the calibration curve.

##### 3.2.36 lr repetition number 36

Figure 3.44: Calibration plot for logistic regression for simulation scenario 5. GAM is used as scatterplot smoother over repetition number 36 to estimate the calibration curve.

##### 3.2.37 lr repetition number 37

Figure 3.45: Calibration plot for logistic regression for simulation scenario 5. GAM is used as scatterplot smoother over repetition number 37 to estimate the calibration curve.

##### 3.2.38 lr repetition number 38

Figure 3.46: Calibration plot for logistic regression for simulation scenario 5. GAM is used as scatterplot smoother over repetition number 38 to estimate the calibration curve.

##### 3.2.39 lr repetition number 39

Figure 3.47: Calibration plot for logistic regression for simulation scenario 5. GAM is used as scatterplot smoother over repetition number 39 to estimate the calibration curve.

##### 3.2.40 lr repetition number 40

Figure 3.48: Calibration plot for logistic regression for simulation scenario 5. GAM is used as scatterplot smoother over repetition number 40 to estimate the calibration curve.

##### 3.2.41 lr repetition number 41

Figure 3.49: Calibration plot for logistic regression for simulation scenario 5. GAM is used as scatterplot smoother over repetition number 41 to estimate the calibration curve.

##### 3.2.42 lr repetition number 42

Figure 3.50: Calibration plot for logistic regression for simulation scenario 5. GAM is used as scatterplot smoother over repetition number 42 to estimate the calibration curve.

##### 3.2.43 lr repetition number 43

Figure 3.51: Calibration plot for logistic regression for simulation scenario 5. GAM is used as scatterplot smoother over repetition number 43 to estimate the calibration curve.

##### 3.2.44 lr repetition number 44

Figure 3.52: Calibration plot for logistic regression for simulation scenario 5. GAM is used as scatterplot smoother over repetition number 44 to estimate the calibration curve.

##### 3.2.45 lr repetition number 45

Figure 3.53: Calibration plot for logistic regression for simulation scenario 5. GAM is used as scatterplot smoother over repetition number 45 to estimate the calibration curve.

##### 3.2.46 lr repetition number 46

Figure 3.54: Calibration plot for logistic regression for simulation scenario 5. GAM is used as scatterplot smoother over repetition number 46 to estimate the calibration curve.

##### 3.2.47 lr repetition number 47

Figure 3.55: Calibration plot for logistic regression for simulation scenario 5. GAM is used as scatterplot smoother over repetition number 47 to estimate the calibration curve.

##### 3.2.48 lr repetition number 48

Figure 3.56: Calibration plot for logistic regression for simulation scenario 5. GAM is used as scatterplot smoother over repetition number 48 to estimate the calibration curve.

##### 3.2.49 lr repetition number 49

Figure 3.57: Calibration plot for logistic regression for simulation scenario 5. GAM is used as scatterplot smoother over repetition number 49 to estimate the calibration curve.

##### 3.2.50 lr repetition number 50

Figure 3.58: Calibration plot for logistic regression for simulation scenario 5. GAM is used as scatterplot smoother over repetition number 50 to estimate the calibration curve.

##### 3.2.51 lr repetition number 51

Figure 3.59: Calibration plot for logistic regression for simulation scenario 5. GAM is used as scatterplot smoother over repetition number 51 to estimate the calibration curve.

##### 3.2.52 lr repetition number 52

Figure 3.60: Calibration plot for logistic regression for simulation scenario 5. GAM is used as scatterplot smoother over repetition number 52 to estimate the calibration curve.

##### 3.2.53 lr repetition number 53

Figure 3.61: Calibration plot for logistic regression for simulation scenario 5. GAM is used as scatterplot smoother over repetition number 53 to estimate the calibration curve.

##### 3.2.54 lr repetition number 54

Figure 3.62: Calibration plot for logistic regression for simulation scenario 5. GAM is used as scatterplot smoother over repetition number 54 to estimate the calibration curve.

##### 3.2.55 lr repetition number 55

Figure 3.63: Calibration plot for logistic regression for simulation scenario 5. GAM is used as scatterplot smoother over repetition number 55 to estimate the calibration curve.

##### 3.2.56 lr repetition number 56

Figure 3.64: Calibration plot for logistic regression for simulation scenario 5. GAM is used as scatterplot smoother over repetition number 56 to estimate the calibration curve.

##### 3.2.57 lr repetition number 57

Figure 3.65: Calibration plot for logistic regression for simulation scenario 5. GAM is used as scatterplot smoother over repetition number 57 to estimate the calibration curve.

##### 3.2.58 lr repetition number 58

Figure 3.66: Calibration plot for logistic regression for simulation scenario 5. GAM is used as scatterplot smoother over repetition number 58 to estimate the calibration curve.

##### 3.2.59 lr repetition number 59

Figure 3.67: Calibration plot for logistic regression for simulation scenario 5. GAM is used as scatterplot smoother over repetition number 59 to estimate the calibration curve.

##### 3.2.60 lr repetition number 60

Figure 3.68: Calibration plot for logistic regression for simulation scenario 5. GAM is used as scatterplot smoother over repetition number 60 to estimate the calibration curve.

##### 3.2.61 lr repetition number 61

Figure 3.69: Calibration plot for logistic regression for simulation scenario 5. GAM is used as scatterplot smoother over repetition number 61 to estimate the calibration curve.

##### 3.2.62 lr repetition number 62

Figure 3.70: Calibration plot for logistic regression for simulation scenario 5. GAM is used as scatterplot smoother over repetition number 62 to estimate the calibration curve.

##### 3.2.63 lr repetition number 63

Figure 3.71: Calibration plot for logistic regression for simulation scenario 5. GAM is used as scatterplot smoother over repetition number 63 to estimate the calibration curve.

##### 3.2.64 lr repetition number 64

Figure 3.72: Calibration plot for logistic regression for simulation scenario 5. GAM is used as scatterplot smoother over repetition number 64 to estimate the calibration curve.

##### 3.2.65 lr repetition number 65

Figure 3.73: Calibration plot for logistic regression for simulation scenario 5. GAM is used as scatterplot smoother over repetition number 65 to estimate the calibration curve.

##### 3.2.66 lr repetition number 66

Figure 3.74: Calibration plot for logistic regression for simulation scenario 5. GAM is used as scatterplot smoother over repetition number 66 to estimate the calibration curve.

##### 3.2.67 lr repetition number 67

Figure 3.75: Calibration plot for logistic regression for simulation scenario 5. GAM is used as scatterplot smoother over repetition number 67 to estimate the calibration curve.

##### 3.2.68 lr repetition number 68

Figure 3.76: Calibration plot for logistic regression for simulation scenario 5. GAM is used as scatterplot smoother over repetition number 68 to estimate the calibration curve.

##### 3.2.69 lr repetition number 69

Figure 3.77: Calibration plot for logistic regression for simulation scenario 5. GAM is used as scatterplot smoother over repetition number 69 to estimate the calibration curve.

##### 3.2.70 lr repetition number 70

Figure 3.78: Calibration plot for logistic regression for simulation scenario 5. GAM is used as scatterplot smoother over repetition number 70 to estimate the calibration curve.

##### 3.2.71 lr repetition number 71

Figure 3.79: Calibration plot for logistic regression for simulation scenario 5. GAM is used as scatterplot smoother over repetition number 71 to estimate the calibration curve.

##### 3.2.72 lr repetition number 72

Figure 3.80: Calibration plot for logistic regression for simulation scenario 5. GAM is used as scatterplot smoother over repetition number 72 to estimate the calibration curve.

##### 3.2.73 lr repetition number 73

Figure 3.81: Calibration plot for logistic regression for simulation scenario 5. GAM is used as scatterplot smoother over repetition number 73 to estimate the calibration curve.

##### 3.2.74 lr repetition number 74

Figure 3.82: Calibration plot for logistic regression for simulation scenario 5. GAM is used as scatterplot smoother over repetition number 74 to estimate the calibration curve.

##### 3.2.75 lr repetition number 75

Figure 3.83: Calibration plot for logistic regression for simulation scenario 5. GAM is used as scatterplot smoother over repetition number 75 to estimate the calibration curve.

##### 3.2.76 lr repetition number 76

Figure 3.84: Calibration plot for logistic regression for simulation scenario 5. GAM is used as scatterplot smoother over repetition number 76 to estimate the calibration curve.

##### 3.2.77 lr repetition number 77

Figure 3.85: Calibration plot for logistic regression for simulation scenario 5. GAM is used as scatterplot smoother over repetition number 77 to estimate the calibration curve.

##### 3.2.78 lr repetition number 78

Figure 3.86: Calibration plot for logistic regression for simulation scenario 5. GAM is used as scatterplot smoother over repetition number 78 to estimate the calibration curve.

##### 3.2.79 lr repetition number 79

Figure 3.87: Calibration plot for logistic regression for simulation scenario 5. GAM is used as scatterplot smoother over repetition number 79 to estimate the calibration curve.

##### 3.2.80 lr repetition number 80

Figure 3.88: Calibration plot for logistic regression for simulation scenario 5. GAM is used as scatterplot smoother over repetition number 80 to estimate the calibration curve.

##### 3.2.81 lr repetition number 81

Figure 3.89: Calibration plot for logistic regression for simulation scenario 5. GAM is used as scatterplot smoother over repetition number 81 to estimate the calibration curve.

##### 3.2.82 lr repetition number 82

Figure 3.90: Calibration plot for logistic regression for simulation scenario 5. GAM is used as scatterplot smoother over repetition number 82 to estimate the calibration curve.

##### 3.2.83 lr repetition number 83

Figure 3.91: Calibration plot for logistic regression for simulation scenario 5. GAM is used as scatterplot smoother over repetition number 83 to estimate the calibration curve.

##### 3.2.84 lr repetition number 84

Figure 3.92: Calibration plot for logistic regression for simulation scenario 5. GAM is used as scatterplot smoother over repetition number 84 to estimate the calibration curve.

##### 3.2.85 lr repetition number 85

Figure 3.93: Calibration plot for logistic regression for simulation scenario 5. GAM is used as scatterplot smoother over repetition number 85 to estimate the calibration curve.

##### 3.2.86 lr repetition number 86

Figure 3.94: Calibration plot for logistic regression for simulation scenario 5. GAM is used as scatterplot smoother over repetition number 86 to estimate the calibration curve.

##### 3.2.87 lr repetition number 87

Figure 3.95: Calibration plot for logistic regression for simulation scenario 5. GAM is used as scatterplot smoother over repetition number 87 to estimate the calibration curve.

##### 3.2.88 lr repetition number 88

Figure 3.96: Calibration plot for logistic regression for simulation scenario 5. GAM is used as scatterplot smoother over repetition number 88 to estimate the calibration curve.

##### 3.2.89 lr repetition number 89

Figure 3.97: Calibration plot for logistic regression for simulation scenario 5. GAM is used as scatterplot smoother over repetition number 89 to estimate the calibration curve.

##### 3.2.90 lr repetition number 90

Figure 3.98: Calibration plot for logistic regression for simulation scenario 5. GAM is used as scatterplot smoother over repetition number 90 to estimate the calibration curve.

##### 3.2.91 lr repetition number 91

Figure 3.99: Calibration plot for logistic regression for simulation scenario 5. GAM is used as scatterplot smoother over repetition number 91 to estimate the calibration curve.

##### 3.2.92 lr repetition number 92

Figure 3.100: Calibration plot for logistic regression for simulation scenario 5. GAM is used as scatterplot smoother over repetition number 92 to estimate the calibration curve.

##### 3.2.93 lr repetition number 93

Figure 3.101: Calibration plot for logistic regression for simulation scenario 5. GAM is used as scatterplot smoother over repetition number 93 to estimate the calibration curve.

##### 3.2.94 lr repetition number 94

Figure 3.102: Calibration plot for logistic regression for simulation scenario 5. GAM is used as scatterplot smoother over repetition number 94 to estimate the calibration curve.

##### 3.2.95 lr repetition number 95

Figure 3.103: Calibration plot for logistic regression for simulation scenario 5. GAM is used as scatterplot smoother over repetition number 95 to estimate the calibration curve.

##### 3.2.96 lr repetition number 96

Figure 3.104: Calibration plot for logistic regression for simulation scenario 5. GAM is used as scatterplot smoother over repetition number 96 to estimate the calibration curve.

##### 3.2.97 lr repetition number 97

Figure 3.105: Calibration plot for logistic regression for simulation scenario 5. GAM is used as scatterplot smoother over repetition number 97 to estimate the calibration curve.

##### 3.2.98 lr repetition number 98

Figure 3.106: Calibration plot for logistic regression for simulation scenario 5. GAM is used as scatterplot smoother over repetition number 98 to estimate the calibration curve.

##### 3.2.99 lr repetition number 99

Figure 3.107: Calibration plot for logistic regression for simulation scenario 5. GAM is used as scatterplot smoother over repetition number 99 to estimate the calibration curve.

##### 3.2.100 lr repetition number 100

Figure 3.108: Calibration plot for logistic regression for simulation scenario 5. GAM is used as scatterplot smoother over repetition number 100 to estimate the calibration curve.

##### 3.2.101 All repetitions overlapped

Figure 3.109: Calibration plot for logistic regression for simulation scenario 5. GAM is used as scatterplot smoother over repetition number 1 to estimate the calibration curve.

### 4 MSE tables and boxplots

#### 4.1 Logistic regression

Table 4.1: Table 4.2: Summary statistics for mean squared error (MSE) for logistic regression. The statistics are computed over 100 simulation replications.

| Scenario | Approach | Mean | Standard deviation | Median | 25th percentile | 75th percentile |
| --- | --- | --- | --- | --- | --- | --- |
| 1 | NoCal | 0.0383662 | 0.0057773 | 0.0381932 | 0.0340784 | 0.0424767 |
| 1 | Logistic | 0.0005136 | 0.0004559 | 0.0003697 | 0.0001623 | 0.0007133 |
| 1 | SBA-10 | 0.0381869 | 0.0058044 | 0.0379801 | 0.0340136 | 0.0424366 |
| 1 | Elkan | 0.0021300 | 0.0012697 | 0.0019404 | 0.0011282 | 0.0028500 |
| 1 | SimCal | 0.0005003 | 0.0004224 | 0.0003470 | 0.0002070 | 0.0006702 |
| 1 | ACP | 0.0398109 | 0.0058478 | 0.0397576 | 0.0355137 | 0.0436159 |
| 5 | NoCal | 0.2577773 | 0.0126522 | 0.2580991 | 0.2490049 | 0.2666260 |
| 5 | Logistic | 0.0004951 | 0.0004167 | 0.0003912 | 0.0001622 | 0.0007538 |
| 5 | SBA-10 | 0.2593206 | 0.0128262 | 0.2596967 | 0.2504777 | 0.2680422 |
| 5 | Elkan | 0.0374255 | 0.0089162 | 0.0373878 | 0.0313712 | 0.0424534 |
| 5 | SimCal | 0.0037163 | 0.0017617 | 0.0034938 | 0.0026536 | 0.0046631 |
| 5 | ACP | 0.2567261 | 0.0129263 | 0.2563695 | 0.2473488 | 0.2654906 |
| 9 | NoCal | 0.0466000 | 0.0060640 | 0.0467779 | 0.0426455 | 0.0512289 |
| 9 | Logistic | 0.0013463 | 0.0006029 | 0.0013323 | 0.0008521 | 0.0017797 |
| 9 | SBA-10 | 0.0456300 | 0.0062043 | 0.0455693 | 0.0417761 | 0.0509850 |
| 9 | Elkan | 0.0020454 | 0.0010744 | 0.0017872 | 0.0012119 | 0.0026426 |
| 9 | SimCal | 0.0015009 | 0.0007010 | 0.0014638 | 0.0009862 | 0.0019398 |
| 9 | ACP | 0.0467880 | 0.0061565 | 0.0468449 | 0.0424765 | 0.0512222 |
| 14 | NoCal | 0.0395118 | 0.0060557 | 0.0398505 | 0.0354449 | 0.0430247 |
| 14 | Logistic | 0.0030596 | 0.0027200 | 0.0021844 | 0.0009728 | 0.0044058 |
| 14 | SBA-10 | 0.0393676 | 0.0060313 | 0.0394427 | 0.0351660 | 0.0426909 |
| 14 | Elkan | 0.0027133 | 0.0030495 | 0.0016529 | 0.0005685 | 0.0034854 |
| 14 | SimCal | 0.0018837 | 0.0020779 | 0.0011786 | 0.0005815 | 0.0019944 |
| 14 | ACP | 0.0411565 | 0.0062325 | 0.0409820 | 0.0377190 | 0.0445742 |
| 16 | NoCal | 0.0262782 | 0.0052054 | 0.0257759 | 0.0225805 | 0.0302991 |
| 16 | Logistic | 0.0002876 | 0.0002925 | 0.0002319 | 0.0001086 | 0.0003718 |
| 16 | SBA-10 | 0.0261820 | 0.0051983 | 0.0256384 | 0.0224883 | 0.0301980 |
| 16 | Elkan | 0.0007199 | 0.0007281 | 0.0004717 | 0.0002061 | 0.0010750 |
| 16 | SimCal | 0.0003262 | 0.0003669 | 0.0002064 | 0.0001010 | 0.0003650 |
| 16 | ACP | 0.0278409 | 0.0052721 | 0.0274080 | 0.0242613 | 0.0319874 |
| 19 | NoCal | 0.0350827 | 0.0087979 | 0.0349806 | 0.0292416 | 0.0404662 |
| 19 | Logistic | 0.0003755 | 0.0003608 | 0.0002414 | 0.0001140 | 0.0005108 |
| 19 | SBA-10 | 0.0329265 | 0.0076812 | 0.0326442 | 0.0279072 | 0.0378920 |
| 19 | Elkan | 0.1017338 | 0.0094232 | 0.1015841 | 0.0951341 | 0.1086317 |
| 19 | SimCal | 0.0039505 | 0.0018103 | 0.0037470 | 0.0025928 | 0.0047794 |
| 19 | ACP | 0.0554418 | 0.0436683 | 0.0420229 | 0.0349222 | 0.0512321 |
| 20 | NoCal | 0.0418256 | 0.0053015 | 0.0417963 | 0.0380847 | 0.0451988 |
| 20 | Logistic | 0.0002330 | 0.0002473 | 0.0001787 | 0.0000829 | 0.0002946 |
| 20 | SBA-10 | 0.0418826 | 0.0053116 | 0.0418633 | 0.0381476 | 0.0452932 |
| 20 | Elkan | 0.0288131 | 0.0041777 | 0.0283011 | 0.0259590 | 0.0315670 |
| 20 | SimCal | 0.0283850 | 0.0045210 | 0.0279490 | 0.0248408 | 0.0318800 |
| 20 | ACP | 0.0446413 | 0.0061899 | 0.0445673 | 0.0407800 | 0.0488142 |
| 21 | NoCal | 0.1895818 | 0.0124368 | 0.1902397 | 0.1810602 | 0.1978097 |
| 21 | Logistic | 0.0003301 | 0.0003306 | 0.0002172 | 0.0000954 | 0.0004279 |
| 21 | SBA-10 | 0.1890908 | 0.0124517 | 0.1899563 | 0.1804528 | 0.1975061 |
| 21 | Elkan | 0.1342200 | 0.0107678 | 0.1329064 | 0.1275667 | 0.1415987 |
| 21 | SimCal | 0.1300365 | 0.0103172 | 0.1300383 | 0.1238162 | 0.1362037 |
| 21 | ACP | 0.1890744 | 0.0127562 | 0.1897911 | 0.1796952 | 0.1969798 |

Figure 4.1: Boxplots of mean square error (MSE) for different calibration approaches for logistic regression.

### 5 LogLoss tables and boxplots

#### 5.1 Logistic regression

Table 4.1: Table 5.1: Summary statistics for LogLoss for logistic regression. The statistics are computed over 100 simulation replications.

| Scenario | Approach | Mean | Standard deviation | Median | 25th percentile | 75th percentile |
| --- | --- | --- | --- | --- | --- | --- |
| 1 | NoCal | 0.6120175 | 0.0150614 | 0.6107429 | 0.6008104 | 0.6211513 |
| 1 | Logistic | 0.5220640 | 0.0053327 | 0.5227021 | 0.5180542 | 0.5262075 |
| 1 | SBA-10 | 0.6112700 | 0.0149539 | 0.6099948 | 0.6004855 | 0.6207271 |
| 1 | Elkan | 0.5259206 | 0.0059605 | 0.5261197 | 0.5213872 | 0.5298153 |
| 1 | SimCal | 0.5220155 | 0.0055574 | 0.5231100 | 0.5176688 | 0.5266537 |
| 1 | ACP | 0.6504199 | 0.0384231 | 0.6416671 | 0.6246356 | 0.6611537 |
| 5 | NoCal | 1.3502440 | 0.0625996 | 1.3473328 | 1.3175474 | 1.3885843 |
| 5 | Logistic | 0.5220151 | 0.0052688 | 0.5226275 | 0.5181602 | 0.5258820 |
| 5 | SBA-10 | 1.3445215 | 0.0618312 | 1.3424561 | 1.3136744 | 1.3797079 |
| 5 | Elkan | 0.6196962 | 0.0281969 | 0.6180239 | 0.6036888 | 0.6348614 |
| 5 | SimCal | 0.5349176 | 0.0091362 | 0.5344188 | 0.5293037 | 0.5403545 |
| 5 | ACP | 2.1250475 | 0.4708346 | 2.0412339 | 1.8184039 | 2.4709328 |
| 9 | NoCal | 0.6558455 | 0.0140861 | 0.6555162 | 0.6471798 | 0.6655992 |
| 9 | Logistic | 0.5545738 | 0.0042278 | 0.5545449 | 0.5515969 | 0.5578473 |
| 9 | SBA-10 | 0.6532054 | 0.0134710 | 0.6532561 | 0.6443230 | 0.6619652 |
| 9 | Elkan | 0.5560614 | 0.0046641 | 0.5558614 | 0.5524392 | 0.5593441 |
| 9 | SimCal | 0.5551798 | 0.0043581 | 0.5549033 | 0.5520482 | 0.5582427 |
| 9 | ACP | 0.6738598 | 0.0254178 | 0.6685190 | 0.6566772 | 0.6880060 |
| 14 | NoCal | 0.6152870 | 0.0147467 | 0.6151437 | 0.6040263 | 0.6245029 |
| 14 | Logistic | 0.5308279 | 0.0092656 | 0.5284800 | 0.5243225 | 0.5352850 |
| 14 | SBA-10 | 0.6146865 | 0.0144959 | 0.6135118 | 0.6034267 | 0.6237397 |
| 14 | Elkan | 0.5280826 | 0.0091340 | 0.5266136 | 0.5212158 | 0.5326534 |
| 14 | SimCal | 0.5264666 | 0.0070641 | 0.5256065 | 0.5212959 | 0.5312327 |
| 14 | ACP | 0.6523721 | 0.0424072 | 0.6434079 | 0.6247007 | 0.6679255 |
| 16 | NoCal | 0.4471733 | 0.0136121 | 0.4453957 | 0.4377501 | 0.4569834 |
| 16 | Logistic | 0.3792023 | 0.0050803 | 0.3790692 | 0.3758405 | 0.3826998 |
| 16 | SBA-10 | 0.4468725 | 0.0135809 | 0.4449851 | 0.4374713 | 0.4565585 |
| 16 | Elkan | 0.3803410 | 0.0053012 | 0.3806582 | 0.3762090 | 0.3843202 |
| 16 | SimCal | 0.3793453 | 0.0050923 | 0.3797014 | 0.3757185 | 0.3827899 |
| 16 | ACP | 0.4738620 | 0.0298812 | 0.4698666 | 0.4509069 | 0.4859398 |
| 19 | NoCal | 0.6321682 | 0.0272301 | 0.6309336 | 0.6134526 | 0.6466563 |
| 19 | Logistic | 0.5416824 | 0.0046570 | 0.5424262 | 0.5381299 | 0.5450958 |
| 19 | SBA-10 | 0.6203109 | 0.0199174 | 0.6180564 | 0.6053720 | 0.6324977 |
| 19 | Elkan | 0.8322501 | 0.0408590 | 0.8321193 | 0.8003582 | 0.8637017 |
| 19 | SimCal | 0.5525030 | 0.0068753 | 0.5519360 | 0.5485592 | 0.5552869 |
| 19 | ACP | 2.4591451 | 1.5417878 | 1.9829521 | 1.5365819 | 2.7053554 |
| 20 | NoCal | 0.4783745 | 0.0143549 | 0.4788993 | 0.4685292 | 0.4874997 |
| 20 | Logistic | 0.3500640 | 0.0070915 | 0.3513778 | 0.3460147 | 0.3552445 |
| 20 | SBA-10 | 0.4787087 | 0.0143735 | 0.4792615 | 0.4687735 | 0.4879661 |
| 20 | Elkan | 0.4474465 | 0.0126519 | 0.4459371 | 0.4379628 | 0.4568073 |
| 20 | SimCal | 0.4446125 | 0.0134971 | 0.4431233 | 0.4357365 | 0.4542900 |
| 20 | ACP | 0.4851393 | 0.0157150 | 0.4845155 | 0.4747878 | 0.4951418 |
| 21 | NoCal | 0.9848744 | 0.0368309 | 0.9854691 | 0.9594041 | 1.0094810 |
| 21 | Logistic | 0.5330884 | 0.0045083 | 0.5327116 | 0.5304301 | 0.5366886 |
| 21 | SBA-10 | 0.9811160 | 0.0363510 | 0.9820322 | 0.9564914 | 1.0064623 |
| 21 | Elkan | 0.8902178 | 0.0331122 | 0.8882667 | 0.8680126 | 0.9125321 |
| 21 | SimCal | 0.8990075 | 0.0360056 | 0.8995792 | 0.8697802 | 0.9232900 |
| 21 | ACP | 1.2247602 | 0.1709790 | 1.1972000 | 1.0804077 | 1.3561196 |

Figure 5.1: Boxplots of LogLoss for different calibration approaches for logistic regression.

### 6 Critical difference plots for the LogLoss per machine

#### 6.1 Logistic regression

##### 6.1.1 Critical difference plots logistic regression scenario 1

Figure 6.1: Critical difference plots for the LogLoss for logistic regression for simulation scenario 1. Lower average ranks are considered better. Groups of calibration approaches connected by a horizontal line segment can not be shown to have a significantly different performance. The p-value from the Iman & Davenport modification of Friedman test is <0.001.

##### 6.1.2 Critical difference plots logistic regression scenario 5

Figure 6.2: Critical difference plots for the LogLoss for logistic regression for simulation scenario 5. Lower average ranks are considered better. Groups of calibration approaches connected by a horizontal line segment can not be shown to have a significantly different performance. The p-value from the Iman & Davenport modification of Friedman test is <0.001.

##### 6.1.3 Critical difference plots logistic regression scenario 9

Figure 6.3: Critical difference plots for the LogLoss for logistic regression for simulation scenario 9. Lower average ranks are considered better. Groups of calibration approaches connected by a horizontal line segment can not be shown to have a significantly different performance. The p-value from the Iman & Davenport modification of Friedman test is <0.001.

##### 6.1.4 Critical difference plots logistic regression scenario 14

Figure 6.4: Critical difference plots for the LogLoss for logistic regression for simulation scenario 14. Lower average ranks are considered better. Groups of calibration approaches connected by a horizontal line segment can not be shown to have a significantly different performance. The p-value from the Iman & Davenport modification of Friedman test is <0.001.

##### 6.1.5 Critical difference plots logistic regression scenario 16

Figure 6.5: Critical difference plots for the LogLoss for logistic regression for simulation scenario 16. Lower average ranks are considered better. Groups of calibration approaches connected by a horizontal line segment can not be shown to have a significantly different performance. The p-value from the Iman & Davenport modification of Friedman test is <0.001.

##### 6.1.6 Critical difference plots logistic regression scenario 19

Figure 6.6: Critical difference plots for the LogLoss for logistic regression for simulation scenario 19. Lower average ranks are considered better. Groups of calibration approaches connected by a horizontal line segment can not be shown to have a significantly different performance. The p-value from the Iman & Davenport modification of Friedman test is <0.001.

##### 6.1.7 Critical difference plots logistic regression scenario 20

Figure 6.7: Critical difference plots for the LogLoss for logistic regression for simulation scenario 20. Lower average ranks are considered better. Groups of calibration approaches connected by a horizontal line segment can not be shown to have a significantly different performance. The p-value from the Iman & Davenport modification of Friedman test is <0.001.

##### 6.1.8 Critical difference plots logistic regression scenario 21

Figure 6.8: Critical difference plots for the LogLoss for logistic regression for simulation scenario 21. Lower average ranks are considered better. Groups of calibration approaches connected by a horizontal line segment can not be shown to have a significantly different performance. The p-value from the Iman & Davenport modification of Friedman test is <0.001.

### 7 Critical difference plots for the LogLoss averaging over machines

#### 7.1 Critical difference plot scenario 1

Figure 7.1: Critical difference plots for the LogLoss for simulation scenario 1. Results are averaged over all probability machines. Lower average ranks are considered better. Groups of calibration approaches connected by a horizontal line segment can not be shown to have a significantly different performance. The p-value from the Iman & Davenport modification of Friedman test is <0.001.

#### 7.2 Critical difference plot scenario 5

Figure 7.2: Critical difference plots for the LogLoss for simulation scenario 5. Results are averaged over all probability machines. Lower average ranks are considered better. Groups of calibration approaches connected by a horizontal line segment can not be shown to have a significantly different performance. The p-value from the Iman & Davenport modification of Friedman test is <0.001.

#### 7.3 Critical difference plot scenario 9

Figure 7.3: Critical difference plots for the LogLoss for simulation scenario 9. Results are averaged over all probability machines. Lower average ranks are considered better. Groups of calibration approaches connected by a horizontal line segment can not be shown to have a significantly different performance. The p-value from the Iman & Davenport modification of Friedman test is <0.001.

#### 7.4 Critical difference plot scenario 14

Figure 7.4: Critical difference plots for the LogLoss for simulation scenario 14. Results are averaged over all probability machines. Lower average ranks are considered better. Groups of calibration approaches connected by a horizontal line segment can not be shown to have a significantly different performance. The p-value from the Iman & Davenport modification of Friedman test is <0.001.

#### 7.5 Critical difference plot scenario 16

Figure 7.5: Critical difference plots for the LogLoss for simulation scenario 16. Results are averaged over all probability machines. Lower average ranks are considered better. Groups of calibration approaches connected by a horizontal line segment can not be shown to have a significantly different performance. The p-value from the Iman & Davenport modification of Friedman test is <0.001.

#### 7.6 Critical difference plot scenario 19

Figure 7.6: Critical difference plots for the LogLoss for simulation scenario 19. Results are averaged over all probability machines. Lower average ranks are considered better. Groups of calibration approaches connected by a horizontal line segment can not be shown to have a significantly different performance. The p-value from the Iman & Davenport modification of Friedman test is <0.001.

#### 7.7 Critical difference plot scenario 20

Figure 7.7: Critical difference plots for the LogLoss for simulation scenario 20. Results are averaged over all probability machines. Lower average ranks are considered better. Groups of calibration approaches connected by a horizontal line segment can not be shown to have a significantly different performance. The p-value from the Iman & Davenport modification of Friedman test is <0.001.

#### 7.8 Critical difference plot scenario 21

Figure 7.8: Critical difference plots for the LogLoss for simulation scenario 21. Results are averaged over all probability machines. Lower average ranks are considered better. Groups of calibration approaches connected by a horizontal line segment can not be shown to have a significantly different performance. The p-value from the Iman & Davenport modification of Friedman test is <0.001.

### 8 R session information

It took 13.1 mins for the R code used to produce the
tables and figures in this file to run.

```
R version 4.5.2 (2025-10-31 ucrt)
Platform: x86_64-w64-mingw32/x64
Running under: Windows 11 x64 (build 26100)

Matrix products: default
  LAPACK version 3.12.1

locale:
[1] LC_COLLATE=English_United States.utf8 
[2] LC_CTYPE=English_United States.utf8   
[3] LC_MONETARY=English_United States.utf8
[4] LC_NUMERIC=C                          
[5] LC_TIME=English_United States.utf8    

time zone: Europe/Zurich
tzcode source: internal

attached base packages:
[1] stats     graphics  grDevices utils     datasets  methods   base     

other attached packages:
[1] kableExtra_1.4.0 forcats_1.0.0    Hmisc_5.2-3      stringr_1.5.1   
[5] gridExtra_2.3    ggplot2_4.0.2    dplyr_1.1.4      tidyr_1.3.1     
[9] here_1.0.2      

loaded via a namespace (and not attached):
 [1] gtable_0.3.6       xfun_0.52          bslib_0.9.0        htmlwidgets_1.6.4 
 [5] lattice_0.22-7     vctrs_0.7.1        tools_4.5.2        generics_0.1.4    
 [9] tibble_3.3.0       cluster_2.1.8.1    pkgconfig_2.0.3    Matrix_1.7-4      
[13] data.table_1.17.8  checkmate_2.3.2    RColorBrewer_1.1-3 S7_0.2.1          
[17] lifecycle_1.0.5    compiler_4.5.2     farver_2.1.2       textshaping_1.0.1 
[21] htmltools_0.5.8.1  sass_0.4.10        yaml_2.3.10        htmlTable_2.4.3   
[25] Formula_1.2-5      pillar_1.11.1      jquerylib_0.1.4    cachem_1.1.0      
[29] rpart_4.1.24       nlme_3.1-168       tidyselect_1.2.1   digest_0.6.37     
[33] stringi_1.8.7      purrr_1.1.0        bookdown_0.44      labeling_0.4.3    
[37] splines_4.5.2      rprojroot_2.1.1    fastmap_1.2.0      grid_4.5.2        
[41] colorspace_2.1-1   cli_3.6.5          magrittr_2.0.3     base64enc_0.1-3   
[45] foreign_0.8-90     withr_3.0.2        scales_1.4.0       backports_1.5.0   
[49] rmarkdown_2.29     nnet_7.3-20        evaluate_1.0.4     knitr_1.50        
[53] viridisLite_0.4.2  mgcv_1.9-3         rlang_1.1.7        glue_1.8.0        
[57] xml2_1.3.8         svglite_2.2.1      rstudioapi_0.17.1  jsonlite_2.0.0    
[61] R6_2.6.1           systemfonts_1.3.1
```
