## Supplementary material for "Calibrating machine learning approaches for probability estimation without calibration data": Supplementary_material_4a.Rmd

````
---
date: "`r Sys.Date()`"
output:  
  bookdown::html_document2:
    toc: true
    toc_float: true
    theme: united
    fig_caption: yes
editor_options: 
  markdown: 
    wrap: 72
---

```{r setup, include=FALSE}
knitr::opts_chunk$set(echo = FALSE)
```

```{r}
# Supplementary Material 4a for 
# "Calibrating machine learning approaches for probability
# estimation in case of the absence of calibration data"

# This script is a modified version of the one found in supplementary material 
# 2 of Ojeda FM, et al., Calibrating machine learning approaches for probability 
# estimation: A comprehensive comparison. Stat Med. 2023 Dec 20;42(29):5451-5478

# Knitting this Rmd file produces Supplementary Material 4b, which 
# contains graphics and tables showing the results of the recalibrating the 
# estimated probabilities in the simulated datasets with misspecifications in 
# mean parameter.
```

```{r}
CAL_VERSION <- "cal" 
```

---
title: "Simulation study 2: performance and comparison of new calibrating machine learning 
approach (Simcal) for probability estimation in case of misspecification of the mean."
subtitle: "Supplementary material 4b"
author: "Eleonora Di Carluccio, Francisco M. Ojeda"
---

# About this file

This html file contains tables and figures presenting results of the
simulation study conducted to compare the performance of different
calibration approaches.

Data from two populations with different disease prevalences were
generated. The data from the first population were used for model
building. Data from the second population were split into training
(cal-training) data and test (cal-test) data.

In all simulation scenarios the sample size of the model building data
and cal-test data was set to 1000 observations.

In `r ifelse(CAL_VERSION == "cal", "(this file)", "")` results are
presented where the number of observations in the cal-training data was
set to 1000 for all scenarios, except simulation scenarios 14 where only
100 observations were used.

This version aims to investigate the effect of misspecification of the mean
parameters on the novel Simcal approach.

```{r, message=FALSE, warning=FALSE}
begin_time <- Sys.time()
# Attaching packages -----------------------------------------------------------
library(here)
library(tidyr)
library(dplyr)
library(ggplot2)
library(stringr)
library(Hmisc)
library(forcats)
library(kableExtra)

# Loading functions -------------------------------------------------------------
source(here( "Calibration_Sup2/04-rmd-functions.R"))
source(here("Calibration_Sup2/03-calibration-functions.R"))
```

```{r}
# To avoid dplyr::sumarise message:
# '`summarise()` regrouping output by ... (override with `.groups` argument)'
# See
# https://stackoverflow.com/questions/62140483/how-to-interpret-dplyr-message-summarise-regrouping-output-by-x-override
options(dplyr.summarise.inform = FALSE) 
```

```{r}
# The objects defined on this code chunk can be used to reduce the time
# this Rmd file takes to run (e.g. by changing the number of scenarios 
# considered or disabling some plots).

# Simulation scenarios examined 
SCENARIOS <- paste0("16.", 1:10)

# Reduce number of replications that are used to produced the graphics
# and tables. For each simulation scenario 100 replications were used.
REDUCE_REPN <- FALSE
if(REDUCE_REPN) N_REPN <- 10 else N_REPN <- 100

# Parameter to be passed to method in 'geom_smooth' for calibration plots. 
# 'loess' takes considerably more time to run than 'gam'.
SMOOTH <- "gam"

# Parameter to be passed to se in 'geom_smooth' for calibration plots
SE_GEOM_SMOOTH <- FALSE

# Should scatterplots of recalibrate probabilities be produced?
DRAW_SCATTERPLOTS <- TRUE

# Should boxplots of MSE and LogLoss be produced?
DRAW_BOXPLOTS <- TRUE

# Should CD plots according to single machines be produced? 
DRAW_CD1 <- TRUE

# Should CD plots averaging over machines be produced?
DRAW_CD2 <- TRUE

# For the purpose of this simulation, only the lr machine is investigated
machine_abbreviations <- "lr"
```

```{r, results='asis'}
if(REDUCE_REPN) {
  cat("<b>The number of replications examined was changed from 100 to ", N_REPN, 
      "</b>", sep = "")
  cat("\n\n")
}
```

# Simulation scenarios

On this section a table summarizing all simulation scenarios is
presented.

```{r}
tab <- rbind(
  #c(1, 2, 0, 0, "-"),
  #c(2, 2, 0, 10, "10 noise variables"),
  #c(3, 2, 0, 50, "50 noise variables"),
  #c(4, 2, 0, 100, "100 noise variables"),
  #c(5, 2, 0, 0, "Lower disease prevalence"),
  #c(6, 2, 0, 10, "Lower disease prevalence"),
  #c(7, 2, 0, 50, "Lower disease prevalence"),
  #c(8, 2, 0, 100, "Lower disease prevalence"),
  #c(9, 5, 2, 0, "Additional covariates"),
  #c(10, 5, 5, 10, "Additional covariates plus noise variables"),
  #c(11, 5, 5, 50, "Additional covariates plus noise variables"),
  #c(12, 5, 5, 100, "Additional covariates plus noise variables"),
  #c(13, 2, 0, 0, "Data generation using Mease model"),
  #c(14, 2, 0, 0, "Smaller calibration data set for training"),
  #c(15, 2, 0, 0, paste0("Smaller calibration data set for ",                
  #               "training, data generation using Mease",
  #               "model")),
  c(16.1, 1, 0, 0, "Covariate distribution: MB N(1, 1), misspecified 
    Cal N(1.1, 1)"),
  c(16.2, 1, 0, 0, "Covariate distribution: MB N(1, 1), misspecified 
    Cal N(1.2, 1)"),
  c(16.3, 1, 0, 0, "Covariate distribution: MB N(1, 1), misspecified 
    Cal N(1.3, 1)"),
  c(16.4, 1, 0, 0, "Covariate distribution: MB N(1, 1), misspecified 
    Cal N(1.4, 1)"),
  c(16.5, 1, 0, 0, "Covariate distribution: MB N(1, 1), misspecified 
    Cal N(1.5, 1)"),
  c(16.6, 1, 0, 0, "Covariate distribution: MB N(1, 1), misspecified 
    Cal N(1.6, 1)"),
  c(16.7, 1, 0, 0, "Covariate distribution: MB N(1, 1), misspecified 
    Cal N(1.7, 1)"),
  c(16.8, 1, 0, 0, "Covariate distribution: MB N(1, 1), misspecified 
    Cal N(1.8, 1)"),
  c(16.9, 1, 0, 0, "Covariate distribution: MB N(1, 1), misspecified 
    Cal N(1.9, 1)"),
  c("16.10", 1, 0, 0, "Covariate distribution: MB N(1, 1), misspecified 
    Cal N(2, 1)"))
  #c(17, 1, 0, 0, "Unequal covariate distribution: MB N(1, 1), Cal N(1, 2)"),
  #c(18, 1, 0, 0, "Unequal covariate distribution: MB N(1, 1), Cal N(-1, 1)"),
  #c(19, 1, 0, 0, "Unequal covariate distribution: MB N(1, 1), Cal N(-1, 2)"),
  #c(20, 1, 0, 0, "Unequal coefficients: MB beta = 1, Cal beta = 3"),
  #c(21, 1, 0, 0, "Unequal coefficients: MB beta = 1, Cal beta = -1"))
  #c(22, 2, 0, 0, paste0("Unequal covariate distribution for both continuous",
  #               " variables: MB N(1, 1), Cal N(-1, 1)")),
  #c(23, 2, 0, 0,  paste0("Unequal covariate distribution for one ",    
  #                        "continuous variable: MB N(1, 1), Cal N(3, 1), ",
  #                        "for the other continuous variable: ",
  #                       "MB N(1, 1), Cal N(-1, 1)")))
  
colnames(tab) <- c("Scenario", "Continuous", "Categorical", "Noise", 
                   "Distinctive feature")

#tab <- tab[1:length(SCENARIOS), ]
kable_wrapper(tab)
# Adding information on prevalences and EPV to table summarizing simulation
# scenarios

new_cols <- NULL

cols_to_keep <- c("prev_build_train", "prev_cal_train", "epv_build_train", 
                  "epv_cal_train")

if(CAL_VERSION == "cal_alt") {
  cols_to_keep <- sub("cal_train", "cal_alt_train", cols_to_keep)
}

for(scenario in SCENARIOS) {
  for (mispec_type in c("mean-mispec")) {
    File <- here(paste0("rds-files/after-recalibration-misspec/", 
               "data-with-cal-probs--scen-", scenario, mispec_type, ".Rds"))
    results <- readRDS(file = File)
    matrix_prev_epv <- results[["matrix_prev_epv"]][, cols_to_keep] %>% 
      colMeans()
    
    n_build_train <- nrow(results$data_list[[1]][["build"]][["train"]])
    n_cal_train <- nrow(results$data_list[[1]][[CAL_VERSION]]$train)
    n_cal_test <- nrow(results$data_list[[1]][[CAL_VERSION]]$test)
    
    col_names <- names(matrix_prev_epv)
    dim(matrix_prev_epv) <- c(1, 4)
    colnames(matrix_prev_epv) <- col_names
    
    matrix_prev_epv <- cbind(n_build_train = n_build_train, 
                             n_cal_train = n_cal_train, 
                             n_cal_test = n_cal_test, matrix_prev_epv)
    
    new_cols <- rbind(new_cols, matrix_prev_epv)
    rm(File, results, matrix_prev_epv)
  }
}
rownames(new_cols) <- NULL
for(i in 4:5) new_cols[, i] <- round(new_cols[, i], 2) 
for(i in 6:7) new_cols[, i] <- round(new_cols[, i], 1)
colnames(new_cols) <- c("MB N", "Cal-training N", "Cal-test N",
                        "MB prevalence", "Cal-training prevalence",
                        "MB EPV", "Cal-training EPV")

tab <- cbind(tab, new_cols)
reordered_cols <- c(setdiff(colnames(tab), "Distinctive feature"))#,
                    #"Distinctive feature")
tab <- tab[, reordered_cols]

caption <- paste0("Main settings in the simulation scenarios: Number of ",
                  "continuous covariates (Continuous), number of categorical ",
                  "covariates (Categorical), number of noise variables (Noise), ",
                  "model building data set sample size (MB N), ",
                  "calibration training data set sample size ",
                  "(Cal-training N), ",
                  "calibration test data set sample size (Cal-test N), ",
                  "model building data set outcome prevalence ",
                  "(MB prevalence), ",
                  "calibration training data set outcome ",
                  "prevalence (Cal-training prevalence), ",
                  "model building data set number of events per ",
                  "variable (MB EVP), ",
                  "calibration training data set number of events per ",
                  "variable (Cal-training EVP) and, if applicable, additional",
                  " distinctive feature (Distinctive feature).")

kable_wrapper(tab, caption) %>% 
  add_footnote("MB: Model building data; EPV: events per variable.", 
               notation = "none")
rm(tab, caption)
```

# Calibration plots

```{r}
# Creating captions for calibration plots 
fig_captions <- NULL

for(machine in machine_abbreviations) {
  machine_name <- machine_full_names[machine]
  
  for(scenario in SCENARIOS) {
    
    if(scenario %in% setdiff(SCENARIOS, 16:21) | machine != "en"){
      caption <- paste0("Calibration plot for ", tolower(machine_name), 
                        " for simulation scenario ", scenario, ". ", 
                        toupper(SMOOTH), " is used as ",
                        "scatterplot smoother over all ", N_REPN, 
                        " replications to estimate the calibration curve.")
      fig_captions <- c(fig_captions, caption)
      rm(caption)
    }
  } 
  rm(machine_name)
}
rm(machine)

```

```{r, echo=FALSE}
# custom functions to exclude unwanted calibration methods
beautify_recal_labels <- function(x, machine) {
 # Levels <- c("NoCal","Refit", "Elkan", "Logistic", "Beta", "Simcal")
  Levels <- c("NoCal", "Simcal",
              "Misspecified Simcal")
  x <- gsub("_", " ", x) %>% 
    sub(paste0("^", machine, " "), "", .) %>%
    sub(" cal$", "", .) %>% 
    sub(paste0("^", machine), "NoCal", .) %>%
    #sub("rcs", "RCS", .) %>%
    sub("simcal mispec", "Misspecified Simcal", .) %>%
    capitalize() %>% factor(levels = Levels)
  if(sum(is.na(x)) > 0) stop("NAs introduced")
  x
}

prepare_data_average_logloss_demsar <- function(scenario, reduce_repn, N_REPN,
                                                cal_version = c("cal", 
                                                                "cal_alt")) {
  cols <- NULL
  
  if(scenario %in% 16:21) {
    machine_abbreviations <- setdiff(machine_abbreviations, "en")
  }
  
  # for(machine in machine_abbreviations){
  #   cols <- c(cols, sub("prob_lr", paste0("prob_", machine), cols_lr))
  # }
  # cols <- c("prob_lr", "prob_lr_refit", "prob_lr_elkan_cal", "prob_lr_logistic_cal", 
  #   "prob_lr_beta_cal", "prob_lr_simcal_cal")
  cols <- c("prob_lr", "prob_lr_simcal_cal", 
            "prob_lr_simcal_mispec_cal")
  df <- prepare_plot_df(scenario = scenario, cols = cols,
                        build_or_cal = cal_version,
                        reduce_repn = reduce_repn, N_REPN = N_REPN)
  
  df[["Method"]] <- df[["machine_and_recal_method"]]
  
  df[["Machine"]] <- (strsplit(df[["machine_and_recal_method"]], "_") %>% 
                        sapply(`[`, 1))
  
  y <- "Method"
  for(machine in machine_abbreviations){
    df[[y]] <- sub(paste0(machine, "$"), "NoCal", df[[y]])
  }
  df[[y]] <- sub("_cal$", "", df[[y]])
  v <- which(str_detect(df[[y]], "_"))
  df[[y]][v] <- (strsplit(df[[y]][v], "_") %>% sapply(`[`, 2) %>% 
                   sub("rcs", "RCS", .)  %>% capitalize()) 
  #Levels <-  c("NoCal","Refit", "Elkan", "Logistic", "Beta", "Simcal")
  Levels <- c("Simcal", "Misspecified Simcal")

  df[[y]] <- factor(df[[y]], levels = Levels)
  
  df <- group_by(df, Machine, Method, repn) %>% 
    summarise(LogLoss = log_loss(prob_true, Probability)) %>%
    group_by(Method, repn) %>% summarise(`AverageLogLoss` = mean(LogLoss))
  
  df_wide <- pivot_wider(df, names_from = Method, 
                         values_from = AverageLogLoss, id_cols = repn)
  
  df_wide[["repn"]] <- NULL
  
  as.matrix(df_wide)
  
}
```

```{r, results='asis', eval = DRAW_SCATTERPLOTS, message=FALSE, warning=FALSE, fig.cap=fig_captions}
# Scatterplots of probabilities (true and recalibrated)

epsilon <- 0.01

for(machine in machine_abbreviations) {
  machine_name <- machine_full_names[machine]
  #cols <- sub("prob_lr", paste0("prob_", machine), cols_lr)
  #cols <- c("prob_lr", "prob_lr_refit", "prob_lr_elkan_cal", "prob_lr_logistic_cal", 
  #  "prob_lr_beta_cal", "prob_lr_simcal_cal")
  cols <- c("prob_lr",
            "prob_lr_simcal_cal",
            "prob_lr_simcal_mispec_cal")
  
  cat("\n\n")
  cat("##", machine_name); cat("\n\n"); cat("<br>");cat("\n\n")
  
  
  for(scenario in SCENARIOS) {
    for (mispec_param in c("mean")) {
          if(scenario %in% setdiff(SCENARIOS, 16:21) | machine != "en"){
      
      cat(paste0("### ", machine_name, " scenario ", scenario, 
                 " misspecified ", 
                 if_else(mispec_param == "both", "mean and variance",
                         mispec_param)))
      cat("\n\n"); cat("<br>"); cat("\n\n")
      df_plot <- prepare_plot_df_mispec(scenario, cols = cols,
                                 build_or_cal = CAL_VERSION,
                                 reduce_repn = REDUCE_REPN, N_REPN = N_REPN,
                                 mispec_param = mispec_param)
      aux <- beautify_recal_labels(df_plot[["machine_and_recal_method"]], 
                                   machine)
      df_plot[["recal_method"]] <- aux; rm(aux)
      
      p <- ggplot(df_plot, aes(x = Probability, y = prob_true)) +
        geom_point(alpha = 0.05, colour = "gray") + 
        geom_abline(slope = 1, 
                    intercept = 0, 
                    lty = 1, 
                    colour = "black") +
        geom_smooth(method = SMOOTH, se = SE_GEOM_SMOOTH, span = 0.75,
                    lty = 2, colour = "black") +
        facet_wrap(vars(recal_method), nrow = 1) +  coord_fixed(ratio = 1) +
        scale_x_continuous(name = "Predicted probability", 
                           limits = c(0 - epsilon, 1 + epsilon)) + 
        scale_y_continuous(name = "True probability", 
                           limits = c(0 - epsilon, 1 + epsilon)) + 
        theme_bw() +  theme(strip.background = element_rect(fill = "gray95"),
                            panel.spacing.x = unit(0.4, "lines"),
                            axis.text = element_text(size = 7))
      print(p)
      
      cat("\n\n"); cat("<br>");cat("\n\n")
      rm(p, df_plot)
    }
    }
    
  }
  
  rm(machine_name, cols)
}

rm(epsilon)
```

```{r}
rm(fig_captions)
```

```{r}
# Creating captions for boxplots of MSE and LogLoss per recalibration method
fig_captions <- NULL

performance_measures <- c("mean square error (MSE)", "LogLoss")
for(perf_meas_index in 1:2) {

  for(machine in machine_abbreviations) {
    machine_name <- machine_full_names[machine]
    caption <- paste0("Boxplots of ", performance_measures[perf_meas_index],
                      " for different calibration approaches for ", 
                      tolower(machine_name), ".")
    fig_captions <- c(fig_captions, caption)
    rm(caption)
    
  }
}  
caption <- c("Boxplots of LogLoss for different mean misspecifications for SimCal calibration for logistic regression.")
fig_captions <- c(fig_captions, caption)
rm(perf_meas_index, performance_measures)
```

```{r, results='asis', fig.width=9, fig.height=9, eval=DRAW_BOXPLOTS, fig.cap=fig_captions}
# Boxplots of MSE and LogLoss per recalibration method

# Tables labels are not updated inside a chunk, see:
# https://github.com/rstudio/bookdown/issues/1097
# The solutions suggested there is for labels to be created manually.


performance_measures <- c("MSE", "LogLoss")
performance_measure_functions <- c("mse_brier", "log_loss")
for(perf_meas_index in 1:2) {
  
  section_index <- perf_meas_index + 3
  table_number <- 0
  
  perf_meas <- performance_measures[perf_meas_index]
  perf_fun <- get(performance_measure_functions[perf_meas_index])
  
  cat("#", perf_meas, "tables and boxplots\n\n")
  
  perf_meas_long <- perf_meas
  if(perf_meas == "MSE") perf_meas_long <- "mean squared error (MSE)"

  for(machine in machine_abbreviations) {
    
    table_number <- table_number + 1
    
    #cols <- sub("prob_lr", paste0("prob_", machine), cols_lr)
   # cols <- c("prob_lr", "prob_lr_refit", "prob_lr_elkan_cal", "prob_lr_logistic_cal",  "prob_lr_beta_cal", "prob_lr_simcal_cal")
    cols <- c("prob_lr",
              "prob_lr_simcal_cal", 
              "prob_lr_simcal_mispec_cal")
    machine_name <- machine_full_names[machine]
    
    cat("##", machine_name, "\n\n")
    
    FUN <- function(x, mispec_param) {
      out <- prepare_plot_df_mispec(scenario = x, cols = cols,
                             build_or_cal = CAL_VERSION,
                             reduce_repn = REDUCE_REPN, N_REPN = N_REPN,
                                 mispec_param = mispec_param)
      y <- "machine_and_recal_method"
      out[["recal_method"]] <- beautify_recal_labels(out[[y]], machine) 
      out <- group_by(out, recal_method, repn, scenario) %>% 
        summarise(perf_ms = perf_fun(prob_true, Probability))
    }
    
    scenarios_available <- SCENARIOS
    
    if(machine == "en") scenarios_available <- setdiff(SCENARIOS, 16:21)
    
    for (mispec_param in c("mean")) {
      cat(paste0("### ", 
                 "Misspecified ", 
                 if_else(mispec_param == "both", "mean and variance",
                         mispec_param)))
      cat("\n\n"); cat("<br>"); cat("\n\n")
      
      df_plot <- lapply(scenarios_available, FUN, mispec_param = mispec_param) %>% do.call("rbind", .)

     df_plot[["scenario_new"]] <- as.factor(rep(paste("Scenario", paste0("16.", 1:length(SCENARIOS))), each = 300))
    
    desired_order <- paste("Scenario", paste0("16.", 1:length(unique(df_plot[["scenario_new"]]))))
    df_plot$scenario_new <- factor(df_plot$scenario_new, levels = desired_order)
    
    
    tab <- summarise_perf_measure(df_plot, perf_measure = perf_ms)
    caption <- paste0("Summary statistics for ", perf_meas_long, " for ", 
                      tolower(machine_name), ". The statistics are ",
                      "computed over ", 
                      N_REPN, " simulation replications.")
    tab_label <- paste0(perf_meas, section_index, "-", table_number)
    kable_wrapper(tab, caption, label = tab_label) %>% cat()
    cat("<br>"); cat("\n\n")     
    
    
    df_plot[["recal_method"]] <- fct_rev(df_plot[["recal_method"]])
    p <- ggplot(df_plot, aes(x = recal_method, y = perf_ms)) +
      geom_boxplot(outlier.size = 0.5) +
      facet_wrap(vars(scenario_new)) + 
      xlab("") + ylab(perf_meas) + coord_flip() +
      theme_bw() + ggtitle(machine_name) + 
      theme(strip.background = element_rect(fill = "gray95"),
            panel.spacing.x = unit(0.75, "lines"))
    print(p)
    
    if (perf_meas == "LogLoss") {

      df_plot_simcal <- df_plot %>% 
      filter(recal_method == "Simcal" & scenario_new == "Scenario 16.1") 
      df_plot_simcal[["scenario_new"]] <- "original"
      
          df_plot_new <- df_plot %>% 
      filter(recal_method == "Misspecified Simcal")
    df_plot_new <- bind_rows(df_plot_new, df_plot_simcal)
      
    df_plot_new <- df_plot_new %>% 
      mutate(misspec_index = case_when(
        scenario_new == "original" ~ 0.00,
        scenario_new == "Scenario 16.1" ~ 0.1,
        scenario_new == "Scenario 16.2" ~ 0.2,
        scenario_new == "Scenario 16.3" ~ 0.3,
        scenario_new == "Scenario 16.4" ~ 0.4,
        scenario_new == "Scenario 16.5" ~ 0.5,
        scenario_new == "Scenario 16.6" ~ 0.6,
        scenario_new == "Scenario 16.7" ~ 0.7,
        scenario_new == "Scenario 16.8" ~ 0.8,
        scenario_new == "Scenario 16.9" ~ 0.9,
        scenario_new == "Scenario 16.10" ~ 1.0))
    
    df_plot_new[["misspec_index"]] <- factor(df_plot_new[["misspec_index"]])

    q <- ggplot(df_plot_new, aes(x = misspec_index, y = perf_ms)) +
      geom_boxplot(outlier.size = 0.5) +
      #facet_wrap(vars(scenario_new)) + 
      xlab("Misspecification of mean") + ylab(perf_meas) + #coord_flip() +
      theme_bw() + 
      theme(strip.background = element_rect(fill = "gray95"),
            panel.spacing.x = unit(0.75, "lines"),
            text = element_text(size = 12)) + 
      theme(axis.title.y = element_text(margin = margin(r = 15)))
   print(q)

    }
    
    rm(p, df_plot, tab, caption, tab_label)
    cat("<br>\n\n")
    }
    
    rm(scenarios_available)
  }
  
  rm(perf_meas, perf_fun, perf_meas_long, section_index, table_number)
}

rm(perf_meas_index)
```

```{r}
rm(fig_captions)
```

# Critical difference plots for the LogLoss per machine

```{r}
nemenyi_per_machine <- function(x, cols, machine, mispec_param) {
  out <- prepare_plot_df_mispec(scenario = x, cols = cols,
                         build_or_cal = CAL_VERSION,
                         reduce_repn = REDUCE_REPN, N_REPN = N_REPN,
                                 mispec_param = mispec_param)
  aux <- beautify_recal_labels(out[["machine_and_recal_method"]],
                               machine)
  out[["recal_method"]] <- aux; rm(aux)
  out <- group_by(out, recal_method, repn, scenario) %>%
    summarise(LogLoss = log_loss(prob_true, Probability))
}
```

```{r}
# Creating captions for machine and scenario specific CD plots

fig_captions <- NULL

for(machine in machine_abbreviations) {
  the_scenarios <- SCENARIOS
  #cols <- sub("prob_lr", paste0("prob_", machine), cols_lr)
  #cols <- c("prob_lr", "prob_lr_refit", "prob_lr_elkan_cal", "prob_lr_logistic_cal", "prob_lr_beta_cal", "prob_lr_simcal_cal")
  cols <- c("prob_lr",
            "prob_lr_simcal_cal", 
            "prob_lr_simcal_mispec_cal")
  machine_name <- machine_full_names[machine]
  if(machine == "en") the_scenarios <- setdiff(SCENARIOS, 16:21)

  for (mispec_param in c("mean")) {

  df_plot <- lapply(the_scenarios, nemenyi_per_machine, cols = cols,
                    machine = machine, mispec_param = mispec_param) %>% do.call("rbind", .)
  df_plot[["scenario"]] <- paste("Scenario", df_plot[["scenario"]])

  for(scenario in the_scenarios) {

    dsub <- df_plot[which(df_plot[["scenario"]] == paste("Scenario",
                                                         scenario)), ]
    dsub[["scenario"]] <- NULL
    dsub_wide <- pivot_wider(dsub, names_from = recal_method,
                             values_from = LogLoss, id_cols = repn)
    dsub_wide[["repn"]] <- NULL
    dsub_wide <- as.matrix(dsub_wide)

    # small_better = FALSE combined with x_axis_1_right = FALSE on
    # 'CD_diagram_Nemenyi' will produce CD plots where the lowest
    # ranks are the better ranks.
    id_test <- iman_davenport_test(dsub_wide, small_better = FALSE)
    pval_iman_dvt <- id_test[["logloss"]][["F.test"]][["pval_iman_davenport"]]


    caption <- paste0("Critical difference plots for the LogLoss for ",
                      tolower(machine_name),
                      " for simulation scenario ", scenario, ". Lower ",
                      "average ranks are ",
                      "considered better. Groups of calibration approaches ",
                      "connected by a horizontal line segment can not be ",
                      "shown to have a significantly different performance. ",
                      "The p-value from the Iman & Davenport modification of ",
                      "Friedman test is ", format_p.ade(pval_iman_dvt, 3),
                      ".")
    fig_captions <- c(fig_captions, caption)
    rm(caption, id_test, pval_iman_dvt, dsub_wide)
  }
  }
  rm(the_scenarios, machine_name, df_plot)
}
rm(machine)
```

```{r, results='asis', eval=DRAW_CD1, fig.cap=fig_captions, warning=FALSE, message=FALSE}

section_index <- 6

subsection_index <- 0

for(machine in machine_abbreviations) {

  subsection_index <- subsection_index + 1

  #cols <- sub("prob_lr", paste0("prob_", machine), cols_lr)
  #cols <- c("prob_lr", "prob_lr_refit", "prob_lr_elkan_cal", "prob_lr_logistic_cal",  "prob_lr_beta_cal", "prob_lr_simcal_cal")
  cols <- c("prob_lr",
            "prob_lr_simcal_cal",
            "prob_lr_simcal_mispec_cal")
  machine_name <- machine_full_names[machine]

  cat("\n\n")
  cat("##", machine_name); cat("\n\n"); cat("<br>"); cat("\n\n")

  the_scenarios <- SCENARIOS
  if(machine == "en") the_scenarios <- setdiff(SCENARIOS, 16:21)


  for (mispec_param in c("mean")) {
      cat(paste0("### ",
                 "Misspecified ",
                 if_else(mispec_param == "both", "mean and variance",
                         mispec_param)))
      cat("\n\n"); cat("<br>"); cat("\n\n")

  df_plot <- lapply(the_scenarios, nemenyi_per_machine, cols = cols,
                    machine = machine, mispec_param = mispec_param) %>% do.call("rbind", .)
  df_plot[["scenario"]] <- paste("Scenario", df_plot[["scenario"]])

  for(scenario in the_scenarios) {
    dsub <- df_plot[which(df_plot[["scenario"]] == paste("Scenario",
                                                         scenario)), ]
    dsub[["scenario"]] <- NULL
    dsub_wide <- pivot_wider(dsub, names_from = recal_method,
                             values_from = LogLoss, id_cols = repn)
    dsub_wide[["repn"]] <- NULL
    dsub_wide <- as.matrix(dsub_wide)


    id_test <- iman_davenport_test(dsub_wide, small_better = FALSE)

    cat(paste0("#### Critical difference plots ", tolower(machine_name),
               " scenario ", scenario)); cat("\n\n")

    p <- CD_diagram_Nemenyi(id_test, coord_fixed_ratio = 0.8,
                            x_axis_1_right = FALSE)
    print(p)

    cat("\n\n"); cat("<br>"); cat("\n\n")

    rm(dsub_wide, dsub, p)
  }
  }
  cat("\n\n"); cat("<br>"); cat("\n\n")
  rm(df_plot, the_scenarios, machine_name)

}

rm(machine, section_index, subsection_index)
```

```{r}
rm(fig_captions)
```


# R session information

```{r}
# Computing time it takes for the code in this Rmd file to run.
# Observe that this does not include running the simulation nor the calibration.

end_time <- Sys.time()
TIME <- end_time - begin_time
x <- as.numeric(unclass(TIME))
UNIT <- attr(TIME, "units")
```

It took `r round(x, 2)` `r UNIT` for the R code used to produce the
tables and figures in this file to run.

```{r, comment="", results='markup'}
sessionInfo()
```
````
