## Supplementary material for "Calibrating machine learning approaches for probability estimation without calibration data": Supplementary_material_4b.html

Simulation study 2: performance and comparison of new calibrating machine learning approach (Simcal) for probability estimation in case of misspecification of the mean.


### Simulation study 2: performance and comparison of new calibrating machine learning approach (Simcal) for probability estimation in case of misspecification of the mean.

##### Supplementary material 4b

###### Eleonora Di Carluccio, Francisco M. Ojeda

#### 2026-06-25

### 1 About this file

This html file contains tables and figures presenting results of the
simulation study conducted to compare the performance of different
calibration approaches.

Data from two populations with different disease prevalences were
generated. The data from the first population were used for model
building. Data from the second population were split into training
(cal-training) data and test (cal-test) data.

In all simulation scenarios the sample size of the model building data
and cal-test data was set to 1000 observations.

In (this file) results are
presented where the number of observations in the cal-training data was
set to 1000 for all scenarios, except simulation scenarios 14 where only
100 observations were used.

This version aims to investigate the effect of misspecification of the mean
parameters on the novel Simcal approach.

### 2 Simulation scenarios

On this section a table summarizing all simulation scenarios is
presented.

| Scenario | Continuous | Categorical | Noise | Distinctive feature |
| --- | --- | --- | --- | --- |
| 16.1 | 1 | 0 | 0 | Covariate distribution: MB N(1, 1), misspecified Cal N(1.1, 1) |
| 16.2 | 1 | 0 | 0 | Covariate distribution: MB N(1, 1), misspecified Cal N(1.2, 1) |
| 16.3 | 1 | 0 | 0 | Covariate distribution: MB N(1, 1), misspecified Cal N(1.3, 1) |
| 16.4 | 1 | 0 | 0 | Covariate distribution: MB N(1, 1), misspecified Cal N(1.4, 1) |
| 16.5 | 1 | 0 | 0 | Covariate distribution: MB N(1, 1), misspecified Cal N(1.5, 1) |
| 16.6 | 1 | 0 | 0 | Covariate distribution: MB N(1, 1), misspecified Cal N(1.6, 1) |
| 16.7 | 1 | 0 | 0 | Covariate distribution: MB N(1, 1), misspecified Cal N(1.7, 1) |
| 16.8 | 1 | 0 | 0 | Covariate distribution: MB N(1, 1), misspecified Cal N(1.8, 1) |
| 16.9 | 1 | 0 | 0 | Covariate distribution: MB N(1, 1), misspecified Cal N(1.9, 1) |
| 16.10 | 1 | 0 | 0 | Covariate distribution: MB N(1, 1), misspecified Cal N(2, 1) |

Table 2.1: Table 2.2: Main settings in the simulation scenarios: Number of continuous covariates (Continuous), number of categorical covariates (Categorical), number of noise variables (Noise), model building data set sample size (MB N), calibration training data set sample size (Cal-training N), calibration test data set sample size (Cal-test N), model building data set outcome prevalence (MB prevalence), calibration training data set outcome prevalence (Cal-training prevalence), model building data set number of events per variable (MB EVP), calibration training data set number of events per variable (Cal-training EVP) and, if applicable, additional distinctive feature (Distinctive feature).

| Scenario | Continuous | Categorical | Noise | MB N | Cal-training N | Cal-test N | MB prevalence | Cal-training prevalence | MB EPV | Cal-training EPV |
| --- | --- | --- | --- | --- | --- | --- | --- | --- | --- | --- |
| 16.1 | 1 | 0 | 0 | 1000 | 1000 | 1000 | 0.7 | 0.84 | 303.1 | 156.4 |
| 16.2 | 1 | 0 | 0 | 1000 | 1000 | 1000 | 0.7 | 0.84 | 303.1 | 156.4 |
| 16.3 | 1 | 0 | 0 | 1000 | 1000 | 1000 | 0.7 | 0.84 | 303.1 | 156.4 |
| 16.4 | 1 | 0 | 0 | 1000 | 1000 | 1000 | 0.7 | 0.84 | 303.1 | 156.4 |
| 16.5 | 1 | 0 | 0 | 1000 | 1000 | 1000 | 0.7 | 0.84 | 303.1 | 156.4 |
| 16.6 | 1 | 0 | 0 | 1000 | 1000 | 1000 | 0.7 | 0.84 | 303.1 | 156.4 |
| 16.7 | 1 | 0 | 0 | 1000 | 1000 | 1000 | 0.7 | 0.84 | 303.1 | 156.4 |
| 16.8 | 1 | 0 | 0 | 1000 | 1000 | 1000 | 0.7 | 0.84 | 303.1 | 156.4 |
| 16.9 | 1 | 0 | 0 | 1000 | 1000 | 1000 | 0.7 | 0.84 | 303.1 | 156.4 |
| 16.10 | 1 | 0 | 0 | 1000 | 1000 | 1000 | 0.7 | 0.84 | 303.1 | 156.4 |
|  |  |  |  |  |  |  |  |  |  |  |
| --- | --- | --- | --- | --- | --- | --- | --- | --- | --- | --- |
| MB: Model building data; EPV: events per variable. |  |  |  |  |  |  |  |  |  |  |

### 3 Calibration plots

#### 3.1 Logistic regression

##### 3.1.1 Logistic regression scenario 16.1 misspecified mean

Figure 3.1: Calibration plot for logistic regression for simulation scenario 16.1. GAM is used as scatterplot smoother over all 100 replications to estimate the calibration curve.

##### 3.1.2 Logistic regression scenario 16.2 misspecified mean

Figure 3.2: Calibration plot for logistic regression for simulation scenario 16.2. GAM is used as scatterplot smoother over all 100 replications to estimate the calibration curve.

##### 3.1.3 Logistic regression scenario 16.3 misspecified mean

Figure 3.3: Calibration plot for logistic regression for simulation scenario 16.3. GAM is used as scatterplot smoother over all 100 replications to estimate the calibration curve.

##### 3.1.4 Logistic regression scenario 16.4 misspecified mean

Figure 3.4: Calibration plot for logistic regression for simulation scenario 16.4. GAM is used as scatterplot smoother over all 100 replications to estimate the calibration curve.

##### 3.1.5 Logistic regression scenario 16.5 misspecified mean

Figure 3.5: Calibration plot for logistic regression for simulation scenario 16.5. GAM is used as scatterplot smoother over all 100 replications to estimate the calibration curve.

##### 3.1.6 Logistic regression scenario 16.6 misspecified mean

Figure 3.6: Calibration plot for logistic regression for simulation scenario 16.6. GAM is used as scatterplot smoother over all 100 replications to estimate the calibration curve.

##### 3.1.7 Logistic regression scenario 16.7 misspecified mean

Figure 3.7: Calibration plot for logistic regression for simulation scenario 16.7. GAM is used as scatterplot smoother over all 100 replications to estimate the calibration curve.

##### 3.1.8 Logistic regression scenario 16.8 misspecified mean

Figure 3.8: Calibration plot for logistic regression for simulation scenario 16.8. GAM is used as scatterplot smoother over all 100 replications to estimate the calibration curve.

##### 3.1.9 Logistic regression scenario 16.9 misspecified mean

Figure 3.9: Calibration plot for logistic regression for simulation scenario 16.9. GAM is used as scatterplot smoother over all 100 replications to estimate the calibration curve.

##### 3.1.10 Logistic regression scenario 16.10 misspecified mean

Figure 3.10: Calibration plot for logistic regression for simulation scenario 16.10. GAM is used as scatterplot smoother over all 100 replications to estimate the calibration curve.

### 4 MSE tables and boxplots

#### 4.1 Logistic regression

##### 4.1.1 Misspecified mean

Table 4.1: Table 4.2: Summary statistics for mean squared error (MSE) for logistic regression. The statistics are computed over 100 simulation replications.

| Scenario | Approach | Mean | Standard deviation | Median | 25th percentile | 75th percentile |
| --- | --- | --- | --- | --- | --- | --- |
| 16.1 | NoCal | 0.0262782 | 0.0052054 | 0.0257759 | 0.0225805 | 0.0302991 |
| 16.1 | Simcal | 0.0003262 | 0.0003669 | 0.0002064 | 0.0001010 | 0.0003650 |
| 16.1 | Misspecified Simcal | 0.0007039 | 0.0006749 | 0.0005564 | 0.0002206 | 0.0009134 |
| 16.10 | NoCal | 0.0262782 | 0.0052054 | 0.0257759 | 0.0225805 | 0.0302991 |
| 16.10 | Simcal | 0.0003262 | 0.0003669 | 0.0002064 | 0.0001010 | 0.0003650 |
| 16.10 | Misspecified Simcal | 0.0316195 | 0.0087235 | 0.0312695 | 0.0260470 | 0.0364434 |
| 16.2 | NoCal | 0.0262782 | 0.0052054 | 0.0257759 | 0.0225805 | 0.0302991 |
| 16.2 | Simcal | 0.0003262 | 0.0003669 | 0.0002064 | 0.0001010 | 0.0003650 |
| 16.2 | Misspecified Simcal | 0.0015325 | 0.0011214 | 0.0013917 | 0.0006932 | 0.0019199 |
| 16.3 | NoCal | 0.0262782 | 0.0052054 | 0.0257759 | 0.0225805 | 0.0302991 |
| 16.3 | Simcal | 0.0003262 | 0.0003669 | 0.0002064 | 0.0001010 | 0.0003650 |
| 16.3 | Misspecified Simcal | 0.0028825 | 0.0016774 | 0.0027772 | 0.0016827 | 0.0035123 |
| 16.4 | NoCal | 0.0262782 | 0.0052054 | 0.0257759 | 0.0225805 | 0.0302991 |
| 16.4 | Simcal | 0.0003262 | 0.0003669 | 0.0002064 | 0.0001010 | 0.0003650 |
| 16.4 | Misspecified Simcal | 0.0048120 | 0.0023369 | 0.0047206 | 0.0032139 | 0.0057670 |
| 16.5 | NoCal | 0.0262782 | 0.0052054 | 0.0257759 | 0.0225805 | 0.0302991 |
| 16.5 | Simcal | 0.0003262 | 0.0003669 | 0.0002064 | 0.0001010 | 0.0003650 |
| 16.5 | Misspecified Simcal | 0.0073783 | 0.0031039 | 0.0072761 | 0.0051965 | 0.0087240 |
| 16.6 | NoCal | 0.0262782 | 0.0052054 | 0.0257759 | 0.0225805 | 0.0302991 |
| 16.6 | Simcal | 0.0003262 | 0.0003669 | 0.0002064 | 0.0001010 | 0.0003650 |
| 16.6 | Misspecified Simcal | 0.0106376 | 0.0039840 | 0.0104969 | 0.0079434 | 0.0125754 |
| 16.7 | NoCal | 0.0262782 | 0.0052054 | 0.0257759 | 0.0225805 | 0.0302991 |
| 16.7 | Simcal | 0.0003262 | 0.0003669 | 0.0002064 | 0.0001010 | 0.0003650 |
| 16.7 | Misspecified Simcal | 0.0146435 | 0.0049825 | 0.0144342 | 0.0112927 | 0.0172491 |
| 16.8 | NoCal | 0.0262782 | 0.0052054 | 0.0257759 | 0.0225805 | 0.0302991 |
| 16.8 | Simcal | 0.0003262 | 0.0003669 | 0.0002064 | 0.0001010 | 0.0003650 |
| 16.8 | Misspecified Simcal | 0.0194463 | 0.0061039 | 0.0191602 | 0.0153622 | 0.0225960 |
| 16.9 | NoCal | 0.0262782 | 0.0052054 | 0.0257759 | 0.0225805 | 0.0302991 |
| 16.9 | Simcal | 0.0003262 | 0.0003669 | 0.0002064 | 0.0001010 | 0.0003650 |
| 16.9 | Misspecified Simcal | 0.0250916 | 0.0073507 | 0.0248242 | 0.0202457 | 0.0290063 |

Figure 4.1: Boxplots of mean square error (MSE) for different calibration approaches for logistic regression.

### 5 LogLoss tables and boxplots

#### 5.1 Logistic regression

##### 5.1.1 Misspecified mean

Table 4.1: Table 5.1: Summary statistics for LogLoss for logistic regression. The statistics are computed over 100 simulation replications.

| Scenario | Approach | Mean | Standard deviation | Median | 25th percentile | 75th percentile |
| --- | --- | --- | --- | --- | --- | --- |
| 16.1 | NoCal | 0.4471733 | 0.0136121 | 0.4453957 | 0.4377501 | 0.4569834 |
| 16.1 | Simcal | 0.3793453 | 0.0050923 | 0.3797014 | 0.3757185 | 0.3827899 |
| 16.1 | Misspecified Simcal | 0.3801739 | 0.0052861 | 0.3801411 | 0.3758641 | 0.3840627 |
| 16.10 | NoCal | 0.4471733 | 0.0136121 | 0.4453957 | 0.4377501 | 0.4569834 |
| 16.10 | Simcal | 0.3793453 | 0.0050923 | 0.3797014 | 0.3757185 | 0.3827899 |
| 16.10 | Misspecified Simcal | 0.4589695 | 0.0214598 | 0.4566367 | 0.4467464 | 0.4724615 |
| 16.2 | NoCal | 0.4471733 | 0.0136121 | 0.4453957 | 0.4377501 | 0.4569834 |
| 16.2 | Simcal | 0.3793453 | 0.0050923 | 0.3797014 | 0.3757185 | 0.3827899 |
| 16.2 | Misspecified Simcal | 0.3823407 | 0.0058225 | 0.3828730 | 0.3779934 | 0.3860681 |
| 16.3 | NoCal | 0.4471733 | 0.0136121 | 0.4453957 | 0.4377501 | 0.4569834 |
| 16.3 | Simcal | 0.3793453 | 0.0050923 | 0.3797014 | 0.3757185 | 0.3827899 |
| 16.3 | Misspecified Simcal | 0.3859774 | 0.0067016 | 0.3858923 | 0.3813675 | 0.3897477 |
| 16.4 | NoCal | 0.4471733 | 0.0136121 | 0.4453957 | 0.4377501 | 0.4569834 |
| 16.4 | Simcal | 0.3793453 | 0.0050923 | 0.3797014 | 0.3757185 | 0.3827899 |
| 16.4 | Misspecified Simcal | 0.3911616 | 0.0078982 | 0.3909663 | 0.3859678 | 0.3956811 |
| 16.5 | NoCal | 0.4471733 | 0.0136121 | 0.4453957 | 0.4377501 | 0.4569834 |
| 16.5 | Simcal | 0.3793453 | 0.0050923 | 0.3797014 | 0.3757185 | 0.3827899 |
| 16.5 | Misspecified Simcal | 0.3979703 | 0.0093950 | 0.3975858 | 0.3921277 | 0.4035862 |
| 16.6 | NoCal | 0.4471733 | 0.0136121 | 0.4453957 | 0.4377501 | 0.4569834 |
| 16.6 | Simcal | 0.3793453 | 0.0050923 | 0.3797014 | 0.3757185 | 0.3827899 |
| 16.6 | Misspecified Simcal | 0.4064796 | 0.0111870 | 0.4060437 | 0.3996936 | 0.4134638 |
| 16.7 | NoCal | 0.4471733 | 0.0136121 | 0.4453957 | 0.4377501 | 0.4569834 |
| 16.7 | Simcal | 0.3793453 | 0.0050923 | 0.3797014 | 0.3757185 | 0.3827899 |
| 16.7 | Misspecified Simcal | 0.4167637 | 0.0132784 | 0.4155844 | 0.4091153 | 0.4253014 |
| 16.8 | NoCal | 0.4471733 | 0.0136121 | 0.4453957 | 0.4377501 | 0.4569834 |
| 16.8 | Simcal | 0.3793453 | 0.0050923 | 0.3797014 | 0.3757185 | 0.3827899 |
| 16.8 | Misspecified Simcal | 0.4288946 | 0.0156792 | 0.4273724 | 0.4199927 | 0.4389839 |
| 16.9 | NoCal | 0.4471733 | 0.0136121 | 0.4453957 | 0.4377501 | 0.4569834 |
| 16.9 | Simcal | 0.3793453 | 0.0050923 | 0.3797014 | 0.3757185 | 0.3827899 |
| 16.9 | Misspecified Simcal | 0.4429414 | 0.0184020 | 0.4408648 | 0.4323329 | 0.4545643 |

Figure 5.1: Boxplots of LogLoss for different calibration approaches for logistic regression.

Figure 5.2: Boxplots of LogLoss for different mean misspecifications for SimCal calibration for logistic regression.

### 6 Critical difference plots for the LogLoss per machine

#### 6.1 Logistic regression

##### 6.1.1 Misspecified mean

###### 6.1.1.1 Critical difference plots logistic regression scenario 16.1

Figure 6.1: Critical difference plots for the LogLoss for logistic regression for simulation scenario 16.1. Lower average ranks are considered better. Groups of calibration approaches connected by a horizontal line segment can not be shown to have a significantly different performance. The p-value from the Iman & Davenport modification of Friedman test is <0.001.

###### 6.1.1.9 Critical difference plots logistic regression scenario 16.9

Figure 6.9: Critical difference plots for the LogLoss for logistic regression for simulation scenario 16.9. Lower average ranks are considered better. Groups of calibration approaches connected by a horizontal line segment can not be shown to have a significantly different performance. The p-value from the Iman & Davenport modification of Friedman test is <0.001.

###### 6.1.1.10 Critical difference plots logistic regression scenario 16.10

Figure 6.10: Critical difference plots for the LogLoss for logistic regression for simulation scenario 16.10. Lower average ranks are considered better. Groups of calibration approaches connected by a horizontal line segment can not be shown to have a significantly different performance. The p-value from the Iman & Davenport modification of Friedman test is <0.001.

### 7 R session information

It took 6.26 mins for the R code used to produce the
tables and figures in this file to run.

```
R version 4.5.2 (2025-10-31 ucrt)
Platform: x86_64-w64-mingw32/x64
Running under: Windows 11 x64 (build 26100)

Matrix products: default
  LAPACK version 3.12.1

locale:
[1] LC_COLLATE=English_United States.utf8 
[2] LC_CTYPE=English_United States.utf8   
[3] LC_MONETARY=English_United States.utf8
[4] LC_NUMERIC=C                          
[5] LC_TIME=English_United States.utf8    

time zone: Europe/Zurich
tzcode source: internal

attached base packages:
[1] stats     graphics  grDevices utils     datasets  methods   base     

other attached packages:
[1] kableExtra_1.4.0 forcats_1.0.0    Hmisc_5.2-3      stringr_1.5.1   
[5] ggplot2_4.0.2    dplyr_1.1.4      tidyr_1.3.1      here_1.0.2      

loaded via a namespace (and not attached):
 [1] gtable_0.3.6       xfun_0.52          bslib_0.9.0        htmlwidgets_1.6.4 
 [5] lattice_0.22-7     vctrs_0.7.1        tools_4.5.2        generics_0.1.4    
 [9] tibble_3.3.0       cluster_2.1.8.1    pkgconfig_2.0.3    Matrix_1.7-4      
[13] data.table_1.17.8  checkmate_2.3.2    RColorBrewer_1.1-3 S7_0.2.1          
[17] lifecycle_1.0.5    compiler_4.5.2     farver_2.1.2       textshaping_1.0.1 
[21] htmltools_0.5.8.1  sass_0.4.10        yaml_2.3.10        htmlTable_2.4.3   
[25] Formula_1.2-5      pillar_1.11.1      jquerylib_0.1.4    cachem_1.1.0      
[29] rpart_4.1.24       nlme_3.1-168       tidyselect_1.2.1   digest_0.6.37     
[33] stringi_1.8.7      purrr_1.1.0        bookdown_0.44      labeling_0.4.3    
[37] splines_4.5.2      rprojroot_2.1.1    fastmap_1.2.0      grid_4.5.2        
[41] colorspace_2.1-1   cli_3.6.5          magrittr_2.0.3     base64enc_0.1-3   
[45] foreign_0.8-90     withr_3.0.2        scales_1.4.0       backports_1.5.0   
[49] rmarkdown_2.29     nnet_7.3-20        gridExtra_2.3      evaluate_1.0.4    
[53] knitr_1.50         viridisLite_0.4.2  mgcv_1.9-3         rlang_1.1.7       
[57] glue_1.8.0         xml2_1.3.8         svglite_2.2.1      rstudioapi_0.17.1 
[61] jsonlite_2.0.0     R6_2.6.1           systemfonts_1.3.1
```
