## Supplementary material for "Calibrating machine learning approaches for probability estimation without calibration data": Supplementary_material_5b.html

Simulation study 3: performance and comparison of new calibrating machine learning approach for probability estimation in case of misspecification of the outcome probability.


### Simulation study 3: performance and comparison of new calibrating machine learning approach for probability estimation in case of misspecification of the outcome probability.

##### Supplementary material 5b

###### Eleonora Di Carluccio, Francisco M. Ojeda

#### 2026-06-25

### 1 About this file

This html file contains tables and figures presenting results of the
simulation study conducted to compare the performance of different
calibration approaches.

Data from two populations with different disease prevalences were
generated. The data from the first population were used for model
building. Data from the second population were split into training
(cal-training) data and test (cal-test) data.

In all simulation scenarios the sample size of the model building data
and cal-test data was set to 1000 observations.

In (this file) results are
presented where the number of observations in the cal-training data was
set to 1000 for all scenarios, except simulation scenarios 14 where only
100 observations were used.

This version aims to investigate the effect of misspecification of the mean,
variance and of both parameters on the novel Simcal approach.

### 2 Simulation scenarios

On this section a table summarizing all simulation scenarios is
presented.

| Scenario | Continuous | Categorical | Noise | Distinctive feature |
| --- | --- | --- | --- | --- |
| 16.1 | 1 | 0 | 0 | Covariate distribution: MB N(1, 1),  Cal N(1, 1), misspecified prevalence 0.76 |
| 16.2 | 1 | 0 | 0 | Covariate distribution: MB N(1, 1),  Cal N(1, 1), misspecified prevalence 0.78 |
| 16.3 | 1 | 0 | 0 | Covariate distribution: MB N(1, 1),  Cal N(1, 1), misspecified prevalence 0.80 |
| 16.4 | 1 | 0 | 0 | Covariate distribution: MB N(1, 1),  Cal N(1, 1), misspecified prevalence 0.82 |
| 16.5 | 1 | 0 | 0 | Covariate distribution: MB N(1, 1),  Cal N(1, 1), misspecified prevalence 0.86 |
| 16.6 | 1 | 0 | 0 | Covariate distribution: MB N(1, 1),  Cal N(1, 1), misspecified prevalence 0.88 |
| 16.7 | 1 | 0 | 0 | Covariate distribution: MB N(1, 1),  Cal N(1, 1), misspecified prevalence 0.90 |
| 16.8 | 1 | 0 | 0 | Covariate distribution: MB N(1, 1),  Cal N(1, 1), misspecified prevalence 0.92 |

### 4 MSE tables and boxplots

#### 4.1 Logistic regression

##### 4.1.1 Misspecified prop

Table 4.1: Table 4.2: Summary statistics for mean squared error (MSE) for logistic regression. The statistics are computed over 100 simulation replications.

| Scenario | Approach | Mean | Standard deviation | Median | 25th percentile | 75th percentile |
| --- | --- | --- | --- | --- | --- | --- |
| 16.1 | NoCal | 0.0262782 | 0.0052054 | 0.0257759 | 0.0225805 | 0.0302991 |
| 16.1 | Simcal | 0.0003262 | 0.0003669 | 0.0002064 | 0.0001010 | 0.0003650 |
| 16.1 | Misspecified Simcal | 0.0077398 | 0.0015108 | 0.0075124 | 0.0066772 | 0.0085441 |
| 16.2 | NoCal | 0.0262782 | 0.0052054 | 0.0257759 | 0.0225805 | 0.0302991 |
| 16.2 | Simcal | 0.0003262 | 0.0003669 | 0.0002064 | 0.0001010 | 0.0003650 |
| 16.2 | Misspecified Simcal | 0.0049609 | 0.0012417 | 0.0047402 | 0.0041419 | 0.0055777 |
| 16.3 | NoCal | 0.0262782 | 0.0052054 | 0.0257759 | 0.0225805 | 0.0302991 |
| 16.3 | Simcal | 0.0003262 | 0.0003669 | 0.0002064 | 0.0001010 | 0.0003650 |
| 16.3 | Misspecified Simcal | 0.0027357 | 0.0009525 | 0.0025117 | 0.0020839 | 0.0031804 |
| 16.4 | NoCal | 0.0262782 | 0.0052054 | 0.0257759 | 0.0225805 | 0.0302991 |
| 16.4 | Simcal | 0.0003262 | 0.0003669 | 0.0002064 | 0.0001010 | 0.0003650 |
| 16.4 | Misspecified Simcal | 0.0009657 | 0.0006195 | 0.0007898 | 0.0005881 | 0.0012241 |
| 16.5 | NoCal | 0.0262782 | 0.0052054 | 0.0257759 | 0.0225805 | 0.0302991 |
| 16.5 | Simcal | 0.0003262 | 0.0003669 | 0.0002064 | 0.0001010 | 0.0003650 |
| 16.5 | Misspecified Simcal | 0.0007312 | 0.0002787 | 0.0006668 | 0.0005349 | 0.0009032 |
| 16.6 | NoCal | 0.0262782 | 0.0052054 | 0.0257759 | 0.0225805 | 0.0302991 |
| 16.6 | Simcal | 0.0003262 | 0.0003669 | 0.0002064 | 0.0001010 | 0.0003650 |
| 16.6 | Misspecified Simcal | 0.0025751 | 0.0005009 | 0.0025278 | 0.0022173 | 0.0029335 |
| 16.7 | NoCal | 0.0262782 | 0.0052054 | 0.0257759 | 0.0225805 | 0.0302991 |
| 16.7 | Simcal | 0.0003262 | 0.0003669 | 0.0002064 | 0.0001010 | 0.0003650 |
| 16.7 | Misspecified Simcal | 0.0051329 | 0.0007209 | 0.0050933 | 0.0046327 | 0.0056665 |
| 16.8 | NoCal | 0.0262782 | 0.0052054 | 0.0257759 | 0.0225805 | 0.0302991 |
| 16.8 | Simcal | 0.0003262 | 0.0003669 | 0.0002064 | 0.0001010 | 0.0003650 |
| 16.8 | Misspecified Simcal | 0.0105909 | 0.0009829 | 0.0106089 | 0.0099807 | 0.0112972 |

Figure 4.1: Boxplots of mean square error (MSE) for different calibration approaches for logistic regression.

### 5 LogLoss tables and boxplots

#### 5.1 Logistic regression

##### 5.1.1 Misspecified prop

Table 4.1: Table 5.1: Summary statistics for LogLoss for logistic regression. The statistics are computed over 100 simulation replications.

| Scenario | Approach | Mean | Standard deviation | Median | 25th percentile | 75th percentile |
| --- | --- | --- | --- | --- | --- | --- |
| 16.1 | NoCal | 0.4471733 | 0.0136121 | 0.4453957 | 0.4377501 | 0.4569834 |
| 16.1 | Simcal | 0.3793453 | 0.0050923 | 0.3797014 | 0.3757185 | 0.3827899 |
| 16.1 | Misspecified Simcal | 0.3995802 | 0.0061126 | 0.3992699 | 0.3954856 | 0.4042289 |
| 16.2 | NoCal | 0.4471733 | 0.0136121 | 0.4453957 | 0.4377501 | 0.4569834 |
| 16.2 | Simcal | 0.3793453 | 0.0050923 | 0.3797014 | 0.3757185 | 0.3827899 |
| 16.2 | Misspecified Simcal | 0.3920413 | 0.0057816 | 0.3921694 | 0.3877790 | 0.3960909 |
| 16.3 | NoCal | 0.4471733 | 0.0136121 | 0.4453957 | 0.4377501 | 0.4569834 |
| 16.3 | Simcal | 0.3793453 | 0.0050923 | 0.3797014 | 0.3757185 | 0.3827899 |
| 16.3 | Misspecified Simcal | 0.3858744 | 0.0054624 | 0.3862358 | 0.3815514 | 0.3895465 |
| 16.4 | NoCal | 0.4471733 | 0.0136121 | 0.4453957 | 0.4377501 | 0.4569834 |
| 16.4 | Simcal | 0.3793453 | 0.0050923 | 0.3797014 | 0.3757185 | 0.3827899 |
| 16.4 | Misspecified Simcal | 0.3808673 | 0.0052184 | 0.3812805 | 0.3769795 | 0.3839868 |
| 16.5 | NoCal | 0.4471733 | 0.0136121 | 0.4453957 | 0.4377501 | 0.4569834 |
| 16.5 | Simcal | 0.3793453 | 0.0050923 | 0.3797014 | 0.3757185 | 0.3827899 |
| 16.5 | Misspecified Simcal | 0.3815880 | 0.0053262 | 0.3817308 | 0.3779677 | 0.3850537 |
| 16.6 | NoCal | 0.4471733 | 0.0136121 | 0.4453957 | 0.4377501 | 0.4569834 |
| 16.6 | Simcal | 0.3793453 | 0.0050923 | 0.3797014 | 0.3757185 | 0.3827899 |
| 16.6 | Misspecified Simcal | 0.3898544 | 0.0057680 | 0.3900426 | 0.3857757 | 0.3935552 |
| 16.7 | NoCal | 0.4471733 | 0.0136121 | 0.4453957 | 0.4377501 | 0.4569834 |
| 16.7 | Simcal | 0.3793453 | 0.0050923 | 0.3797014 | 0.3757185 | 0.3827899 |
| 16.7 | Misspecified Simcal | 0.4022554 | 0.0064011 | 0.4025225 | 0.3975471 | 0.4062729 |
| 16.8 | NoCal | 0.4471733 | 0.0136121 | 0.4453957 | 0.4377501 | 0.4569834 |
| 16.8 | Simcal | 0.3793453 | 0.0050923 | 0.3797014 | 0.3757185 | 0.3827899 |
| 16.8 | Misspecified Simcal | 0.4343040 | 0.0079029 | 0.4340722 | 0.4283904 | 0.4399177 |

Figure 5.1: Boxplots of LogLoss for different calibration approaches for logistic regression.

Figure 5.2: Boxplots of LogLoss for different proportion misspecifications for SimCal calibration for logistic regression.
