## Supplementary material for "Calibrating machine learning approaches for probability estimation without calibration data": Supplementary_material_7.qmd

````
---
title: "Calibration Simulation - Cleveland Data"
format: 
  html:
    self-contained: true    
    toc: true
    toc-float: yes
    smooth-scroll: true
    code-fold: true
    code-tools: true
  pdf:
    fig-dpi: 600
editor: 
  markdown: 
    wrap: 72
---


```{r setup, include=FALSE}
knitr::opts_chunk$set(echo = FALSE, warning = FALSE, message = FALSE, results='hide')
```

```{r, include=FALSE}
# Supplementary Material 6 for 
# "Calibrating machine learning approaches for probability
# estimation in case of the absence of calibration data"

# This script is adapted from the one found in supplementary material 
# 4 of Ojeda FM, et al., Calibrating machine learning approaches for probability 
# estimation: A comprehensive comparison. Stat Med. 2023 Dec 20;42(29):5451-5478
```

```{r libraries}
suppressWarnings({
  library(GLDEX)
  library(dplyr)
  library(kableExtra)
  library(readr)
  library(wesanderson)
  library(tidymodels)
  library(optparse)
  library(modgo)
  library(stringr)
  library(rms)
  library(here)
  library(patchwork)
  library(epade)
  library(FNN)
  library(resamplr)
  library(naniar)
  library(tableone)
  library(VIM)
})
```

```{r functions}
impCon <- function (df, conVars){
  for (i in conVars){
    naPos <- which(is.na(df[,i]))
    if(length(naPos) != 0){
      mean_df <- mean(df[,i], na.rm = TRUE)
      sd_df <- sd(df[,i], na.rm=T)
      n_df <- length(naPos)
      set.seed(50)
      df[naPos,i] <- rnorm(n=n_df, mean = mean_df, sd = sd_df)
    }
  }
  return (df)
}
impCat <- function (df, catVars){
  for (i in catVars){
    naPos <- which(is.na(df[,i]))
    if(length(naPos) != 0){
      temp <- df[,i]
      var <- unique(temp[!is.na(temp)])
      pvar <- NULL
      set.seed(50)
      for (j in 1:length(var)){
        pvar[j] <- length(temp[!is.na(temp) & temp == var[j]]) / length(temp)
      }
      imp <- sample(var, size=length(naPos), replace = TRUE, prob = pvar)
      df[naPos, i] <- imp
    }
  }
  return (df)
}

# Function that creates, recodes and reorders variables in Cleveland data
#===============================================================================
renameVars <- function(df){
  df$Sex <- ifelse(df$Sex == 'Female',0,1)
  df$cp <- 0
  df$cp[df$Chestpaintype == 'Typical angina'] <- 1
  df$cp[df$Chestpaintype == 'Atypical angina'] <- 2
  df$cp[df$Chestpaintype == 'Non-anginal pain'] <- 3
  df$cp[df$Chestpaintype == 'Asymptomatic'] <- 4
  df$HighFastBloodSugar<-ifelse(df$HighFastBloodSugar == 'No', 0, 1)
  df$restECG <- 0
  df$restECG[df$RestingECG == 'Normal'] <- 0
  df$restECG[df$RestingECG == 'ST-T wave abnorm'] <- 1
  df$restECG[df$RestingECG == 'Leftvent hypertrophy'] <- 2
  df$ExInducedAngina<-ifelse(df$ExInducedAngina == 'No', 0, 1)
  df$CAD <- ifelse(df$CAD == 'No', 0, 1)
  df <- df[,c(1, 2, 12, 4:6, 13, 8:11)]
  names(df)[3] <- 'Chestpaintype'
  names(df)[7] <- 'RestingECG'
  return(df)
}

# Function to reads and process Cleveland data ---------------------
# ==============================================================================
read_process_df <- function(url) {
  col.names <- c("Age", "Sex", "Chestpaintype", "RestingBP", "Cholsterol",
                 "HighFastBloodSugar", "RestingECG", "MaxHeartRate", 
                 "ExInducedAngina", "STDepression", "SlopePeakST", 
                 "NumbMajorVessels", "Thal", "CAD")
  df <- read.table(url, header = FALSE, sep = ",", col.names = col.names, 
                   na.strings = "?") %>% 
    mutate(Chestpaintype = recode(as.factor(Chestpaintype), 
                                  "1" = "Typical angina", 
                                  "2" = "Atypical angina", 
                                  "3" = "Non-anginal pain", 
                                  "4" = "Asymptomatic") ) %>% 
    mutate(HighFastBloodSugar = recode(as.factor(HighFastBloodSugar), 
                                       "1" = "Yes", "0" = "No") ) %>% 
    mutate(RestingECG = recode(as.factor(RestingECG), "0" = "Normal", 
                               "1" = "ST-T wave abnorm", 
                               "2" = "Leftvent hypertrophy") ) %>% 
    mutate(ExInducedAngina = recode(as.factor(ExInducedAngina), 
                                    "1" = "Yes", "0" = "No") ) %>% 
    mutate(SlopePeakST = recode(as.factor(SlopePeakST), 
                                "1" = "Upsloping", "2" = "Flat", 
                                "3" = "Downsloping") ) %>% 
    mutate(Thal = recode(as.factor(Thal), "6" = "Fixed defect", "3" = "Normal", 
                         "7" = "Reversable defect") ) %>% 
    mutate(NumbMajorVessels = as.numeric(NumbMajorVessels) ) %>%
    mutate(Sex = recode(as.factor(Sex), "1" = "Male", "0" = "Female") ) %>%
    mutate( CAD = as.factor( (case_when( CAD > 0 ~ "Yes", CAD == 0 ~ "No" ) ) )) 
  # Variables with too many missing values
  df <- subset(df, select=-c(SlopePeakST, Thal, NumbMajorVessels))
  # Impute missing value
  continuousVars <- c (1, 4, 5, 8, 10)
  df<-impCon(df, continuousVars)
  categoricalVars <- c(2, 3, 6, 7, 9)
  df <- impCat(df, categoricalVars)
  # creating, recoding and reordering variables
  df <- renameVars(df)
  # Adding factor version of CAD
  df[["CAD_fac"]] <- factor(df[["CAD"]])
  df
}

read_process_df_noimp <- function(url) {
  col.names <- c("Age", "Sex", "Chestpaintype", "RestingBP", "Cholsterol",
                 "HighFastBloodSugar", "RestingECG", "MaxHeartRate", 
                 "ExInducedAngina", "STDepression", "SlopePeakST", 
                 "NumbMajorVessels", "Thal", "CAD")
  df <- read.table(url, header = FALSE, sep = ",", col.names = col.names, 
                   na.strings = "?") %>% 
    mutate(Chestpaintype = recode(as.factor(Chestpaintype), 
                                  "1" = "Typical angina", 
                                  "2" = "Atypical angina", 
                                  "3" = "Non-anginal pain", 
                                  "4" = "Asymptomatic") ) %>% 
    mutate(HighFastBloodSugar = recode(as.factor(HighFastBloodSugar), 
                                       "1" = "Yes", "0" = "No") ) %>% 
    mutate(RestingECG = recode(as.factor(RestingECG), "0" = "Normal", 
                               "1" = "ST-T wave abnorm", 
                               "2" = "Leftvent hypertrophy") ) %>% 
    mutate(ExInducedAngina = recode(as.factor(ExInducedAngina), 
                                    "1" = "Yes", "0" = "No") ) %>% 
    mutate(SlopePeakST = recode(as.factor(SlopePeakST), 
                                "1" = "Upsloping", "2" = "Flat", 
                                "3" = "Downsloping") ) %>% 
    mutate(Thal = recode(as.factor(Thal), "6" = "Fixed defect", "3" = "Normal", 
                         "7" = "Reversable defect") ) %>% 
    mutate(NumbMajorVessels = as.numeric(NumbMajorVessels) ) %>%
    mutate(Sex = recode(as.factor(Sex), "1" = "Male", "0" = "Female") ) %>%
    mutate( CAD = as.factor( (case_when( CAD > 0 ~ "Yes", CAD == 0 ~ "No" ) ) )) 
  # Variables with too many missing values
  df <- subset(df, select=-c(SlopePeakST, Thal, NumbMajorVessels))
  # Impute missing value
  #continuousVars <- c (1, 4, 5, 8, 10)
  #df<-impCon(df, continuousVars)
  #categoricalVars <- c(2, 3, 6, 7, 9)
  #df <- impCat(df, categoricalVars)
  # creating, recoding and reordering variables
  df <- renameVars(df)
  # Adding factor version of CAD
  df[["CAD_fac"]] <- factor(df[["CAD"]])
  df
}


brier <- function(p_true, p_hat, winsorize = TRUE) {
  if(winsorize) {
    p_hat <- winsorize_probs(p_hat)
  } 
  
  sqrt(mean((p_true  - p_hat)^2, na.rm = TRUE))
}

logloss <- function(p_true, p_hat, winsorize = TRUE) {
  if(winsorize) {
    p_hat <- winsorize_probs(p_hat)
  } 
  
  (-1) * mean(p_true * log(p_hat) + (1 - p_true) * log(1 - p_hat), na.rm = TRUE)
}


# Functions for calibration ----------------------------------------------------
# ==============================================================================
# Functions to deal with 0 or 1 predictions
winsorize_probs <- function(x) {
  eps <- .Machine$double.eps
  pmax(pmin(x, 1 - eps), eps)
}

# Functions for calibration
cal_elkan <- function(response, traindata, caldata, newdata, prob) {
  br0 <- mean(traindata[, response])
  br1 <- mean(caldata[, response])
  prob.new <- newdata[, prob] %>% winsorize_probs()
  br1 * (prob.new - prob.new * br0) / (br0 - prob.new * br0 + br1 * prob.new - br0 * br1)
}
cal_logistic <- function(response, traindata, caldata, newdata, prob) {
  prob1 <- caldata[, prob]%>% winsorize_probs()
  prob2 <- newdata[, prob]%>% winsorize_probs()
  caldata[["lp"]] <- log(prob1 / (1 - prob1))
  newdata[["lp"]] <- log(prob2 / (1 - prob2))
  Formula <- paste0(response, " ~ lp") %>% as.formula
  fit <- glm(Formula, data = caldata, family = binomial())
  stats::predict(fit, newdata = newdata, type = "response")
}

cal_knearest <- function(train_data, test_data, prob_col, n.neighbors = 10){
  p_cal <- NULL
  # loop observations (translate t lapply later)
  for (i in 1:nrow(test_data)) {
    query_point <- test_data[i, c(vars_to_use, prob_col), drop = FALSE]
    neighbors <- get.knnx(data = train_data[,c(vars_to_use, prob_col)], query = query_point, k = n.neighbors)  
    neighbor_indices <- neighbors$nn.index
    neighbor_train <- train_data[neighbor_indices,]
    p_cal[i] <- sum(neighbor_train[prob_col])/n.neighbors
  }
  
  return(p_cal)
}


invlogit <- function(x) 
{ 
 return( exp(x) / (1 + exp(x) ) ) 
}

cal_acp <- function(train_data, train.outcome_col, test_data, alpha=0.05) 
{ 
 train.x = train_data 
 train.y = train.outcome_col 
 test.x = test_data 
 # (1-alpha)% critical term 
 crit.term = qnorm(1 - alpha/2) 
 
 # create a training set 
 da.train <- data.frame(train.x, response=train.y) 
 # train the model 
 m.lr = glm( response ~ ., data=da.train, family="binomial") 
 train = list() 
 train$pred = predict(m.lr, newdata=da.train, se.fit=TRUE) 
 train$yhat = invlogit(train$pred$fit) 
 
 # apply the trained model to a test data 
 test = list() 
 test$pred = predict(m.lr, newdata=test.x, se.fit=TRUE) 
 n.test = nrow(test.x) 
 # prediction interval for the test set 
 test$yhat = invlogit( test$pred$fit ) 
 test$lower = invlogit( test$pred$fit - crit.term * test$pred$se) 
 test$upper = invlogit( test$pred$fit + crit.term * test$pred$se) 
 test$ublb = cbind(lower=test$lower, upper=test$upper) 
 # prediction range 
 r = max(train$yhat) - min(train$yhat) 
 # covariance matrix for coefficients
 beta.covmat = as.matrix( summary(m.lr)$cov.unscaled ) 
 # Equation 5 
 X.mat = as.matrix( cbind(1, test.x) ) 
 logit.covmat = X.mat %*% beta.covmat %*% t(X.mat) 
 vxb = diag(logit.covmat) 
 idxSE = sqrt(vxb) 
 w = as.numeric( m.lr$coef ) 
 idxLower = X.mat %*% w - crit.term * idxSE * r 
 idxUpper = X.mat %*% w + crit.term * idxSE * r 
 # Step 2 
 lb = invlogit( idxLower ) 
 ub = invlogit( idxUpper ) 
 # Step 3 & 4 
 p.calib = rep(0, nrow(test.x)) 
 for(i in 1:n.test) { 
 idx.WithinCI = ( lb[i] < train$yhat) & (train$yhat < ub[i]) 
 n.trainedPredWithinCI = sum(idx.WithinCI) 
 
 if(n.trainedPredWithinCI == 0 ) { 
 dist.testpoint = (train$yhat - test$yhat[i])^2 
 dist.minimum = min(dist.testpoint) 
 isMinimizer = dist.testpoint == dist.minimum 
 idx.WithinCI = c(1:length(dist.testpoint))[isMinimizer] 
 n.trainedPredWithinCI = length(idx.WithinCI)
 } 
 p.calib[i] = sum(m.lr$y[idx.WithinCI]) / n.trainedPredWithinCI 
 } 
 return(p.calib) 
}


format_mean_sd <- function (Mean, Sd, round_dec) 
{
  paste0(epade::round_n.ade(Mean, round_dec), " ± ", epade::round_n.ade(Sd, 
    round_dec))
}

# SimCal calibration functions ------------------------------------------------- 

add_recal_probs <- function(df_original,
                            df_sim,
                            pred_model = c("lr", "rf", "gb"),
                            outcome_col) {
  pred_model <- match.arg(pred_model)
  prob_col <- paste0("prob_", pred_model)
  
  Formula <-
    paste0(outcome_col, " ~ I( log(", prob_col , "/(1-", prob_col, ")))") %>%
    as.formula()
   
  df_sim[[prob_col]][df_sim[[prob_col]] == 0] <- 10^(-8)

  
  fit <- glm(Formula, data = df_sim, family = binomial())
  prob_col_new <- paste0(prob_col, "_recal")
  
  df_original[[prob_col_new]] <-
      stats::predict.glm(fit, newdata = df_original,
                         type = "response")
  
  rm(prob_col, prob_col_new)
  df_original
    
}

add_recal_wrapper <-
  function(df_sim,
           df_original,
           df_calibrator,
           vars_modgo_cont,
           vars_modgo_bin,
           vars_modgo_cat,
           outcome_col) {
    
    tab_mean_sd <- NULL
    for (j in vars_modgo_cont) {
      df_sim[[j]] <-
        modgo::rbi_normal_transform_inv(df_sim[[j]], df_calibrator[[j]])
      
      # updating mean and sd of simulated data for continuous variables
      # using new study information
      
      mean_new <- mean(df_calibrator[[j]])
      mean_study <- mean(df_original[[j]])
      
      
      sd_new <- sd(df_calibrator[[j]])
      sd_study <- sd(df_original[[j]])
      
      df_sim[[j]] <-
        ((df_sim[[j]] - mean_new) / sd_new) * sd_study +
        mean_study
      
      
      if (j == "Age") {
        df_sim[[j]] <- as.integer(df_sim[[j]])
        }
      
    }
    
    # processing binary variables using the cutoffs of the new study
    
    for (j in c(vars_modgo_bin, vars_modgo_cat)) {
      df_sim[[j]] <-
        modgo::rbi_normal_transform_inv(df_sim[[j]], df_original[[j]])
    }
    
    df_sim[["prob_rf"]] <- stats::predict(fit_rf, new_data = df_sim,
                                   type = "prob")[[".pred_1"]]
    df_sim$prob_lr <-
      plogis(stats::predict(lr.cleve[["fit"]], newdata = df_sim))
    df_sim[["prob_gb"]] <- stats::predict(fit_gb, new_data = df_sim,
                                   type = "prob")[[".pred_1"]]
    
    for (j in c("lr", "rf", "gb")) {
    df_original <- add_recal_probs(df_original, df_sim,  pred_model = j, outcome_col = outcome_col)
    }
    
    df_original 

  }
```

```{r data-prep}

# Reading and processing datasets ----------------------------------------
# ==============================================================================
url_root <- paste0("https://archive.ics.uci.edu/ml/machine-learning-databases/",
                   "heart-disease/processed.")
url_df <- data.frame(df_name = c("Cleve", "Hung", "Swiss", "VA"),
                     url = c(paste0(url_root, c("cleveland", "hungarian", 
                                                "switzerland", "va"), ".data")), 
                     stringsAsFactors = FALSE)
url_df_miss <- data.frame(df_name = c("Cleve_miss", "Hung_miss", "Swiss_miss", "VA_miss"),
                     url = c(paste0(url_root, c("cleveland", "hungarian", 
                                                "switzerland", "va"), ".data")), 
                     stringsAsFactors = FALSE)

for(i in 1:nrow(url_df)) {
  assign(url_df[["df_name"]][i], read_process_df(url_df[["url"]][i]))
}

for(i in 1:nrow(url_df)) {
  assign(url_df_miss[["df_name"]][i], read_process_df_noimp(url_df[["url"]][i]))
}


# Fitting machines on the Cleveland data ---------------------------------------
# ==============================================================================
# Train and test data 
df_train <- Cleve
for(i in 2:nrow(url_df)) {
  study_name <- url_df[["df_name"]][i]
  df <- get(study_name)
  df[["study"]] <- study_name
  assign(study_name, df); rm(df, study_name)
}
df_test <- rbind(Hung, Swiss, VA)
# Creating formula
Formula <- paste0("CAD_fac ~ ", paste(setdiff(names(df_train), 
                                              c("CAD", "CAD_fac")), 
                                      collapse = "+")) %>% as.formula
# Creating folds for cross-validation
set.seed(50)
folds <- vfold_cv(df_train, v = 2, repeats = 2)

# Missingness plots -----------------------------------------------------------

for(i in 1:nrow(url_df_miss)) {
  study_name <- url_df_miss[["df_name"]][i]
  df <- get(study_name)
  study_label <- url_df[["df_name"]][i]
  df[["study"]] <- study_label
  assign(study_name, df); rm(df, study_name)
}


df_full_miss <- rbind(Cleve_miss, Hung_miss, Swiss_miss, VA_miss)

gg_miss_fct(x = df_full_miss, fct = study) +
  scale_fill_gradient(
    low = "steelblue",
    high = "goldenrod",
    name = "% missing"
  )

fac <- c("Sex", "Chestpaintype", "HighFastBloodSugar", "RestingECG", "ExInducedAngina")
df_full_miss[fac] <- lapply(df_full_miss[fac], factor)
descriptives <- data.frame(print(CreateTableOne(vars = colnames(df_full_miss), 
                                             data = df_full_miss, 
                                             strata = "study"),
                           catDigits = 2, contDigits = 2, nonnormal=F))[1:4]

# Cleveland center has no missing data

#Hung center
vis_miss(Hung_miss[,1:11]) + theme(
  axis.text.x = element_text(
    angle = 90,
    hjust = 0,
    vjust = 0
  ))
gg_miss_var(Hung_miss[,1:11])
p <- gg_miss_upset(Hung_miss[,1:11])
p

#Swiss center
vis_miss(Swiss_miss[,1:11]) + theme(
  axis.text.x = element_text(
    angle = 90,
    hjust = 0,
    vjust = 0
  ))
gg_miss_var(Swiss_miss[,1:11])
p <- gg_miss_upset(Swiss_miss[,1:11])
p

#VA center
vis_miss(VA_miss[,1:11]) + theme(
  axis.text.x = element_text(
    angle = 90,
    hjust = 0,
    vjust = 0
  ))
gg_miss_var(VA_miss[,1:11])
p <- gg_miss_upset(VA_miss[,1:11])
p
```


```{r fitting-machines}
grid_size <- 50
# Random forest ----------------------------------------------------------------
# ==============================================================================
set.seed(50)
spec_rf <- rand_forest(mtry = tune(), trees = 500,
                       min_n = tune()) %>%
  set_engine("ranger") %>%
  set_mode("classification") %>% set_args(seed = NULL)
# random forest grid. Adding the parameters used in the original publication to the grid
vars <- all.vars(Formula)[-1] # these are the predictors
grid_rf <- dials::grid_latin_hypercube(
  finalize(dials::mtry(), 
           x = df_train[, vars, drop = FALSE]),
  dials::min_n(),
  size = grid_size
)
grid_rf_aux <- grid_rf[1, ]
grid_rf_aux[["mtry"]] <- floor(sqrt(ncol(df_train[, vars, 
                                                  drop = FALSE])))
grid_rf_aux[["min_n"]] <- floor(nrow(df_train) * 0.05)
grid_rf <- rbind(grid_rf, grid_rf_aux)
grid_rf <- grid_rf[!duplicated(grid_rf), ]
rm(grid_rf_aux)
# Creating workflow
wf_rf <- workflows::workflow() %>%
  add_model(spec_rf) %>% 
  add_formula(Formula)
# tuning model
rf_tuned <- tune::tune_grid(
  object = wf_rf,
  resamples = folds,
  grid = grid_rf,
  metrics = metric_set(mn_log_loss),
  control = tune::control_grid(verbose = TRUE, 
                               allow_par = FALSE))
best_log_loss_rf <- select_best(rf_tuned, metric ="mn_log_loss")
tuned_final_rf <- finalize_workflow(wf_rf, best_log_loss_rf)
fit_rf <- tuned_final_rf  %>% fit(data = df_train)
df_test[["prob_rf"]] <- predict(fit_rf, new_data = df_test, 
                                type = "prob")[[".pred_1"]]

# Gradient boost ---------------------------------------------------------------
# ==============================================================================
grid_size <- 50
set.seed(50)
# specifying engine and parameters to tune
spec_gb <- boost_tree(learn_rate = tune(),
                      tree_depth = tune()) %>% 
  set_engine("xgboost") %>% 
  set_mode("classification")
# specifying grid for hyperparameter tuning
grid_gb <- dials::grid_latin_hypercube(
  dials::learn_rate(),tree_depth(),
  size = grid_size)
# Creating workflow
wf_gb <- workflows::workflow() %>%
  add_model(spec_gb) %>% 
  add_formula(Formula)
# tuning model
gb_tuned <- tune::tune_grid(
  object = wf_gb,
  resamples = folds,
  grid = grid_gb,
  metrics = metric_set(mn_log_loss),
  control = tune::control_grid(verbose = TRUE, 
                               allow_par = FALSE))
best_log_loss_gb <- select_best(gb_tuned, metric = "mn_log_loss")
tuned_final_gb <- finalize_workflow(wf_gb, best_log_loss_gb) 
fit_gb <- tuned_final_gb  %>% fit(data = df_train)
df_test[["prob_gb"]] <- predict(fit_gb, new_data = df_test, 
                                type = "prob")[[".pred_1"]]

# Logistic regression ----------------------------------------------------------
# ==============================================================================
stepPvalue <- function(df, p.lim){
  fit <- glm(as.factor(CAD)~., data = df, family = binomial)
  coef <- summary(fit)$coefficients
  while(any(coef[-1,4] >= p.lim)){
    fit <- glm(as.factor(CAD)~., data = df, family = binomial)
    coef <- summary(fit)$coefficients
    removeVar <- which(max(coef[-1,4]) == coef[,4])
    df <- df[-which(names(df) == names(removeVar))]
  }
  returnList <- list(fit = fit, df = df, coef = coef)
  return(returnList)
}
library(mfp)
mfp(CAD ~ Age + RestingBP + Cholsterol + MaxHeartRate + STDepression, Cleve)
lr.cleve<-stepPvalue(Cleve[,-12],0.001)
df_test[["prob_lr"]] <- plogis(predict(lr.cleve[["fit"]], newdata=df_test))


```

```{r validation-calibration}
# Creating folds for 10-fold cross-validation
# ==============================================================================
set.seed(50)
fold_hung<-vfold_cv(Hung, v=10, repeats = 1, strata='CAD')
Hung$cv<-0
for (i in 1:10){
  Hung$cv[-fold_hung$splits[[i]]$in_id]=i
}
fold_swiss<-vfold_cv(Swiss, v=10, repeats = 1, strata='CAD')
Swiss$cv<-0
for (i in 1:10){
  Swiss$cv[-fold_swiss$splits[[i]]$in_id]=i
}
fold_va<-vfold_cv(VA, v=10, repeats = 1, strata='CAD')
VA$cv<-0
for (i in 1:10){
  VA$cv[-fold_va$splits[[i]]$in_id]=i
}
df_test$cv<-c(Hung$cv, Swiss$cv, VA$cv)
df_test$id<-1:nrow(df_test)

# Calibration
# ==============================================================================
df_app <- df_test
df_test <- df_app
vars_to_use <-c("Age", "Sex", "Chestpaintype", "RestingBP", "Cholsterol", "HighFastBloodSugar", 
"RestingECG", "MaxHeartRate", "ExInducedAngina", "STDepression")
machine<-c('lr','rf','gb')
cal.out<-df_test
set.seed(50)
for (j in machine){
  out<-NULL
  for (i in 1:10){
    cal<-cal.out[cal.out$cv!=i,]
    new<-cal.out[cal.out$cv==i,]
    new[[paste0('prob_',j,'_elkan')]]<-cal_elkan ('CAD', df_train, cal, new, paste0('prob_',j))
    new[[paste0('prob_',j,'_log')]]<-cal_logistic ('CAD', df_train, cal, new, paste0('prob_',j))
    new[[paste0("prob_",j,"_sba")]]<-cal_knearest(train_data = cal[,c(vars_to_use, "CAD", paste0('prob_',j))],
                                                 test_data = new[,c(vars_to_use, "CAD", paste0('prob_',j))],
                                                 prob_col = paste0('prob_',j),
                                                 n.neighbors = 10)
    if (j == "lr") {
      new[[paste0('prob_',j,'_acp')]]<-cal_acp(train_data = cal[,c(vars_to_use)], 
                                               train.outcome_col = cal[,"CAD"], 
                                               test_data = new[,c(vars_to_use)])
      }
    out<-rbind(out,new)
  }
  df_test <- merge(df_test, out[, c(
    "id",
    paste0('prob_', j, '_elkan'),
    paste0('prob_', j, '_log'),
    paste0('prob_', j, '_sba')
  )], by = 'id')
  if (j == "lr") {
    df_test <- merge(df_test, out[, c("id", paste0('prob_', j, '_acp'))], by =
                       'id')
  }
}
# Refit
refit<-df_test[,c(1:18)]
refit.out<-NULL
for (i in 1:10){
  cal<-refit[refit$cv!=i,]
  new<-refit[refit$cv==i,]
  set.seed(50)
  folds <- vfold_cv(cal, v = 2, repeats = 2)
  grid_size <- 50
  set.seed(50)
  spec_rf <- rand_forest(mtry = tune(), trees = 500,
                         min_n = tune()) %>%
    set_engine("ranger") %>%
    set_mode("classification") %>% set_args(seed = NULL)
  grid_rf <- dials::grid_latin_hypercube(
    finalize(dials::mtry(),
             x = cal[, vars, drop = FALSE]),
    dials::min_n(),
    size = grid_size
  )
  grid_rf_aux <- grid_rf[1, ]
  grid_rf_aux[["mtry"]] <- floor(sqrt(ncol(cal[, vars,
                                               drop = FALSE])))
  grid_rf_aux[["min_n"]] <- floor(nrow(cal) * 0.05)
  grid_rf <- rbind(grid_rf, grid_rf_aux)
  grid_rf <- grid_rf[!duplicated(grid_rf), ]
  rm(grid_rf_aux)
  wf_rf <- workflows::workflow() %>%
    add_model(spec_rf) %>%
    add_formula(Formula)
  rf_tuned <- tune::tune_grid(
    object = wf_rf,
    resamples = folds,
    grid = grid_rf,
    metrics = metric_set(mn_log_loss),
    control = tune::control_grid(verbose = TRUE,
                                 allow_par = FALSE))
  best_log_loss_rf <- select_best(rf_tuned, metric = "mn_log_loss")
  tuned_final_rf <- finalize_workflow(wf_rf, best_log_loss_rf)
  fit_rf <- tuned_final_rf  %>% fit(data = cal)
  new[["prob_rf_refit"]] <- predict(fit_rf, new_data = new,
                                    type = "prob")[[".pred_1"]]
  grid_size <- 50
  set.seed(50)
  spec_gb <- boost_tree(learn_rate = tune(),
                        tree_depth = tune()) %>%
    set_engine("xgboost") %>%
    set_mode("classification")
  grid_gb <- dials::grid_latin_hypercube(
    dials::learn_rate(),tree_depth(),
    size = grid_size)
  wf_gb <- workflows::workflow() %>%
    add_model(spec_gb) %>%
    add_formula(Formula)
  gb_tuned <- tune::tune_grid(
    object = wf_gb,
    resamples = folds,
    grid = grid_gb,
    metrics = metric_set(mn_log_loss),
    control = tune::control_grid(verbose = TRUE,
                                 allow_par = FALSE))
  best_log_loss_gb <- select_best(gb_tuned, metric = "mn_log_loss")
  tuned_final_gb <- finalize_workflow(wf_gb, best_log_loss_gb)
  fit_gb <- tuned_final_gb  %>% fit(data = cal)
  new[["prob_gb_refit"]] <- predict(fit_gb, new_data = new,
                                    type = "prob")[[".pred_1"]]
  lr.refit<-stepPvalue(cal[,c(2:12)],0.001)
  new[["prob_lr_refit"]] <- plogis(predict(lr.refit[["fit"]], newdata=new))
  refit.out<-rbind(refit.out, new)
}

df_test <- merge(df_test, refit.out[, c(
    "id",
    'prob_lr_refit',
    'prob_rf_refit',
    'prob_gb_refit')])
```

```{r simcal, results='asis'}
df_train.sim <- df_train %>% dplyr::select(-CAD_fac)
vars_modgo <- colnames(df_train.sim)
vars_modgo_bin <- c("Sex", "HighFastBloodSugar", "ExInducedAngina", "CAD")
vars_modgo_cont <- c("Age", "RestingBP", "Cholsterol", "MaxHeartRate", "STDepression" )
vars_modgo_cat <- c("Chestpaintype", "RestingECG")
vars_modgo_cont <- setdiff(vars_modgo_cont, vars_modgo_cat)
# SimCal Preparation -----------------------------------------------------------
# simulate data
res_modgo <- modgo(data = df_train.sim, 
                   variables = vars_modgo, 
                   bin_variables = vars_modgo_bin,
                   categ_variables = vars_modgo_cat, nrep = 1)
norm_cor_matrix <- res_modgo[["covariance_matrix"]]

n_samples <- 100000L 
set.seed(1234)

# Simulating data using that correlation matrix estimated by modgo
norm_sim <- MASS::mvrnorm(n = n_samples, mu = rep(0, ncol(norm_cor_matrix)), 
                          Sigma = norm_cor_matrix) %>% data.frame() 

# Add SimCal probabilities =====================================================

df_res <- add_recal_wrapper(
  df_sim = norm_sim,            # Dataset simulated from the correlation matrix.
  df_original = df_test,        # Dataset with model variables and 
                                # probabilities to calibrate.
  df_calibrator = df_train.sim, # Dataset used to calibrate, i.e. training data.
  vars_modgo_cont,              # List of continuous model variables.
  vars_modgo_bin,               # List of binary model variables.
  vars_modgo_cat,               # List of categorical model variables.
  outcome_col = "CAD"           # Model outcome. 
)

```

# Calibration plots 

```{r figures, results='asis'}
#Fig ===========================================================================
df_plot <- c()
machine_abb <- c("lr", "rf", "gb")
machine_labels <- c("Logistic regression", "Random forest", "Gradient boosting")
for (j in 1:3) {
  
  if (j ==1) {
    vars <- c(
      "study",
      "CAD",
      paste0("prob_", machine_abb[[j]]),
      paste0("prob_", machine_abb[[j]], "_log"),
      paste0("prob_", machine_abb[[j]], "_refit"),
      paste0("prob_", machine_abb[[j]], "_sba"),
      paste0("prob_", machine_abb[[j]], "_elkan"),
      paste0("prob_", machine_abb[[j]], "_recal"),
      paste0("prob_", machine_abb[[j]], "_acp")
    )
    
        Levels <-
    paste0("prob_", machine_abb[[j]], c("",  "_log", "_refit", "_sba","_elkan", "_recal",  "_acp"))
  Labels <- c("No recalibration",
              "Logistic",
              "Refit",
             "SBA-10",
              "Elkan",
              "SimCal",
             "ACP")
  } else {
    vars <- c(
      "study",
      "CAD",
      paste0("prob_", machine_abb[[j]]),
      paste0("prob_", machine_abb[[j]], "_log"),
      paste0("prob_", machine_abb[[j]], "_refit"),
      paste0("prob_", machine_abb[[j]], "_sba"),
      paste0("prob_", machine_abb[[j]], "_elkan"),
      paste0("prob_", machine_abb[[j]], "_recal")
    )
      
        Levels <-
    paste0("prob_", machine_abb[[j]], c("",  "_log", "_refit", "_sba" , "_elkan", "_recal"))
  Labels <- c("No recalibration",
              "Logistic",
              "Refit",
             "SBA-10",
              "Elkan",
              "SimCal")
  }
  
  
  df_long <-
    pivot_longer(data = df_res[, vars], cols = starts_with("prob"))
  
  df_long[["method"]] <- factor(df_long[["name"]],
                                levels = Levels, labels = Labels)
  df_long[["machine"]] <- machine_labels[[j]]
  
  df_plot <- bind_rows(df_plot, df_long)
  
}

df_plot <- transform(df_plot,
      machine=factor(machine,levels=c("Logistic regression", "Random forest", "Gradient boosting")))
  
epsilon <- 0.01


```

## Logistic regression 

### Overall

```{r}
df_plot_lr <- df_plot %>%  filter(machine == "Logistic regression")
df_plot_lr$study <- as.factor(df_plot_lr$study)
COLS = c("steelblue","darkgreen","orange")
names(COLS) <- levels(df_plot_lr$study)
# 
# pdf(
#   "CleveClinicsLR.pdf",
#   width = 10,
#   height = 6
# )

ggplot(df_plot_lr,
       aes(x = value, y = CAD,
           colour = study,
           fill = study)) +
  # Center-specific smooths
  geom_smooth(method = "gam",
              span = 0.75,
              lty = 1,
              se = FALSE,
              linewidth = 0.7) +
  # Overall pooled smooth
geom_smooth(
  data = df_plot_lr,
  aes(x = value, y = CAD,
      group = 1,
      colour = "Average"),
  inherit.aes = FALSE,
  method = "gam",
  linewidth = 1.2,
  linetype = 2,
  se = FALSE
) +
  facet_wrap(~method, ncol = 4, nrow = 2) +
  coord_fixed(ratio = 1) +
  scale_x_continuous(
    name = "Predicted probability",
    limits = c(0 - epsilon, 1 + epsilon)
  ) +
  scale_y_continuous(
    name = "True probability",
    limits = c(0 - epsilon, 1 + epsilon)
  ) +
  theme_bw() +
  theme(
    strip.background = element_rect(fill = "gray95"),
    panel.spacing.x = unit(0.6, "lines"),
    axis.text = element_text(size = 6.75)
  ) +
  scale_color_manual(
  name = "Clinic",
  values = c(COLS, Average = "black"),
  breaks = c(names(COLS), "Average")
) +
guides(
  fill = "none"
)

#dev.off()

```

### Hung

```{r}
df_plot_lr <- df_plot %>%  filter(machine == "Logistic regression" & study == "Hung")
ggplot(df_plot_lr, aes(x = value, y = CAD)) + 
        geom_abline(slope = 1, 
                    intercept = 0, 
                    lty = 1, 
                    colour = "black") +
        geom_smooth(method = "gam", span = 0.75,
                    lty = 2, colour = "black") +
       facet_wrap(~ method, ncol = 4, nrow = 2) +
        coord_fixed(ratio = 1) +
        scale_x_continuous(name = "Predicted probability", 
                           limits = c(0 - epsilon, 1 + epsilon)) + 
        scale_y_continuous(name = "True probability", 
                           limits = c(0 - epsilon, 1 + epsilon)) + 
        theme_bw() +  theme(strip.background = element_rect(fill = "gray95"),
                            panel.spacing.x = unit(0.6, "lines"),
                            axis.text = element_text(size = 6.75))
```

### Swiss

```{r}
df_plot_lr <- df_plot %>%  filter(machine == "Logistic regression" & study == "Swiss")
ggplot(df_plot_lr, aes(x = value, y = CAD)) + 
        geom_abline(slope = 1, 
                    intercept = 0, 
                    lty = 1, 
                    colour = "black") +
        geom_smooth(method = "gam", span = 0.75,
                    lty = 2, colour = "black") +
       facet_wrap(~ method, ncol = 4, nrow = 2) +
        coord_fixed(ratio = 1) +
        scale_x_continuous(name = "Predicted probability", 
                           limits = c(0 - epsilon, 1 + epsilon)) + 
        scale_y_continuous(name = "True probability", 
                           limits = c(0 - epsilon, 1 + epsilon)) + 
        theme_bw() +  theme(strip.background = element_rect(fill = "gray95"),
                            panel.spacing.x = unit(0.6, "lines"),
                            axis.text = element_text(size = 6.75))
```

### VA

```{r}
df_plot_lr <- df_plot %>%  filter(machine == "Logistic regression" & study == "VA")
ggplot(df_plot_lr, aes(x = value, y = CAD)) + 
        geom_abline(slope = 1, 
                    intercept = 0, 
                    lty = 1, 
                    colour = "black") +
        geom_smooth(method = "gam", span = 0.75,
                    lty = 2, colour = "black") +
       facet_wrap(~ method, ncol = 4, nrow = 2) +
        coord_fixed(ratio = 1) +
        scale_x_continuous(name = "Predicted probability", 
                           limits = c(0 - epsilon, 1 + epsilon)) + 
        scale_y_continuous(name = "True probability", 
                           limits = c(0 - epsilon, 1 + epsilon)) + 
        theme_bw() +  theme(strip.background = element_rect(fill = "gray95"),
                            panel.spacing.x = unit(0.6, "lines"),
                            axis.text = element_text(size = 6.75))
```

## Random forest

### Overall

```{r}
df_plot_rf <- df_plot %>%  filter(machine == "Random forest")
df_plot_rf$study <- as.factor(df_plot_rf$study)
COLS = c("steelblue","darkgreen","orange")
names(COLS) <- levels(df_plot_lr$study)
# 
# pdf(
#   "CleveClinicsRF.pdf",
#   width = 10,
#   height = 6
# )
ggplot(df_plot_rf,
       aes(x = value, y = CAD,
           colour = study,
           fill = study)) +
  # geom_abline(slope = 1,
  #             intercept = 0,
  #             lty = 1,
  #             colour = "black",
  #             linewidth = 0.5) +
  # Center-specific smooths
  geom_smooth(method = "gam",
              span = 0.75,
              lty = 1,
              se = FALSE,
              linewidth = 0.7) +
  # Overall pooled smooth
geom_smooth(
  data = df_plot_rf,
  aes(x = value, y = CAD,
      group = 1,
      colour = "Average"),
  inherit.aes = FALSE,
  method = "gam",
  linewidth = 1.2,
  linetype = 2,
  se = FALSE
) +
  facet_wrap(~method, ncol = 4, nrow = 2) +
  coord_fixed(ratio = 1) +
  scale_x_continuous(
    name = "Predicted probability",
    limits = c(0 - epsilon, 1 + epsilon)
  ) +
  scale_y_continuous(
    name = "True probability",
    limits = c(0 - epsilon, 1 + epsilon)
  ) +
  theme_bw() +
  theme(
    strip.background = element_rect(fill = "gray95"),
    panel.spacing.x = unit(0.6, "lines"),
    axis.text = element_text(size = 6.75)
  ) +
  #scale_fill_manual(name = "Clinic", values = c(COLS, Average = "black")) +
  scale_color_manual(
  name = "Clinic",
  values = c(COLS, Average = "black"),
  breaks = c(names(COLS), "Average")
) +
guides(
  fill = "none"
)

#dev.off()

```

### Hung

```{r}
df_plot_rf <- df_plot %>%  filter(machine == "Random forest" & study == "Hung")
ggplot(df_plot_rf, aes(x = value, y = CAD)) + 
        geom_abline(slope = 1, 
                    intercept = 0, 
                    lty = 1, 
                    colour = "black") +
        geom_smooth(method = "gam", span = 0.75,
                    lty = 2, colour = "black") +
       facet_wrap(~ method, ncol = 4, nrow = 2) +
        coord_fixed(ratio = 1) +
        scale_x_continuous(name = "Predicted probability", 
                           limits = c(0 - epsilon, 1 + epsilon)) + 
        scale_y_continuous(name = "True probability", 
                           limits = c(0 - epsilon, 1 + epsilon)) + 
        theme_bw() +  theme(strip.background = element_rect(fill = "gray95"),
                            panel.spacing.x = unit(0.6, "lines"),
                            axis.text = element_text(size = 6.75))
```

### Swiss

```{r}
df_plot_rf <- df_plot %>%  filter(machine == "Random forest" & study == "Swiss")
ggplot(df_plot_rf, aes(x = value, y = CAD)) + 
        geom_abline(slope = 1, 
                    intercept = 0, 
                    lty = 1, 
                    colour = "black") +
        geom_smooth(method = "gam", span = 0.75,
                    lty = 2, colour = "black") +
       facet_wrap(~ method, ncol = 4, nrow = 2) +
        coord_fixed(ratio = 1) +
        scale_x_continuous(name = "Predicted probability", 
                           limits = c(0 - epsilon, 1 + epsilon)) + 
        scale_y_continuous(name = "True probability", 
                           limits = c(0 - epsilon, 1 + epsilon)) + 
        theme_bw() +  theme(strip.background = element_rect(fill = "gray95"),
                            panel.spacing.x = unit(0.6, "lines"),
                            axis.text = element_text(size = 6.75))
```

### VA

```{r}
df_plot_rf <- df_plot %>%  filter(machine == "Random forest" & study == "VA")
ggplot(df_plot_rf, aes(x = value, y = CAD)) + 
        geom_abline(slope = 1, 
                    intercept = 0, 
                    lty = 1, 
                    colour = "black") +
        geom_smooth(method = "gam", span = 0.75,
                    lty = 2, colour = "black") +
       facet_wrap(~ method, ncol = 4, nrow = 2) +
        coord_fixed(ratio = 1) +
        scale_x_continuous(name = "Predicted probability", 
                           limits = c(0 - epsilon, 1 + epsilon)) + 
        scale_y_continuous(name = "True probability", 
                           limits = c(0 - epsilon, 1 + epsilon)) + 
        theme_bw() +  theme(strip.background = element_rect(fill = "gray95"),
                            panel.spacing.x = unit(0.6, "lines"),
                            axis.text = element_text(size = 6.75))
```

## Gradient boosting

### Overall

```{r}
df_plot_gb <- df_plot %>%  filter(machine == "Gradient boosting")
df_plot_gb$study <- as.factor(df_plot_gb$study)
COLS = c("steelblue","darkgreen","orange")
names(COLS) <- levels(df_plot_gb$study)

# pdf(
#   "CleveClinicsGB.pdf",
#   width = 10,
#   height = 6
# )
ggplot(df_plot_gb,
       aes(x = value, y = CAD,
           colour = study,
           fill = study)) +
  # geom_abline(slope = 1,
  #             intercept = 0,
  #             lty = 1,
  #             colour = "black",
  #             linewidth = 0.5) +
  # Center-specific smooths
  geom_smooth(method = "gam",
              span = 0.75,
              lty = 1,
              se = FALSE,
              linewidth = 0.7) +
  # Overall pooled smooth
geom_smooth(
  data = df_plot_gb,
  aes(x = value, y = CAD,
      group = 1,
      colour = "Average"),
  inherit.aes = FALSE,
  method = "gam",
  linewidth = 1.2,
  linetype = 2,
  se = FALSE
) +
  facet_wrap(~method, ncol = 4, nrow = 2) +
  coord_fixed(ratio = 1) +
  scale_x_continuous(
    name = "Predicted probability",
    limits = c(0 - epsilon, 1 + epsilon)
  ) +
  scale_y_continuous(
    name = "True probability",
    limits = c(0 - epsilon, 1 + epsilon)
  ) +
  theme_bw() +
  theme(
    strip.background = element_rect(fill = "gray95"),
    panel.spacing.x = unit(0.6, "lines"),
    axis.text = element_text(size = 6.75)
  ) +
  #scale_fill_manual(name = "Clinic", values = c(COLS, Average = "black")) +
  scale_color_manual(
  name = "Clinic",
  values = c(COLS, Average = "black"),
  breaks = c(names(COLS), "Average")
) +
guides(
  fill = "none"
)

# dev.off()

```


### Hung

```{r}
df_plot_gb <- df_plot %>%  filter(machine == "Gradient boosting" & study == "Hung")
ggplot(df_plot_gb, aes(x = value, y = CAD)) + 
        geom_abline(slope = 1, 
                    intercept = 0, 
                    lty = 1, 
                    colour = "black") +
        geom_smooth(method = "gam", span = 0.75,
                    lty = 2, colour = "black") +
       facet_wrap(~ method, ncol = 4, nrow = 2) +
        coord_fixed(ratio = 1) +
        scale_x_continuous(name = "Predicted probability", 
                           limits = c(0 - epsilon, 1 + epsilon)) + 
        scale_y_continuous(name = "True probability", 
                           limits = c(0 - epsilon, 1 + epsilon)) + 
        theme_bw() +  theme(strip.background = element_rect(fill = "gray95"),
                            panel.spacing.x = unit(0.6, "lines"),
                            axis.text = element_text(size = 6.75))
```

### Swiss

```{r}
df_plot_gb <- df_plot %>%  filter(machine == "Gradient boosting" & study == "Swiss")
ggplot(df_plot_gb, aes(x = value, y = CAD)) + 
        geom_abline(slope = 1, 
                    intercept = 0, 
                    lty = 1, 
                    colour = "black") +
        geom_smooth(method = "gam", span = 0.75,
                    lty = 2, colour = "black") +
       facet_wrap(~ method, ncol = 4, nrow = 2) +
        coord_fixed(ratio = 1) +
        scale_x_continuous(name = "Predicted probability", 
                           limits = c(0 - epsilon, 1 + epsilon)) + 
        scale_y_continuous(name = "True probability", 
                           limits = c(0 - epsilon, 1 + epsilon)) + 
        theme_bw() +  theme(strip.background = element_rect(fill = "gray95"),
                            panel.spacing.x = unit(0.6, "lines"),
                            axis.text = element_text(size = 6.75))
```

### VA

```{r}
df_plot_gb <- df_plot %>%  filter(machine == "Random forest" & study == "VA")
ggplot(df_plot_gb, aes(x = value, y = CAD)) + 
        geom_abline(slope = 1, 
                    intercept = 0, 
                    lty = 1, 
                    colour = "black") +
        geom_smooth(method = "gam", span = 0.75,
                    lty = 2, colour = "black") +
       facet_wrap(~ method, ncol = 4, nrow = 2) +
        coord_fixed(ratio = 1) +
        scale_x_continuous(name = "Predicted probability", 
                           limits = c(0 - epsilon, 1 + epsilon)) + 
        scale_y_continuous(name = "True probability", 
                           limits = c(0 - epsilon, 1 + epsilon)) + 
        theme_bw() +  theme(strip.background = element_rect(fill = "gray95"),
                            panel.spacing.x = unit(0.6, "lines"),
                            axis.text = element_text(size = 6.75))
```


```{r boxplot-function}
# Function necessary for logloss boxplot

bootstrap_LogLoss_plot <- function(boot_data, repetitions = 100) {
  
NoCal_RMSE <- matrix(data=NA,nrow=100,ncol=3); NoCal_LogLoss <- matrix(data=NA,nrow=100,ncol=3); NoCal_BS <- matrix(data=NA,nrow=100,ncol=3)
logreg_RMSE <- matrix(data=NA,nrow=100,ncol=3); logreg_LogLoss <- matrix(data=NA,nrow=100,ncol=3); logreg_BS <- matrix(data=NA,nrow=100,ncol=3)
refit_RMSE <- matrix(data=NA,nrow=100,ncol=3); refit_LogLoss <- matrix(data=NA,nrow=100,ncol=3); refit_BS <- matrix(data=NA,nrow=100,ncol=3)
elkan_RMSE <- matrix(data=NA,nrow=100,ncol=3); elkan_LogLoss <- matrix(data=NA,nrow=100,ncol=3); elkan_BS <- matrix(data=NA,nrow=100,ncol=3)
SimCal_RMSE <- matrix(data=NA,nrow=100,ncol=3); SimCal_LogLoss <- matrix(data=NA,nrow=100,ncol=3); SimCal_BS <- matrix(data=NA,nrow=100,ncol=3)
SBA_RMSE <- matrix(data=NA,nrow=100,ncol=3); SBA_LogLoss <- matrix(data=NA,nrow=100,ncol=3); SBA_BS <- matrix(data=NA,nrow=100,ncol=3)
ACP_RMSE <- matrix(data=NA,nrow=100,ncol=1); ACP_LogLoss <- matrix(data=NA,nrow=100,ncol=1); ACP_BS <- matrix(data=NA,nrow=100,ncol=1)

set.seed(43)

bootstrap_res <- balanced_bootstrap(boot_data, R = repetitions, stratify = "outcome")
machines_used <- c("lr", "rf", "gb")
k <- 0
for (machine in machines_used) {
  k <- k + 1
  for (i in 1:repetitions) {
  
  idx <- bootstrap_res[[1]][[i]][["idx"]]
  in_bag_ids <- unique(idx)
  
  #No Calibration
  NoCal_RMSE[i,k] <- sqrt(mean((boot_data$outcome[-in_bag_ids] - boot_data[[paste0("prob_", machine)]][-in_bag_ids])^2, 
                             na.rm=TRUE))
  NoCal_BS[i,k] <- mean((boot_data$outcome[-in_bag_ids] - boot_data[[paste0("prob_", machine)]][-in_bag_ids])^2, 
                             na.rm=TRUE)
  NoCal_LogLoss[i,k] <-  (-1) * mean(boot_data$outcome[-in_bag_ids] * log(boot_data[[paste0("prob_", machine)]][-in_bag_ids]) + 
                                     (1 - boot_data$outcome[-in_bag_ids]) * 
                                     log(1 - boot_data[[paste0("prob_", machine)]][-in_bag_ids]), na.rm = TRUE)
  
  # Logistic Regression
  logreg_RMSE[i,k] <- sqrt(mean((boot_data$outcome[-in_bag_ids] - boot_data[[paste0("prob_", machine,"_log")]][-in_bag_ids])^2, 
                             na.rm=TRUE))
  logreg_BS[i,k] <- mean((boot_data$outcome[-in_bag_ids] - boot_data[[paste0("prob_", machine,"_log")]][-in_bag_ids])^2, 
                             na.rm=TRUE)
  logreg_LogLoss[i,k] <-  (-1) * mean(boot_data$outcome[-in_bag_ids] * log(boot_data[[paste0("prob_", machine,"_log")]][-in_bag_ids]) + 
                                     (1 - boot_data$outcome[-in_bag_ids]) * 
                                     log(1 - boot_data[[paste0("prob_", machine,"_log")]][-in_bag_ids]),   na.rm = TRUE)
  
    # Refit
  refit_RMSE[i,k] <- sqrt(mean((boot_data$outcome[-in_bag_ids] - boot_data[[paste0("prob_", machine,"_refit")]][-in_bag_ids])^2, 
                             na.rm=TRUE))
  refit_BS[i,k] <- mean((boot_data$outcome[-in_bag_ids] - boot_data[[paste0("prob_", machine,"_refit")]][-in_bag_ids])^2, 
                             na.rm=TRUE)
  refit_LogLoss[i,k] <-  (-1) * mean(boot_data$outcome[-in_bag_ids] * log(boot_data[[paste0("prob_", machine,"_refit")]][-in_bag_ids]) + 
                                     (1 - boot_data$outcome[-in_bag_ids]) * 
                                     log(1 - boot_data[[paste0("prob_", machine,"_refit")]][-in_bag_ids]),   na.rm = TRUE)

  # Elkan
  elkan_RMSE[i,k] <- sqrt(mean((boot_data$outcome[-in_bag_ids] - boot_data[[paste0("prob_", machine,"_elkan")]][-in_bag_ids])^2, 
                             na.rm=TRUE))
  elkan_BS[i,k] <- mean((boot_data$outcome[-in_bag_ids] - boot_data[[paste0("prob_", machine,"_elkan")]][-in_bag_ids])^2, 
                             na.rm=TRUE)
  #propcal <- winsorize_probs(propcal)
  elkan_LogLoss[i,k] <-  (-1) * mean(boot_data$outcome[-in_bag_ids] * log(boot_data[[paste0("prob_", machine,"_elkan")]][-in_bag_ids]) + 
                                     (1 - boot_data$outcome[-in_bag_ids]) * 
                                     log(1 - boot_data[[paste0("prob_", machine,"_elkan")]][-in_bag_ids]),   na.rm = TRUE)

  
  # SimCal
  SimCal_RMSE[i,k] <- sqrt(mean((boot_data$outcome[-in_bag_ids] - boot_data[[paste0("prob_", machine,"_recal")]][-in_bag_ids])^2, 
                             na.rm=TRUE))
  SimCal_BS[i,k] <- mean((boot_data$outcome[-in_bag_ids] - boot_data[[paste0("prob_", machine,"_recal")]][-in_bag_ids])^2, 
                             na.rm=TRUE)
  #propcal <- winsorize_probs(propcal)
  SimCal_LogLoss[i,k] <-  (-1) * mean(boot_data$outcome[-in_bag_ids] * log(boot_data[[paste0("prob_", machine,"_recal")]][-in_bag_ids]) + 
                                     (1 - boot_data$outcome[-in_bag_ids]) * 
                                     log(1 - boot_data[[paste0("prob_", machine,"_recal")]][-in_bag_ids]),   na.rm = TRUE)
  
    # SBA
  SBA_RMSE[i,k] <- sqrt(mean((boot_data$outcome[-in_bag_ids] - boot_data[[paste0("prob_", machine,"_sba")]][-in_bag_ids])^2, 
                             na.rm=TRUE))
  SBA_BS[i,k] <- mean((boot_data$outcome[-in_bag_ids] - boot_data[[paste0("prob_", machine,"_sba")]][-in_bag_ids])^2, 
                             na.rm=TRUE)
  SBA_LogLoss[i,k] <-  (-1) * mean(boot_data$outcome[-in_bag_ids] * log(boot_data[[paste0("prob_", machine,"_sba")]][-in_bag_ids]) + 
                                     (1 - boot_data$outcome[-in_bag_ids]) * 
                                     log(1 - boot_data[[paste0("prob_", machine,"_sba")]][-in_bag_ids]),   na.rm = TRUE)
  
  if (machine == "lr") {
  # ACP
  ACP_RMSE[i,k] <- sqrt(mean((boot_data$outcome[-in_bag_ids] - boot_data[[paste0("prob_", machine,"_acp")]][-in_bag_ids])^2, 
                             na.rm=TRUE))
  ACP_BS[i,k] <- mean((boot_data$outcome[-in_bag_ids] - boot_data[[paste0("prob_", machine,"_acp")]][-in_bag_ids])^2, 
                             na.rm=TRUE)
  ACP_LogLoss[i,k] <-  (-1) * mean(boot_data$outcome[-in_bag_ids] * log(boot_data[[paste0("prob_", machine,"_acp")]][-in_bag_ids]) + 
                                     (1 - boot_data$outcome[-in_bag_ids]) * 
                                     log(1 - boot_data[[paste0("prob_", machine,"_acp")]][-in_bag_ids]),   na.rm = TRUE)
  }
  
}
}

NoCal_LogLoss <- as.data.frame(NoCal_LogLoss)
names(NoCal_LogLoss) <- c("Logistic Regression", "Random Forest", "Gradient Boosting")
NoCal_LogLoss$Method <- "NoCal"

logreg_LogLoss <- as.data.frame(logreg_LogLoss)
names(logreg_LogLoss) <- c("Logistic Regression", "Random Forest", "Gradient Boosting")
logreg_LogLoss$Method <- "Logistic"

refit_LogLoss <- as.data.frame(refit_LogLoss)
names(refit_LogLoss) <- c("Logistic Regression", "Random Forest", "Gradient Boosting")
refit_LogLoss$Method <- "Refit"

elkan_LogLoss <- as.data.frame(elkan_LogLoss)
names(elkan_LogLoss) <- c("Logistic Regression", "Random Forest", "Gradient Boosting")
elkan_LogLoss$Method <- "Elkan"

SimCal_LogLoss <- as.data.frame(SimCal_LogLoss)
names(SimCal_LogLoss) <- c("Logistic Regression", "Random Forest", "Gradient Boosting")
SimCal_LogLoss$Method <- "SimCal"

SBA_LogLoss <- as.data.frame(SBA_LogLoss)
names(SBA_LogLoss) <- c("Logistic Regression", "Random Forest", "Gradient Boosting")
SBA_LogLoss$Method <- "SBA-10"

ACP_LogLoss <- as.data.frame(ACP_LogLoss)
names(ACP_LogLoss) <- c("Logistic Regression")
ACP_LogLoss$Method <- "ACP"
ACP_LogLoss[["Random Forest"]] <- NA
ACP_LogLoss[["Gradient Boosting"]] <- NA

test <- rbind(NoCal_LogLoss, logreg_LogLoss, refit_LogLoss,  SBA_LogLoss,  elkan_LogLoss, SimCal_LogLoss, ACP_LogLoss)

df_long <- test %>%
  pivot_longer(cols = c("Logistic Regression", "Random Forest", "Gradient Boosting"),
               names_to = "Model",
               values_to = "LogLoss")
df_long$Model <- factor(df_long$Model, levels = c(
  "Logistic Regression",
  "Random Forest",
  "Gradient Boosting"
))

df_long$Method <- factor(df_long$Method, levels = rev(c(
  "NoCal", "Logistic", "Refit", "SBA-10", "Elkan", "SimCal","ACP"
)))

# Plot
p <- ggplot(df_long, aes(x = Method, y = LogLoss)) +
  geom_boxplot(outlier.size = 0.5) +
  facet_wrap(vars(Model)) +
  xlab("") + ylab("LogLoss") +
  coord_flip() +
  theme_bw() +
  theme(
    strip.background = element_rect(fill = "gray95"),
    panel.spacing.x = unit(0.75, "lines")
  )  +
  theme(
    text = element_text(size = 12),         # all text
    axis.title = element_text(size = 12),
    axis.text = element_text(size = 12),
    legend.text = element_text(size = 12),
    legend.title = element_text(size = 12)
  )


print(p)

}
```


# Critical difference plots (single machines)


```{r cd-functions}
# Functions necessary for critical difference plots

iman_davenport_test <- function(input_matrix, p.value = 0.05, 
                                small_better = TRUE) {
  
  rankMatrix <- function(data, small_better) {
    if (!small_better){
      f <- function(x){
        rank(-x, ties.method="average")
      }
    } else {
      f <- function(x){
        rank(x, ties.method="average")
      }
    }
    rankings <- t(apply(data, MARGIN = 1, FUN = f))
    colnames(rankings) <- colnames(data)
    rownames(rankings) <- rownames(data)
    rankings
  }
  
  nts <- nrow(input_matrix)
  nws <- ncol(input_matrix)
  mr <- colMeans(rankMatrix(input_matrix, small_better = small_better))
  
  chi <- 12 * nts/(nws * (nws + 1)) * (sum(mr^2) - (nws * (nws + 1)^2)/4)
  FF <- (nts - 1) * chi/(nts * (nws - 1) - chi)
  critVal <- df(1 - p.value, nws - 1, (nws - 1) * (nts - 1))
  rejNull <- FF > critVal
  pval_id <- 1 - pf(FF, df1 = nws - 1, df2 = (nws - 1) * (nts - 1))
  
  output <- list(F.test = list(chi = chi, FF = FF, 
                               critVal = critVal, rejNull = rejNull, 
                               pval_iman_davenport = pval_id),
                 Nemenyi.test = NULL,
                 avgRksWFs = mr
  )
  
  if (rejNull) {
    CD.n <- qtukey(1 - p.value, nws, df = Inf)/sqrt(2) * 
      sqrt(nws * (nws + 1)/(6 * nts))
    allRkDifs <- outer(mr, mr, function(x, y) abs(x - y))
    signifDifs <- allRkDifs >= CD.n
    output[["Nemenyi.test"]] <- list(critDif = CD.n, rkDifs = allRkDifs, 
                                     signifDifs = signifDifs)
  }
  
  list(logloss = output)
}


CD_diagram_Nemenyi <- function(r, metric = names(r)[1], coord_fixed_ratio = 0.5, 
                               bw = FALSE, x_axis_1_right = TRUE,
                               size_cd_text = 3,
                               size_cd_segment = 1.5, 
                               size_segments_joining_methods = 1.2, 
                               show_cd_segment_val = FALSE, dist_cd_segment = 1,
                               base_size = 11, 
                               size_methods_text = 4,
                               axis_x_text_face = "plain",
                               axis_x_text_size = 12) {
  
  if (is.null(r[[metric]]$F.test) | 
      is.null(r[[metric]]$Nemenyi.test)) stop(paste0("Results of both the F ",
                                                     "and Nemenyi tests are ",
                                                     "required for these ",
                                                     "diagrams."))
  
  o <- rank(r[[metric]]$avgRksWFs, ties.method = "first")
  mxl <- ceiling(length(r[[metric]]$avgRksWFs)/2)
  data <- data.frame(avgRk = r[[metric]]$avgRksWFs, 
                     invRk = length(r[[metric]]$avgRksWFs) + 
                       1 - r[[metric]]$avgRksWFs, 
                     sys = names(r[[metric]]$avgRksWFs), 
                     line = o%%mxl + ifelse(o%%mxl == 0, mxl, 0), 
                     side = ifelse(o <= mxl, -1, 1))
  data$line <- ifelse(data$side == 1, mxl + 1 - data$line, 
                      data$line)
  len <- length(r[[metric]]$avgRksWFs)
  cd <- r[[metric]]$Nemenyi.test$critDif
  
  col_arg <- "sys"; if(bw){col_arg <- NULL}
  
  g <- ggplot(data, aes_string(x = "invRk", y = "line")) + 
    geom_segment(aes_string(x = "invRk", y = 0, xend = "invRk", yend = "line", 
                            col = col_arg)) + 
    geom_text(data = data[data$side == -1, ], 
              aes_string(label = "sys", x = +Inf, y = "line", col = col_arg), 
              hjust = 0, size = size_methods_text) + 
    geom_text(data = data[data$side == 1, ], 
              aes_string(label = "sys", x = -Inf, y = "line", col = col_arg), 
              hjust = 1, 
              size = size_methods_text) + 
    geom_segment(data = data[data$side == -1, ], 
                 aes_string(x = max(o) + 0.1, y = "line", xend = "invRk", 
                            yend = "line", col = col_arg)) + 
    geom_segment(data = data[data$side == 1, ], 
                 aes_string(x = "invRk", y = "line", 
                            xend = 1 - 0.1, yend = "line", col = col_arg)) 
  
  labels_scale_x <- paste(1:len)
  if(x_axis_1_right) {
    labels_scale_x <- paste(len:1)
  }
  g <- g +  scale_x_continuous(limits = c(1 - 0.15, len + 0.15), 
                               breaks = 1:len, 
                               labels = labels_scale_x) 
  
  g <-  g +  scale_y_continuous(limits = c(0, mxl + 1)) + 
    xlab("Average Rank") + ylab("") + theme_grey(base_size = base_size) +
    theme(axis.text.y = element_blank(), 
          axis.ticks.y = element_blank(), axis.title.y = element_blank(),
          axis.text.x = element_text(size = axis_x_text_size, 
                                     face = axis_x_text_face),
          #axis.title.x = element_text(size = 5),
          legend.position = "none", panel.background = element_blank(), 
          panel.border = element_blank(), 
          axis.line = element_line(size = 1), 
          axis.line.y = element_blank(), panel.grid.major = element_blank(), 
          plot.background = element_blank(), 
          plot.margin = grid::unit(c(3, 6, 1, 5), "lines")) + 
    coord_fixed(ratio = coord_fixed_ratio, clip = "off")
  
  if(show_cd_segment_val) {
    g <- g + annotate("segment", x = 1, xend = cd + 1, 
                      y = mxl + dist_cd_segment, 
                      yend = mxl + dist_cd_segment, 
                      size = size_cd_segment) + 
      annotate("text", x = 1, y = mxl + dist_cd_segment, 
               label = paste("Critical Difference", round(cd, 1), sep = " = "), 
               vjust = -0.5, hjust = 0, size = size_cd_text)
  }  
  wfsOrd <- names(r[[metric]]$avgRksWFs[order(r[[metric]]$avgRksWFs)])
  ss <- r[[metric]]$Nemenyi.test$signifDifs[wfsOrd, wfsOrd]
  mx <- ncol(ss)
  pos <- rep(1:mxl, each = mxl) - seq(0, 1, by = 1/(mxl + 1))[-c(1, mxl + 2)]
  frees <- rep(1, mxl)
  from <- 1
  currTill <- 0
  for (i in 1:nrow(ss)) {
    
    # add i-1 to which(ss[i,i:mx]) to get the correct which-values (since the 
    # column selection removes i-1 values)
    till <- which(ss[i,i:mx]) + (i-1) 
    till <- if (length(till)>0) till[1]-1 else mx
    if (till > i && till > currTill) {
      theLine <- min(data[wfsOrd[i], "line"], data[wfsOrd[till], 
                                                   "line"])
      ypos <- pos[(theLine - 1) * mxl + frees[theLine]]
      frees[theLine] <- frees[theLine] + 1
      g <- g + annotate("segment", x = data[wfsOrd[till], "invRk"] - 0.1, 
                        xend = data[wfsOrd[i], "invRk"] + 0.1, y = ypos, 
                        yend = ypos, size = size_segments_joining_methods)
      currTill <- till
    }
    if (till == mx) break
  }
  
  g
}


bootstrap_CD_plot <- function(boot_data, repetitions = 100,  machine = c("lr", "rf", "gb"),
                              multiplot = FALSE) {
  
NoCal_RMSE <- rep(NA,100); NoCal_LogLoss <- rep(NA,100); NoCal_BS <- rep(NA,100)
refit_RMSE <- rep(NA,100); refit_LogLoss <- rep(NA,100); refit_BS <- rep(NA,100)
logreg_RMSE <- rep(NA,100); logreg_LogLoss <- rep(NA,100); logreg_BS <- rep(NA,100)
elkan_RMSE <- rep(NA,100); elkan_LogLoss <- rep(NA,100); elkan_BS <- rep(NA,100)
SimCal_RMSE <- rep(NA,100); SimCal_LogLoss <- rep(NA,100); SimCal_BS <- rep(NA,100)
SBA_RMSE <- rep(NA,100); SBA_LogLoss <- rep(NA,100); SBA_BS <- rep(NA,100)
ACP_RMSE <- rep(NA,100); ACP_LogLoss <- rep(NA,100); ACP_BS <- rep(NA,100)

set.seed(43)

bootstrap_res <- balanced_bootstrap(boot_data, R = repetitions, stratify = "outcome")

for (i in 1:repetitions) {
  
  idx <- bootstrap_res[[1]][[i]][["idx"]]
  in_bag_ids <- unique(idx)
  
  #No Calibration
  NoCal_RMSE[i] <- sqrt(mean((boot_data$outcome[-in_bag_ids] - boot_data[[paste0("prob_", machine)]][-in_bag_ids])^2, 
                             na.rm=TRUE))
  NoCal_BS[i] <- mean((boot_data$outcome[-in_bag_ids] - boot_data[[paste0("prob_", machine)]][-in_bag_ids])^2, 
                             na.rm=TRUE)
  NoCal_LogLoss[i] <-  (-1) * mean(boot_data$outcome[-in_bag_ids] * log(boot_data[[paste0("prob_", machine)]][-in_bag_ids]) + 
                                     (1 - boot_data$outcome[-in_bag_ids]) * 
                                     log(1 - boot_data[[paste0("prob_", machine)]][-in_bag_ids]), na.rm = TRUE)
  
  # Logistic Regression
  logreg_RMSE[i] <- sqrt(mean((boot_data$outcome[-in_bag_ids] - boot_data[[paste0("prob_", machine,"_log")]][-in_bag_ids])^2, 
                             na.rm=TRUE))
  logreg_BS[i] <- mean((boot_data$outcome[-in_bag_ids] - boot_data[[paste0("prob_", machine,"_log")]][-in_bag_ids])^2, 
                             na.rm=TRUE)
  logreg_LogLoss[i] <-  (-1) * mean(boot_data$outcome[-in_bag_ids] * log(boot_data[[paste0("prob_", machine,"_log")]][-in_bag_ids]) + 
                                     (1 - boot_data$outcome[-in_bag_ids]) * 
                                     log(1 - boot_data[[paste0("prob_", machine,"_log")]][-in_bag_ids]),   na.rm = TRUE)
  
    # Refit
  refit_RMSE[i] <- sqrt(mean((boot_data$outcome[-in_bag_ids] - boot_data[[paste0("prob_", machine,"_refit")]][-in_bag_ids])^2, 
                             na.rm=TRUE))
  refit_BS[i] <- mean((boot_data$outcome[-in_bag_ids] - boot_data[[paste0("prob_", machine,"_refit")]][-in_bag_ids])^2, 
                             na.rm=TRUE)
  refit_LogLoss[i] <-  (-1) * mean(boot_data$outcome[-in_bag_ids] * log(boot_data[[paste0("prob_", machine,"_refit")]][-in_bag_ids]) + 
                                     (1 - boot_data$outcome[-in_bag_ids]) * 
                                     log(1 - boot_data[[paste0("prob_", machine,"_refit")]][-in_bag_ids]),   na.rm = TRUE)

  
  # Elkan 
  elkan_RMSE[i] <- sqrt(mean((boot_data$outcome[-in_bag_ids] - boot_data[[paste0("prob_", machine,"_elkan")]][-in_bag_ids])^2, 
                             na.rm=TRUE))
  elkan_BS[i] <- mean((boot_data$outcome[-in_bag_ids] - boot_data[[paste0("prob_", machine,"_elkan")]][-in_bag_ids])^2, 
                             na.rm=TRUE)
  elkan_LogLoss[i] <-  (-1) * mean(boot_data$outcome[-in_bag_ids] * log(boot_data[[paste0("prob_", machine,"_elkan")]][-in_bag_ids]) + 
                                     (1 - boot_data$outcome[-in_bag_ids]) * 
                                     log(1 - boot_data[[paste0("prob_", machine,"_elkan")]][-in_bag_ids]),   na.rm = TRUE)

  
  # SimCal
  SimCal_RMSE[i] <- sqrt(mean((boot_data$outcome[-in_bag_ids] - boot_data[[paste0("prob_", machine,"_recal")]][-in_bag_ids])^2, 
                             na.rm=TRUE))
  SimCal_BS[i] <- mean((boot_data$outcome[-in_bag_ids] - boot_data[[paste0("prob_", machine,"_recal")]][-in_bag_ids])^2, 
                             na.rm=TRUE)
  SimCal_LogLoss[i] <-  (-1) * mean(boot_data$outcome[-in_bag_ids] * log(boot_data[[paste0("prob_", machine,"_recal")]][-in_bag_ids]) + 
                                     (1 - boot_data$outcome[-in_bag_ids]) * 
                                     log(1 - boot_data[[paste0("prob_", machine,"_recal")]][-in_bag_ids]),   na.rm = TRUE)
  
    # SBA
  SBA_RMSE[i] <- sqrt(mean((boot_data$outcome[-in_bag_ids] - boot_data[[paste0("prob_", machine,"_sba")]][-in_bag_ids])^2, 
                             na.rm=TRUE))
  SBA_BS[i] <- mean((boot_data$outcome[-in_bag_ids] - boot_data[[paste0("prob_", machine,"_sba")]][-in_bag_ids])^2, 
                             na.rm=TRUE)
  SBA_LogLoss[i] <-  (-1) * mean(boot_data$outcome[-in_bag_ids] * log(boot_data[[paste0("prob_", machine,"_sba")]][-in_bag_ids]) + 
                                     (1 - boot_data$outcome[-in_bag_ids]) * 
                                     log(1 - boot_data[[paste0("prob_", machine,"_sba")]][-in_bag_ids]),   na.rm = TRUE)
                                                                                                                                                                        
  if (machine == "lr") {
  # ACP
  ACP_RMSE[i] <- sqrt(mean((boot_data$outcome[-in_bag_ids] - boot_data[[paste0("prob_", machine,"_acp")]][-in_bag_ids])^2, 
                             na.rm=TRUE))
  ACP_BS[i] <- mean((boot_data$outcome[-in_bag_ids] - boot_data[[paste0("prob_", machine,"_acp")]][-in_bag_ids])^2, 
                             na.rm=TRUE)
  ACP_LogLoss[i] <-  (-1) * mean(boot_data$outcome[-in_bag_ids] * log(boot_data[[paste0("prob_", machine,"_acp")]][-in_bag_ids]) + 
                                     (1 - boot_data$outcome[-in_bag_ids]) * 
                                     log(1 - boot_data[[paste0("prob_", machine,"_acp")]][-in_bag_ids]),   na.rm = TRUE)
  }

}

if (machine == "lr") {
  test <- cbind(NoCal_LogLoss, logreg_LogLoss, refit_LogLoss, elkan_LogLoss, SimCal_LogLoss, SBA_LogLoss, ACP_LogLoss)
  colnames(test) <- c("NoCal", "Logistic", "Refit",  "Elkan", "SimCal", "SBA-10", "ACP")
} else {
  test <- cbind(NoCal_LogLoss, logreg_LogLoss, refit_LogLoss, elkan_LogLoss, SimCal_LogLoss, SBA_LogLoss)
  colnames(test) <- c("NoCal", "Logistic", "Refit",  "Elkan", "SimCal", "SBA-10")
}

dsub_wide_test <- as.matrix(test) 
id_test_check <- iman_davenport_test(dsub_wide_test, small_better = FALSE)
pval_iman_dvt <- id_test_check[["logloss"]][["F.test"]][["pval_iman_davenport"]]

if (multiplot) {
  p <- CD_diagram_Nemenyi(id_test_check, coord_fixed_ratio = 0.3, bw = TRUE, 
                            x_axis_1_right = FALSE, size_cd_segment = 0.7, 
                            size_segments_joining_methods = 0.6, 
                            show_cd_segment_val = FALSE,
                            axis_x_text_face = "bold",
                            axis_x_text_size = 6.5)
} else {
  p <- CD_diagram_Nemenyi(id_test_check, coord_fixed_ratio = 0.8, bw = TRUE,
                            x_axis_1_right = FALSE)
}
print(p)

}

ci_fun <- function(x){
  margin_e <- qt(0.975, df = length(x) - 1)*sd(x)/sqrt(length(x))
  c(mean(x) - margin_e, mean(x) + margin_e)
}

bootstrap_average_CD_plot <- function(boot_data, repetitions = 100, multiplot = FALSE) {
  
NoCal_RMSE <- matrix(data=NA,nrow=100,ncol=3); NoCal_LogLoss <- matrix(data=NA,nrow=100,ncol=3); NoCal_BS <- matrix(data=NA,nrow=100,ncol=3)
logreg_RMSE <- matrix(data=NA,nrow=100,ncol=3); logreg_LogLoss <- matrix(data=NA,nrow=100,ncol=3); logreg_BS <- matrix(data=NA,nrow=100,ncol=3)
refit_RMSE <- matrix(data=NA,nrow=100,ncol=3); refit_LogLoss <- matrix(data=NA,nrow=100,ncol=3); refit_BS <- matrix(data=NA,nrow=100,ncol=3)
elkan_RMSE <- matrix(data=NA,nrow=100,ncol=3); elkan_LogLoss <- matrix(data=NA,nrow=100,ncol=3); elkan_BS <- matrix(data=NA,nrow=100,ncol=3)
SimCal_RMSE <- matrix(data=NA,nrow=100,ncol=3); SimCal_LogLoss <- matrix(data=NA,nrow=100,ncol=3); SimCal_BS <- matrix(data=NA,nrow=100,ncol=3)
SBA_RMSE <- matrix(data=NA,nrow=100,ncol=3); SBA_LogLoss <- matrix(data=NA,nrow=100,ncol=3); SBA_BS <- matrix(data=NA,nrow=100,ncol=3)

set.seed(43)

bootstrap_res <- balanced_bootstrap(boot_data, R = repetitions, stratify = "outcome")
machines_used <- c("lr", "rf", "gb")
for (k in 1:3) {
  
  for (i in 1:repetitions) {
  
  idx <- bootstrap_res[[1]][[i]][["idx"]]
  in_bag_ids <- unique(idx)
  
  #No Calibration
  NoCal_RMSE[i,k] <- sqrt(mean((boot_data$outcome[-in_bag_ids] - boot_data[[paste0("prob_", machines_used[k])]][-in_bag_ids])^2, 
                             na.rm=TRUE))
  NoCal_BS[i,k] <- mean((boot_data$outcome[-in_bag_ids] - boot_data[[paste0("prob_", machines_used[k])]][-in_bag_ids])^2, 
                             na.rm=TRUE)
  NoCal_LogLoss[i,k] <-  (-1) * mean(boot_data$outcome[-in_bag_ids] * log(boot_data[[paste0("prob_", machines_used[k])]][-in_bag_ids]) + 
                                     (1 - boot_data$outcome[-in_bag_ids]) * 
                                     log(1 - boot_data[[paste0("prob_", machines_used[k])]][-in_bag_ids]), na.rm = TRUE)
  
  
  # Logistic Regression
  logreg_RMSE[i,k] <- sqrt(mean((boot_data$outcome[-in_bag_ids] - boot_data[[paste0("prob_", machines_used[k],"_log")]][-in_bag_ids])^2, 
                             na.rm=TRUE))
  logreg_BS[i,k] <- mean((boot_data$outcome[-in_bag_ids] - boot_data[[paste0("prob_", machines_used[k],"_log")]][-in_bag_ids])^2, 
                             na.rm=TRUE)
  logreg_LogLoss[i,k] <-  (-1) * mean(boot_data$outcome[-in_bag_ids] * log(boot_data[[paste0("prob_", machines_used[k],"_log")]][-in_bag_ids]) + 
                                     (1 - boot_data$outcome[-in_bag_ids]) * 
                                     log(1 - boot_data[[paste0("prob_", machines_used[k],"_log")]][-in_bag_ids]),   na.rm = TRUE)

  
  # Elkan
  elkan_RMSE[i,k] <- sqrt(mean((boot_data$outcome[-in_bag_ids] - boot_data[[paste0("prob_", machines_used[k],"_elkan")]][-in_bag_ids])^2, 
                             na.rm=TRUE))
  elkan_BS[i,k] <- mean((boot_data$outcome[-in_bag_ids] - boot_data[[paste0("prob_", machines_used[k],"_elkan")]][-in_bag_ids])^2, 
                             na.rm=TRUE)
  elkan_LogLoss[i,k] <-  (-1) * mean(boot_data$outcome[-in_bag_ids] * log(boot_data[[paste0("prob_", machines_used[k],"_elkan")]][-in_bag_ids]) + 
                                     (1 - boot_data$outcome[-in_bag_ids]) * 
                                     log(1 - boot_data[[paste0("prob_", machines_used[k],"_elkan")]][-in_bag_ids]),   na.rm = TRUE)

  
  # SimCal
  SimCal_RMSE[i,k] <- sqrt(mean((boot_data$outcome[-in_bag_ids] - boot_data[[paste0("prob_", machines_used[k],"_recal")]][-in_bag_ids])^2, 
                             na.rm=TRUE))
  SimCal_BS[i,k] <- mean((boot_data$outcome[-in_bag_ids] - boot_data[[paste0("prob_", machines_used[k],"_recal")]][-in_bag_ids])^2, 
                             na.rm=TRUE)
  SimCal_LogLoss[i,k] <-  (-1) * mean(boot_data$outcome[-in_bag_ids] * log(boot_data[[paste0("prob_", machines_used[k],"_recal")]][-in_bag_ids]) + 
                                     (1 - boot_data$outcome[-in_bag_ids]) * 
                                     log(1 - boot_data[[paste0("prob_", machines_used[k],"_recal")]][-in_bag_ids]),   na.rm = TRUE)
  
    # SBA
  SBA_RMSE[i,k] <- sqrt(mean((boot_data$outcome[-in_bag_ids] - boot_data[[paste0("prob_", machines_used[k],"_sba")]][-in_bag_ids])^2, 
                             na.rm=TRUE))
  SBA_BS[i,k] <- mean((boot_data$outcome[-in_bag_ids] - boot_data[[paste0("prob_", machines_used[k],"_sba")]][-in_bag_ids])^2, 
                             na.rm=TRUE)
  SBA_LogLoss[i,k] <-  (-1) * mean(boot_data$outcome[-in_bag_ids] * log(boot_data[[paste0("prob_", machines_used[k],"_sba")]][-in_bag_ids]) + 
                                     (1 - boot_data$outcome[-in_bag_ids]) * 
                                     log(1 - boot_data[[paste0("prob_", machines_used[k],"_sba")]][-in_bag_ids]),   na.rm = TRUE)
  
  
}
  
}

NoCal_LogLoss <- as.data.frame(NoCal_LogLoss) %>% mutate( AverageLogLoss = rowMeans(.))
logreg_LogLoss <- as.data.frame(logreg_LogLoss) %>% mutate( AverageLogLoss = rowMeans(.))
refit_LogLoss <- as.data.frame(refit_LogLoss) %>% mutate( AverageLogLoss = rowMeans(.))
elkan_LogLoss <- as.data.frame(elkan_LogLoss) %>% mutate( AverageLogLoss = rowMeans(.))
SimCal_LogLoss <- as.data.frame(SimCal_LogLoss) %>% mutate( AverageLogLoss = rowMeans(.))
SBA_LogLoss <- as.data.frame(SBA_LogLoss) %>% mutate( AverageLogLoss = rowMeans(.))

test <- cbind(NoCal_LogLoss[4], logreg_LogLoss[4], refit_LogLoss[4], elkan_LogLoss[4], SimCal_LogLoss[4], SBA_LogLoss[4])
colnames(test) <- c("NoCal", "Logistic", "Refit", "Elkan", "SimCal", "SBA-10")


dsub_wide_test <- as.matrix(test) 
id_test_check <- iman_davenport_test(dsub_wide_test, small_better = FALSE)
pval_iman_dvt <- id_test_check[["logloss"]][["F.test"]][["pval_iman_davenport"]]
if (multiplot) {
  p <- CD_diagram_Nemenyi(id_test_check, coord_fixed_ratio = 0.3, bw = TRUE, 
                            x_axis_1_right = FALSE, size_cd_segment = 0.7, 
                            size_segments_joining_methods = 0.6, 
                            show_cd_segment_val = FALSE,
                            axis_x_text_face = "bold",
                            axis_x_text_size = 6.5)
} else {
  p <- CD_diagram_Nemenyi(id_test_check, coord_fixed_ratio = 0.8, bw = TRUE,
                            x_axis_1_right = FALSE)
}
print(p)

}

```

## Logistic regression

```{r}
boot_data <- df_res %>% rename(outcome = CAD)
p1 <- bootstrap_CD_plot(boot_data, repetitions = 100,  machine = c("lr"), multiplot = TRUE)
```

## Random forest

```{r}
p2 <- bootstrap_CD_plot(boot_data, repetitions = 100,  machine = c("rf"), multiplot = TRUE)
```

## Gradient boosting

```{r}
p3 <- bootstrap_CD_plot(boot_data, repetitions = 100,  machine = c("gb"), multiplot = TRUE)
```

# Critical difference plots for the LogLoss averaging over machines and performance measure plots

```{r}
p4 <- bootstrap_average_CD_plot(boot_data, repetitions = 100, multiplot = TRUE)
p5 <- bootstrap_LogLoss_plot(boot_data, repetitions = 100)
```
````
