## Supplementary material for "Calibrating machine learning approaches for probability estimation without calibration data": README.txt

Calibration Comparison Code

Supplementary Material for
"Calibrating machine learning approaches for probability estimation in case of the absence of calibration data"
by Eleonora Di Carluccio, Georgios Koliopanos, Francisco M. Ojeda, Christian Weimar and Andreas Ziegler.

A description of the folder structure is shown below:

Supplementary_material
|
|-----Calibration_Sup1
|     |  Supplementary_material_1.pdf :  TRIPOD checklist for stroke data, LogReg regression coefficients
|     |                                  for stroke data and Cleveland Clinic data, graphics for real data
|     |                                  application (stroke and Cleveland Clinic data)
|  
|-----Calibration_Sup2
|     |  01-data-generation-functions.R : list of functions used to generate simulated data
|     |  02-machine-fitting-functions.R : list of functions used to fit machines
|     |  03-calibration-functions.R     : list of functions used in calibration process
|     |  04-rmd-functions.R             : list of functions for presenting simulation results in .Rmd file
|     |  05-fitting-machines-estimating-probs.R : simulates data and fits machines
|     |  06-calibrating-predictions.R : calibrates output data from 05
|
|-----Calibration_Sup3                  : files for simulation study 1
|     |  Supplementary_material_3a.Rmd  : graphics and tables for correctly calibrated data (markdown)
|     |  Supplementary_material_3b.html : graphics and tables for correctly calibrated data (html)
|
|-----Calibration_Sup4                  : files for simulation study 2
|     |  Supplementary_material_4a.Rmd  : graphics and tables for misspecified mean calibrated data (markdown)
|     |  Supplementary_material_4b.html : graphics and tables for misspecified mean calibrated data (html)
|
|-----Calibration_Sup5                  : files for simulation study 3
|     |  Supplementary_material_5a.Rmd  : graphics and tables for misspecified proportion calibrated data (markdown)
|     |  Supplementary_material_5b.html : graphics and tables for misspecified proportion calibrated data (html)
|
|-----Calibration_Sup6                  : files for simulation study 4
|     |  Supplementary_material_6a.Rmd  : graphics and tables for misspecified mean and proportion calibrated data (markdown)
|     |  Supplementary_material_6b.html : graphics and tables for misspecified mean and proportion calibrated data (html)
|
|-----Calibration_Sup7
|     |  Supplementary_material_7.qmd  : generates graphics and tables for real data application (Cleveland Clinic data)
|
|-----rds-files                        : folder where simulated data will get saved when the code is run
|     |  before-recalibration          : simulated data with probabilities before recalibration
|     |  after-recalibration           : simulated data with probabilities after recalibration
|     |  after-recalibration-misspec   : simulated data with probabilities after misspecified recalibration
|
|-----Supplementary_material.Rproj     : R project file
|
|-----README.md                        : the currently open README file.
|
| renv, .Rprofile and renv.lock are all files relative to the "renv" R package, used to create a reproducible R environment.


To reproduce results, please follow these instructions:
- Open the R project file "Supplementary_material.Rproj". This will ensure the correct folder path to
  save and read files.
- Run renv::restore() to install all necessary packages.
- File 05 in the Calibration_Sup2 folder will simulate the data for all simulation scenarios, fit the machines and
  estimate probabilities using those machines. The results will be saved in the folder 'rds-files/before-recalibration'.
- File 06 in the Calibration_Sup2 folder will recalibrate the probabilities estimated from file 05:
1) When setting both the parameters MISSPEC_MEAN and MISSPEC_PROP to FALSE, the correctly recalibrated data will be
   saved in folder 'rds-files/after-recalibration' (Simulation study 1).
2) Setting the parameter MISSPEC_MEAN to TRUE and MISSPEC_PROP to FALSE will generate calibrations with misspecified mean
   values and save the results in the subfolder 'after-recalibration-misspec', all ending with the suffix
   'mean-misspec' (Simulation study 2).
3) Setting the parameter MISSPEC_PROP to TRUE and  MISSPEC_MEAN to FALSE will generate calibrations with misspecified
   proportion values and save the results in the subfolder 'after-recalibration-misspec', all ending with the suffix
   'prop-misspec' (Simulation study 3).
4) Setting both the parameters MISSPEC_PROP and MISSPEC_MEAN to TRUE will generate calibrations with misspecified
   proportion and misspecified mean values and save the results in the subfolder 'after-recalibration-misspec',
   all ending with the suffix 'mean-prop-misspec' (Simulation study 4).
- By default both parameters are set to FALSE.
- 'Knitting' files Supplementary_material_3a.Rmd, Supplementary_material_4a.Rmd,
- Supplementary_material_5a.Rmd and Supplementary_material_6a.Rmd will produce an html file that
  contains figures and tables presenting the simulation results in the case of no misspecification,
  misspecification of the mean, misspecification of the proportion, and misspecification of both.

Please note: computational time for the recalibration of all simulated datasets is quite extensive.
Estimated run times are presented as comments in the parameter selection section of file 06 (lines 70-100).
For this reason, we have uploaded a copy of the rds-files folder containing all the necessary data to Zenodo
(zenodo.org/records/21099064). To access the data correctly, please download the folder, unzip it, and move the
innermost rds-files folder to the Supplementary_materials folder, replacing the empty rds-files folder.
