## Supplementary material for "Calibrating machine learning approaches for probability estimation without calibration data": activate.R

```
local({

  # the requested version of renv
  version <- "1.2.3"
  attr(version, "md5") <- "1bd9f58e1cfe27ce035933937c6f03de"
  attr(version, "sha") <- NULL

  # the project directory
  project <- Sys.getenv("RENV_PROJECT")
  if (!nzchar(project))
    project <- getwd()

  # use start-up diagnostics if enabled
  diagnostics <- Sys.getenv("RENV_STARTUP_DIAGNOSTICS", unset = "FALSE")
  if (diagnostics) {
    start <- Sys.time()
    profile <- tempfile("renv-startup-", fileext = ".Rprof")
    utils::Rprof(profile)
    on.exit({
      utils::Rprof(NULL)
      elapsed <- signif(difftime(Sys.time(), start, units = "auto"), digits = 2L)
      writeLines(sprintf("- renv took %s to run the autoloader.", format(elapsed)))
      writeLines(sprintf("- Profile: %s", profile))
      print(utils::summaryRprof(profile))
    }, add = TRUE)
  }

  # figure out whether the autoloader is enabled
  enabled <- local({

    # first, check config option
    override <- getOption("renv.config.autoloader.enabled")
    if (!is.null(override))
      return(override)

    # if we're being run in a context where R_LIBS is already set,
    # don't load -- presumably we're being run as a sub-process and
    # the parent process has already set up library paths for us
    rcmd <- Sys.getenv("R_CMD", unset = NA)
    rlibs <- Sys.getenv("R_LIBS", unset = NA)
    if (!is.na(rlibs) && !is.na(rcmd))
      return(FALSE)

    # next, check environment variables
    # prefer using the configuration one in the future
    envvars <- c(
      "RENV_CONFIG_AUTOLOADER_ENABLED",
      "RENV_AUTOLOADER_ENABLED",
      "RENV_ACTIVATE_PROJECT"
    )

    for (envvar in envvars) {
      envval <- Sys.getenv(envvar, unset = NA)
      if (!is.na(envval))
        return(tolower(envval) %in% c("true", "t", "1"))
    }

    # enable by default
    TRUE

  })

  # bail if we're not enabled
  if (!enabled) {

    # if we're not enabled, we might still need to manually load
    # the user profile here
    profile <- Sys.getenv("R_PROFILE_USER", unset = "~/.Rprofile")
    if (file.exists(profile)) {
      cfg <- Sys.getenv("RENV_CONFIG_USER_PROFILE", unset = "TRUE")
      if (tolower(cfg) %in% c("true", "t", "1"))
        sys.source(profile, envir = globalenv())
    }

    return(FALSE)

  }

  # avoid recursion
  if (identical(getOption("renv.autoloader.running"), TRUE)) {
    warning("ignoring recursive attempt to run renv autoloader")
    return(invisible(TRUE))
  }

  # signal that we're loading renv during R startup
  options(renv.autoloader.running = TRUE)
  on.exit(options(renv.autoloader.running = NULL), add = TRUE)

  # signal that we've consented to use renv
  options(renv.consent = TRUE)

  # load the 'utils' package eagerly -- this ensures that renv shims, which
  # mask 'utils' packages, will come first on the search path
  library(utils, lib.loc = .Library)

  # unload renv if it's already been loaded
  if ("renv" %in% loadedNamespaces())
    unloadNamespace("renv")

  # load bootstrap tools   
  ansify <- function(text) {
    if (renv_ansify_enabled())
      renv_ansify_enhanced(text)
    else
      renv_ansify_default(text)
  }
  
  renv_ansify_enabled <- function() {
  
    override <- Sys.getenv("RENV_ANSIFY_ENABLED", unset = NA)
    if (!is.na(override))
      return(as.logical(override))
  
    pane <- Sys.getenv("RSTUDIO_CHILD_PROCESS_PANE", unset = NA)
    if (identical(pane, "build"))
      return(FALSE)
  
    testthat <- Sys.getenv("TESTTHAT", unset = "false")
    if (tolower(testthat) %in% "true")
      return(FALSE)
  
    iderun <- Sys.getenv("R_CLI_HAS_HYPERLINK_IDE_RUN", unset = "false")
    if (tolower(iderun) %in% "false")
      return(FALSE)
  
    TRUE
  
  }
  
  renv_ansify_default <- function(text) {
    text
  }
  
  renv_ansify_enhanced <- function(text) {
  
    # R help links
    pattern <- "`\\?(renv::(?:[^`])+)`"
    replacement <- "`\033]8;;x-r-help:\\1\a?\\1\033]8;;\a`"
    text <- gsub(pattern, replacement, text, perl = TRUE)
  
    # runnable code
    pattern <- "`(renv::(?:[^`])+)`"
    replacement <- "`\033]8;;x-r-run:\\1\a\\1\033]8;;\a`"
    text <- gsub(pattern, replacement, text, perl = TRUE)
  
    # return ansified text
    text
  
  }
  
  renv_ansify_init <- function() {
  
    envir <- renv_envir_self()
    if (renv_ansify_enabled())
      assign("ansify", renv_ansify_enhanced, envir = envir)
    else
      assign("ansify", renv_ansify_default, envir = envir)
  
  }
  
  `%||%` <- function(x, y) {
    if (is.null(x)) y else x
  }
  
  catf <- function(fmt, ..., appendLF = TRUE) {
  
    quiet <- getOption("renv.bootstrap.quiet", default = FALSE)
    if (quiet)
      return(invisible())
  
    # also check for config environment variables that should suppress messages
    # https://github.com/rstudio/renv/issues/2214
    enabled <- Sys.getenv("RENV_CONFIG_STARTUP_QUIET", unset = NA)
    if (!is.na(enabled) && tolower(enabled) %in% c("true", "1"))
      return(invisible())
  
    enabled <- Sys.getenv("RENV_CONFIG_SYNCHRONIZED_CHECK", unset = NA)
    if (!is.na(enabled) && tolower(enabled) %in% c("false", "0"))
      return(invisible())
  
    msg <- sprintf(fmt, ...)
    cat(msg, file = stdout(), sep = if (appendLF) "\n" else "")
  
    invisible(msg)
  
  }
  
  header <- function(label,
                     ...,
                     prefix = "#",
                     suffix = "-",
                     n = min(getOption("width"), 78))
  {
    label <- sprintf(label, ...)
    n <- max(n - nchar(label) - nchar(prefix) - 2L, 8L)
    if (n <= 0)
      return(paste(prefix, label))
  
    tail <- paste(rep.int(suffix, n), collapse = "")
    paste0(prefix, " ", label, " ", tail)
  
  }
  
  heredoc <- function(text, leave = 0) {
  
    # remove leading, trailing whitespace
    trimmed <- gsub("^\\s*\\n|\\n\\s*$", "", text)
  
    # split into lines
    lines <- strsplit(trimmed, "\n", fixed = TRUE)[[1L]]
  
    # compute common indent
    indent <- regexpr("[^[:space:]]", lines)
    common <- min(setdiff(indent, -1L)) - leave
    text <- paste(substring(lines, common), collapse = "\n")
  
    # substitute in ANSI links for executable renv code
    ansify(text)
  
  }
  
  bootstrap <- function(version, library) {
  
    friendly <- renv_bootstrap_version_friendly(version)
    section <- header(sprintf("Bootstrapping renv %s", friendly))
    catf(section)
  
    # ensure the target library path exists; required for file.copy(..., recursive = TRUE)
    dir.create(library, showWarnings = FALSE, recursive = TRUE)
  
    # try to install renv from cache
    md5 <- attr(version, "md5", exact = TRUE)
    if (length(md5)) {
      pkgpath <- renv_bootstrap_find(version)
      if (length(pkgpath) && file.exists(pkgpath)) {
        ok <- file.copy(pkgpath, library, recursive = TRUE)
        if (isTRUE(ok))
          return(invisible())
      }
    }
  
    # attempt to download renv
    catf("- Downloading renv ... ", appendLF = FALSE)
    withCallingHandlers(
      tarball <- renv_bootstrap_download(version),
      error = function(err) {
        catf("FAILED")
        stop("failed to download:\n", conditionMessage(err))
      }
    )
    catf("OK")
    on.exit(unlink(tarball), add = TRUE)
  
    # now attempt to install
    catf("- Installing renv  ... ", appendLF = FALSE)
    withCallingHandlers(
      status <- renv_bootstrap_install(version, tarball, library),
      error = function(err) {
        catf("FAILED")
        stop("failed to install:\n", conditionMessage(err))
      }
    )
    catf("OK")
  
    # add empty line to break up bootstrapping from normal output
    catf("")
    return(invisible())
  }
  
  renv_bootstrap_tests_running <- function() {
    getOption("renv.tests.running", default = FALSE)
  }
  
  renv_bootstrap_repos <- function() {
  
    # get CRAN repository
    cran <- getOption("renv.repos.cran", "https://cloud.r-project.org")
  
    # check for repos override
    repos <- Sys.getenv("RENV_CONFIG_REPOS_OVERRIDE", unset = NA)
    if (!is.na(repos)) {
  
      # split on ';' if present
      parts <- strsplit(repos, ";", fixed = TRUE)[[1L]]
  
      # split into named repositories if present
      idx <- regexpr("=", parts, fixed = TRUE)
      keys <- substring(parts, 1L, idx - 1L)
      vals <- substring(parts, idx + 1L)
      names(vals) <- keys
  
      # if we have a single unnamed repository, call it CRAN
      if (length(vals) == 1L && identical(keys, ""))
        names(vals) <- "CRAN"
  
      return(vals)
  
    }
  
    # check for lockfile repositories
    repos <- tryCatch(renv_bootstrap_repos_lockfile(), error = identity)
    if (!inherits(repos, "error") && length(repos))
      return(repos)
  
    # retrieve current repos
    repos <- getOption("repos")
  
    # ensure @CRAN@ entries are resolved
    repos[repos == "@CRAN@"] <- cran
  
    # add in renv.bootstrap.repos if set
    default <- c(FALLBACK = "https://cloud.r-project.org")
    extra <- getOption("renv.bootstrap.repos", default = default)
    repos <- c(repos, extra)
  
    # remove duplicates that might've snuck in
    dupes <- duplicated(repos) | duplicated(names(repos))
    repos[!dupes]
  
  }
  
  renv_bootstrap_repos_lockfile <- function() {
  
    lockpath <- Sys.getenv("RENV_PATHS_LOCKFILE", unset = "renv.lock")
    if (!file.exists(lockpath))
      return(NULL)
  
    lockfile <- tryCatch(renv_json_read(lockpath), error = identity)
    if (inherits(lockfile, "error")) {
      warning(lockfile)
      return(NULL)
    }
  
    repos <- lockfile$R$Repositories
    if (length(repos) == 0)
      return(NULL)
  
    keys <- vapply(repos, `[[`, "Name", FUN.VALUE = character(1))
    vals <- vapply(repos, `[[`, "URL", FUN.VALUE = character(1))
    names(vals) <- keys
  
    return(vals)
  
  }
  
  renv_bootstrap_download <- function(version) {
  
    sha <- attr(version, "sha", exact = TRUE)
  
    methods <- if (!is.null(sha)) {
  
      # attempting to bootstrap a development version of renv
      c(
        function() renv_bootstrap_download_tarball(sha),
        function() renv_bootstrap_download_github(sha)
      )
  
    } else {
  
      # attempting to bootstrap a release version of renv
      c(
        function() renv_bootstrap_download_tarball(version),
        function() renv_bootstrap_download_cran_latest(version),
        function() renv_bootstrap_download_cran_archive(version)
      )
  
    }
  
    for (method in methods) {
      path <- tryCatch(method(), error = identity)
      if (is.character(path) && file.exists(path))
        return(path)
    }
  
    stop("All download methods failed")
  
  }
  
  renv_bootstrap_download_impl <- function(url, destfile) {
  
    mode <- "wb"
  
    # https://bugs.r-project.org/bugzilla/show_bug.cgi?id=17715
    fixup <-
      Sys.info()[["sysname"]] == "Windows" &&
      substring(url, 1L, 5L) == "file:"
  
    if (fixup)
      mode <- "w+b"
  
    args <- list(
      url      = url,
      destfile = destfile,
      mode     = mode,
      quiet    = TRUE
    )
  
    if ("headers" %in% names(formals(utils::download.file))) {
      headers <- renv_bootstrap_download_custom_headers(url)
      if (length(headers) && is.character(headers))
        args$headers <- headers
    }
  
    do.call(utils::download.file, args)
  
  }
  
  renv_bootstrap_download_custom_headers <- function(url) {
  
    headers <- getOption("renv.download.headers")
    if (is.null(headers))
      return(character())
  
    if (!is.function(headers))
      stopf("'renv.download.headers' is not a function")
  
    headers <- headers(url)
    if (length(headers) == 0L)
      return(character())
  
    if (is.list(headers))
      headers <- unlist(headers, recursive = FALSE, use.names = TRUE)
  
    ok <-
      is.character(headers) &&
      is.character(names(headers)) &&
      all(nzchar(names(headers)))
  
    if (!ok)
      stop("invocation of 'renv.download.headers' did not return a named character vector")
  
    headers
  
  }
  
  renv_bootstrap_download_cran_latest <- function(version) {
  
    spec <- renv_bootstrap_download_cran_latest_find(version)
    type  <- spec$type
    repos <- spec$repos
  
    baseurl <- utils::contrib.url(repos = repos, type = type)
    ext <- if (identical(type, "source"))
      ".tar.gz"
    else if (Sys.info()[["sysname"]] == "Windows")
      ".zip"
    else
      ".tgz"
    name <- sprintf("renv_%s%s", version, ext)
    url <- paste(baseurl, name, sep = "/")
  
    destfile <- file.path(tempdir(), name)
    status <- tryCatch(
      renv_bootstrap_download_impl(url, destfile),
      condition = identity
    )
  
    if (inherits(status, "condition"))
      return(FALSE)
  
    # report success and return
    destfile
  
  }
  
  renv_bootstrap_download_cran_latest_find <- function(version) {
  
    # check whether binaries are supported on this system
    binary <-
      getOption("renv.bootstrap.binary", default = TRUE) &&
      !identical(.Platform$pkgType, "source") &&
      !identical(getOption("pkgType"), "source") &&
      Sys.info()[["sysname"]] %in% c("Darwin", "Windows")
  
    types <- c(if (binary) "binary", "source")
  
    # iterate over types + repositories
    for (type in types) {
      for (repos in renv_bootstrap_repos()) {
  
        # build arguments for utils::available.packages() call
        args <- list(type = type, repos = repos)
  
        # add custom headers if available -- note that
        # utils::available.packages() will pass this to download.file()
        if ("headers" %in% names(formals(utils::download.file))) {
          headers <- renv_bootstrap_download_custom_headers(repos)
          if (length(headers) && is.character(headers))
            args$headers <- headers
        }
  
        # retrieve package database
        db <- tryCatch(
          as.data.frame(
            do.call(utils::available.packages, args),
            stringsAsFactors = FALSE
          ),
          error = identity
        )
  
        if (inherits(db, "error"))
          next
  
        # check for compatible entry
        entry <- db[db$Package %in% "renv" & db$Version %in% version, ]
        if (nrow(entry) == 0)
          next
  
        # found it; return spec to caller
        spec <- list(entry = entry, type = type, repos = repos)
        return(spec)
  
      }
    }
  
    # if we got here, we failed to find renv
    fmt <- "renv %s is not available from your declared package repositories"
    stop(sprintf(fmt, version))
  
  }
  
  renv_bootstrap_download_cran_archive <- function(version) {
  
    name <- sprintf("renv_%s.tar.gz", version)
    repos <- renv_bootstrap_repos()
    urls <- file.path(repos, "src/contrib/Archive/renv", name)
    destfile <- file.path(tempdir(), name)
  
    for (url in urls) {
  
      status <- tryCatch(
        renv_bootstrap_download_impl(url, destfile),
        condition = identity
      )
  
      if (identical(status, 0L))
        return(destfile)
  
    }
  
    return(FALSE)
  
  }
  
  renv_bootstrap_find <- function(version) {
  
    path <- renv_bootstrap_find_cache(version)
    if (length(path) && file.exists(path)) {
      catf("- Using renv %s from global package cache", version)
      return(path)
    }
  
  }
  
  renv_bootstrap_find_cache <- function(version) {
  
    md5 <- attr(version, "md5", exact = TRUE)
    if (is.null(md5))
      return()
  
    # infer path to renv cache
    cache <- Sys.getenv("RENV_PATHS_CACHE", unset = "")
    if (!nzchar(cache)) {
      root <- Sys.getenv("RENV_PATHS_ROOT", unset = NA)
      if (!is.na(root))
        cache <- file.path(root, "cache")
    }
  
    if (!nzchar(cache)) {
      tools <- asNamespace("tools")
      if (is.function(tools$R_user_dir)) {
        root <- tools$R_user_dir("renv", "cache")
        cache <- file.path(root, "cache")
      }
    }
  
    # start completing path to cache
    file.path(
      cache,
      renv_bootstrap_cache_version(),
      renv_bootstrap_platform_prefix(),
      "renv",
      version,
      md5,
      "renv"
    )
  
  }
  
  renv_bootstrap_download_tarball <- function(version) {
  
    # if the user has provided the path to a tarball via
    # an environment variable, then use it
    tarball <- Sys.getenv("RENV_BOOTSTRAP_TARBALL", unset = NA)
    if (is.na(tarball))
      return()
  
    # allow directories
    if (dir.exists(tarball)) {
      name <- sprintf("renv_%s.tar.gz", version)
      tarball <- file.path(tarball, name)
    }
  
    # bail if it doesn't exist
    if (!file.exists(tarball)) {
  
      # let the user know we weren't able to honour their request
      fmt <- "- RENV_BOOTSTRAP_TARBALL is set (%s) but does not exist."
      msg <- sprintf(fmt, tarball)
      warning(msg)
  
      # bail
      return()
  
    }
  
    catf("- Using local tarball '%s'.", tarball)
    tarball
  
  }
  
  renv_bootstrap_github_token <- function() {
    for (envvar in c("GITHUB_TOKEN", "GITHUB_PAT", "GH_TOKEN")) {
      envval <- Sys.getenv(envvar, unset = NA)
      if (!is.na(envval))
        return(envval)
    }
  }
  
  renv_bootstrap_download_github <- function(version) {
  
    enabled <- Sys.getenv("RENV_BOOTSTRAP_FROM_GITHUB", unset = "TRUE")
    if (!identical(enabled, "TRUE"))
      return(FALSE)
  
    # prepare download options
    token <- renv_bootstrap_github_token()
    if (is.null(token))
      token <- ""
  
    if (nzchar(Sys.which("curl")) && nzchar(token)) {
      fmt <- "--location --fail --header \"Authorization: token %s\""
      extra <- sprintf(fmt, token)
      saved <- options("download.file.method", "download.file.extra")
      options(download.file.method = "curl", download.file.extra = extra)
      on.exit(do.call(base::options, saved), add = TRUE)
    } else if (nzchar(Sys.which("wget")) && nzchar(token)) {
      fmt <- "--header=\"Authorization: token %s\""
      extra <- sprintf(fmt, token)
      saved <- options("download.file.method", "download.file.extra")
      options(download.file.method = "wget", download.file.extra = extra)
      on.exit(do.call(base::options, saved), add = TRUE)
    }
  
    url <- file.path("https://api.github.com/repos/rstudio/renv/tarball", version)
    name <- sprintf("renv_%s.tar.gz", version)
    destfile <- file.path(tempdir(), name)
  
    status <- tryCatch(
      renv_bootstrap_download_impl(url, destfile),
      condition = identity
    )
  
    if (!identical(status, 0L))
      return(FALSE)
  
    renv_bootstrap_download_augment(destfile)
  
    return(destfile)
  
  }
  
  # Add Sha to DESCRIPTION. This is stop gap until #890, after which we
  # can use renv::install() to fully capture metadata.
  renv_bootstrap_download_augment <- function(destfile) {
    sha <- renv_bootstrap_git_extract_sha1_tar(destfile)
    if (is.null(sha)) {
      return()
    }
  
    # Untar
    tempdir <- tempfile("renv-github-")
    on.exit(unlink(tempdir, recursive = TRUE), add = TRUE)
    untar(destfile, exdir = tempdir)
    pkgdir <- dir(tempdir, full.names = TRUE)[[1]]
  
    # Modify description
    desc_path <- file.path(pkgdir, "DESCRIPTION")
    desc_lines <- readLines(desc_path)
    remotes_fields <- c(
      "RemoteType: github",
      "RemoteHost: api.github.com",
      "RemoteRepo: renv",
      "RemoteUsername: rstudio",
      "RemotePkgRef: rstudio/renv",
      paste("RemoteRef: ", sha),
      paste("RemoteSha: ", sha)
    )
    writeLines(c(desc_lines[desc_lines != ""], remotes_fields), con = desc_path)
  
    # Re-tar
    local({
      old <- setwd(tempdir)
      on.exit(setwd(old), add = TRUE)
  
      tar(destfile, compression = "gzip")
    })
    invisible()
  }
  
  # Extract the commit hash from a git archive. Git archives include the SHA1
  # hash as the comment field of the tarball pax extended header
  # (see https://www.kernel.org/pub/software/scm/git/docs/git-archive.html)
  # For GitHub archives this should be the first header after the default one
  # (512 byte) header.
  renv_bootstrap_git_extract_sha1_tar <- function(bundle) {
  
    # open the bundle for reading
    # We use gzcon for everything because (from ?gzcon)
    # > Reading from a connection which does not supply a 'gzip' magic
    # > header is equivalent to reading from the original connection
    conn <- gzcon(file(bundle, open = "rb", raw = TRUE))
    on.exit(close(conn))
  
    # The default pax header is 512 bytes long and the first pax extended header
    # with the comment should be 51 bytes long
    # `52 comment=` (11 chars) + 40 byte SHA1 hash
    len <- 0x200 + 0x33
    res <- rawToChar(readBin(conn, "raw", n = len)[0x201:len])
  
    if (grepl("^52 comment=", res)) {
      sub("52 comment=", "", res)
    } else {
      NULL
    }
  }
  
  renv_bootstrap_install <- function(version, tarball, library) {
  
    # attempt to install it into project library
    dir.create(library, showWarnings = FALSE, recursive = TRUE)
    output <- renv_bootstrap_install_impl(library, tarball)
  
    # check for successful install
    status <- attr(output, "status")
    if (is.null(status) || identical(status, 0L))
      return(status)
  
    # an error occurred; report it
    header <- "installation of renv failed"
    lines <- paste(rep.int("=", nchar(header)), collapse = "")
    text <- paste(c(header, lines, output), collapse = "\n")
    stop(text)
  
  }
  
  renv_bootstrap_install_impl <- function(library, tarball) {
  
    # invoke using system2 so we can capture and report output
    bin <- R.home("bin")
    exe <- if (Sys.info()[["sysname"]] == "Windows") "R.exe" else "R"
    R <- file.path(bin, exe)
  
    args <- c(
      "--vanilla", "CMD", "INSTALL", "--no-multiarch",
      "-l", shQuote(path.expand(library)),
      shQuote(path.expand(tarball))
    )
  
    system2(R, args, stdout = TRUE, stderr = TRUE)
  
  }
  
  renv_bootstrap_platform_prefix_default <- function() {
  
    # read version component
    version <- Sys.getenv("RENV_PATHS_VERSION", unset = "R-%v")
  
    # expand placeholders
    placeholders <- list(
      list("%v", format(getRversion()[1, 1:2])),
      list("%V", format(getRversion()[1, 1:3]))
    )
  
    for (placeholder in placeholders)
      version <- gsub(placeholder[[1L]], placeholder[[2L]], version, fixed = TRUE)
  
    # include SVN revision for development versions of R
    # (to avoid sharing platform-specific artefacts with released versions of R)
    devel <-
      identical(R.version[["status"]],   "Under development (unstable)") ||
      identical(R.version[["nickname"]], "Unsuffered Consequences")
  
    if (devel)
      version <- paste(version, R.version[["svn rev"]], sep = "-r")
  
    version
  
  }
  
  renv_bootstrap_platform_prefix <- function() {
  
    # construct version prefix
    version <- renv_bootstrap_platform_prefix_default()
  
    # build list of path components
    components <- c(version, R.version$platform)
  
    # include prefix if provided by user
    prefix <- renv_bootstrap_platform_prefix_impl()
    if (!is.na(prefix) && nzchar(prefix))
      components <- c(prefix, components)
  
    # build prefix
    paste(components, collapse = "/")
  
  }
  
  renv_bootstrap_platform_prefix_impl <- function() {
  
    # if an explicit prefix has been supplied, use it
    prefix <- Sys.getenv("RENV_PATHS_PREFIX", unset = NA)
    if (!is.na(prefix))
      return(prefix)
  
    # if the user has requested an automatic prefix, generate it
    auto <- Sys.getenv("RENV_PATHS_PREFIX_AUTO", unset = NA)
    if (is.na(auto) && getRversion() >= "4.4.0")
      auto <- "TRUE"
  
    if (auto %in% c("TRUE", "True", "true", "1"))
      return(renv_bootstrap_platform_prefix_auto())
  
    # empty string on failure
    ""
  
  }
  
  renv_bootstrap_platform_prefix_auto <- function() {
  
    prefix <- tryCatch(renv_bootstrap_platform_os(), error = identity)
    if (inherits(prefix, "error") || prefix %in% "unknown") {
  
      msg <- paste(
        "failed to infer current operating system",
        "please file a bug report at https://github.com/rstudio/renv/issues",
        sep = "; "
      )
  
      warning(msg)
  
    }
  
    prefix
  
  }
  
  renv_bootstrap_platform_os <- function() {
  
    sysinfo <- Sys.info()
    sysname <- sysinfo[["sysname"]]
  
    # handle Windows + macOS up front
    if (sysname == "Windows")
      return("windows")
    else if (sysname == "Darwin")
      return("macos")
  
    # check for os-release files
    for (file in c("/etc/os-release", "/usr/lib/os-release"))
      if (file.exists(file))
        return(renv_bootstrap_platform_os_via_os_release(file, sysinfo))
  
    # check for redhat-release files
    if (file.exists("/etc/redhat-release"))
      return(renv_bootstrap_platform_os_via_redhat_release())
  
    "unknown"
  
  }
  
  renv_bootstrap_platform_os_via_os_release <- function(file, sysinfo) {
  
    # read /etc/os-release
    release <- utils::read.table(
      file             = file,
      sep              = "=",
      quote            = c("\"", "'"),
      col.names        = c("Key", "Value"),
      comment.char     = "#",
      stringsAsFactors = FALSE
    )
  
    vars <- as.list(release$Value)
    names(vars) <- release$Key
  
    # get os name
    os <- tolower(sysinfo[["sysname"]])
  
    # read id
    id <- "unknown"
    for (field in c("ID", "ID_LIKE")) {
      if (field %in% names(vars) && nzchar(vars[[field]])) {
        id <- vars[[field]]
        break
      }
    }
  
    # read version
    version <- "unknown"
    for (field in c("UBUNTU_CODENAME", "VERSION_CODENAME", "VERSION_ID", "BUILD_ID")) {
      if (field %in% names(vars) && nzchar(vars[[field]])) {
        version <- vars[[field]]
        break
      }
    }
  
    # join together
    paste(c(os, id, version), collapse = "-")
  
  }
  
  renv_bootstrap_platform_os_via_redhat_release <- function() {
  
    # read /etc/redhat-release
    contents <- readLines("/etc/redhat-release", warn = FALSE)
  
    # infer id
    id <- if (grepl("centos", contents, ignore.case = TRUE))
      "centos"
    else if (grepl("redhat", contents, ignore.case = TRUE))
      "redhat"
    else
      "unknown"
  
    # try to find a version component (very hacky)
    version <- "unknown"
  
    parts <- strsplit(contents, "[[:space:]]")[[1L]]
    for (part in parts) {
  
      nv <- tryCatch(numeric_version(part), error = identity)
      if (inherits(nv, "error"))
        next
  
      version <- nv[1, 1]
      break
  
    }
  
    paste(c("linux", id, version), collapse = "-")
  
  }
  
  renv_bootstrap_library_root_name <- function(project) {
  
    # use project name as-is if requested
    asis <- Sys.getenv("RENV_PATHS_LIBRARY_ROOT_ASIS", unset = "FALSE")
    if (asis)
      return(basename(project))
  
    # otherwise, disambiguate based on project's path
    id <- substring(renv_bootstrap_hash_text(project), 1L, 8L)
    paste(basename(project), id, sep = "-")
  
  }
  
  renv_bootstrap_library_root <- function(project) {
  
    prefix <- renv_bootstrap_profile_prefix()
  
    path <- Sys.getenv("RENV_PATHS_LIBRARY", unset = NA)
    if (!is.na(path))
      return(paste(c(path, prefix), collapse = "/"))
  
    path <- renv_bootstrap_library_root_impl(project)
    if (!is.null(path)) {
      name <- renv_bootstrap_library_root_name(project)
      return(paste(c(path, prefix, name), collapse = "/"))
    }
  
    renv_bootstrap_paths_renv("library", project = project)
  
  }
  
  renv_bootstrap_library_root_impl <- function(project) {
  
    root <- Sys.getenv("RENV_PATHS_LIBRARY_ROOT", unset = NA)
    if (!is.na(root))
      return(root)
  
    type <- renv_bootstrap_project_type(project)
    if (identical(type, "package")) {
      userdir <- renv_bootstrap_user_dir()
      return(file.path(userdir, "library"))
    }
  
  }
  
  renv_bootstrap_validate_version <- function(version, description = NULL) {
  
    # resolve description file
    #
    # avoid passing lib.loc to `packageDescription()` below, since R will
    # use the loaded version of the package by default anyhow. note that
    # this function should only be called after 'renv' is loaded
    # https://github.com/rstudio/renv/issues/1625
    description <- description %||% packageDescription("renv")
  
    # check whether requested version 'version' matches loaded version of renv
    sha <- attr(version, "sha", exact = TRUE)
    valid <- if (!is.null(sha))
      renv_bootstrap_validate_version_dev(sha, description)
    else
      renv_bootstrap_validate_version_release(version, description)
  
    if (valid)
      return(TRUE)
  
    # the loaded version of renv doesn't match the requested version;
    # give the user instructions on how to proceed
    dev <- identical(description[["RemoteType"]], "github")
    remote <- if (dev)
      paste("rstudio/renv", description[["RemoteSha"]], sep = "@")
    else
      paste("renv", description[["Version"]], sep = "@")
  
    # display both loaded version + sha if available
    friendly <- renv_bootstrap_version_friendly(
      version = description[["Version"]],
      sha     = if (dev) description[["RemoteSha"]]
    )
  
    fmt <- heredoc("
      renv %1$s was loaded from project library, but this project is configured to use renv %2$s.
      - Use `renv::record(\"%3$s\")` to record renv %1$s in the lockfile.
      - Use `renv::restore(packages = \"renv\")` to install renv %2$s into the project library.
    ")
    catf(fmt, friendly, renv_bootstrap_version_friendly(version), remote)
  
    FALSE
  
  }
  
  renv_bootstrap_validate_version_dev <- function(version, description) {
  
    expected <- description[["RemoteSha"]]
    if (!is.character(expected))
      return(FALSE)
  
    pattern <- sprintf("^\\Q%s\\E", version)
    grepl(pattern, expected, perl = TRUE)
  
  }
  
  renv_bootstrap_validate_version_release <- function(version, description) {
    expected <- description[["Version"]]
    is.character(expected) && identical(c(expected), c(version))
  }
  
  renv_bootstrap_hash_text <- function(text) {
  
    hashfile <- tempfile("renv-hash-")
    on.exit(unlink(hashfile), add = TRUE)
  
    writeLines(text, con = hashfile)
    tools::md5sum(hashfile)
  
  }
  
  renv_bootstrap_load <- function(project, libpath, version) {
  
    # try to load renv from the project library
    if (!requireNamespace("renv", lib.loc = libpath, quietly = TRUE))
      return(FALSE)
  
    # warn if the version of renv loaded does not match
    renv_bootstrap_validate_version(version)
  
    # execute renv load hooks, if any
    hooks <- getHook("renv::autoload")
    for (hook in hooks)
      if (is.function(hook))
        tryCatch(hook(), error = warnify)
  
    # load the project
    renv::load(project)
  
    TRUE
  
  }
  
  renv_bootstrap_profile_load <- function(project) {
  
    # if RENV_PROFILE is already set, just use that
    profile <- Sys.getenv("RENV_PROFILE", unset = NA)
    if (!is.na(profile) && nzchar(profile))
      return(profile)
  
    # check for a profile file (nothing to do if it doesn't exist)
    path <- renv_bootstrap_paths_renv("profile", profile = FALSE, project = project)
    if (!file.exists(path))
      return(NULL)
  
    # read the profile, and set it if it exists
    contents <- readLines(path, warn = FALSE)
    if (length(contents) == 0L)
      return(NULL)
  
    # set RENV_PROFILE
    profile <- contents[[1L]]
    if (!profile %in% c("", "default"))
      Sys.setenv(RENV_PROFILE = profile)
  
    profile
  
  }
  
  renv_bootstrap_profile_prefix <- function() {
    profile <- renv_bootstrap_profile_get()
    if (!is.null(profile))
      return(file.path("profiles", profile, "renv"))
  }
  
  renv_bootstrap_profile_get <- function() {
    profile <- Sys.getenv("RENV_PROFILE", unset = "")
    renv_bootstrap_profile_normalize(profile)
  }
  
  renv_bootstrap_profile_set <- function(profile) {
    profile <- renv_bootstrap_profile_normalize(profile)
    if (is.null(profile))
      Sys.unsetenv("RENV_PROFILE")
    else
      Sys.setenv(RENV_PROFILE = profile)
  }
  
  renv_bootstrap_profile_normalize <- function(profile) {
  
    if (is.null(profile) || profile %in% c("", "default"))
      return(NULL)
  
    profile
  
  }
  
  renv_bootstrap_path_absolute <- function(path) {
  
    substr(path, 1L, 1L) %in% c("~", "/", "\\") || (
      substr(path, 1L, 1L) %in% c(letters, LETTERS) &&
      substr(path, 2L, 3L) %in% c(":/", ":\\")
    )
  
  }
  
  renv_bootstrap_paths_renv <- function(..., profile = TRUE, project = NULL) {
    renv <- Sys.getenv("RENV_PATHS_RENV", unset = "renv")
    root <- if (renv_bootstrap_path_absolute(renv)) NULL else project
    prefix <- if (profile) renv_bootstrap_profile_prefix()
    components <- c(root, renv, prefix, ...)
    paste(components, collapse = "/")
  }
  
  renv_bootstrap_project_type <- function(path) {
  
    descpath <- file.path(path, "DESCRIPTION")
    if (!file.exists(descpath))
      return("unknown")
  
    desc <- tryCatch(
      read.dcf(descpath, all = TRUE),
      error = identity
    )
  
    if (inherits(desc, "error"))
      return("unknown")
  
    type <- desc$Type
    if (!is.null(type))
      return(tolower(type))
  
    package <- desc$Package
    if (!is.null(package))
      return("package")
  
    "unknown"
  
  }
  
  renv_bootstrap_user_dir <- function() {
    dir <- renv_bootstrap_user_dir_impl()
    path.expand(chartr("\\", "/", dir))
  }
  
  renv_bootstrap_user_dir_impl <- function() {
  
    # use local override if set
    override <- getOption("renv.userdir.override")
    if (!is.null(override))
      return(override)
  
    # use R_user_dir if available
    tools <- asNamespace("tools")
    if (is.function(tools$R_user_dir))
      return(tools$R_user_dir("renv", "cache"))
  
    # try using our own backfill for older versions of R
    envvars <- c("R_USER_CACHE_DIR", "XDG_CACHE_HOME")
    for (envvar in envvars) {
      root <- Sys.getenv(envvar, unset = NA)
      if (!is.na(root))
        return(file.path(root, "R/renv"))
    }
  
    # use platform-specific default fallbacks
    if (Sys.info()[["sysname"]] == "Windows")
      file.path(Sys.getenv("LOCALAPPDATA"), "R/cache/R/renv")
    else if (Sys.info()[["sysname"]] == "Darwin")
      "~/Library/Caches/org.R-project.R/R/renv"
    else
      "~/.cache/R/renv"
  
  }
  
  renv_bootstrap_version_friendly <- function(version, shafmt = NULL, sha = NULL) {
    sha <- sha %||% attr(version, "sha", exact = TRUE)
    parts <- c(version, sprintf(shafmt %||% " [sha: %s]", substring(sha, 1L, 7L)))
    paste(parts, collapse = "")
  }
  
  renv_bootstrap_exec <- function(project, libpath, version) {
    if (!renv_bootstrap_load(project, libpath, version))
      renv_bootstrap_run(project, libpath, version)
  }
  
  renv_bootstrap_run <- function(project, libpath, version) {
    tryCatch(
      renv_bootstrap_run_impl(project, libpath, version),
      error = function(e) {
        msg <- paste(
          "failed to bootstrap renv: the project will not be loaded.",
          paste("Reason:", conditionMessage(e)),
          "Use `renv::activate()` to re-initialize the project.",
          sep = "\n"
        )
        warning(msg, call. = FALSE)
      }
    )
  }
  
  renv_bootstrap_run_impl <- function(project, libpath, version) {
  
    # perform bootstrap
    bootstrap(version, libpath)
  
    # exit early if we're just testing bootstrap
    if (!is.na(Sys.getenv("RENV_BOOTSTRAP_INSTALL_ONLY", unset = NA)))
      return(TRUE)
  
    # try again to load
    if (requireNamespace("renv", lib.loc = libpath, quietly = TRUE)) {
      return(renv::load(project = project))
    }
  
    # failed to download or load renv; warn the user
    msg <- c(
      "Failed to find an renv installation: the project will not be loaded.",
      "Use `renv::activate()` to re-initialize the project."
    )
  
    warning(paste(msg, collapse = "\n"), call. = FALSE)
  
  }
  
  renv_bootstrap_cache_version <- function() {
    # NOTE: users should normally not override the cache version;
    # this is provided just to make testing easier
    Sys.getenv("RENV_CACHE_VERSION", unset = "v5")
  }
  
  renv_bootstrap_cache_version_previous <- function() {
    version <- renv_bootstrap_cache_version()
    number <- as.integer(substring(version, 2L))
    paste("v", number - 1L, sep = "")
  }
  
  renv_json_read <- function(file = NULL, text = NULL) {
  
    jlerr <- NULL
  
    # if jsonlite is loaded, use that instead
    if ("jsonlite" %in% loadedNamespaces()) {
  
      json <- tryCatch(renv_json_read_jsonlite(file, text), error = identity)
      if (!inherits(json, "error"))
        return(json)
  
      jlerr <- json
  
    }
  
    # otherwise, fall back to the default JSON reader
    json <- tryCatch(renv_json_read_default(file, text), error = identity)
    if (!inherits(json, "error"))
      return(json)
  
    # report an error
    if (!is.null(jlerr))
      stop(jlerr)
    else
      stop(json)
  
  }
  
  renv_json_read_jsonlite <- function(file = NULL, text = NULL) {
    text <- paste(text %||% readLines(file, warn = FALSE), collapse = "\n")
    jsonlite::fromJSON(txt = text, simplifyVector = FALSE)
  }
  
  renv_json_read_patterns <- function() {
  
    list(
  
      # objects
      list("{", "\t\n\tobject(\t\n\t", TRUE),
      list("}", "\t\n\t)\t\n\t",       TRUE),
  
      # arrays
      list("[", "\t\n\tarray(\t\n\t", TRUE),
      list("]", "\n\t\n)\n\t\n",      TRUE),
  
      # maps
      list(":", "\t\n\t=\t\n\t", TRUE),
  
      # newlines
      list("\\u000a", "\n", FALSE)
  
    )
  
  }
  
  renv_json_read_envir <- function() {
  
    envir <- new.env(parent = emptyenv())
  
    envir[["+"]] <- `+`
    envir[["-"]] <- `-`
  
    envir[["object"]] <- function(...) {
      result <- list(...)
      names(result) <- as.character(names(result))
      result
    }
  
    envir[["array"]] <- list
  
    envir[["true"]]  <- TRUE
    envir[["false"]] <- FALSE
    envir[["null"]]  <- NULL
  
    envir
  
  }
  
  renv_json_read_remap <- function(object, patterns) {
  
    # repair names if necessary
    if (!is.null(names(object))) {
  
      nms <- names(object)
      for (pattern in patterns)
        nms <- gsub(pattern[[2L]], pattern[[1L]], nms, fixed = TRUE)
      names(object) <- nms
  
    }
  
    # repair strings if necessary
    if (is.character(object)) {
      for (pattern in patterns)
        object <- gsub(pattern[[2L]], pattern[[1L]], object, fixed = TRUE)
    }
  
    # recurse for other objects
    if (is.recursive(object))
      for (i in seq_along(object))
        object[i] <- list(renv_json_read_remap(object[[i]], patterns))
  
    # return remapped object
    object
  
  }
  
  renv_json_read_default <- function(file = NULL, text = NULL) {
  
    # read json text
    text <- paste(text %||% readLines(file, warn = FALSE), collapse = "\n")
  
    # convert into something the R parser will understand
    patterns <- renv_json_read_patterns()
    transformed <- text
    for (pattern in patterns)
      transformed <- gsub(pattern[[1L]], pattern[[2L]], transformed, fixed = TRUE)
  
    # parse it
    rfile <- tempfile("renv-json-", fileext = ".R")
    on.exit(unlink(rfile), add = TRUE)
    writeLines(transformed, con = rfile)
    json <- parse(rfile, keep.source = FALSE, srcfile = NULL)[[1L]]
  
    # evaluate in safe environment
    result <- eval(json, envir = renv_json_read_envir())
  
    # fix up strings if necessary -- do so only with reversible patterns
    patterns <- Filter(function(pattern) pattern[[3L]], patterns)
    renv_json_read_remap(result, patterns)
  
  }
  

  # load the renv profile, if any
  renv_bootstrap_profile_load(project)

  # construct path to library root
  root <- renv_bootstrap_library_root(project)

  # construct library prefix for platform
  prefix <- renv_bootstrap_platform_prefix()

  # construct full libpath
  libpath <- file.path(root, prefix)

  # run bootstrap code
  renv_bootstrap_exec(project, libpath, version)

  invisible()

})
```
